# Positive and negative affect, related mental health traits, and cognitive performance: shared genetic architecture and potential causality

**DOI:** 10.1101/2024.11.01.24316562

**Authors:** Chloe Slaney, Naoise Mac Giollabhui, Peter J. van der Most, Ensor R. Palacios, Harold Snieder, Michel Nivard, Gibran Hemani, Catharina A. Hartman, Golam M. Khandaker

**Author notes:** Joint Senior Author (Catharina A. Hartman, Golam M. Khandaker). **Corresponding author:** Dr Chloe Slaney, MRC Integrative Epidemiology Unit, University of Bristol, Oakfield House, Oakfield Grove, Bristol, BS8 2BN,.

## Abstract

Altered affect and cognitive dysfunction are transdiagnostic, burdensome, and pervasive features of many psychiatric conditions which remain poorly understood and have few efficacious treatments. Research on the genetic architecture of these phenotypes and causal relationships between them may provide insight into their aetiology and comorbidity. Using data from the Lifelines Cohort Study, we conducted genome-wide association studies (GWAS) on positive and negative affect and four cognitive domains (working memory, reaction time, visual learning and memory, executive function). Using publicly available large GWAS on related - albeit distinct-phenotypes (depression, anxiety, wellbeing, general cognitive ability [GCA]) we conducted genetic correlation and Mendelian randomization (MR) analyses to examine genetic overlap and causal relationships. We identified one genome-wide hit (*p*<5×10^-8^) for reaction time, and many loci with suggestive associations (*p*<5×10^-6^; N range= 11-20 independent hits) for other phenotypes. For most phenotypes, gene mapping and tissue expression analysis of suggestive hits from the GWAS showed increased gene expression in brain tissue compared to other tissues. As predicted, negative affect is genetically correlated with mental health phenotypes (depression *r*_g_=0.51; anxiety *r*_g_=0.70; wellbeing *r*_g_ = −0.71) and cognitive domains are genetically correlated with GCA and brain volume (*r*_g_ ≤ 0.66). Genetic correlations between negative and positive affect suggest that they are dissociable constructs (*r*_g_ = −0.18) with negative affect having higher genetic overlap with GCA than positive affect (*r*_g_ =-0.19 vs −0.06). This could indicate that negative affect has a higher shared neural basis with GCA than positive affect and/or GCA and negative affect may exhibit causal relationships. MR analyses suggest potential causal effects of higher GCA on reduced negative affect, reduced risk of depression and anxiety, and higher wellbeing, but little impact on positive affect. We also report evidence for potential causal effects of depression and lower wellbeing on reduced GCA. Taken together, these results suggests that GCA may be a valid target for negative affect (but not positive affect) and depression and wellbeing may be valid targets for GCA.

## 1. Introduction

Positive and negative affect reflect the extent to which a person feels positive or negative emotions (e.g., excited, interested; nervous, distressed), respectively. Positive and negative affect can be experienced in a state (temporary) or trait (stable) manner (Watson et al., 1988) and are distinct transdiagnostic features of many health conditions, particularly depression and anxiety. Another critical yet often overlooked transdiagnostic feature of many health conditions is cognitive dysfunction (Colwell et al., 2022; Hilton et al., 2024). Despite altered affect and cognitive dysfunction being burdensome and pervasive features of many conditions, they remain poorly understood and have few efficacious treatments (Colwell et al., 2022; Rush et al., 2006).

Understanding the genetic architecture of mental health and cognitive phenotypes, and their relation to each other, can provide aetiological and biological insights which may facilitate identification of novel treatment targets (Carey et al., 2021; Minikel et al., 2024). Although observational studies have found associations between mental health and cognition (Dam et al., 2021; Rock et al., 2014; Zainal & Newman, 2021; although see Ball et al., 2024;), causality remains unclear (Suddell et al., 2023). Reported associations may reflect: (1) poorer mental health causing poorer cognition, (2) poorer cognition causing poorer mental health, (3) confounding via shared risk factors (e.g., stress, sedentary behaviour) (Mac Giollabhui, 2021). Crucially, studies testing causal relationships between more narrowly defined mental health phenotypes (e.g., affect) and cognitive domains (e.g., memory, attention) are needed (Chavez-Baldini et al., 2023). Evidence of bidirectional causality would strengthen the case that targeting poor mental health (either diagnostic conditions or transdiagnostic phenotypes) may prevent and treat cognitive dysfunction, and *vice versa*. Two approaches that can shed light on the aetiology, comorbidity, and/or causal relationships between phenotypes are genetic correlations (using all genome-wide variants) (Bulik-Sullivan, Finucane, et al., 2015) and Mendelian randomization (MR) (typically using genome-wide association study hits) (Davey Smith & Ebrahim, 2003). MR can test for causality given certain assumptions are met (see Supplementary Methods). It does this by using genetic variants robustly associated with the exposure as a proxy for it. The properties of genetic variants (random assignment from parents, fixed at conception) mean that they are less likely to be associated with confounders and overcome issues of reverse causality (Davey Smith & Ebrahim, 2003; Sanderson et al., 2022). These methods provide powerful tools given the availability of large genome-wide association studies (GWAS) on well-defined phenotypes.

To date, GWAS on mental health phenotypes have largely been conducted on diagnostic categories such as depression and anxiety (Carey et al., 2021). However, these conditions are heterogeneous, with diagnosed individuals showing diverse symptom profiles. For example, a diagnosis of depression requires ≥5/9 *diverse* symptoms to be present within a two-week period, one of which must be low mood or anhedonia (Regier et al., 2013). In addition, symptoms are often not specific to a condition, meaning that GWAS on a condition may in reality capture a broader phenotype. One potential approach to reducing phenotypic heterogeneity while addressing the overlap with other conditions, is to focus on *more narrowly defined* traits that more closely map onto biological systems (positive and negative affect, specific cognitive domains) (Wendt et al., 2020). In line with the Research Domain Criteria framework (Cuthbert, 2014; Insel et al., 2010), these are often transdiagnostic. GWAS on these more narrowly defined transdiagnostic traits may potentially facilitate insights into aetiology, comorbidity, and novel therapeutics.

In this study, we first conducted GWAS of negative and positive affect (assessed using the Positive and Negative Affect Schedule [PANAS]) and cognitive task performance in several domains (executive function, working memory, visual learning and memory, and reaction time). Second, we conducted gene mapping and tissue expression analysis to gain greater insight into associated genetic variants. Third, we investigated genetic overlap between mental health (negative affect, positive affect, depression, anxiety, wellbeing) and cognitive (general cognitive ability [GCA], specific cognitive domains) phenotypes using genetic correlations. Fourth, we tested evidence of potential causality between mental health and cognitive phenotypes using MR analyses. This is the first GWAS conducted on positive and negative affect using well-validated measures of these phenotypes. To achieve this, we use a large Dutch population-based cohort, the Lifelines Cohort. The application of MR to test evidence of causal relationships between affect and cognition is critical for improving our conceptual understanding of these phenotypes.

We hypothesised that: (i) follow-up analyses on all GWAS show increased gene expression in brain tissue compared to other tissues; (ii) affect and cognitive measures in the Lifelines Cohort genetically correlate with related (albeit distinct) phenotypes from external GWAS. Specifically, negative affect positively correlates with depression/anxiety and negatively correlates with wellbeing (reverse direction for positive affect), and higher cognitive performance on all domains correlates with GCA; (iii) there are bidirectional causal relationships between mental health and cognitive phenotypes.

## 2. Methods and Materials

### 2.1. Description of the Lifelines Cohort

Lifelines is a multi-disciplinary prospective population-based cohort study examining in a unique three-generation design the health and health-related behaviours of 167,729 persons living in the north of the Netherlands. It employs a broad range of investigative procedures in assessing the biomedical, socio-demographic, behavioural, physical and psychological factors which contribute to the health and disease of the general population, with a special focus on multi-morbidity and complex genetics. Participants were recruited between 2006-2013 via their GP (49%), participating family members (38%), and self-registration on the Lifelines website (13%) (Scholtens et al., 2015). Exclusion criteria for GP recruitment were: insufficient knowledge of Dutch language, severe psychiatric or physical illness, limited life expectancy (<5 years) (Scholtens et al., 2015). Baseline data included approximately: 15,000 children (0-17 years), 140,000 adults (18-65 years), 12,000 elderly individuals (65+ years). Following baseline, participants completed follow-up questionnaires every 1.5 years and study visits every 5 years (1st follow-up visit [2014-2017], 2nd follow-up visit [2019-2023]) (Scholtens et al., 2015; Sijtsma et al., 2022). In this study, we included participants ≥18 years who were genotyped on a GWAS array and excluded participants who have conditions with a significant cognitive sequel: Alzheimer’s disease, other dementia, epilepsy, multiple sclerosis, Parkinson’s disease, and stroke.

### 2.2. Assessment of Positive and Negative Affect

The Positive and Negative Affect Scale (PANAS) was used. PANAS assesses positive and negative affect using two separate sub-scales, each containing 10 items (example items: excited, upset, nervous) (Crawford & Henry, 2004; Watson et al., 1988). Participants rate the extent they experienced each item during the last four weeks on a five-point scale (ranging from ‘not at all’ to ‘extremely’). The outcome is the summed score on each subscale, which ranges from 10-50 (higher values reflect higher positive or negative affect). The scales have high internal consistency (*a*=.87) and moderate test-retest reliability over 8-weeks (positive *r*=.58; negative *r*=.48) (Watson et al., 1988).

### 2.3. Assessment of Cognitive Performance

Ruff Figural Fluency Test (RFFT) was used, which is a valid and reliable measure of executive functioning (Ross, 2014). The task consists of five parts, each containing 35 identical five-dot patterns. Participants draw as many unique designs as possible within one minute by connecting dots in different patterns (Kuiper et al., 2017). The primary outcome is total number of unique designs.

Four tasks from the CogState Test Battery were administered in Lifelines, which tap into specific cognitive domains: visual learning & memory (one card learning task), reaction time/attention (identification task and detection task) and working memory (one-back task). As the detection and identification task both assess reaction time, we include only the identification task in this study which has a larger sample size. We used the recommended primary outcomes for all tasks. All outcomes were transformed (reaction time [ms] on the identification task was log10 transformed, and accuracy on the one-back and one card learning task were arcsine transformed). On both memory tasks, higher values reflect better memory; on the reaction time task higher values reflect poorer performance (i.e., slower reaction time). See Supplementary Methods for further information on dataset preparation.

### 2.4. Genetics Data and Quality Control

Genotyping was conducted using three chip arrays in three subsets of the Lifelines cohort: (i) Illumina CytoSNP-12 Bead Chip v2 array (N=17,033), (ii) Infinium Global Screening Array (GSA) Beadchip-24 v1.0 (N=38,030), (iii) FinnGen Thermo Fisher Axiom® custom array (Affymetrix; N=29,166). See Supplementary for quality control (QC) and imputation procedures conducted by Lifelines. Following Lifelines QCs, 73,086 participants were potentially eligible for this study (CytoSNP N=14,942; GSA N=31,810; Affymetrix N=26,334). We applied additional QCs, removing: (i) duplicates (individuals genotyped on >1 chip) and 1st-degree relatives between chips, (ii) non-European individuals, and (iii) genetic outliers, see Supplementary Figure S1. Following additional QCs, 58,713 participants with genetics data were included in this study (CytoSNP N=7,632; GSA N=24,975; Affymetrix N=26,106).

### 2.5. GWAS Summary Statistics for Related Mental Health and Cognitive Phenotypes

To examine genetic overlap and potential causality between affect and cognitive performance, we used summary statistics from large publicly available GWAS of depression (N Cases=294,322; N Controls=741,438) (Als et al., 2023), anxiety (N Cases=7,016; N Controls=14,745)(Otowa et al., 2016), wellbeing (N=2,370,390) (Baselmans et al., 2019), general cognitive ability (GCA; N=373,617)(Lam et al., 2021), and brain volume (N=47,316) (Jansen et al., 2020). See Supplementary Methods and Table S1-S2 for further details on these GWAS.

### 2.6. Statistical Analyses

#### 2.6.1. GWAS of Negative Affect, Positive Affect, and Cognitive Performance in Lifelines

We conducted GWAS (negative affect score, positive affect score, and cognitive performance score on each task) in each genotyped subset separately using REGENIE, which accounts for relatedness (Mbatchou et al., 2021). Prior to conducting GWAS, within each subset, we standardised all outcomes (mean=0; SD=1) and used PLINK (Purcell et al., 2007) to clean the genetics data for REGENIE Step 1. We included variants that met the following criteria: call rate (0.95), Hardy-Weinberg equilibrium (1e-6), minor allele count (100), minor allele frequency (0.01), not multi-allelic; and restricted to individuals with low missingness (0.95). In each GWAS, covariates included age, sex, and top 10 genetic principal components. We excluded poorly imputed variants (INFO score <0.8). For each outcome, we meta-analysed GWAS across the three subsets using STDERR model in METAL and applied genomic corrections (Willer et al., 2010). GWAS quality was inspected using GWAS Inspector 1.6.4.0 (Ani et al., 2021). The standard p-value threshold of 5×10^-8^ was used to determine genome-wide significance (for suggestive significance: p-value <5×10^-6^). Variants with MAF < 0.01 were excluded from all follow-up analyses.

#### 2.6.2. Tissue Specificity of Prioritised Genes

We used gene and tissue mapping to explore associated variants in each GWAS. We used an online platform (FUMA) which integrates resources to annotate, prioritize, and visualise the summary statistics (Watanabe et al., 2017). The SNP2GENE function was used to annotate SNPs and prioritise genes at each locus using positional mapping. Using these prioritised genes, we used GENE2FUNC to investigate tissue specificity of differentially expressed gene (DEG) sets using GTEx v8 data on 54 non-diseased tissue types (N≤838 adults). For further details, see Supplementary Methods.

### 2.6.3. Genetic Correlations

Linkage Disequilibrium Score Regression (LDSR) (Bulik-Sullivan et al., 2015; Bulik-Sullivan et al., 2015) was used to estimate genetic correlations of mental health and cognitive phenotypes. This included phenotypes from Lifelines (positive and negative affect, cognitive performance), and phenotypes from large external GWAS (depression, anxiety, wellbeing, GCA, brain volume). We used precomputed LD scores calculated using 1000G European data. As low imputation quality may confound LDSR we filtered to HapMap3 panel SNPs which tend to be well-imputed in most studies (Bulik-Sullivan, Finucane, et al., 2015). If GWAS did not have a sample size column for SNPs, we assumed the same sample size for all SNPs. We checked all phenotypes had heritability z-scores > 4 in line with standard procedure (Bulik-Sullivan et al., 2015). For LDSR cleaning filters applied, see (Bulik-Sullivan et al., 2015).

### 2.7. Bidirectional Mendelian Randomization

MR was performed to investigate potential causal effect of mental health phenotypes on cognition and *vice versa*. This was done in R v4.2.2 using *TwoSampleMR* (Hemani et al., 2017) and *CAUSE* v1.2.0.0335 (Morrison et al., 2020).

We applied the following criteria to identify genetic instruments: (1) *p*<5×10^-8^ (if not available, a lenient *p*<5×10^-6^ threshold was used) (2) independent (r^2^ < 0.01, kb = 1000; based on European clustering in the 1000 genomes reference panel using *ld_clump()* in the *ieugwasr* package (Hemani et al., 2024)), (3) minor allele frequency (MAF) ≥1%. Any SNPs not available in the outcome GWAS were excluded. For primary analyses, we used the Inverse-Variance Weighted method (>1 SNP available) or Wald ratio (if only 1 SNP was available). Where multiple genetic variants were available, we conducted sensitivity analyses using different MR methods which have different assumptions regarding the validity of the genetic instruments: MR-Egger (Bowden et al., 2015), weighted median (Bowden et al., 2016), weighted mode (Hartwig et al., 2017).

We also applied Steiger filtering to assess whether genetic variants have stronger associations with exposures than outcomes (Hemani et al., 2017). Given the possibility of correlated pleiotropy, we additionally applied causal analysis using summary effect estimates (CAUSE) which accounts for correlated and uncorrelated pleiotropy (Morrison et al., 2020). For CAUSE, we included genome-wide variants pruned using default criteria (r^2^=0.01, p-value=0.003) based on European 1000 genomes reference panel. Given the possibility of population-level confounding (e.g., dynastic effects), we also conducted within-sibship MR in the MR-base platform (Hemani et al., 2018) using publicly available within-sibship GWAS on wellbeing, depressive symptoms, and cognitive ability (Howe et al., 2022); see Supplementary methods for more detail. We also checked for heterogeneity (Cochran’s Q-statistic) and pleiotropy (Egger intercept). For further details on assumptions of different MR methods used here, see Supplementary methods.

In our primary analysis, we included phenotypes which have well-powered GWAS as exposures (>1 SNP available: depression, wellbeing, GCA). In secondary analysis, we included phenotypes which have less well-powered GWAS as exposures (1 SNP or lenient p-value criteria: anxiety, negative affect, positive affect) to investigate whether similar patterns of effects are observed for these phenotypes.

## 3. Results

### 3.1. GWAS of Negative Affect, Positive Affect, and Cognitive Performance in the Lifelines Cohort

No SNPs met genome-wide significance threshold (*p*<5×10^-8^), except one SNP for reaction time (rs2920287, MAF=0.04, *p*=1.907e-08). Visual inspection of the locus zoom plot of the region around rs2920287 shows many nearby genes, with PSCA being in closest proximity (Figure S4). Follow-up analyses of this SNP using the GWAS Catalog did not show associations with any traits. Lowering the threshold to *p*<5×10^-6^ yielded associated SNPs for all phenotypes (Table 1). For Manhattan plots, see Figures S2-S3.

**Table 1.**
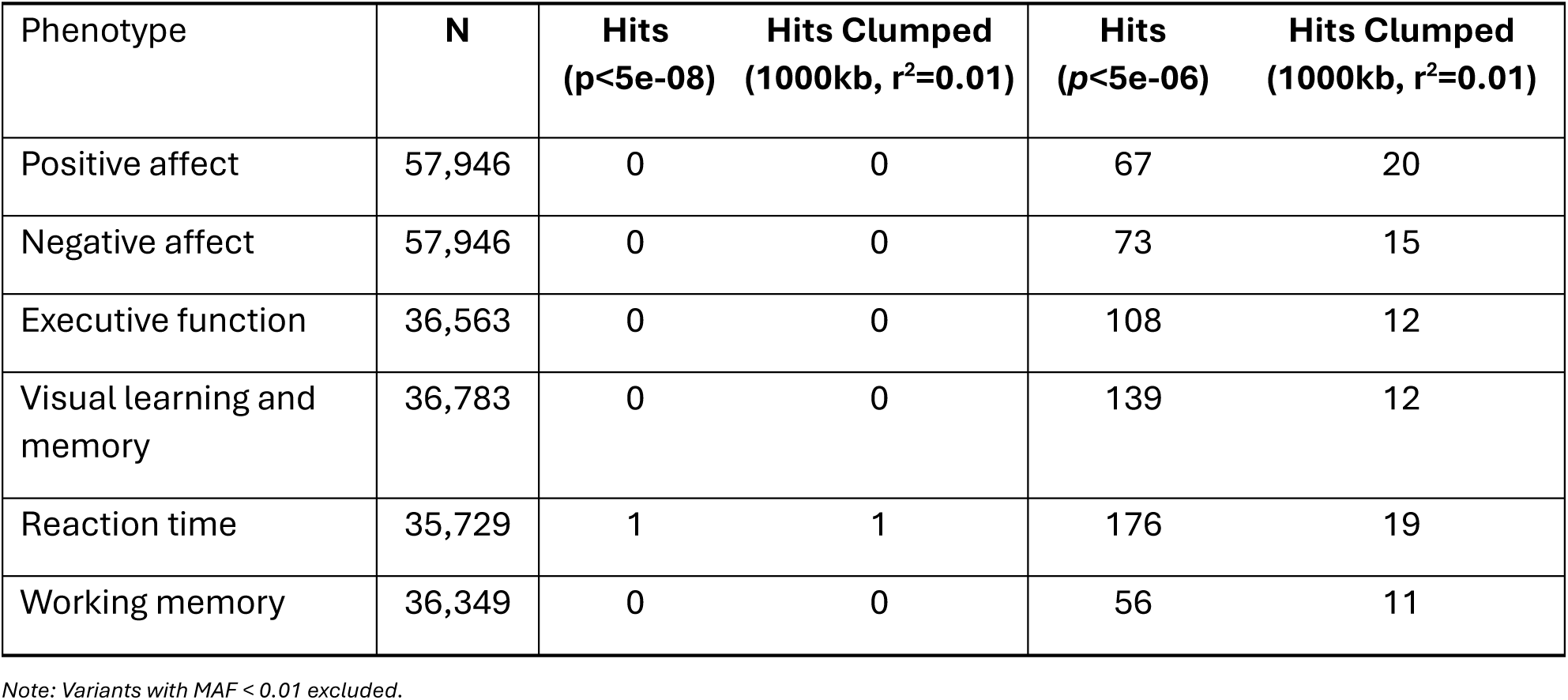
GWAS Results of Positive Affect, Negative Affect, and Cognitive Performance in the Lifelines cohort.

### 3.2. Tissue Specificity of Prioritised Genes

We used a lenient p-value threshold (*p*<5×10^-6^) to identify SNPs for follow-up analysis in FUMA. For all phenotype’s, prioritised genes were upregulated across multiple tissue types, particularly brain tissue (see Supplementary Figures S5-S6 and Tables S18-S23). Following FUMAs default Bonferroni corrections for multiple comparisons, there was significant upregulation in brain tissue for positive affect (substantia nigra) and visual learning and memory (cerebellum and cerebellar hemispheres) compared to other tissue types. For visualisation of tissue specificity of prioritised genes, see Figures S5-S6.

### 3.3. Genetic Correlations between Affect and Mental Health Phenotypes

#### 3.3.1. Negative Affect is Genetically Correlated with Mental Health Phenotypes

Negative affect score was genetically correlated with depression (*r*_g_ = 0.51 [SE=0.05], *p*=2.80E-28), wellbeing (*r*_g_ = −0.71 [SE=0.05], *p*=8.65E-41), and anxiety (*r*_g_ = 0.70 [SE=0.18], *p*=0.0002). Positive affect showed weaker genetic correlations with depression (*r*_g_ = −0.11 [SE=0.04], *p*=0.005), wellbeing (*r*_g_ = 0.30 [SE=0.05], *p*=2.38E-09), with little evidence for anxiety (*r*_g_ = −0.16 [SE=0.17], *p*=0.33).

#### 3.3.2. Cognitive Domains are Genetically Correlated with GCA

Cognitive performance in Lifelines tasks were genetically correlated with the largest GWAS for GCA: executive function (*r*_g_=0.66 [SE=0.07], *p*=1.39E-23), visual learning & memory (*r*_g_=0.54 [SE=0.05], *p*=6.90E-29), working memory (*r*_g_=0.53 [SE=0.06], *p*=1.23E-18), and reaction time (*r*_g_= −0.39 [SE=0.06], *p*=1.48E-12).

#### 3.3.3. Compared to Positive Affect, Negative Affect has Stronger Genetic Correlation with GCA

There was a weak negative genetic correlation between positive and negative affect scores (*r*_g_ = −0.18 [SE=0.08], *p*=0.016). There was stronger evidence of genetic correlations between GCA and negative affect (*r*_g_ = −0.19 [SE=0.04], *p*=7.56E-06) compared to GCA and positive affect (*r*_g_ = −0.06 [SE=0.04], *p*=0.18).

For all results, see Figure 2 and Supplementary Table S3.

**Figure 1.**
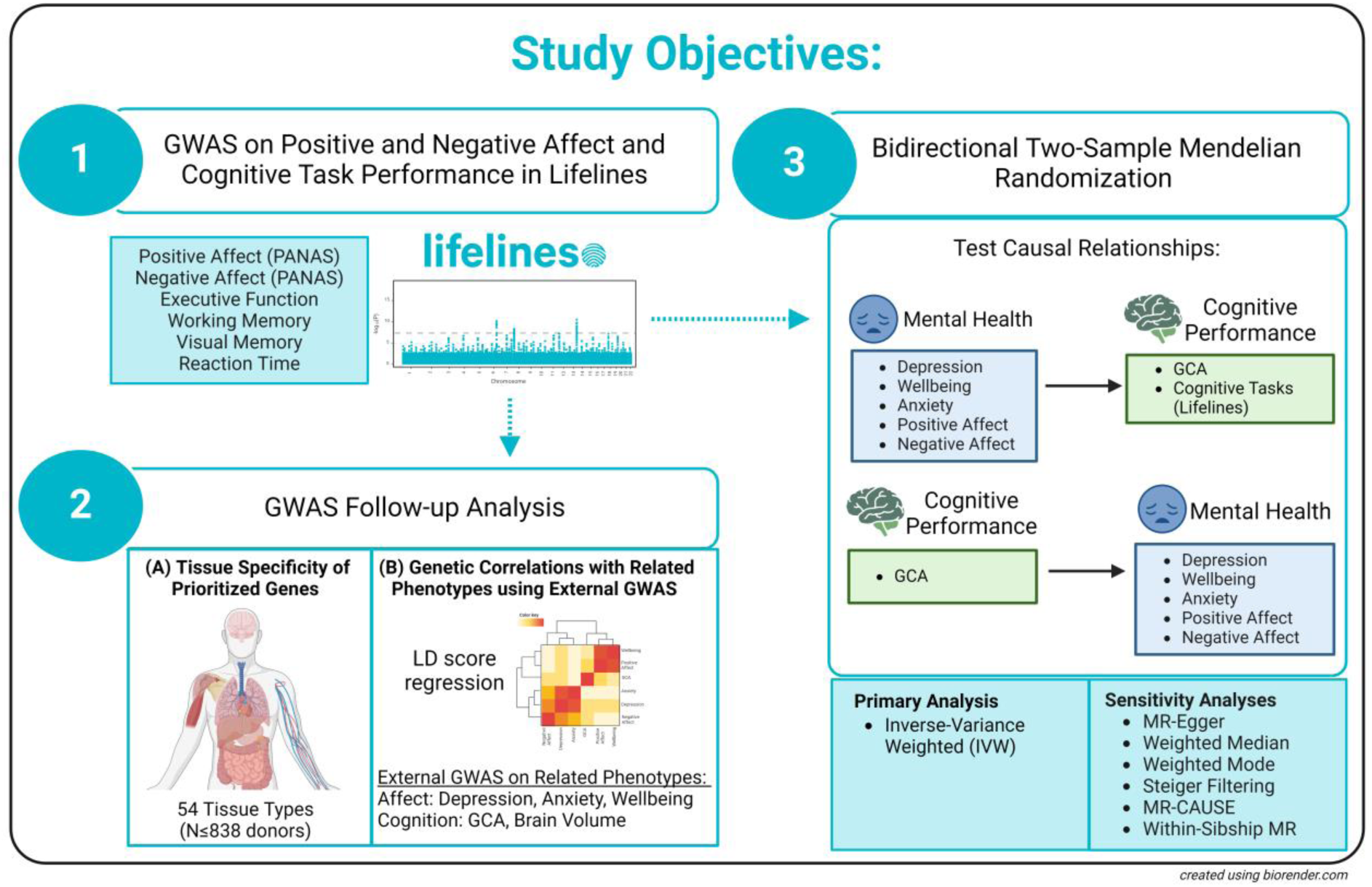
Study Objectives.

**Figure 2.**
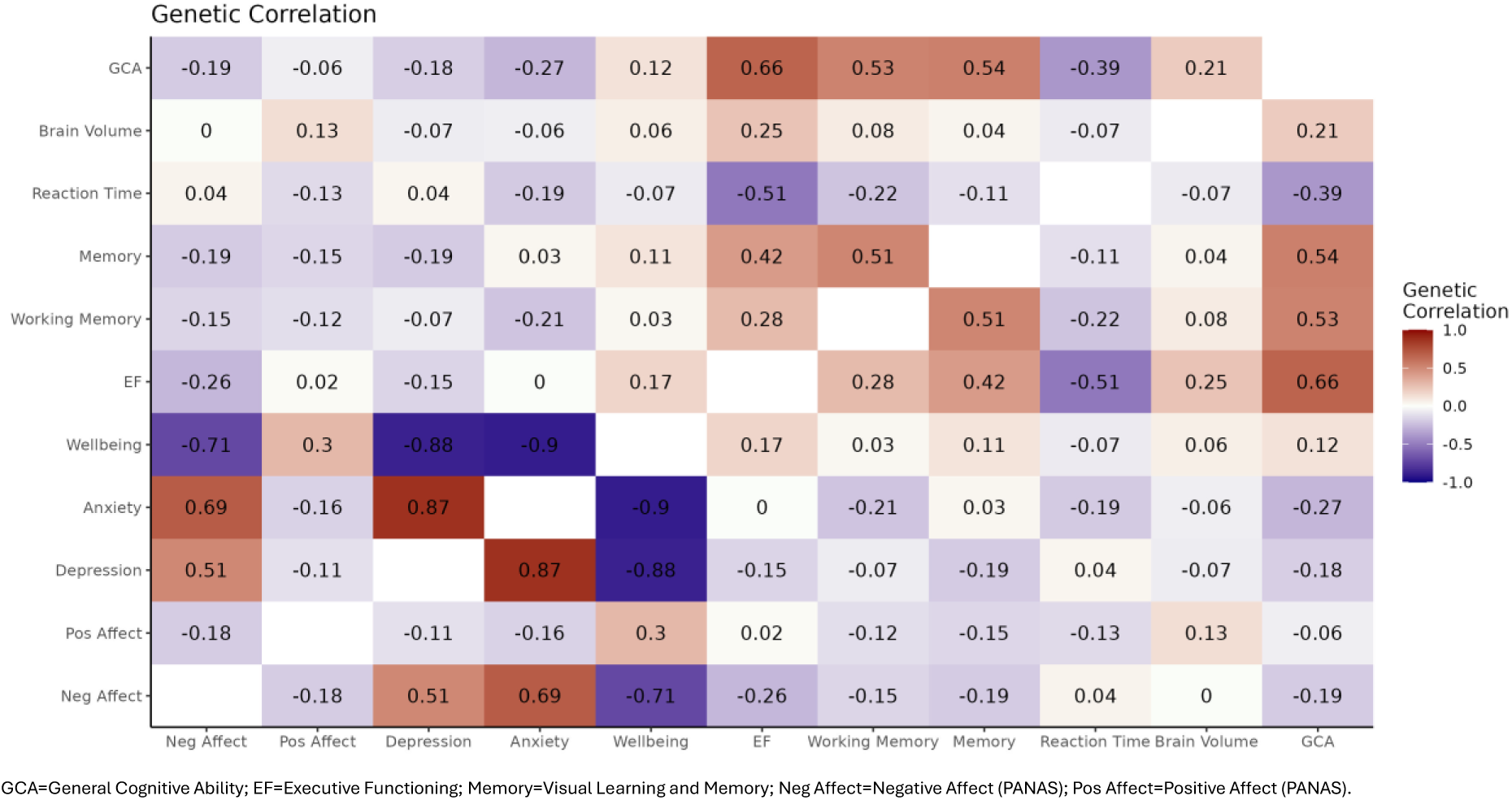
Genetic correlations between Lifelines phenotypes (negative affect, positive affect, cognitive performance) and external phenotypes (depression, anxiety, wellbeing, GCA, brain volume) using LDSR.

### 3.4. Potential Causal Effects: Results from Mendelian Randomization

#### 3.4.1. Effect of Mental Health Phenotypes on Cognition

In primary analyses, across all MR methods there was consistent evidence for a potential causal effect of depression on lower GCA (IVW estimate: −0.14 [95% CI= −0.19 to −0.09], *p*=0.00000006) and of one SD increase in wellbeing on higher GCA (IVW estimate: 0.30 [95% CI= 0.13 to 0.46], *p*=0.0004) (Figure 3). These effects remained after applying Steiger Filtering (Figure 4; Supplementary Tables S6-S9) and were supported by MR CAUSE (Supplementary Table S24-S25). There was also evidence for a potential causal effect of one SD increase in wellbeing on better executive functioning (IVW estimate: 0.24 [95% CI= 0.03 to 0.44], p=0.024), and weak evidence of an effect on memory (IVW estimate: 0.19 [95% CI= −0.02 to 0.41], p=0.076). However, these effects were not consistent across MR methods and were not supported following Steiger filtering (Figure 4; Supplementary Tables S6-S9).

**Figure 3.**
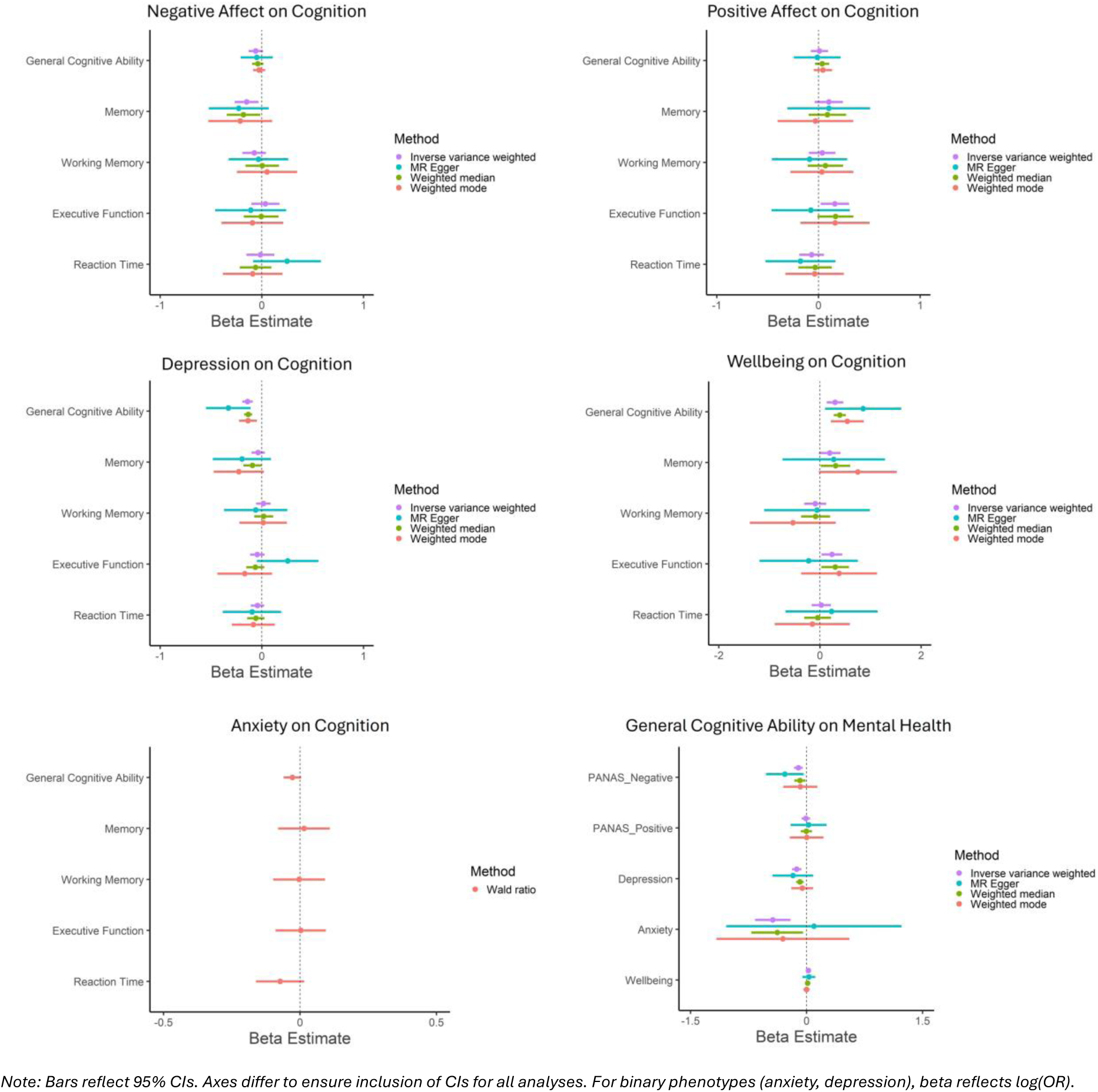
Mendelian Randomization Analyses Testing Evidence of Potential Causality between Mental Health and Cognition Phenotypes.

**Figure 4.**
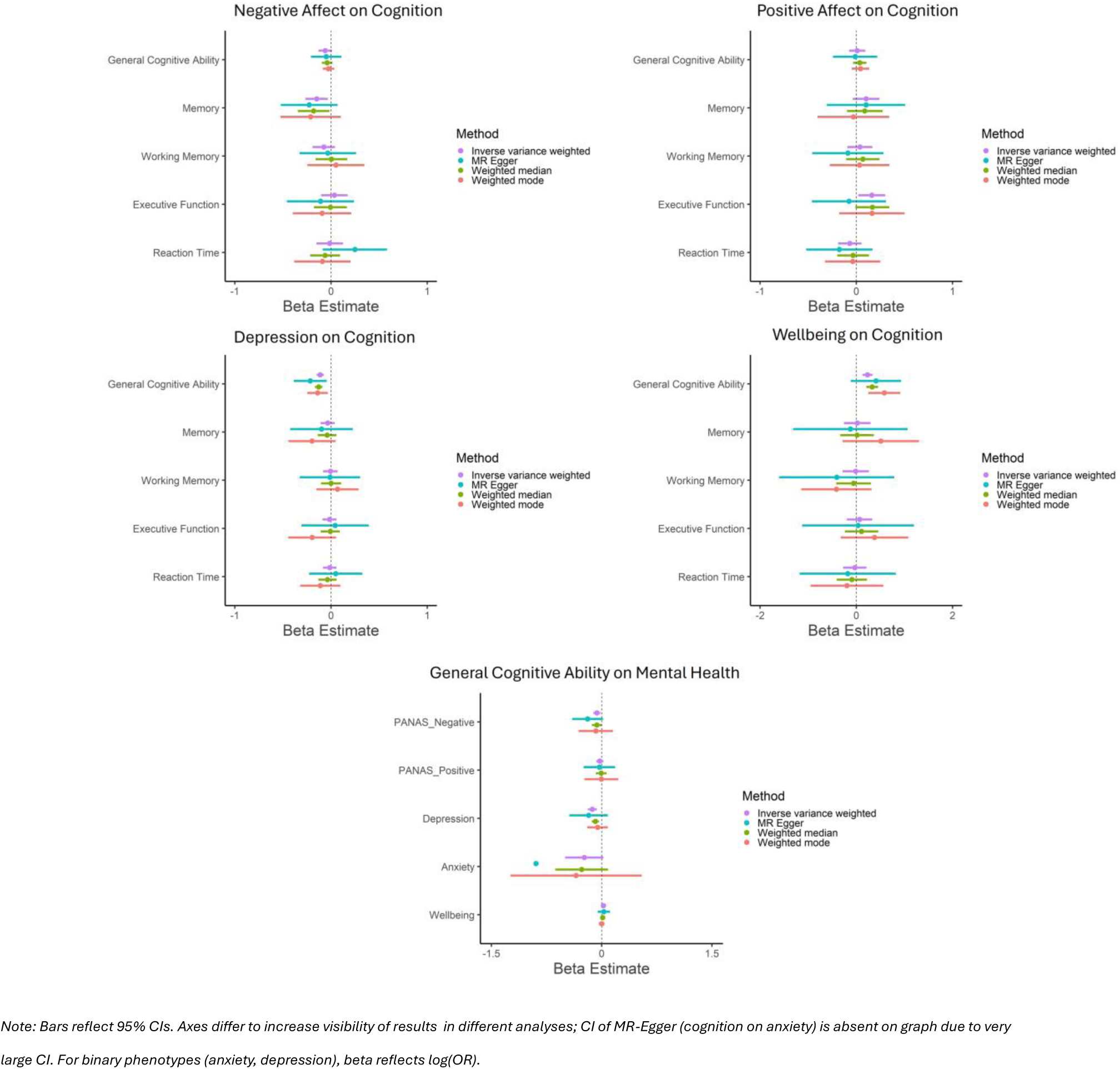
Steiger Filtered Mendelian Randomization Analyses Testing Evidence of Potential Causality between Mental Health and Cognition Phenotypes.

In secondary analyses, there was evidence for a potential causal effect of one SD increase in negative affect on poorer memory (IVW estimate: −0.15 [95% CI= −0.27 to −0.03], *p*=0.012) and one SD increase in positive affect on better executive functioning (IVW estimate: 0.16 [95% CI= 0.02 to 0.30], *p*= 0.024). However, these effects were not consistent across MR methods, see Figures 4 and Supplementary Tables S10-S13.

For other mental health and cognitive phenotypes, there was little evidence of causality based on IVW estimates (Figures 4; Supplementary Tables S6-S14). Confidence intervals (CIs) using within-sibship GWAS were very large with imprecise estimates (Figure S16).

There was evidence of heterogeneity, with the strongest evidence in analyses of depression and wellbeing on GCA, see Supplementary Table S15. There was little evidence of horizontal pleiotropy (based on Egger Intercept; *p*s≥0.14), except for weak evidence in MR analyses of depression on GCA (*p*=0.084) and executive function (*p*=0.048); see Supplementary Table S16.

#### 3.4.2. Effect of GCA on Mental Health Phenotypes

There was evidence for a potential causal effect of one SD increase in GCA on lower negative affect (IVW estimate: −0.11 [95% CI= −0.16 to −0.05], p=0.0002), reduced risk of depression (OR: 0.88 [95% CI=0.83 to 0.93], *p*=0.00003), and anxiety (OR: 0.64 [95% CI=0.51 to 0.81], *p*= 0.0002), and higher wellbeing (IVW estimate: 0.02 [95% CI= 0.01 to 0.04], *p*=0.011) (Figure 3; Supplementary Table S4). However, effects were not consistent across all MR methods: there was more consistent evidence for GCA on negative affect (3/4 methods provide evidence supporting causality) compared to depression (2/4), anxiety (2/4), and wellbeing (1/4).

Following Steiger Filtering, there was still evidence for a potential causal effect of one SD increase in GCA on lower negative affect (IVW estimate: −0.07 [95% CI= −0.11 to −0.02], *p*=0.007) but weaker evidence for one SD increase in GCA on reduced risk of anxiety (IVW estimate: −0.24 [95% CI= −0.50 to 0.02], *p*=0.072), although CIs were large (see Figure 4 and Supplementary Table S5). For GCA on depression and wellbeing, results were unaltered as no variants were removed in Steiger Filtering. MR CAUSE support that the data are consistent with a causal effect of GCA on negative affect, depression, and wellbeing (See Supplementary Tables S24-S25). Although MR CAUSE suggests the data for GCA on anxiety fit the causal model better than the null or sharing model, this did not meet conventional *p*-value criteria (*p*>0.05). In within-sibship MR, CI were very large with imprecise estimates (Figure S16).

There was strong evidence of heterogeneity in MR analyses testing effects of GCA on mental health phenotypes, except for GCA on anxiety (*p*=0.34); see Supplementary Table S15. There was little evidence of horizontal pleiotropy based on MR Egger Intercept (*p*s ≥ .15) (Supplementary Table S16).

## 4. Discussion

We conducted genome-wide association studies (GWAS) on negative and positive affect using a well-validated scale (PANAS) and four cognitive domains (executive function, working memory, visual learning and memory, reaction time) in the Lifelines Cohort. We identified one genome-wide hit (*p*<5×10^-8^) for reaction time, and many loci with suggestive associations (*p*<5×10^-6^) for other phenotypes. As predicted, gene mapping and tissue expression analysis of suggestive hits show higher gene expression in brain tissue compared to other tissues for most phenotypes; negative affect is genetically correlated with mental health phenotypes (depression *r*_g_=0.51; anxiety *r*_g_=0.70; wellbeing *r*_g_ = −0.71) and cognitive domains are genetically correlated with GCA and brain volume (*r*_g_ ≤ 0.66). Genetic correlations between negative and positive affect suggest that they are dissociable constructs (*r*_g_ = −0.18); with negative affect having higher genetic overlap with GCA than positive affect (*r*_g_ =-0.19 vs −0.06). Importantly, MR results suggest evidence of potential causal effects of higher GCA on reduced negative affect, reduced risk of depression and anxiety, and higher wellbeing (with the most robust result being higher GCA on reduced negative affect), but little impact of GCA on positive affect. We also report evidence for potential causal effects of depression and lower wellbeing on reduced GCA. Taken together, these results suggests that GCA may be a valid target for reducing negative affect, but not for increasing positive affect; and depression and wellbeing may be valid targets for GCA.

### 4.1. Potential Causality between Mental Health and Cognitive Phenotypes

Across different MR methods, we found evidence of a potential causal effect of higher GCA on many mental health outcomes (reduced negative affect, reduced risk of depression and anxiety, increased wellbeing). There was also evidence of potential causal effects of depression and wellbeing on GCA. Whilst we did not observe evidence for negative affect and anxiety on GCA, given the consistent direction of effect, this may be due to these GWAS being smaller and having decreased statistical power when used as exposures. Our findings complement recent studies using MR analyses and complementary designs (e.g., within-sibship analyses) reporting evidence of causality between mental health phenotypes (depression, wellbeing, and/or anxiety) and constructs related to cognition [e.g., educational attainment (Demange et al., 2024), education duration (Van De Weijer et al., 2024)]).

However, few studies have directly tested causality between mental health and cognition. Using MR analyses in the ALSPAC cohort, Suddell et al. (2023) tested causality between mental health conditions (depression, anxiety) and specific cognitive domains (response inhibition, working memory, emotion recognition) but reported that estimates were imprecise likely due to limited statistical power (Suddell et al., 2023). Marchi et al. (2024) used multivariable MR to test the causal effect of GCA and poverty on several mental health conditions (including depression and anxiety) (Marchi et al., 2024). After adjusting for poverty, they found evidence that higher GCA may causally reduce risk of depression and anxiety. Through inclusion of additional GWAS (negative affect, positive affect, larger depression GWAS), our findings lend support to these findings and additionally suggest: (1) higher GCA may also cause lower negative affect (but not influence positive affect), (2) evidence of bidirectional effects (i.e., higher risk of depression and lower wellbeing may also cause poorer GCA). Taken together, these MR findings suggest a potential bidirectional causal relationship between mental health and cognition.

Whilst our results could indicate a bidirectional causal effect, other explanations (which violate MR assumptions) may account for these results including: (1) correlated pleiotropy and (2) population-level confounding. Correlated pleiotropy occurs when genetic variants used in MR analyses affect both the exposure and outcome via a shared heritable factor (Morrison et al., 2020). An example of how this could occur here is through genetic variants impacting brain-related processes (e.g., synaptic plasticity) which directly affect both cognition and mental health. Whilst we did include several MR methods which allow for some correlated pleiotropy (e.g., weighted median, weighted mode, MR CAUSE) (Morrison et al., 2020), these methods could still give biased estimates (and lead to incorrect conclusions regarding causality) if the *majority* of instruments exhibit correlated pleiotropy. Another factor which may account for these results is population-level confounding (e.g., assortative mating, dynastic effects) (Brumpton et al., 2020). Whilst we are interested in direct effects from GWAS (i.e., genetic variants effect on phenotypic variation), for many phenotypes GWAS will also pick up indirect effects (e.g., dynastic effects: parental genotype affecting offspring phenotype via environmental factors). This is problematic as it violates MR assumptions and could lead to incorrect conclusions (Brumpton et al., 2020). One approach proposed to overcome this is to use within-family GWAS (Brumpton et al., 2020; Howe et al., 2022). Howe et al. (2022) found that GWAS estimates for some phenotypes, including cognitive ability and depressive symptoms, were attenuated when using within-sibship GWAS compared to population-level GWAS (Howe et al., 2022). To try to address this, we also conducted MR using within-sibship GWAS (depressive symptoms, wellbeing, and cognitive ability) to test whether this impacted our findings.

Unfortunately, confidence intervals (CIs) were very large with imprecise estimates (Supplementary Methods and Figure S16). This is unsurprising given the much smaller sample size of within-sibship GWAS compared to population-level GWAS and highlights the need for larger within-family GWAS on complex phenotypes like cognitive ability and depressive symptoms.

### 4.2. Additional Insights from Genetic Analyses on Negative and Positive Affect, and Cognitive Performance

Genetic correlation analyses reveal key insights into the genetic architecture of these phenotypes. First, as predicted, there was moderate genetic overlap between negative affect and mental health phenotypes (depression *r*_g_=0.51; anxiety *r*_g_=0.70; wellbeing *r*_g_ = −0.71), and between cognitive domains and cognition-related phenotypes (GCA, brain volume; *r*_g_ ≤ 0.66). This suggests that although these phenotypes are related, they are not interchangeable but rather have partly distinct genetic components. Second, as expected based on phenotypic correlations, genetic correlation between negative and positive affect suggest they are dissociable constructs as opposed to being opposite ends of the same spectrum (*r*_g_ = −0.18) (Watson et al., 1988). Third, compared to positive affect, negative affect has higher genetic overlap with GCA (*r*_g_ =-0.19 vs −0.06). This could indicate that negative affect has a higher shared neural basis with GCA than positive affect and/or GCA and negative affect may exhibit causal relationships (as suggested by our MR results).

Despite the negative affect GWAS having a much smaller sample size than the depression GWAS (N=57,946 versus N=1,035,760), when used as an outcome, we found more consistent evidence that GCA may play a causal role in negative affect across MR methods. Speculatively, this may be because the GWAS on negative affect consists of a more homogeneous phenotype which may increase statistical power and impact effect estimates (Manchia et al., 2013). This highlights the importance of future studies conducting GWAS on more homogeneous phenotypes (Nagel et al., 2018). Whilst we focused on transdiagnostic features of positive and negative affect and specific cognitive domains, there is a need for GWAS on other transdiagnostic features [e.g., sleep disturbances, anhedonia, hot cognition (Roiser & Sahakian, 2013)]. It is likely that advances will also be gained by parsing heterogeneity using other approaches. For example, GWAS on depressed patients with specific characteristics [e.g., immune-metabolic depression (Milaneschi et al., 2020)]. Research focusing on improving the validity of subtypes within and across psychiatric conditions will be necessary for advancing our understanding of these conditions (Hammen, 2018).

### 4.3. Limitations

Limitations of this study must be considered when interpreting the results. First, smaller sample sizes for some GWAS resulted in a lack of genome-wide significant variants (*p*<5×10^-8^) and/or larger CIs in MR analyses (positive and negative affect, specific cognitive domains, anxiety). Consortia combining data from several large datasets are necessary to provide well-powered GWAS on these phenotypes; our GWAS in the Lifelines Cohort will provide a useful contribution to this endeavour. Second, MR estimates lifetime effect of an exposure (e.g., depression) on an outcome (e.g., GCA) (Sanderson et al., 2022). Whilst this study is informative for understanding lifetime risk, it is unclear what time periods would be best to intervene on. This requires either a randomised controlled trial (RCT; which would be expensive and time consuming) or MR with large GWAS on exposures and outcomes at specific ages in the lifespan (Power et al., 2023). Third, cognitive performance is highly related to other socioeconomic phenotypes (e.g., education, socioeconomic status). Future studies testing independent effects and interactions between these phenotypes on mental health using other methods (e.g., multivariable MR) would be useful (see Marchi et al., 2024). Fourth, as discussed above, observed bidirectional causal relationships in MR analyses between mental health and GCA could instead be due to violation of MR assumptions. Triangulating results from standard MR analyses with other methods (e.g., within-sibship MR) may help to increase confidence in conclusions drawn. Fifth, many GWAS use data from large population-based cohorts which are less representative of some populations (e.g., less affluent people) which may hinder generalizability of the findings. Sixth, for continuous variables (e.g., GCA, negative affect, wellbeing), our study cannot shed light on whether these relationships are nonlinear. As currently available nonlinear MR approaches have provided implausible results (Wade et al., 2023), there is a need for other methods to be used to characterise the shape of these relationships (see Pines et al., 2024). Seventh, we focus on a subset of phenotypes, future studies should expand this to provide insight into other mental health and cognitive phenotypes which may show different relationships (e.g., schizophrenia, hot cognition) (Danhauer et al., 2013). Finally, many GWAS on psychiatric conditions do not exclude people with comorbidities. For example, the depression GWAS includes UK Biobank which defines depression based on the following question: “Have you ever seen a general practitioner (or psychiatrist) for nerves, anxiety, tension, or depression?”. This may result in many individuals with anxiety being characterised as having depression and makes it challenging to conduct subsequent analyses testing genetic overlap/causality between different conditions.

### 4.4. Implications

Our GWAS on positive and negative affect and cognitive domains in the Lifelines Cohort provide valuable resources which may facilitate insights into aetiology, comorbidity, and causal risk factors for these phenotypes. We found evidence of potential causal relationships between mental health phenotypes (negative affect, depression, anxiety, wellbeing) and GCA. This may suggest that strategies targeting poor mental health may prevent/treat cognitive dysfunction, and *vice versa*. However, to increase confidence in this finding, triangulation using other methods which have different strengths/limitations to MR, and consideration of highly related phenotypes (e.g., education), are needed. Additionally, GWAS on other transdiagnostic phenotypes are necessary to enable clearer insights into potential causal relationships. If multiple lines of evidence support causality, careful consideration of potential interventions (e.g., age to intervene, length of intervention, whether interventions targeting depression to reduce poorer GCA [or *vice versa*] could also have their own direct effect of GCA) would be necessary. This could have important implications for clinical practice (e.g., targeting depression may help prevent/treat cognitive impairments in health conditions such as dementia; cognitive remediation therapy may help prevent/treat depression). Considering the broader literature (Demange et al., 2024; Marchi et al., 2024), policy changes targeting factors impacting GCA [e.g., education (Anderson et al., 2020)] would potentially be promising for reducing future mental health challenges in the general population.

Nevertheless, there is also evidence suggesting that cognitive impairments can persist in remitted depressed individuals (Semkovska et al., 2019). This may appear to contrast the idea that treating depression may help to prevent/treat cognitive impairments. Speculatively, this could be because: (1) once depressive symptoms have decreased, cognitive impairments reduce but require a longer time to observe effects (RCTs may not have long enough follow-up lengths), (2) reducing depression may help to prevent cognitive impairments, but may not help treat them once they are already experienced (i.e., a ‘scar effect’), (3) treating depression may improve cognition in a subset of people (not all individuals), and/or (4) some symptoms of depression when treated may improve cognition (but not all symptoms when treated will improve cognition). There is a need for research testing these different theories to better understand the dynamic relationship between depression and cognition.

### 4.5. Conclusions

In summary, we conducted GWAS on transdiagnostic features of many health conditions (positive and negative affect, four specific cognitive domains). We identified one genome-wide hit (*p*<5×10^-8^) for reaction time, and many loci with suggestive associations (*p*<5×10^-6^) for other cognitive phenotypes. Follow-up gene mapping and tissue expression analyses of suggestive hits show higher gene expression in brain tissue compared to other tissues for most phenotypes. Genetic correlation analyses show that negative and positive affect are dissociable constructs, with negative affect having higher genetic overlap with GCA than positive affect. Importantly, in MR analyses, we found evidence of a potential causal effect of higher GCA on multiple mental health phenotypes (reduced negative affect, depression, and anxiety; and increased wellbeing), with little evidence on positive affect. We also report evidence of potential causal effects of depression and lower wellbeing on reduced GCA. Taken together, as the most robust evidence was for GCA on negative affect, with little effect on positive affect, this suggests that GCA may be a valid target for negative affect (but not positive affect) and depression and wellbeing may be valid targets for GCA. Further research testing the relationship between depression and cognition using complementary research designs is warranted, particularly there is a need for studies to test different theories we proposed in the discussion.

## Data Availability

For details on how to access publicly available GWAS summary statistics used in this paper, please see Supplementary Materials. New GWAS summary statistics conducted in the Lifelines Cohort and used in this paper will be made available upon publication. Conditions of data access to the Lifelines Cohort Study can be found at www.lifelines-biobank.com/researchers/working-with-us.

## Acknowledgements

The generation and management of GWAS genotype data for the Lifelines Cohort Study is supported by the UMCG Genetics Lifelines Initiative (UGLI). UGLI is partly supported by a Spinoza Grant from NWO, awarded to Cisca Wijmenga. The authors wish to acknowledge the services of the Lifelines Cohort Study, the contributing research centers delivering data to Lifelines, and all the study participants.

The Genotype-Tissue Expression (GTEx) Project was supported by the <u>Common Fund</u> of the Office of the Director of the National Institutes of Health, and by NCI, NHGRI, NHLBI, NIDA, NIMH, and NINDS. The final data used for the analyses described in this manuscript were obtained from: the GTEx Portal via FUMA on 01/07/2024 and 18/07/2024.

## Ethics approval

The general Lifelines protocol has been approved by the UMCG Medical ethical committee under number 2007/152.

## Contributions

CS, GK, CH, NMG and GH conceptualized and designed the study. CS analyzed the data and drafted the manuscript. All authors advised on the project/analysis and approved the final version of the manuscript.

