## Supplementary Materials for "Positive and negative affect, related mental health traits, and cognitive performance: shared genetic architecture and potential causality"

### Supplementary Material

Figure S16. Comparison of Mendelian randomization results using GWAS data used in this study, alongside population and within-sibship GWAS estimates from Howe et al., (2022). 145
REFERENCES........ 146

#### SUPPLEMENTARY LIFELINES GROUP AUTHOR INFORMATION

UMCG Genetics Lifelines Initiative (UGLI) group author: LifeLines Cohort Study

Raul Aguirre-Gamboa (1), Patrick Deelen (1), Lude Franke (1), Jan A Kuivenhoven (2), Esteban A Lopera Maya (1), Ilja M Nolte (3), Serena Sanna (1), Harold Snieder (3), Morris A Swertz (1), Peter M. Visscher (3,4), Judith M Vonk (3), Cisca Wijmenga (1), Naomi Wray (4).

(1) Department of Genetics, University of Groningen, University Medical Center Groningen, The Netherlands.

(2) Department of Pediatrics, University of Groningen, University Medical Center Groningen, The Netherlands.

(3) Department of Epidemiology, University of Groningen, University Medical Center Groningen, The Netherlands.

(4) Institute for Molecular Bioscience, The University of Queensland, Brisbane, Queensland, Australia.

#### SUPPLEMENTARY METHODS

***Lifelines Genotype Data*** *[Taken from (Giollabhui et al., 2024)]*

*CytoSNP*. In total, ~17,000 participants were genotyped in different batches using Illumina HumanCytoSNP-12v2.0 (n~16,500) and HumanCytoSNP-12v2.1 (n~500). Only probes present on both platforms were included. Genotyping was done using OptiCall and calls were refined using Beagle. CytoSNP originally used Genome Build 36, the probes were remapped to Genome Build 37 using SHRiMP2, with all probes are mapped on the forward strand. In total, 264,922 variants were present on both versions of the used HumanCytoSNP. Quality controls included excluding individuals with: (1) gender mismatches, (2) minimal or excessive heterozygosity, (3) duplicate sample identification, (4) missingness (call-rate < 95%), (5) non-Caucasian (determined by self-report in Lifelines phenotype database, Outlier [IBS] analysis, population stratification using Eigenstrat), (6) cryptic relationships (if a pair of samples were indicated as first-degree relatives using genetic similarity, the sample with the best genotyping quality was included). This resulted in 15,422 participants being available. Variants with minor allele frequency (MAF) of < 1%, call rate < 95%, or evidence for violations of Hardy-Weinberg Equilibrium ($p < 0.001$) were removed. Phasing was done using SHAPEIT2 and imputation using IMPUTE2 (combined reference panel of both genomes Genome of the Netherlands release 5 and 1000 Genomes phase1 v3 was used). For further details, please see: http://wiki-lifelines.web.rug.nl/doku.php?id=gwas.

*GSA [UGLI cohort]*. In total, 38,030 participants were genotyped in 31 batches using the Infinium Global Screening Array® (GSA) MultiEthnic Disease version 1.0. In total, 691,072 variants were genotyped, of which 571,420 markers met quality control steps. Quality controls included excluding individuals with: (1) gender mismatches, (2) heterozygosity (>4 standard deviations from mean), (3) duplicate samples, (4) missingness (two-step process: first removed individuals > 20% missingness and then > 1% missingness). Participants were of Caucasian-ancestry, although PCA analysis detected 35 participants who were non-European. Variants which were monomorphic (MAF = 0), call rate < 1%, HWE ($p \leq 1x10^{-6}$) were removed. A final set of 36,339 participants and 571,420 variants on autosomal and X chromosomes passed quality steps described above and were used for genetic imputation. Genetic imputation was done through the Sanger imputation service using the Haplotype Reference Consortium (http://www.haplotype-reference-consortium.org) panel. Following format instructions from the Sanger webpage (https://www.sanger.ac.uk/science/tools/sanger-imputation-service), 152 tri-allelic variants and 1608 insertions/deletions were removed. To facilitate researchers, Lifelines provide a list of non-European participants and a list of poorly imputed SNPs. For further details, please see: http://wiki-lifelines.web.rug.nl/lib/exe/fetch.php?media=qc_report_ugli_r1.pdf and http://wiki-lifelines.web.rug.nl/doku.php?id=ugli.

*Affymetrix [UGLI2 cohort]*. In total, 29,166 participants were genotyped in 12 batches using the FinnGen Thermo Fisher Axiom® Custom array. Genotyping was done in Human

Genome Build hg38. Quality controls included excluding individuals with: (1) sample mix-ups (using gender mismatches and pedigree concordance), (2) heterozygosity (> 4 SD from the mean), (3) duplicate sample, (4) missingness (two-step process: first removed individuals with > 20% missingness and then > 3% missingness). A genetic relationship matrix was created for the 1000G cohort (without the admixed AMR population samples) (https://www.internationalgenome.org/) and used for principle-component-analysis (PCA) of up to 20 principal components to generate PC-loadings that were projected onto the UGLI2 cohort. The PC analysis of all 1000G superpopulations identified 142 non-Europeans (>4 SDs from centroid 1000G European population for first five PCs), and PC analysis of only 1000G European population identified 161 genetic outliers (>4 SDs from centroid of all UGLI2 samples for first two PCs). In this study, we removed non-European and genetic outliers from the dataset. Variants with MAF < 0.02%, call rate < 1%, HWE in all samples ($p<1x10^{-10}$), HWE in unrelated samples ($p<1x10^{-6}$; defined as no 1st or 2nd degree relations) were excluded. There were no SNPs with > 1% Mendelian errors across all parent-offspring pairs. Prior to imputation, genetic markers were lifted over to Genome Build GRCh37 and aligned with Haplotype Reference Consortium (HRC) v1.1 (http://www.haplotype-referenceconsortium.org/site). A final set of 28,250 samples and 462,731 markers on autosomal and X chromosomes passing quality check steps above were used for genetic imputation using the Sanger imputation service using HRC panel. For further details, please see: http://wiki-lifelines.web.rug.nl/lib/exe/fetch.php?media=qc_report_ugli2_release_1_-v1.pdf and http://wiki-lifelines.web.rug.nl/doku.php?id=ugli.

*Principal Components.* Lifelines provides genetic PCs for each chip separately.

##### *Lifelines Cognitive Measures Included in this Study*

*Cogstate One Card Learning Task ['Visual Learning & Memory'].* This task is designed to measure visual learning and memory (Kuiper et al., 2017). In this task, participants attend to a card in the centre of the screen and respond to the question "Have you seen this card before in this task?" with "Yes" or "No". The task ends after 42 trials. The primary outcome is proportion of correct answers normalized using arcsine transformation.

*Cogstate Identification Task ['Reaction Time].* This task is designed to measure reaction time/visual attention (Kuiper et al., 2017). Participants attend to a card in the centre of the screen and respond to the question "Is the card red?" with "Yes" or "No". The task ends after 30 correct trials. The primary outcome is reaction time (ms) normalized using log10 transformation.

*Cogstate One-back Task ['Working Memory'].* This task is designed to measure of attention and working memory (Kuiper et al., 2017). In this task, participants attend to a card in the centre of the screen and respond to the question "Is this card the same as that on the immediately previous trial?" with "Yes" or "No". The task ends after 30 correct trials. The primary outcome is proportion of correct answers, normalized using arcsine transformation.

Data cleaning involved excluding participants with implausibly low accuracy rates which indicate poor effort, failure to comprehend task instructions, or technical errors. Specifically, accuracy rates <25% on the one card learning task (N=231), <40% on the identification task (N=2,878), and <35% on the one back task (N=1,330).

*Ruff Figural Fluency Task [‘Executive Function’].* The RFFT provides a valid and reliable measure of executive functioning (Ross, 2014). The task consists of five parts, each containing 35 identical five-dot patterns. Participants draw as many unique designs as possible within one minute by connecting dots in different patterns (Kuiper et al., 2017). The primary outcome is total number of unique designs. All Lifelines participants completed the RFFT until April 2012 (when it was subsequently administered to a random 50% of the sample). We removed participants who did not generate one unique design per trial and were deemed invalid (N=181).

##### *Details of Publicly Available GWAS*

**General Cognitive Ability** (Lam et al., 2021). Meta-analysis of two GWAS:

> Davies et al. (2018), a GCA score was derived from two consortia (COGENT and CHARGE) and UK Biobank. For each consortia cohort, a GCA score was constructed from cognitive tasks (required ≥ 3 domains) using principal component analysis (PCA). In UK Biobank, score on the verbal-numerical reasoning test (13-item multiple-choice question) was used. Includes four samples: baseline (N=107,586), second assessment (N=11,123), third MRI assessment (N=3,002), fourth (web-based) (N=46,322). Exclusions were clinical stroke (including self-reported stroke) or prevalent dementia.
>
> Savage et al. (2018), a GCA score was derived from each cohort (except High IQ/Health and Retirement Study where a logistic regression was run predicting whether participants were drawn from a population of very high intelligence). Cohorts had either a single sum score, mean score, or factor score from a battery of cognitive tests (for example, IQ score, fluid intelligence test, and cognitive tasks such as digit span/processing speed. Different cohorts applied different exclusion criteria, see papers supplementary for details.

**Depression** (Als et al., 2023). This GWAS was a meta-analysis of six datasets (summary statistics used in this study exclude 23andMe). The study combined iPSYCH2015 and FinnGen with publicly available GWAS summary statistics from (Howard et al., 2019) and (Wray et al., 2018) which included the following datasets: Million Veteran Program (MVP), 23andMe, UK Biobank, Psychiatric Genetics Consortium (PGC). For details on sample sizes, definitions of depression and controls, proportions of males/females and additional exclusion criteria of individual datasets, please see original papers. Exclusions included individuals with a diagnosis of bipolar disorder. Briefly, depression diagnosis included both meeting diagnostic criteria (e.g., ICD/DSM) and broader definition using self-report questions.

> iPSYCH2015 is a nationwide population sample including all children who were (i) born in Denmark between 1st May 1981 and 31st December 2008, (ii) lived in Denmark on their 1st birthday and (ii) have a known mother. Diagnosis of depression was determined using ICD-10 (F32-F33 codes) and were reported from psychiatric hospitals and outpatient clinics. FinnGen includes Finland health register data, and diagnosis of depression was determined using ICD-10/ICD-9 (F32-F33 codes).
>
> In Howard et al., (2019) depression cases were determined using a broad definition. Briefly, this was based on response to the questions 'Have you ever seen a general practitioner for nerves, anxiety, tension, or depression?' or 'Have you ever seen a psychiatrist for nerves, anxiety, tension or depression?'. Exclusions included people with bipolar disorder, schizophrenia, or personality disorder using self-report, or prescription of antipsychotic medication.
>
> In Wray et al. (2018), depression cases were determined as either meeting diagnostic criteria (DSM-IV, ICD-9, ICD-10) for lifetime diagnosis of MDD assessed by interviewers, clinician-administered checklists, or medical record review within the PGC dataset. Additional cohorts used a range of methods for assessing MDD or major depression and applied their own exclusion/inclusion criteria; see Wray et al. (2018) for details.

**Anxiety Disorders** (Otowa et al., 2016). Meta-analysis of results from seven cohorts. Includes five core anxiety disorders: Generalised Anxiety Disorder (GAD), Panic Disorder (PD), social phobia, agoraphobia, and specific phobias. Conducted GWAS with anxiety phenotype as (i) case-controls, or (ii) quantitative factor score using confirmatory factor analysis, separately. In this study, we used the case-control GWAS summary statistics. Anxiety was determined using standardized assessment instruments to generate DSM-based anxiety disorder diagnosis, with some exceptions. DSM-based anxiety disorder diagnostic assessment was available for all cohorts except Rotterdam Study, in which only one-year prevalence was assessed. In case-control comparison, anxiety cases met criteria for lifetime anxiety disorder; and controls had few or no clinical anxiety symptoms.

**Wellbeing Spectrum** (Baselmans et al., 2019). Multivariate GWAS meta-analysis including life satisfaction, positive affect, neuroticism, and depressive symptoms. Leveraged univariate GWAS meta-analyses of life satisfaction (N=80,852; 2 studies), positive affect (N=410,603; 3 studies), neuroticism (N=582,989; 6 studies), and depressive symptoms (N=1,295,946; 10 studies). For additional details, please see original paper.

**Brain Volume** (Jansen et al., 2020). GWAS meta-analysis of brain volume from UKB (N=17,062) from structural MRI (total grey and white matter volume, ventricular cerebrospinal fluid volume). GWAS was corrected for Townsend Deprivation Index (TDI), age, sex, genotype array, assessment centre, standing height, and top 10 genetic PCs. UKB GWAS meta-analysed with GWAS from two studies: Intracranial volume from ENIGMA consortium (N=11,373), head circumference (proxy for brain volume) from a meta-analysis of adults and children (N=18,881). Total sample size of 47,316 unrelated European.

##### *Functional Mapping and Annotation (FUMA) of Genome-Wide Association Studies*

For each GWAS we conducted in Lifelines, we used FUMA v1.5.2 to identify SNPs (significant independent and candidate), map the SNPs to genes ('prioritised genes'), and examine tissue specific expression patterns of the prioritised genes. For all analyses, we used default criteria in FUMA unless stated otherwise. First, to identify SNPs, we restricted to SNPs with a minor allele frequency (MAF) >=0.01 and used a lenient p-value threshold ($p<5x10^{-6}$). Second, to map SNPs to genes (providing a list of prioritised genes), our primary analyses included only positional mapping (maps SNPs to genes based on physical proximity). As a secondary analysis, we re-ran the analysis including three mapping methods: (1) positional mapping, (2) eQTL mapping (maps SNPs to genes based on eQTL information), (3) chromatin interaction mapping (maps SNPs to genes based on chromatin interactions). This was an exploratory analysis done to test whether including these additional resources (eQTL in brain regions and chromatin interaction in brain regions) impacted subsequent tissue specific expression patterns results. For a list of data sources used in each of these mapping methods, please see below and https://fuma.ctglab.nl/tutorial#eQTLs. Third, we examined tissue specific expression patterns using GTEx.

<u>When using eQTL mapping, we restricted to the following data sources:</u>
(i) eQTL catalogue (BrainSeq brain), (ii) PsychENCODE (Wang et al., 2018) (1387 individuals), (iii) BRAINEAC (Ramasamy et al., 2014) (134 post-mortem brains from individuals of European descent free of known neurological disorders) (includes cerebellar cortex, frontal cortex, hippocampus, inferior olivary nucleus, occipital cortex, putamen, substantia nigra, temporal cortex, thalamus, intralobular white matter, averaged expression of 10 brain regions), (iv) GTEx v6, v7, and v8 Brain Tissue (Aguet et al., 2017, 2020; Ardlie et al., 2015) (includes amygdala, anterior cingulate cortex BA24, caudate basal ganglia, cerebellar hemisphere, cerebellum, cortex, frontal cortex BA9, hippocampus, hypothalamus, nucleus accumbens basal ganglia, putamen basal ganglia, spinal cord cervical c-1, substantia nigra).

<u>When using 3D Chromatin Interaction mapping, we restricted to the following data sources:</u> (i) PsychENCODE EP links (one way and promoter anchored loops), (ii) HiC: adult and fetal cortex (Giusti-Rodríguez et al., 2019) (dorsolateral prefrontal cortex and hippocampus (GSE87112).

For each expression dataset, FUMA has pre-calculated differentially expressed gene sets (DEG). DEG sets are based on a t-test (2-sided) of a given tissue versus all other tissue types (direction is considered: up-regulated vs down-regulated) (Watanabe et al., 2017). DEG are genes which meet the following criteria: (1) Bonferroni-corrected $p<0.05$ and (2) Absolute log fold change ≥0.58 (Watanabe et al., 2017). The -log10(P) in the graph (Figure 2) reflects probability of the hypergeometric test (Watanabe et al., 2017).

##### *MR Assumptions*

MR is an epidemiological method used to assess potential causality (Davey Smith & Ebrahim, 2003). Three core assumptions of MR include: (i) genetic variant(s) are robustly associated with the exposure, (ii) there is no confounding of the genetic variants(s) and the outcome, (iii) the genetic variant(s) are independent of the outcome given the exposure (Sanderson et al., 2022). The validity of the causal inferences drawn relies on these assumptions being met. An additional assumption for two-sample MR used here (exception: secondary analyses using PANAS as exposure on cognitive task performance in Lifelines as outcome) is that samples come from the same underlying population but are non-overlapping (Lawlor, 2016). However, recent work suggests that sample overlap may not bias MR results as much as previously thought (Burgess et al., 2016; Sanderson et al., 2022).

##### *Summary-level MR Methods*

**Inverse Variance Weighted (IVW).** In meta-analyses, the IVW method is often used to combine results from individual studies to estimate an average effect, in which studies are weighted by the inverse of their variance (Burgess et al., 2013, 2020). In MR, IVW is used to combine individual SNP effects (Wald ratios – SNP outcome divided by SNP exposure association). In MR, this method assumes there is no horizontal pleiotropy (assumes all SNPs are associated with the outcome only via the exposure) and forces the intercept through zero. The IVW method will provide a consistent estimate if all SNPs are valid instrumental variables (Burgess et al., 2013, 2020).

**MR-Egger.** Unlike IVW method, MR-Egger does not force the intercept through zero. Thus, this method provide an estimate in the presence of invalid SNPs (Bowden et al., 2015). The slope provides a causal effect estimate and the intercept can be used to indicate the degree of horizontal pleiotropy.

**Weighted Median.** Uses the median of the ratio estimates and provides a consistent estimate if ≥ 50% of the weights come from valid SNPs (Bowden et al., 2016).

**Weighted Mode.** Provides a consistent estimate if the most common causal effect estimates come from valid SNPs (i.e., even if most SNPs are not valid) (Hartwig et al., 2017).

##### *MR CAUSE*

Causal Analysis using Summary Effect Estimates (CAUSE) is an MR method which accounts for correlated and uncorrelated horizontal pleiotropic effects (Morrison et al., 2020). Correlated pleiotropy occurs when genetic variants impact a shared heritable factor which affects both the exposure and outcome. Correlated pleiotropy can lead to false positives if not accounted for (Morrison et al., 2020). In this study, it is possible that genetic variants from GWAS on cognitive (e.g., general cognitive ability) and mental health phenotypes (e.g., negative affect, depression) may impact a shared heritable factor (e.g. brain-related processes such as synaptic plasticity, synaptic pruning) which affects both the cognitive and mental health phenotypes, and therefore it is important to further test this using MR CAUSE. MR-CAUSE is based on the idea that a shared heritable factor will induce correlations between the exposure and outcome beta in a *subset* of variants; whereas a causal effect will lead to correlations between exposure and outcome beta for *all* variants that have nonzero effect on the exposure. Based on this, MR CAUSE tests whether the GWAS summary statistics for both the exposure and outcome (using all genome-wide variants) are consistent with a causal effect by using a Bayesian model comparison approach (expected log pointwise posterior density; ELPD) to compare different models: (1) causal model and (2) sharing model (Morrison et al., 2020). Advantages of MR CAUSE include: (i) reduces false positive rate in the presence of correlated pleiotropy compared to many other MR methods, (ii) increases power by using genome-wide variants (as opposed to restricting to variants meeting a stringent p-value threshold on the exposure) (Morrison et al., 2020).

##### *Bidirectional Mendelian Randomization using Within-Sibship Data*

GWAS population-level estimates capture both direct effects, indirect effects (e.g., dynastic), and demographic effects (e.g., assortative mating) (Howe et al., 2022). Some phenotypes will be more influenced by indirect and demographic effects. This is problematic for Mendelian randomization (MR) analyses which assume no population-level confounding and could bias MR results. As family-based GWAS can control for indirect and demographic effects, comparing effects from population-level GWAS with effects from within-family GWAS may be useful (Howe et al., 2022).

Howe et al. (2022) conducted both population-level (between-family) and within-sibship (within-family) GWAS on 25 phenotypes (including some phenotypes used in this study) from 178,086 siblings. In this study, we re-ran the analysis using within-sibship GWAS (and population GWAS) reported in (Howe et al., 2022) for comparison. We used GWAS on: (1) depressive symptoms (population estimate N=47,517 [ieu-b-4840]; within-sibship estimate N=16,782 [ieu-b-4839]), (2) cognitive function (population estimate N=22,593 [ieu-b-4838]; within-sibship estimate N=9,997 [ieu-b-4837) and (3) wellbeing (population estimate N=63,392 [ieu-b-4852]; within-sibship estimate N=22,656 [ieu-b-4851]). As these GWAS are much smaller compared to the population-level analyses used in this study, we aimed to examine consistency in direction of effects as opposed to replicating results of this study.

Analyses were run using the MR-Base platform (Elsworth et al., 2020; Hemani et al., 2018) . Genetic variants were identified using the following criteria: (1) p-value <5e-06, (2) independent ($r^2$=0.01, kb=1000). Proxy SNPs ($R^2 \geq 0.8$) were identified for exposure SNPs not available in the outcome GWAS.

#### SUPPLEMENTARY TABLES

##### *Table S1. Publicly available GWAS used in genetic correlation and MR analyses.*

| Phenotype | Paper | Ancestry | Cohort/ studies(s) | Age & Sex (% Female) | N GWAS | Access |
|---|---|---|---|---|---|---|
| **General Cognitive Ability** | (Lam et al., 2021) | European | Meta-analysis of two GWAS: Davies et al. (2018) and Savage et al. (2018) | Age Range (5-102 years) | 373,617 | Contacted Author |
| **Depression** | (Als et al., 2023) | European | Meta-analysis of six datasets. Data included here excludes 23andMe (cases and controls) | Mean Age (22-67 years)<br><br>Cohorts:<br>iPSYCH2015 (57%)<br>FinnGen (56%)<br>MVP (7%)<br>UKB (54%)<br>PGC2 (Not stated) | Cases=294,322 Controls=741,438 | https://ipsych.dk/en/research/downloads/ |
| **Anxiety** | (Otowa et al., 2016) | European | Meta-analysis of seven cohorts: MGS controls PsyCoLaus, RS, SHIP, QIMR, TRAILS, NESDA/NTR. | Mean Age (18.7-66.5 years) | Cases=7016 Controls=14,745 | https://pgc.unc.edu/for-researchers/download-results/ |
| **Wellbeing** | (Baselmans et al., 2019) | European | Meta-analysis of life satisfaction, positive affect, depressive symptoms, neuroticism GWAS.<br><br>Cohorts/Consortia: SSGAC, US, UKB, 23andMe, CHARGE. | Not stated. | 2,370,390 | https://surfdrive.surf.nl/files/index.php/s/Ow1qCDpFT421ZOO |
| **Brain Volume** | (Jansen et al., 2020) | European | Meta-analysis in UKB, ENIGMA consortium, 11 population-based cohorts. | Not stated. | 47,316 | https://cncr.nl/research/summary_statistics/ |

Abbreviations: MGS=molecular genetics of schizophrenia; NESDA/NTR=The Netherlands study of depression and anxiety/Netherlands twin registry; QIMR=Queensland institute of medical research; RS=Rotterdam study; SHIP=study of health in Pomerania; TRAILS=tracking adolescents individual lives survey; SSGAC=Social Science Genetic Association Consortium; US=Understanding Society; UKB=UK Biobank; CHARGE=Cohorts for Heart and Aging Research in Genomic Epidemiology Consortium. All GWAS on Build GRCh37.

#### SUPPLEMENTARY RESULTS

***Table S2. Instruments used in MR analyses.***

| Phenotype | Author | Max N | Outcome | Hits (p<5e-08) | Independent (r2=0.01, kb=1000) |
|---|---|---|---|---|---|
| Depression | (Als et al., 2023) | Cases: 294,322 Controls: 741,438 | OR converted to Log(OR) | 11,465 | 165 |
| Wellbeing | (Baselmans et al., 2019) | 2,311,184 | Continuous | 13,345 | 161 |
| GCA | (Lam et al., 2021) | 373,617 | Continuous | 16,696 | 250 |
| Anxiety | (Otowa et al., 2016) | 17,310 | Log(OR) | 7 | 1 |
| Phenotype | Author | Max N | Outcome | Hits (p<5e-06) | Independent (r2=0.01, kb=1000) |
| PANAS Negative | Current study | 57,946 | Continuous | 73 | 20 |
| PANAS Positive | Current study | 57,946 | Continuous | 67 | 15 |

*Max N=Maximum sample size based on GWAS summary statistics (except for GCA which did not contain a sample size column so sample size was taken from paper); OR=Odds Ratio.*

***Table S3. Genetic Correlation Results.***

| Phenotype 1 | Phenotype 2 | rg | se | z | p | h2_obs | h2_obs_se | h2_int | h2_int_se |
|---|---|---|---|---|---|---|---|---|---|
| PANAS-Neg | PANAS-Pos | -0.184 | 0.076 | -2.419 | 1.56E-02 | 0.071 | 0.009 | 0.986 | 0.007 |
| PANAS-Neg | Exec Func | -0.263 | 0.102 | -2.588 | 9.65E-03 | 0.082 | 0.014 | 0.996 | 0.007 |
| PANAS-Neg | WM | -0.151 | 0.113 | -1.335 | 1.82E-01 | 0.066 | 0.012 | 0.986 | 0.007 |
| PANAS-Neg | Memory | -0.189 | 0.087 | -2.168 | 3.02E-02 | 0.108 | 0.014 | 0.990 | 0.007 |
| PANAS-Neg | RT | 0.045 | 0.102 | 0.439 | 6.60E-01 | 0.079 | 0.014 | 0.991 | 0.006 |
| PANAS-Neg | GCA | -0.189 | 0.042 | -4.477 | 7.56E-06 | 0.146 | 0.005 | 1.034 | 0.015 |
| PANAS-Neg | Depression | 0.506 | 0.046 | 11.028 | 2.80E-28 | 0.045 | 0.002 | 1.018 | 0.013 |
| PANAS-Neg | Anxiety | 0.695 | 0.183 | 3.787 | 1.52E-04 | 0.061 | 0.022 | 1.001 | 0.007 |
| PANAS-Neg | Wellbeing | -0.712 | 0.053 | -13.373 | 8.65E-41 | 0.022 | 0.001 | 1.006 | 0.017 |
| PANAS-Neg | Brain Vol | -0.005 | 0.066 | -0.069 | 9.45E-01 | 0.217 | 0.018 | 1.020 | 0.009 |
| PANAS-Pos | PANAS-Neg | -0.184 | 0.076 | -2.419 | 1.56E-02 | 0.080 | 0.008 | 0.992 | 0.007 |
| PANAS-Pos | Exec Func | 0.016 | 0.105 | 0.152 | 8.79E-01 | 0.082 | 0.014 | 0.996 | 0.007 |
| PANAS-Pos | WM | -0.118 | 0.107 | -1.111 | 2.67E-01 | 0.066 | 0.012 | 0.986 | 0.007 |
| PANAS-Pos | Memory | -0.146 | 0.083 | -1.752 | 7.99E-02 | 0.108 | 0.014 | 0.990 | 0.007 |
| PANAS-Pos | RT | -0.132 | 0.112 | -1.179 | 2.39E-01 | 0.079 | 0.014 | 0.991 | 0.006 |
| PANAS-Pos | GCA | -0.057 | 0.042 | -1.353 | 1.76E-01 | 0.146 | 0.005 | 1.034 | 0.015 |
| PANAS-Pos | Depression | -0.111 | 0.040 | -2.807 | 5.01E-03 | 0.045 | 0.002 | 1.018 | 0.013 |
| PANAS-Pos | Anxiety | -0.163 | 0.168 | -0.967 | 3.33E-01 | 0.061 | 0.022 | 1.001 | 0.007 |
| PANAS-Pos | Wellbeing | 0.297 | 0.050 | 5.970 | 2.38E-09 | 0.022 | 0.001 | 1.006 | 0.017 |
| PANAS-Pos | Brain Vol | 0.134 | 0.064 | 2.100 | 3.57E-02 | 0.217 | 0.018 | 1.020 | 0.009 |
| Exec Func | PANAS-Pos | 0.016 | 0.105 | 0.152 | 8.79E-01 | 0.071 | 0.009 | 0.986 | 0.007 |
| Exec Func | PANAS-Neg | -0.263 | 0.102 | -2.588 | 9.65E-03 | 0.080 | 0.008 | 0.992 | 0.007 |
| Exec Func | WM | 0.283 | 0.146 | 1.938 | 5.26E-02 | 0.067 | 0.012 | 0.986 | 0.007 |
| Exec Func | Memory | 0.421 | 0.117 | 3.616 | 2.99E-04 | 0.108 | 0.014 | 0.990 | 0.007 |
| Exec Func | RT | -0.513 | 0.116 | -4.424 | 9.71E-06 | 0.079 | 0.014 | 0.991 | 0.006 |
| Exec Func | GCA | 0.663 | 0.066 | 10.009 | 1.39E-23 | 0.146 | 0.005 | 1.034 | 0.015 |
| Exec Func | Depression | -0.154 | 0.046 | -3.328 | 8.75E-04 | 0.045 | 0.002 | 1.018 | 0.013 |
| Exec Func | Anxiety | -0.003 | 0.172 | -0.020 | 9.84E-01 | 0.061 | 0.022 | 1.001 | 0.007 |

| | | | | | | | | | |
|---|---|---|---|---|---|---|---|---|---|
| Exec Func | Wellbeing | 0.172 | 0.047 | 3.689 | 2.26E-04 | 0.022 | 0.001 | 1.006 | 0.017 |
| Exec Func | Brain Vol | 0.247 | 0.070 | 3.503 | 4.59E-04 | 0.217 | 0.018 | 1.020 | 0.009 |
| WM | Exec Func | 0.283 | 0.146 | 1.938 | 5.26E-02 | 0.083 | 0.014 | 0.996 | 0.007 |
| WM | PANAS-Pos | -0.118 | 0.107 | -1.111 | 2.67E-01 | 0.071 | 0.009 | 0.986 | 0.007 |
| WM | PANAS-Neg | -0.151 | 0.113 | -1.335 | 1.82E-01 | 0.080 | 0.009 | 0.992 | 0.007 |
| WM | Memory | 0.511 | 0.110 | 4.642 | 3.45E-06 | 0.108 | 0.014 | 0.990 | 0.007 |
| WM | RT | -0.216 | 0.126 | -1.712 | 8.68E-02 | 0.080 | 0.014 | 0.990 | 0.006 |
| WM | GCA | 0.527 | 0.060 | 8.812 | 1.23E-18 | 0.146 | 0.005 | 1.034 | 0.015 |
| WM | Depression | -0.068 | 0.054 | -1.245 | 2.13E-01 | 0.045 | 0.002 | 1.018 | 0.013 |
| WM | Anxiety | -0.213 | 0.217 | -0.981 | 3.27E-01 | 0.061 | 0.022 | 1.001 | 0.007 |
| WM | Wellbeing | 0.029 | 0.065 | 0.444 | 6.57E-01 | 0.022 | 0.001 | 1.006 | 0.017 |
| WM | Brain Vol | 0.078 | 0.082 | 0.952 | 3.41E-01 | 0.217 | 0.018 | 1.020 | 0.009 |
| Memory | WM | 0.511 | 0.110 | 4.642 | 3.45E-06 | 0.066 | 0.012 | 0.986 | 0.007 |
| Memory | Exec Func | 0.421 | 0.117 | 3.616 | 2.99E-04 | 0.083 | 0.014 | 0.996 | 0.007 |
| Memory | PANAS-Pos | -0.146 | 0.083 | -1.752 | 7.99E-02 | 0.071 | 0.009 | 0.986 | 0.007 |
| Memory | PANAS-Neg | -0.189 | 0.087 | -2.168 | 3.02E-02 | 0.080 | 0.009 | 0.992 | 0.007 |
| Memory | RT | -0.112 | 0.106 | -1.059 | 2.90E-01 | 0.079 | 0.014 | 0.991 | 0.006 |
| Memory | GCA | 0.537 | 0.048 | 11.153 | 6.90E-29 | 0.146 | 0.005 | 1.034 | 0.015 |
| Memory | Depression | -0.187 | 0.043 | -4.395 | 1.11E-05 | 0.045 | 0.002 | 1.018 | 0.013 |
| Memory | Anxiety | 0.033 | 0.166 | 0.199 | 8.42E-01 | 0.061 | 0.022 | 1.001 | 0.007 |
| Memory | Wellbeing | 0.108 | 0.039 | 2.814 | 4.90E-03 | 0.022 | 0.001 | 1.006 | 0.017 |
| Memory | Brain Vol | 0.040 | 0.061 | 0.667 | 5.05E-01 | 0.217 | 0.018 | 1.020 | 0.009 |
| RT | Memory | -0.112 | 0.106 | -1.059 | 2.90E-01 | 0.108 | 0.014 | 0.989 | 0.007 |
| RT | WM | -0.216 | 0.126 | -1.712 | 8.68E-02 | 0.066 | 0.012 | 0.986 | 0.007 |
| RT | Exec Func | -0.513 | 0.116 | -4.424 | 9.71E-06 | 0.083 | 0.014 | 0.996 | 0.007 |
| RT | PANAS-Pos | -0.132 | 0.112 | -1.179 | 2.39E-01 | 0.071 | 0.009 | 0.986 | 0.007 |
| RT | PANAS-Neg | 0.045 | 0.102 | 0.439 | 6.60E-01 | 0.080 | 0.008 | 0.992 | 0.007 |
| RT | GCA | -0.390 | 0.055 | -7.077 | 1.48E-12 | 0.146 | 0.005 | 1.034 | 0.015 |
| RT | Depression | 0.036 | 0.049 | 0.731 | 4.65E-01 | 0.045 | 0.002 | 1.018 | 0.013 |
| RT | Anxiety | -0.193 | 0.173 | -1.117 | 2.64E-01 | 0.061 | 0.022 | 1.001 | 0.007 |
| RT | Wellbeing | -0.072 | 0.049 | -1.476 | 1.40E-01 | 0.022 | 0.001 | 1.006 | 0.017 |

| | | | | | | | | | |
|---|---|---|---|---|---|---|---|---|---|
| RT | Brain Vol | -0.074 | 0.079 | -0.934 | 3.50E-01 | 0.217 | 0.018 | 1.020 | 0.009 |
| GCA | RT | -0.390 | 0.055 | -7.077 | 1.48E-12 | 0.079 | 0.014 | 0.991 | 0.007 |
| GCA | Memory | 0.537 | 0.048 | 11.153 | 6.90E-29 | 0.107 | 0.014 | 0.990 | 0.007 |
| GCA | WM | 0.527 | 0.060 | 8.812 | 1.23E-18 | 0.065 | 0.012 | 0.987 | 0.007 |
| GCA | Exec Func | 0.663 | 0.066 | 10.009 | 1.39E-23 | 0.083 | 0.014 | 0.996 | 0.007 |
| GCA | PANAS-Pos | -0.057 | 0.042 | -1.353 | 1.76E-01 | 0.070 | 0.009 | 0.987 | 0.007 |
| GCA | PANAS-Neg | -0.189 | 0.042 | -4.477 | 7.56E-06 | 0.080 | 0.009 | 0.992 | 0.007 |
| GCA | Depression | -0.176 | 0.017 | -10.537 | 5.81E-26 | 0.045 | 0.002 | 1.017 | 0.012 |
| GCA | Anxiety | -0.268 | 0.082 | -3.279 | 1.04E-03 | 0.060 | 0.022 | 1.002 | 0.007 |
| GCA | Wellbeing | 0.117 | 0.023 | 5.046 | 4.52E-07 | 0.022 | 0.001 | 1.007 | 0.017 |
| GCA | Brain Vol | 0.213 | 0.030 | 7.165 | 7.77E-13 | 0.218 | 0.019 | 1.020 | 0.009 |
| Depression | GCA | -0.176 | 0.017 | -10.537 | 5.81E-26 | 0.146 | 0.005 | 1.035 | 0.014 |
| Depression | RT | 0.036 | 0.049 | 0.731 | 4.65E-01 | 0.079 | 0.014 | 0.991 | 0.007 |
| Depression | Memory | -0.187 | 0.043 | -4.395 | 1.11E-05 | 0.107 | 0.014 | 0.990 | 0.007 |
| Depression | WM | -0.068 | 0.054 | -1.245 | 2.13E-01 | 0.065 | 0.011 | 0.987 | 0.007 |
| Depression | Exec Func | -0.154 | 0.046 | -3.328 | 8.75E-04 | 0.084 | 0.014 | 0.995 | 0.007 |
| Depression | PANAS-Pos | -0.111 | 0.040 | -2.807 | 5.01E-03 | 0.071 | 0.009 | 0.986 | 0.007 |
| Depression | PANAS-Neg | 0.506 | 0.046 | 11.028 | 2.80E-28 | 0.080 | 0.009 | 0.992 | 0.007 |
| Depression | Anxiety | 0.872 | 0.168 | 5.206 | 1.93E-07 | 0.059 | 0.022 | 1.002 | 0.007 |
| Depression | Wellbeing | -0.877 | 0.011 | -83.883 | 0.00E+00 | 0.022 | 0.001 | 1.007 | 0.017 |
| Depression | Brain Vol | -0.066 | 0.028 | -2.409 | 1.60E-02 | 0.218 | 0.019 | 1.020 | 0.009 |
| Anxiety | GCA | -0.268 | 0.082 | -3.279 | 1.04E-03 | 0.146 | 0.006 | 1.050 | 0.017 |
| Anxiety | RT | -0.193 | 0.173 | -1.117 | 2.64E-01 | 0.086 | 0.015 | 0.987 | 0.007 |
| Anxiety | Memory | 0.033 | 0.166 | 0.199 | 8.42E-01 | 0.108 | 0.015 | 0.991 | 0.008 |
| Anxiety | WM | -0.213 | 0.217 | -0.981 | 3.27E-01 | 0.060 | 0.012 | 0.991 | 0.007 |
| Anxiety | Exec Func | -0.003 | 0.172 | -0.020 | 9.84E-01 | 0.078 | 0.015 | 1.001 | 0.008 |
| Anxiety | PANAS-Pos | -0.163 | 0.168 | -0.967 | 3.33E-01 | 0.067 | 0.009 | 0.991 | 0.008 |
| Anxiety | PANAS-Neg | 0.695 | 0.183 | 3.787 | 1.52E-04 | 0.082 | 0.009 | 0.990 | 0.008 |
| Anxiety | Depression | 0.872 | 0.168 | 5.206 | 1.93E-07 | 0.046 | 0.002 | 1.025 | 0.014 |
| Anxiety | Wellbeing | -0.897 | 0.182 | -4.937 | 7.95E-07 | 0.022 | 0.001 | 1.010 | 0.018 |
| Anxiety | Brain Vol | -0.059 | 0.112 | -0.531 | 5.96E-01 | 0.212 | 0.021 | 1.029 | 0.011 |

| | | | | | | | | | |
|---|---|---|---|---|---|---|---|---|---|
| Wellbeing | GCA | 0.117 | 0.023 | 5.046 | 4.52E-07 | 0.149 | 0.006 | 1.025 | 0.019 |
| Wellbeing | RT | -0.072 | 0.049 | -1.476 | 1.40E-01 | 0.100 | 0.017 | 0.976 | 0.008 |
| Wellbeing | Memory | 0.108 | 0.039 | 2.814 | 4.90E-03 | 0.109 | 0.016 | 0.990 | 0.008 |
| Wellbeing | WM | 0.029 | 0.065 | 0.444 | 6.57E-01 | 0.057 | 0.014 | 0.993 | 0.009 |
| Wellbeing | Exec Func | 0.172 | 0.047 | 3.689 | 2.26E-04 | 0.092 | 0.015 | 0.993 | 0.008 |
| Wellbeing | PANAS-Pos | 0.297 | 0.050 | 5.970 | 2.38E-09 | 0.067 | 0.010 | 0.993 | 0.009 |
| Wellbeing | PANAS-Neg | -0.712 | 0.053 | -13.373 | 8.65E-41 | 0.076 | 0.009 | 0.995 | 0.008 |
| Wellbeing | Depression | -0.877 | 0.011 | -83.883 | 0.00E+00 | 0.046 | 0.002 | 1.009 | 0.017 |
| Wellbeing | Anxiety | -0.897 | 0.182 | -4.937 | 7.95E-07 | 0.058 | 0.023 | 1.001 | 0.008 |
| Wellbeing | Brain Vol | 0.064 | 0.031 | 2.043 | 4.11E-02 | 0.216 | 0.020 | 1.020 | 0.011 |
| Brain Vol | GCA | 0.213 | 0.030 | 7.165 | 7.77E-13 | 0.146 | 0.005 | 1.035 | 0.015 |
| Brain Vol | RT | -0.074 | 0.079 | -0.934 | 3.50E-01 | 0.080 | 0.016 | 0.990 | 0.007 |
| Brain Vol | Memory | 0.040 | 0.061 | 0.667 | 5.05E-01 | 0.106 | 0.014 | 0.990 | 0.007 |
| Brain Vol | WM | 0.078 | 0.082 | 0.952 | 3.41E-01 | 0.062 | 0.013 | 0.989 | 0.007 |
| Brain Vol | Exec Func | 0.247 | 0.070 | 3.503 | 4.59E-04 | 0.084 | 0.014 | 0.994 | 0.007 |
| Brain Vol | PANAS-Pos | 0.134 | 0.064 | 2.100 | 3.57E-02 | 0.070 | 0.009 | 0.986 | 0.007 |
| Brain Vol | PANAS-Neg | -0.005 | 0.066 | -0.069 | 9.45E-01 | 0.078 | 0.009 | 0.995 | 0.007 |
| Brain Vol | Depression | -0.066 | 0.028 | -2.409 | 1.60E-02 | 0.045 | 0.002 | 1.018 | 0.013 |
| Brain Vol | Anxiety | -0.059 | 0.112 | -0.531 | 5.96E-01 | 0.061 | 0.022 | 1.002 | 0.007 |
| Brain Vol | Wellbeing | 0.064 | 0.031 | 2.043 | 4.11E-02 | 0.022 | 0.001 | 1.006 | 0.018 |

PANAS-Neg=PANAS Negative Score; PANAS-POS=PANAS Positive Score; Executive Function=RFFT; Working Memory=One-Back Task; Memory=One-Card Learning Task; GCA=General Cognitive Ability; Brain Vol=Brain Volume.

***Table S4. Summary-level data Mendelian randomization results (GCA on mental health phenotypes).***

| Exposure | Outcome | Method | N SNPs | b | se | p-value | Lower CI | Higher CI |
|---|---|---|---|---|---|---|---|---|
| GCA | PANAS Negative | IVW | 249 | -0.106 | 0.029 | 0.00021 | -0.162 | -0.050 |
| GCA | PANAS Negative | MR Egger | 249 | -0.282 | 0.124 | 0.02448 | -0.526 | -0.038 |
| GCA | PANAS Negative | Weighted median | 249 | -0.086 | 0.037 | 0.02049 | -0.158 | -0.013 |
| GCA | PANAS Negative | Weighted mode | 249 | -0.081 | 0.112 | 0.47065 | -0.301 | 0.139 |
| GCA | PANAS Positive | IVW | 249 | -0.009 | 0.027 | 0.74921 | -0.062 | 0.045 |
| GCA | PANAS Positive | MR Egger | 249 | 0.026 | 0.119 | 0.82417 | -0.207 | 0.260 |
| GCA | PANAS Positive | Weighted median | 249 | -0.003 | 0.036 | 0.93973 | -0.073 | 0.068 |
| GCA | PANAS Positive | Weighted mode | 249 | 0.001 | 0.111 | 0.99230 | -0.217 | 0.219 |
| GCA | Depression | IVW | 227 | -0.127 | 0.031 | 0.00003 | -0.187 | -0.067 |
| GCA | Depression | MR Egger | 227 | -0.177 | 0.133 | 0.18536 | -0.439 | 0.084 |
| GCA | Depression | Weighted median | 227 | -0.086 | 0.025 | 0.00064 | -0.135 | -0.036 |
| GCA | Depression | Weighted mode | 227 | -0.056 | 0.072 | 0.43402 | -0.198 | 0.085 |
| GCA | Anxiety | IVW | 243 | -0.438 | 0.117 | 0.00017 | -0.666 | -0.209 |
| GCA | Anxiety | MR Egger | 243 | 0.095 | 0.579 | 0.86979 | -1.039 | 1.229 |
| GCA | Anxiety | Weighted median | 243 | -0.379 | 0.170 | 0.02617 | -0.713 | -0.045 |
| GCA | Anxiety | Weighted mode | 243 | -0.307 | 0.439 | 0.48537 | -1.166 | 0.553 |
| GCA | Wellbeing | IVW | 213 | 0.024 | 0.009 | 0.01113 | 0.005 | 0.042 |
| GCA | Wellbeing | MR Egger | 213 | 0.030 | 0.042 | 0.47908 | -0.053 | 0.113 |
| GCA | Wellbeing | Weighted median | 213 | 0.012 | 0.008 | 0.10046 | -0.002 | 0.027 |
| GCA | Wellbeing | Weighted mode | 213 | -0.001 | 0.021 | 0.97955 | -0.041 | 0.040 |

***Table S5. Summary-level data Mendelian randomization results (GCA on mental health phenotypes) [Steiger Filtered].***

| Exposure | Outcome | Method | N SNPs | b | se | p-value | Lower CI | Higher CI |
|---|---|---|---|---|---|---|---|---|
| GCA | PANAS Negative | IVW | 229 | -0.065 | 0.024 | 0.00695 | -0.113 | -0.018 |
| GCA | PANAS Negative | MR Egger | 229 | -0.193 | 0.106 | 0.07156 | -0.401 | 0.016 |
| GCA | PANAS Negative | Weighted median | 229 | -0.066 | 0.034 | 0.05308 | -0.133 | 0.001 |
| GCA | PANAS Negative | Weighted mode | 229 | -0.080 | 0.119 | 0.50094 | -0.313 | 0.153 |
| GCA | PANAS Positive | IVW | 242 | -0.025 | 0.025 | 0.31828 | -0.075 | 0.024 |
| GCA | PANAS Positive | MR Egger | 242 | -0.031 | 0.110 | 0.77488 | -0.246 | 0.183 |
| GCA | PANAS Positive | Weighted median | 242 | -0.007 | 0.037 | 0.85425 | -0.079 | 0.065 |
| GCA | PANAS Positive | Weighted mode | 242 | -0.005 | 0.117 | 0.96931 | -0.234 | 0.225 |
| GCA | Depression | IVW | 227 | -0.127 | 0.031 | 0.00003 | -0.187 | -0.067 |
| GCA | Depression | MR Egger | 227 | -0.177 | 0.133 | 0.18536 | -0.439 | 0.084 |
| GCA | Depression | Weighted median | 227 | -0.086 | 0.025 | 0.00064 | -0.135 | -0.036 |
| GCA | Depression | Weighted mode | 227 | -0.056 | 0.072 | 0.43402 | -0.198 | 0.085 |
| GCA | Anxiety | IVW | 178 | -0.238 | 0.132 | 0.07191 | -0.497 | 0.021 |
| GCA | Anxiety | MR Egger | 178 | -0.893 | 0.640 | 0.16456 | -2.147 | 0.361 |
| GCA | Anxiety | Weighted median | 178 | -0.272 | 0.183 | 0.13575 | -0.630 | 0.085 |
| GCA | Anxiety | Weighted mode | 178 | -0.349 | 0.455 | 0.44504 | -1.241 | 0.544 |
| GCA | Wellbeing | IVW | 213 | 0.024 | 0.009 | 0.01113 | 0.005 | 0.042 |
| GCA | Wellbeing | MR Egger | 213 | 0.030 | 0.042 | 0.47908 | -0.053 | 0.113 |
| GCA | Wellbeing | Weighted median | 213 | 0.012 | 0.008 | 0.10046 | -0.002 | 0.027 |
| GCA | Wellbeing | Weighted mode | 213 | -0.001 | 0.021 | 0.97955 | -0.041 | 0.040 |

***Table S6. Summary-level data Mendelian randomization results (Depression on cognition)***

| Exposure | Outcome | Method | N SNPs | b | se | p-value | Lower CI | Higher CI |
|---|---|---|---|---|---|---|---|---|
| Depression | GCA | IVW | 130 | -0.140 | 0.026 | 5.78E-08 | -0.191 | -0.090 |
| Depression | GCA | MR Egger | 130 | -0.330 | 0.112 | 3.77E-03 | -0.549 | -0.111 |
| Depression | GCA | Weighted median | 130 | -0.134 | 0.020 | 1.87E-11 | -0.173 | -0.095 |
| Depression | GCA | Weighted mode | 130 | -0.137 | 0.045 | 2.72E-03 | -0.224 | -0.049 |
| Depression | Memory | IVW | 129 | -0.036 | 0.033 | 2.86E-01 | -0.101 | 0.030 |
| Depression | Memory | MR Egger | 129 | -0.196 | 0.146 | 1.82E-01 | -0.482 | 0.090 |
| Depression | Memory | Weighted median | 129 | -0.092 | 0.045 | 4.15E-02 | -0.181 | -0.004 |
| Depression | Memory | Weighted mode | 129 | -0.226 | 0.126 | 7.46E-02 | -0.472 | 0.020 |
| Depression | Working Memory | IVW | 129 | 0.015 | 0.036 | 6.76E-01 | -0.056 | 0.086 |
| Depression | Working Memory | MR Egger | 129 | -0.061 | 0.159 | 7.03E-01 | -0.372 | 0.251 |
| Depression | Working Memory | Weighted median | 129 | 0.018 | 0.048 | 6.99E-01 | -0.075 | 0.112 |
| Depression | Working Memory | Weighted mode | 129 | 0.013 | 0.119 | 9.17E-01 | -0.221 | 0.246 |
| Depression | Executive Function | IVW | 129 | -0.045 | 0.036 | 2.11E-01 | -0.114 | 0.025 |
| Depression | Executive Function | MR Egger | 129 | 0.254 | 0.154 | 1.01E-01 | -0.048 | 0.556 |
| Depression | Executive Function | Weighted median | 129 | -0.064 | 0.045 | 1.54E-01 | -0.153 | 0.024 |
| Depression | Executive Function | Weighted mode | 129 | -0.169 | 0.137 | 2.21E-01 | -0.437 | 0.100 |
| Depression | Reaction Time | IVW | 129 | -0.042 | 0.033 | 2.05E-01 | -0.108 | 0.023 |
| Depression | Reaction Time | MR Egger | 129 | -0.097 | 0.146 | 5.09E-01 | -0.384 | 0.190 |
| Depression | Reaction Time | Weighted median | 129 | -0.060 | 0.043 | 1.67E-01 | -0.144 | 0.025 |
| Depression | Reaction Time | Weighted mode | 129 | -0.084 | 0.108 | 4.36E-01 | -0.295 | 0.127 |

***Table S7. Summary-level data Mendelian randomization results (Depression on cognition) [Steiger Filtered]***

| Exposure | Outcome | Method | N SNPs | b | se | p-value | Lower CI | Higher CI |
|---|---|---|---|---|---|---|---|---|
| Depression | GCA | IVW | 108 | -0.113 | 0.019 | 1.87E-09 | -0.150 | -0.076 |
| Depression | GCA | MR Egger | 108 | -0.216 | 0.087 | 1.51E-02 | -0.387 | -0.045 |
| Depression | GCA | Weighted median | 108 | -0.128 | 0.020 | 7.16E-11 | -0.167 | -0.090 |
| Depression | GCA | Weighted mode | 108 | -0.140 | 0.054 | 1.11E-02 | -0.246 | -0.034 |
| Depression | Memory | IVW | 88 | -0.035 | 0.037 | 3.45E-01 | -0.108 | 0.038 |
| Depression | Memory | MR Egger | 88 | -0.099 | 0.166 | 5.50E-01 | -0.424 | 0.225 |
| Depression | Memory | Weighted median | 88 | -0.041 | 0.049 | 4.04E-01 | -0.138 | 0.056 |
| Depression | Memory | Weighted mode | 88 | -0.198 | 0.124 | 1.16E-01 | -0.442 | 0.046 |
| Depression | Working Memory | IVW | 84 | -0.007 | 0.038 | 8.56E-01 | -0.082 | 0.068 |
| Depression | Working Memory | MR Egger | 84 | -0.011 | 0.160 | 9.44E-01 | -0.325 | 0.302 |
| Depression | Working Memory | Weighted median | 84 | 0.000 | 0.053 | 9.96E-01 | -0.104 | 0.105 |
| Depression | Working Memory | Weighted mode | 84 | 0.067 | 0.112 | 5.52E-01 | -0.152 | 0.286 |
| Depression | Executive Function | IVW | 86 | -0.016 | 0.037 | 6.66E-01 | -0.089 | 0.057 |
| Depression | Executive Function | MR Egger | 86 | 0.042 | 0.178 | 8.14E-01 | -0.308 | 0.392 |
| Depression | Executive Function | Weighted median | 86 | -0.007 | 0.051 | 8.82E-01 | -0.107 | 0.092 |
| Depression | Executive Function | Weighted mode | 86 | -0.197 | 0.127 | 1.24E-01 | -0.445 | 0.051 |
| Depression | Reaction Time | IVW | 87 | -0.014 | 0.035 | 6.86E-01 | -0.083 | 0.055 |
| Depression | Reaction Time | MR Egger | 87 | 0.048 | 0.141 | 7.32E-01 | -0.228 | 0.324 |
| Depression | Reaction Time | Weighted median | 87 | -0.037 | 0.048 | 4.41E-01 | -0.131 | 0.057 |
| Depression | Reaction Time | Weighted mode | 87 | -0.112 | 0.106 | 2.95E-01 | -0.320 | 0.096 |

***Table S8. Summary-level data Mendelian randomization results (Wellbeing on cognition)***

| Exposure | Outcome | Method | N SNPs | b | se | p-value | Lower CI | Higher CI |
|---|---|---|---|---|---|---|---|---|
| Wellbeing | GCA | IVW | 157 | 0.297 | 0.084 | 3.96E-04 | 0.133 | 0.462 |
| Wellbeing | GCA | MR Egger | 157 | 0.853 | 0.384 | 2.79E-02 | 0.100 | 1.606 |
| Wellbeing | GCA | Weighted median | 157 | 0.391 | 0.061 | 1.52E-10 | 0.271 | 0.511 |
| Wellbeing | GCA | Weighted mode | 157 | 0.540 | 0.166 | 1.38E-03 | 0.215 | 0.866 |
| Wellbeing | Memory | IVW | 157 | 0.193 | 0.108 | 7.56E-02 | -0.020 | 0.405 |
| Wellbeing | Memory | MR Egger | 157 | 0.273 | 0.517 | 5.99E-01 | -0.740 | 1.286 |
| Wellbeing | Memory | Weighted median | 157 | 0.308 | 0.148 | 3.75E-02 | 0.018 | 0.598 |
| Wellbeing | Memory | Weighted mode | 157 | 0.748 | 0.393 | 5.87E-02 | -0.022 | 1.517 |
| Wellbeing | Working Memory | IVW | 158 | -0.096 | 0.111 | 3.90E-01 | -0.314 | 0.123 |
| Wellbeing | Working Memory | MR Egger | 158 | -0.058 | 0.533 | 9.13E-01 | -1.103 | 0.986 |
| Wellbeing | Working Memory | Weighted median | 158 | -0.087 | 0.146 | 5.53E-01 | -0.373 | 0.200 |
| Wellbeing | Working Memory | Weighted mode | 158 | -0.536 | 0.433 | 2.17E-01 | -1.385 | 0.312 |
| Wellbeing | Executive Function | IVW | 158 | 0.235 | 0.104 | 2.43E-02 | 0.031 | 0.440 |
| Wellbeing | Executive Function | MR Egger | 158 | -0.224 | 0.496 | 6.52E-01 | -1.196 | 0.748 |
| Wellbeing | Executive Function | Weighted median | 158 | 0.299 | 0.139 | 3.13E-02 | 0.027 | 0.572 |
| Wellbeing | Executive Function | Weighted mode | 158 | 0.377 | 0.383 | 3.26E-01 | -0.373 | 1.127 |
| Wellbeing | Reaction Time | IVW | 157 | 0.026 | 0.097 | 7.93E-01 | -0.165 | 0.216 |
| Wellbeing | Reaction Time | MR Egger | 157 | 0.229 | 0.465 | 6.23E-01 | -0.682 | 1.140 |
| Wellbeing | Reaction Time | Weighted median | 157 | -0.047 | 0.135 | 7.26E-01 | -0.312 | 0.217 |
| Wellbeing | Reaction Time | Weighted mode | 157 | -0.152 | 0.379 | 6.89E-01 | -0.895 | 0.591 |

***Table S9. Summary-level data Mendelian randomization results (Wellbeing on cognition) [Steiger Filtered]***

| Exposure | Outcome | Method | N SNPs | b | se | p-value | Lower CI | Higher CI |
|---|---|---|---|---|---|---|---|---|
| Wellbeing | GCA | IVW | 110 | 0.232 | 0.053 | 1.058E-05 | 0.129 | 0.335 |
| Wellbeing | GCA | MR Egger | 110 | 0.407 | 0.267 | 1.297E-01 | -0.116 | 0.931 |
| Wellbeing | GCA | Weighted median | 110 | 0.330 | 0.062 | 1.066E-07 | 0.208 | 0.451 |
| Wellbeing | GCA | Weighted mode | 110 | 0.581 | 0.169 | 8.307E-04 | 0.250 | 0.912 |
| Wellbeing | Memory | IVW | 75 | 0.019 | 0.141 | 8.911E-01 | -0.257 | 0.296 |
| Wellbeing | Memory | MR Egger | 75 | -0.124 | 0.607 | 8.389E-01 | -1.313 | 1.065 |
| Wellbeing | Memory | Weighted median | 75 | 0.015 | 0.178 | 9.340E-01 | -0.335 | 0.364 |
| Wellbeing | Memory | Weighted mode | 75 | 0.508 | 0.405 | 2.138E-01 | -0.286 | 1.303 |
| Wellbeing | Working Memory | IVW | 80 | -0.012 | 0.139 | 9.300E-01 | -0.284 | 0.260 |
| Wellbeing | Working Memory | MR Egger | 80 | -0.408 | 0.609 | 5.051E-01 | -1.600 | 0.785 |
| Wellbeing | Working Memory | Weighted median | 80 | -0.056 | 0.184 | 7.598E-01 | -0.417 | 0.304 |
| Wellbeing | Working Memory | Weighted mode | 80 | -0.414 | 0.372 | 2.689E-01 | -1.144 | 0.315 |
| Wellbeing | Executive Function | IVW | 75 | 0.068 | 0.135 | 6.145E-01 | -0.196 | 0.332 |
| Wellbeing | Executive Function | MR Egger | 75 | 0.036 | 0.592 | 9.521E-01 | -1.125 | 1.196 |
| Wellbeing | Executive Function | Weighted median | 75 | 0.108 | 0.176 | 5.387E-01 | -0.237 | 0.454 |
| Wellbeing | Executive Function | Weighted mode | 75 | 0.378 | 0.358 | 2.953E-01 | -0.325 | 1.080 |
| Wellbeing | Reaction Time | IVW | 83 | -0.031 | 0.124 | 7.988E-01 | -0.274 | 0.211 |
| Wellbeing | Reaction Time | MR Egger | 83 | -0.179 | 0.509 | 7.260E-01 | -1.176 | 0.818 |
| Wellbeing | Reaction Time | Weighted median | 83 | -0.092 | 0.160 | 5.628E-01 | -0.405 | 0.220 |
| Wellbeing | Reaction Time | Weighted mode | 83 | -0.195 | 0.385 | 6.136E-01 | -0.950 | 0.560 |

***Table S10. Summary-level data Mendelian randomization results (PANAS Negative on cognition)***

| Exposure | Outcome | Method | N SNPs | b | se | p-value | Lower CI | Higher CI |
|---|---|---|---|---|---|---|---|---|
| PANAS Negative | GCA | IVW | 20 | -0.060 | 0.036 | 0.093 | -0.130 | 0.010 |
| PANAS Negative | GCA | MR Egger | 20 | -0.050 | 0.080 | 0.543 | -0.208 | 0.108 |
| PANAS Negative | GCA | Weighted median | 20 | -0.040 | 0.028 | 0.158 | -0.095 | 0.015 |
| PANAS Negative | GCA | Weighted mode | 20 | -0.026 | 0.030 | 0.394 | -0.086 | 0.033 |
| PANAS Negative | Memory | IVW | 20 | -0.150 | 0.060 | 0.012 | -0.266 | -0.033 |
| PANAS Negative | Memory | MR Egger | 20 | -0.227 | 0.150 | 0.148 | -0.522 | 0.067 |
| PANAS Negative | Memory | Weighted median | 20 | -0.181 | 0.084 | 0.031 | -0.345 | -0.017 |
| PANAS Negative | Memory | Weighted mode | 20 | -0.212 | 0.160 | 0.200 | -0.526 | 0.101 |
| PANAS Negative | Working Memory | IVW | 20 | -0.075 | 0.059 | 0.205 | -0.191 | 0.041 |
| PANAS Negative | Working Memory | MR Egger | 20 | -0.034 | 0.150 | 0.824 | -0.327 | 0.260 |
| PANAS Negative | Working Memory | Weighted median | 20 | 0.004 | 0.084 | 0.962 | -0.161 | 0.169 |
| PANAS Negative | Working Memory | Weighted mode | 20 | 0.051 | 0.151 | 0.741 | -0.245 | 0.347 |
| PANAS Negative | Executive Function | IVW | 20 | 0.035 | 0.070 | 0.619 | -0.103 | 0.173 |
| PANAS Negative | Executive Function | MR Egger | 20 | -0.110 | 0.178 | 0.544 | -0.458 | 0.238 |
| PANAS Negative | Executive Function | Weighted median | 20 | -0.006 | 0.087 | 0.941 | -0.177 | 0.165 |
| PANAS Negative | Executive Function | Weighted mode | 20 | -0.093 | 0.155 | 0.554 | -0.397 | 0.210 |
| PANAS Negative | Reaction Time | IVW | 20 | -0.014 | 0.070 | 0.841 | -0.152 | 0.124 |
| PANAS Negative | Reaction Time | MR Egger | 20 | 0.248 | 0.170 | 0.163 | -0.086 | 0.581 |
| PANAS Negative | Reaction Time | Weighted median | 20 | -0.062 | 0.079 | 0.433 | -0.218 | 0.093 |
| PANAS Negative | Reaction Time | Weighted mode | 20 | -0.090 | 0.149 | 0.553 | -0.383 | 0.202 |

***Table S11. Summary-level data Mendelian randomization results (PANAS Negative on cognition) [Steiger Filtered]***

| Exposure | Outcome | Method | N SNPs | b | se | p-value | Lower CI | Higher CI |
|---|---|---|---|---|---|---|---|---|
| PANAS Negative | GCA | IVW | 20 | -0.060 | 0.036 | 0.093 | -0.130 | 0.010 |
| PANAS Negative | GCA | MR Egger | 20 | -0.050 | 0.080 | 0.543 | -0.208 | 0.108 |
| PANAS Negative | GCA | Weighted median | 20 | -0.040 | 0.028 | 0.158 | -0.095 | 0.015 |
| PANAS Negative | GCA | Weighted mode | 20 | -0.026 | 0.030 | 0.394 | -0.086 | 0.033 |
| PANAS Negative | Memory | IVW | 20 | -0.150 | 0.060 | 0.012 | -0.266 | -0.033 |
| PANAS Negative | Memory | MR Egger | 20 | -0.227 | 0.150 | 0.148 | -0.522 | 0.067 |
| PANAS Negative | Memory | Weighted median | 20 | -0.181 | 0.084 | 0.031 | -0.345 | -0.017 |
| PANAS Negative | Memory | Weighted mode | 20 | -0.212 | 0.160 | 0.200 | -0.526 | 0.101 |
| PANAS Negative | Working Memory | IVW | 20 | -0.075 | 0.059 | 0.205 | -0.191 | 0.041 |
| PANAS Negative | Working Memory | MR Egger | 20 | -0.034 | 0.150 | 0.824 | -0.327 | 0.260 |
| PANAS Negative | Working Memory | Weighted median | 20 | 0.004 | 0.084 | 0.962 | -0.161 | 0.169 |
| PANAS Negative | Working Memory | Weighted mode | 20 | 0.051 | 0.151 | 0.741 | -0.245 | 0.347 |
| PANAS Negative | Executive Function | IVW | 20 | 0.035 | 0.070 | 0.619 | -0.103 | 0.173 |
| PANAS Negative | Executive Function | MR Egger | 20 | -0.110 | 0.178 | 0.544 | -0.458 | 0.238 |
| PANAS Negative | Executive Function | Weighted median | 20 | -0.006 | 0.087 | 0.941 | -0.177 | 0.165 |
| PANAS Negative | Executive Function | Weighted mode | 20 | -0.093 | 0.155 | 0.554 | -0.397 | 0.210 |
| PANAS Negative | Reaction Time | IVW | 20 | -0.014 | 0.070 | 0.841 | -0.152 | 0.124 |
| PANAS Negative | Reaction Time | MR Egger | 20 | 0.248 | 0.170 | 0.163 | -0.086 | 0.581 |
| PANAS Negative | Reaction Time | Weighted median | 20 | -0.062 | 0.079 | 0.433 | -0.218 | 0.093 |
| PANAS Negative | Reaction Time | Weighted mode | 20 | -0.090 | 0.149 | 0.553 | -0.383 | 0.202 |

***Table S12. Summary-level data Mendelian randomization results (PANAS Positive on cognition)***

| **Exposure** | **Outcome** | **Method** | **N SNPs** | **b** | **se** | **p-value** | **Lower CI** | **Higher CI** |
|---|---|---|---|---|---|---|---|---|
| PANAS Positive | GCA | IVW | 14 | 0.009 | 0.043 | 0.829 | -0.075 | 0.094 |
| PANAS Positive | GCA | MR Egger | 14 | -0.012 | 0.117 | 0.919 | -0.242 | 0.217 |
| PANAS Positive | GCA | Weighted median | 14 | 0.036 | 0.036 | 0.312 | -0.034 | 0.106 |
| PANAS Positive | GCA | Weighted mode | 14 | 0.043 | 0.046 | 0.363 | -0.047 | 0.133 |
| PANAS Positive | Memory | IVW | 15 | 0.102 | 0.071 | 0.150 | -0.037 | 0.240 |
| PANAS Positive | Memory | MR Egger | 15 | 0.101 | 0.208 | 0.635 | -0.306 | 0.508 |
| PANAS Positive | Memory | Weighted median | 15 | 0.088 | 0.094 | 0.349 | -0.096 | 0.272 |
| PANAS Positive | Memory | Weighted mode | 15 | -0.031 | 0.190 | 0.874 | -0.403 | 0.341 |
| PANAS Positive | Working Memory | IVW | 15 | 0.037 | 0.067 | 0.578 | -0.093 | 0.167 |
| PANAS Positive | Working Memory | MR Egger | 15 | -0.088 | 0.188 | 0.648 | -0.457 | 0.281 |
| PANAS Positive | Working Memory | Weighted median | 15 | 0.068 | 0.088 | 0.441 | -0.105 | 0.242 |
| PANAS Positive | Working Memory | Weighted mode | 15 | 0.033 | 0.158 | 0.835 | -0.276 | 0.343 |
| PANAS Positive | Executive Function | IVW | 15 | 0.161 | 0.071 | 0.024 | 0.021 | 0.300 |
| PANAS Positive | Executive Function | MR Egger | 15 | -0.077 | 0.196 | 0.703 | -0.461 | 0.308 |
| PANAS Positive | Executive Function | Weighted median | 15 | 0.167 | 0.090 | 0.063 | -0.009 | 0.343 |
| PANAS Positive | Executive Function | Weighted mode | 15 | 0.163 | 0.173 | 0.364 | -0.177 | 0.503 |
| PANAS Positive | Reaction Time | IVW | 15 | -0.068 | 0.062 | 0.272 | -0.190 | 0.053 |
| PANAS Positive | Reaction Time | MR Egger | 15 | -0.177 | 0.176 | 0.332 | -0.521 | 0.167 |
| PANAS Positive | Reaction Time | Weighted median | 15 | -0.034 | 0.084 | 0.686 | -0.198 | 0.130 |
| PANAS Positive | Reaction Time | Weighted mode | 15 | -0.038 | 0.147 | 0.800 | -0.325 | 0.249 |

***Table S13. Summary-level data Mendelian randomization results (PANAS Positive on cognition) [Steiger Filtered]***

| Exposure | Outcome | Method | N SNPs | b | se | p-value | Lower CI | Higher CI |
|---|---|---|---|---|---|---|---|---|
| PANAS Positive | GCA | IVW | 14 | 0.009 | 0.043 | 0.829 | -0.075 | 0.094 |
| PANAS Positive | GCA | MR Egger | 14 | -0.012 | 0.117 | 0.919 | -0.242 | 0.217 |
| PANAS Positive | GCA | Weighted median | 14 | 0.036 | 0.036 | 0.312 | -0.034 | 0.106 |
| PANAS Positive | GCA | Weighted mode | 14 | 0.043 | 0.046 | 0.363 | -0.047 | 0.133 |
| PANAS Positive | Memory | IVW | 15 | 0.102 | 0.071 | 0.150 | -0.037 | 0.240 |
| PANAS Positive | Memory | MR Egger | 15 | 0.101 | 0.208 | 0.635 | -0.306 | 0.508 |
| PANAS Positive | Memory | Weighted median | 15 | 0.088 | 0.094 | 0.349 | -0.096 | 0.272 |
| PANAS Positive | Memory | Weighted mode | 15 | -0.031 | 0.190 | 0.874 | -0.403 | 0.341 |
| PANAS Positive | Working Memory | IVW | 15 | 0.037 | 0.067 | 0.578 | -0.093 | 0.167 |
| PANAS Positive | Working Memory | MR Egger | 15 | -0.088 | 0.188 | 0.648 | -0.457 | 0.281 |
| PANAS Positive | Working Memory | Weighted median | 15 | 0.068 | 0.088 | 0.441 | -0.105 | 0.242 |
| PANAS Positive | Working Memory | Weighted mode | 15 | 0.033 | 0.158 | 0.835 | -0.276 | 0.343 |
| PANAS Positive | Executive Function | IVW | 15 | 0.161 | 0.071 | 0.024 | 0.021 | 0.300 |
| PANAS Positive | Executive Function | MR Egger | 15 | -0.077 | 0.196 | 0.703 | -0.461 | 0.308 |
| PANAS Positive | Executive Function | Weighted median | 15 | 0.167 | 0.090 | 0.063 | -0.009 | 0.343 |
| PANAS Positive | Executive Function | Weighted mode | 15 | 0.163 | 0.173 | 0.364 | -0.177 | 0.503 |
| PANAS Positive | Reaction Time | IVW | 15 | -0.068 | 0.062 | 0.272 | -0.190 | 0.053 |
| PANAS Positive | Reaction Time | MR Egger | 15 | -0.177 | 0.176 | 0.332 | -0.521 | 0.167 |
| PANAS Positive | Reaction Time | Weighted median | 15 | -0.034 | 0.084 | 0.686 | -0.198 | 0.130 |
| PANAS Positive | Reaction Time | Weighted mode | 15 | -0.038 | 0.147 | 0.800 | -0.325 | 0.249 |

***Table S14. Summary-level data Mendelian randomization results (Anxiety on cognition)***

| Exposure | Outcome | Method | N SNPs | b | se | p-value | Lower CI | Higher CI |
|---|---|---|---|---|---|---|---|---|
| Anxiety | General Cognitive Ability | Wald ratio | 1 | -0.028 | 0.016 | 0.089 | -0.060 | 0.004 |
| Anxiety | Memory | Wald ratio | 1 | 0.015 | 0.048 | 0.763 | -0.080 | 0.109 |
| Anxiety | Working Memory | Wald ratio | 1 | -0.003 | 0.048 | 0.945 | -0.098 | 0.092 |
| Anxiety | Executive Function | Wald ratio | 1 | 0.003 | 0.047 | 0.955 | -0.090 | 0.095 |

***Table S15. Tests for heterogeneity of MR results.***

| Exposure | Outcome | Method | Q | Q DF | p-value |
|---|---|---|---|---|---|
| GCA | PANAS_Negative | MR Egger | 372.079 | 247 | 4.43E-07 |
| GCA | PANAS_Negative | Inverse variance weighted | 375.240 | 248 | 3.15E-07 |
| GCA | PANAS_Positive | MR Egger | 327.719 | 247 | 4.39E-04 |
| GCA | PANAS_Positive | Inverse variance weighted | 327.842 | 248 | 5.03E-04 |
| GCA | Depression | MR Egger | 1173.521 | 225 | 4.76E-128 |
| GCA | Depression | Inverse variance weighted | 1174.292 | 226 | 7.97E-128 |
| GCA | Anxiety | MR Egger | 249.793 | 241 | 3.35E-01 |
| GCA | Anxiety | Inverse variance weighted | 250.709 | 242 | 3.37E-01 |
| GCA | Wellbeing | MR Egger | 1143.904 | 211 | 6.50E-128 |
| GCA | Wellbeing | Inverse variance weighted | 1144.033 | 212 | 1.44E-127 |
| Depression | General Cognitive Ability | MR Egger | 785.80 | 128 | 3.83E-95 |
| Depression | General Cognitive Ability | Inverse variance weighted | 804.40 | 129 | 3.84E-98 |
| Depression | Memory | MR Egger | 143.92 | 127 | 1.45E-01 |
| Depression | Memory | Inverse variance weighted | 145.37 | 128 | 1.40E-01 |
| Depression | Working Memory | MR Egger | 172.75 | 127 | 4.34E-03 |
| Depression | Working Memory | Inverse variance weighted | 173.07 | 128 | 4.91E-03 |
| Depression | Executive Function | MR Egger | 171.45 | 127 | 5.26E-03 |
| Depression | Executive Function | Inverse variance weighted | 176.82 | 128 | 2.78E-03 |
| Depression | Reaction Time | MR Egger | 169.09 | 127 | 7.42E-03 |
| Depression | Reaction Time | Inverse variance weighted | 169.29 | 128 | 8.50E-03 |
| Wellbeing | General Cognitive Ability | MR Egger | 1037.178 | 155 | 2.06E-130 |
| Wellbeing | General Cognitive Ability | Inverse variance weighted | 1051.864 | 156 | 1.02E-132 |
| Wellbeing | Memory | MR Egger | 189.367 | 155 | 3.14E-02 |
| Wellbeing | Memory | Inverse variance weighted | 189.397 | 156 | 3.53E-02 |
| Wellbeing | Working Memory | MR Egger | 205.014 | 156 | 5.16E-03 |
| Wellbeing | Working Memory | Inverse variance weighted | 205.020 | 157 | 6.02E-03 |
| Wellbeing | Executive Function | MR Egger | 188.128 | 156 | 4.06E-02 |
| Wellbeing | Executive Function | Inverse variance weighted | 189.210 | 157 | 4.06E-02 |
| Wellbeing | Reaction Time | MR Egger | 177.598 | 155 | 1.03E-01 |

| Exposure | Outcome | Method | Q | Q DF | p-value |
|---|---|---|---|---|---|
| Wellbeing | Reaction Time | Inverse variance weighted | 177.828 | 156 | 1.11E-01 |
| PANAS Negative | General Cognitive Ability | MR Egger | 82.437 | 18 | 3.20E-10 |
| PANAS Negative | General Cognitive Ability | Inverse variance weighted | 82.530 | 19 | 6.79E-10 |
| PANAS Negative | Memory | MR Egger | 17.919 | 18 | 4.61E-01 |
| PANAS Negative | Memory | Inverse variance weighted | 18.235 | 19 | 5.07E-01 |
| PANAS Negative | Working Memory | MR Egger | 16.732 | 18 | 5.42E-01 |
| PANAS Negative | Working Memory | Inverse variance weighted | 16.823 | 19 | 6.02E-01 |
| PANAS Negative | Executive Function | MR Egger | 27.324 | 18 | 7.31E-02 |
| PANAS Negative | Executive Function | Inverse variance weighted | 28.524 | 19 | 7.39E-02 |
| PANAS Negative | Reaction Time | MR Egger | 26.686 | 18 | 8.51E-02 |
| PANAS Negative | Reaction Time | Inverse variance weighted | 30.851 | 19 | 4.19E-02 |
| PANAS Positive | General Cognitive Ability | MR Egger | 47.910 | 12 | 3.24E-06 |
| PANAS Positive | General Cognitive Ability | Inverse variance weighted | 48.066 | 13 | 6.38E-06 |
| PANAS Positive | Episodic Memory | MR Egger | 15.629 | 13 | 2.70E-01 |
| PANAS Positive | Episodic Memory | Inverse variance weighted | 15.629 | 14 | 3.37E-01 |
| PANAS Positive | Working Memory | MR Egger | 6.952 | 13 | 9.05E-01 |
| PANAS Positive | Working Memory | Inverse variance weighted | 7.456 | 14 | 9.16E-01 |
| PANAS Positive | Executive Function | MR Egger | 14.709 | 13 | 3.26E-01 |
| PANAS Positive | Executive Function | Inverse variance weighted | 16.605 | 14 | 2.78E-01 |
| PANAS Positive | Reaction Time | MR Egger | 12.149 | 13 | 5.15E-01 |
| PANAS Positive | Reaction Time | Inverse variance weighted | 12.588 | 14 | 5.59E-01 |

***Table S16. Test of horizontal pleiotropy of MR results (using Egger Intercept)***

| Exposure | Outcome | Egger Intercept | se | p-value |
|---|---|---|---|---|
| GCA | PANAS Negative | 0.003 | 0.002 | 0.149 |
| GCA | PANAS Positive | -0.001 | 0.002 | 0.762 |
| GCA | Depression | 0.001 | 0.002 | 0.701 |
| GCA | Anxiety | -0.009 | 0.010 | 0.348 |
| Depression | General Cognitive Ability | 0.005 | 0.003 | 0.084 |
| Depression | Episodic Memory | 0.004 | 0.003 | 0.261 |
| Depression | Working Memory | 0.002 | 0.004 | 0.625 |
| Depression | Executive Function | -0.007 | 0.004 | 0.048 |
| Wellbeing | General Cognitive Ability | -0.004 | 0.003 | 0.141 |
| Wellbeing | Episodic Memory | -0.001 | 0.003 | 0.874 |
| Wellbeing | Working Memory | 0.000 | 0.004 | 0.943 |
| Wellbeing | Executive Function | 0.003 | 0.003 | 0.345 |
| PANAS Negative | General Cognitive Ability | -0.001 | 0.004 | 0.888 |
| PANAS Negative | Episodic Memory | 0.003 | 0.006 | 0.581 |
| PANAS Negative | Working Memory | -0.002 | 0.006 | 0.767 |
| PANAS Negative | Executive Function | 0.006 | 0.007 | 0.386 |
| PANAS Positive | General Cognitive Ability | 0.001 | 0.005 | 0.846 |
| PANAS Positive | Episodic Memory | 0.000 | 0.009 | 0.997 |
| PANAS Positive | Working Memory | 0.006 | 0.008 | 0.490 |
| PANAS Positive | Executive Function | 0.010 | 0.008 | 0.218 |

***Table S17. Summary-level data Mendelian randomization results comparing current results and within-sibship results***
(current study vs population estimates *(Howe et al., 2022)* vs within-sibship estimates *(Howe et al., 2022)*).

| Exposure | Outcome | N SNPs | Method | b | se | p-value | Lower CI | Higher CI |
|---|---|---|---|---|---|---|---|---|
| Depression (current study) | Cognition (current study) | 130 | IVW | -0.140 | 0.026 | 5.78E-08 | -0.191 | -0.090 |
| Depression (current study) | Cognition (current study) | 130 | MR Egger | -0.330 | 0.112 | 3.77E-03 | -0.549 | -0.111 |
| Depression (current study) | Cognition (current study) | 130 | Weighted median | -0.134 | 0.020 | 1.87E-11 | -0.173 | -0.095 |
| Depression (current study) | Cognition (current study) | 130 | Weighted mode | -0.137 | 0.045 | 2.72E-03 | -0.224 | -0.049 |
| Depression (Howe et al., 2022: pop est) | Cognition (Howe et al., 2022: pop est) | 22 | MR Egger | -0.145 | 0.163 | 3.86E-01 | -0.460 | 0.180 |
| Depression (Howe et al., 2022: pop est) | Cognition (Howe et al., 2022: pop est) | 22 | Weighted median | -0.045 | 0.088 | 6.05E-01 | -0.220 | 0.130 |
| Depression (Howe et al., 2022: pop est) | Cognition (Howe et al., 2022: pop est) | 22 | IVW | -0.024 | 0.064 | 7.11E-01 | -0.150 | 0.100 |
| Depression (Howe et al., 2022: pop est) | Cognition (Howe et al., 2022: pop est) | 22 | Weighted mode | -0.090 | 0.154 | 5.65E-01 | -0.390 | 0.210 |
| Depression (Howe et al., 2022: sib est) | Cognition (Howe et al., 2022: sib est) | 6 | MR Egger | -0.410 | 0.452 | 4.16E-01 | -1.300 | 0.480 |
| Depression (Howe et al., 2022: sib est) | Cognition (Howe et al., 2022: sib est) | 6 | Weighted median | 0.102 | 0.137 | 4.58E-01 | -0.170 | 0.370 |
| Depression (Howe et al., 2022: sib est) | Cognition (Howe et al., 2022: sib est) | 6 | IVW | 0.006 | 0.166 | 9.72E-01 | -0.320 | 0.330 |

| | | | | | | | | |
|---|---|---|---|---|---|---|---|---|
| Depression (Howe et al., 2022: sib est) | Cognition (Howe et al., 2022: sib est) | 6 | Weighted mode | 0.244 | 0.195 | 2.66E-01 | -0.140 | 0.630 |
| Wellbeing (current study) | Cognition (current study) | 157 | IVW | 0.297 | 0.084 | 3.96E-04 | 0.133 | 0.462 |
| Wellbeing (current study) | Cognition (current study) | 157 | MR Egger | 0.853 | 0.384 | 2.79E-02 | 0.100 | 1.606 |
| Wellbeing (current study) | Cognition (current study) | 157 | Weighted median | 0.391 | 0.061 | 1.52E-10 | 0.271 | 0.511 |
| Wellbeing (current study) | Cognition (current study) | 157 | Weighted mode | 0.540 | 0.166 | 1.38E-03 | 0.215 | 0.866 |
| Wellbeing (Howe et al., 2022: pop est) | Cognition (Howe et al., 2022: pop est) | 23 | MR Egger | -0.315 | 0.214 | 1.55E-01 | -0.730 | 0.100 |
| Wellbeing (Howe et al., 2022: pop est) | Cognition (Howe et al., 2022: pop est) | 23 | Weighted median | 0.029 | 0.101 | 7.74E-01 | -0.170 | 0.230 |
| Wellbeing (Howe et al., 2022: pop est) | Cognition (Howe et al., 2022: pop est) | 23 | IVW | 0.015 | 0.072 | 8.33E-01 | -0.130 | 0.160 |
| Wellbeing (Howe et al., 2022: pop est) | Cognition (Howe et al., 2022: pop est) | 23 | Weighted mode | -0.038 | 0.181 | 8.36E-01 | -0.390 | 0.320 |
| Wellbeing (Howe et al., 2022: sib est) | Cognition (Howe et al., 2022: sib est) | 6 | MR Egger | -0.693 | 0.413 | 1.68E-01 | -1.500 | 0.120 |
| Wellbeing (Howe et al., 2022: sib est) | Cognition (Howe et al., 2022: sib est) | 6 | Weighted median | -0.065 | 0.163 | 6.91E-01 | -0.390 | 0.260 |
| Wellbeing (Howe et al., 2022: sib est) | Cognition (Howe et al., 2022: sib est) | 6 | IVW | -0.093 | 0.141 | 5.09E-01 | -0.370 | 0.180 |

| | | | | | | | | |
|---|---|---|---|---|---|---|---|---|
| Wellbeing (Howe et al., 2022: sib est) | Cognition (Howe et al., 2022: sib est) | 6 | Weighted mode | 0.018 | 0.250 | 9.44E-01 | -0.470 | 0.510 |
| Cognition (current study) | Depression (current study) | 227 | IVW | -0.127 | 0.031 | 3.15E-05 | -0.187 | -0.067 |
| Cognition (current study) | Depression (current study) | 227 | MR Egger | -0.177 | 0.133 | 1.85E-01 | -0.439 | 0.084 |
| Cognition (current study) | Depression (current study) | 227 | Weighted median | -0.086 | 0.025 | 6.42E-04 | -0.135 | -0.036 |
| Cognition (current study) | Depression (current study) | 227 | Weighted mode | -0.056 | 0.072 | 4.34E-01 | -0.198 | 0.085 |
| Cognition (Howe et al., 2022: pop est) | Depression (Howe et al., 2022: pop est) | 23 | MR Egger | 0.085 | 0.087 | 3.41E-01 | -0.090 | 0.260 |
| Cognition (Howe et al., 2022: pop est) | Depression (Howe et al., 2022: pop est) | 23 | Weighted median | -0.012 | 0.042 | 7.69E-01 | -0.090 | 0.070 |
| Cognition (Howe et al., 2022: pop est) | Depression (Howe et al., 2022: pop est) | 23 | IVW | -0.035 | 0.030 | 2.51E-01 | -0.090 | 0.020 |
| Cognition (Howe et al., 2022: pop est) | Depression (Howe et al., 2022: pop est) | 23 | Weighted mode | -0.003 | 0.077 | 9.68E-01 | -0.150 | 0.150 |
| Cognition (Howe et al., 2022: sib est) | Depression (Howe et al., 2022: sib est) | 11 | MR Egger | 0.023 | 0.179 | 8.99E-01 | -0.330 | 0.370 |
| Cognition (Howe et al., 2022: sib est) | Depression (Howe et al., 2022: sib est) | 11 | Weighted median | 0.072 | 0.075 | 3.37E-01 | -0.080 | 0.220 |
| Cognition (Howe et al., 2022: sib est) | Depression (Howe et al., 2022: sib est) | 11 | IVW | 0.053 | 0.059 | 3.70E-01 | -0.060 | 0.170 |

| | | | | | | | | |
|---|---|---|---|---|---|---|---|---|
| Cognition (Howe et al., 2022: sib est) | Depression (Howe et al., 2022: sib est) | 11 | Weighted mode | 0.124 | 0.130 | 3.64E-01 | -0.130 | 0.380 |
| Wellbeing (current study) | Cognition (current study) | 157 | IVW | 0.297 | 0.084 | 3.96E-04 | 0.133 | 0.462 |
| Wellbeing (current study) | Cognition (current study) | 157 | MR Egger | 0.853 | 0.384 | 2.79E-02 | 0.100 | 1.606 |
| Wellbeing (current study) | Cognition (current study) | 157 | Weighted median | 0.391 | 0.061 | 1.52E-10 | 0.271 | 0.511 |
| Wellbeing (current study) | Cognition (current study) | 157 | Weighted mode | 0.540 | 0.166 | 1.38E-03 | 0.215 | 0.866 |
| Wellbeing (Howe et al., 2022: pop est) | Cognition (Howe et al., 2022: pop est) | 23 | MR Egger | -0.315 | 0.214 | 1.55E-01 | -0.730 | 0.100 |
| Wellbeing (Howe et al., 2022: pop est) | Cognition (Howe et al., 2022: pop est) | 23 | Weighted median | 0.029 | 0.101 | 7.74E-01 | -0.170 | 0.230 |
| Wellbeing (Howe et al., 2022: pop est) | Cognition (Howe et al., 2022: pop est) | 23 | IVW | 0.015 | 0.072 | 8.33E-01 | -0.130 | 0.160 |
| Wellbeing (Howe et al., 2022: pop est) | Cognition (Howe et al., 2022: pop est) | 23 | Weighted mode | -0.038 | 0.181 | 8.36E-01 | -0.390 | 0.320 |
| Wellbeing (Howe et al., 2022: sib est) | Cognition (Howe et al., 2022: sib est) | 6 | MR Egger | -0.693 | 0.413 | 1.68E-01 | -1.500 | 0.120 |
| Wellbeing (Howe et al., 2022: sib est) | Cognition (Howe et al., 2022: sib est) | 6 | Weighted median | -0.065 | 0.163 | 6.91E-01 | -0.390 | 0.260 |
| Wellbeing (Howe et al., 2022: sib est) | Cognition (Howe et al., 2022: sib est) | 6 | IVW | -0.093 | 0.141 | 5.09E-01 | -0.370 | 0.180 |

| | | | | | | | | |
|---|---|---|---|---|---|---|---|---|
| Wellbeing (Howe et al., 2022: sib est) | Cognition (Howe et al., 2022: sib est) | 6 | Weighted mode | 0.018 | 0.250 | 9.44E-01 | -0.470 | 0.510 |

***Table S18. Tissue Specificity of Prioritised Genes from FUMA (using GTEx v8 54 Tissue Types): PANAS Positive.***

| Category | GeneSet | N_genes | N_overlap | p | adjP | genes |
|---|---|---|---|---|---|---|
| DEG.up | Adipose_Subcutaneous | 1689 | 0 | 1.000 | 1.000 | |
| DEG.up | Adipose_Visceral_Omentum | 1428 | 0 | 1.000 | 1.000 | |
| DEG.up | Adrenal_Gland | 1204 | 2 | 0.316 | 1.000 | ENSG00000084628:ENSG00000121769 |
| DEG.up | Artery_Aorta | 2103 | 1 | 0.879 | 1.000 | ENSG00000121769 |
| DEG.up | Artery_Coronary | 1593 | 2 | 0.452 | 1.000 | ENSG00000121769:ENSG00000054938 |
| DEG.up | Artery_Tibial | 2175 | 2 | 0.627 | 1.000 | ENSG00000121769:ENSG00000054938 |
| DEG.up | Bladder | 728 | 1 | 0.505 | 1.000 | ENSG00000077514 |
| DEG.up | Brain_Amygdala | 1438 | 5 | 0.009 | 0.479 | ENSG00000204624:ENSG00000084628:ENSG00000157103:ENSG00000145335:ENSG00000181790 |
| DEG.up | Brain_Anterior_cingulate_cortex_BA24 | 1866 | 5 | 0.025 | 1.000 | ENSG00000204624:ENSG00000121769:ENSG00000157103:ENSG00000145335:ENSG00000181790 |
| DEG.up | Brain_Caudate_basal_ganglia | 1703 | 5 | 0.018 | 0.957 | ENSG00000204624:ENSG00000084628:ENSG00000157103:ENSG00000145335:ENSG00000181790 |
| DEG.up | Brain_Cerebellar_Hemisphere | 4248 | 5 | 0.373 | 1.000 | ENSG00000204624:ENSG00000084628:ENSG00000157103:ENSG00000145335:ENSG00000181790 |
| DEG.up | Brain_Cerebellum | 4055 | 5 | 0.332 | 1.000 | ENSG00000204624:ENSG00000084628:ENSG00000157103:ENSG00000145335:ENSG00000181790 |
| DEG.up | Brain_Cortex | 2159 | 6 | 0.011 | 0.612 | ENSG00000204624:ENSG00000084628:ENSG00000121769:ENSG00000157103:ENSG00000145335:ENSG00000181790 |
| DEG.up | Brain_Frontal_Cortex_BA9 | 2422 | 6 | 0.019 | 1.000 | ENSG00000204624:ENSG00000084628:ENSG00000121769:ENSG00000157103:ENSG00000145335:ENSG00000181790 |

| | | | | | | |
|---|---|---|---|---|---|---|
| DEG.up | Brain_Hippocampus | 1542 | 6 | 0.002 | 0.114 | ENSG00000204624:ENSG00000084628:ENSG00000121769:ENSG00000157103:ENSG00000145335:ENSG00000181790 |
| DEG.up | Brain_Hypothalamus | 1979 | 5 | 0.032 | 1.000 | ENSG00000204624:ENSG00000084628:ENSG00000157103:ENSG00000145335:ENSG00000181790 |
| DEG.up | Brain_Nucleus_accumbens_basal_ganglia | 1834 | 5 | 0.024 | 1.000 | ENSG00000204624:ENSG00000084628:ENSG00000157103:ENSG00000145335:ENSG00000181790 |
| DEG.up | Brain_Putamen_basal_ganglia | 1346 | 5 | 0.007 | 0.363 | ENSG00000204624:ENSG00000084628:ENSG00000157103:ENSG00000145335:ENSG00000181790 |
| DEG.up | Brain_Spinal_cord_cervical_c-1 | 1611 | 3 | 0.191 | 1.000 | ENSG00000084628:ENSG00000157103:ENSG00000145335 |
| DEG.up | Brain_Substantia_nigra | 1277 | 6 | 0.001 | 0.043 | ENSG00000204624:ENSG00000084628:ENSG00000121769:ENSG00000157103:ENSG00000145335:ENSG00000181790 |
| DEG.up | Breast_Mammary_Tissue | 1428 | 1 | 0.755 | 1.000 | ENSG00000168528 |
| DEG.up | Cells_Cultured_fibroblasts | 3686 | 4 | 0.471 | 1.000 | ENSG00000060688:ENSG00000168528:ENSG00000197081:ENSG00000077514 |
| DEG.up | Cells_EBV-transformed_lymphocytes | 3833 | 3 | 0.735 | 1.000 | ENSG00000060688:ENSG00000197081:ENSG00000077514 |
| DEG.up | Cervix_Ectocervix | 187 | 0 | 1.000 | 1.000 | |
| DEG.up | Cervix_Endocervix | 1723 | 0 | 1.000 | 1.000 | |
| DEG.up | Colon_Sigmoid | 1264 | 3 | 0.113 | 1.000 | ENSG00000068781:ENSG00000242441:ENSG00000054938 |
| DEG.up | Colon_Transverse | 1128 | 4 | 0.019 | 1.000 | ENSG00000168528:ENSG00000068781:ENSG00000242441:ENSG00000054938 |
| DEG.up | Esophagus_Gastroesophageal_Junction | 1016 | 4 | 0.013 | 0.719 | ENSG00000068781:ENSG00000242441:ENSG00000138039:ENSG00000054938 |
| DEG.up | Esophagus_Mucosa | 1755 | 1 | 0.825 | 1.000 | ENSG00000168528 |

| | | | | | | |
|---|---|---|---|---|---|---|
| DEG.up | Esophagus_Muscularis | 1043 | 4 | 0.015 | 0.786 | ENSG00000068781:ENSG00000242441:ENSG00000138039:ENSG00000054938 |
| DEG.up | Fallopian_Tube | 451 | 0 | 1.000 | 1.000 | |
| DEG.up | Heart_Atrial_Appendage | 572 | 2 | 0.100 | 1.000 | ENSG00000121769:ENSG00000054938 |
| DEG.up | Heart_Left_Ventricle | 395 | 1 | 0.314 | 1.000 | ENSG00000121769 |
| DEG.up | Kidney_Cortex | 948 | 2 | 0.225 | 1.000 | ENSG00000121769:ENSG00000168528 |
| DEG.up | Kidney_Medulla | 93 | 2 | 0.003 | 0.184 | ENSG00000121769:ENSG00000168528 |
| DEG.up | Liver | 1090 | 2 | 0.275 | 1.000 | ENSG00000168528:ENSG00000157103 |
| DEG.up | Lung | 3029 | 2 | 0.807 | 1.000 | ENSG00000168528:ENSG00000197081 |
| DEG.up | Minor_Salivary_Gland | 1285 | 1 | 0.716 | 1.000 | ENSG00000168528 |
| DEG.up | Muscle_Skeletal | 879 | 4 | 0.008 | 0.436 | ENSG00000084628:ENSG00000121769:ENSG00000168528:ENSG00000197081 |
| DEG.up | Nerve_Tibial | 4157 | 4 | 0.577 | 1.000 | ENSG00000060688:ENSG00000138039:ENSG00000157103:ENSG00000145335 |
| DEG.up | Ovary | 3674 | 1 | 0.979 | 1.000 | ENSG00000138039 |
| DEG.up | Pancreas | 549 | 0 | 1.000 | 1.000 | |
| DEG.up | Pituitary | 3253 | 2 | 0.840 | 1.000 | ENSG00000204624:ENSG00000181790 |
| DEG.up | Prostate | 2019 | 2 | 0.584 | 1.000 | ENSG00000168528:ENSG00000054938 |
| DEG.up | Skin_Not_Sun_Exposed_Suprapubic | 2037 | 1 | 0.870 | 1.000 | ENSG00000168528 |
| DEG.up | Skin_Sun_Exposed_Lower_leg | 2163 | 1 | 0.887 | 1.000 | ENSG00000168528 |
| DEG.up | Small_Intestine_Terminal_Ileum | 1855 | 2 | 0.535 | 1.000 | ENSG00000168528:ENSG00000054938 |
| DEG.up | Spleen | 2666 | 1 | 0.935 | 1.000 | ENSG00000197081 |
| DEG.up | Stomach | 780 | 3 | 0.035 | 1.000 | ENSG00000168528:ENSG00000068781:ENSG00000054938 |
| DEG.up | Testis | 5947 | 5 | 0.710 | 1.000 | ENSG00000204624:ENSG00000060688:ENSG00000242441:ENSG00000077514:ENSG00000204952 |
| DEG.up | Thyroid | 3732 | 1 | 0.981 | 1.000 | ENSG00000168528 |
| DEG.up | Uterus | 3999 | 7 | 0.063 | 1.000 | ENSG00000060688:ENSG00000121769:ENSG00000068781:ENSG00000242441:ENSG000 |

| | | | | | | |
|---|---|---|---|---|---|---|
| | | | | | | 00145335:ENSG00000077514:ENSG00000054938 |
| DEG.up | Vagina | 2013 | 2 | 0.582 | 1.000 | ENSG00000168528:ENSG00000054938 |
| DEG.up | Whole_Blood | 1299 | 2 | 0.350 | 1.000 | ENSG00000145335:ENSG00000197081 |
| DEG.down | Adipose_Subcutaneous | 1307 | 4 | 0.031 | 1.000 | ENSG00000168528:ENSG00000157103:ENSG00000145335:ENSG00000054938 |
| DEG.down | Adipose_Visceral_Omentum | 1400 | 4 | 0.038 | 1.000 | ENSG00000121769:ENSG00000168528:ENSG00000157103:ENSG00000181790 |
| DEG.down | Adrenal_Gland | 2994 | 2 | 0.801 | 1.000 | ENSG00000145335:ENSG00000077514 |
| DEG.down | Artery_Aorta | 1323 | 4 | 0.032 | 1.000 | ENSG00000168528:ENSG00000157103:ENSG00000145335:ENSG00000181790 |
| DEG.down | Artery_Coronary | 863 | 3 | 0.045 | 1.000 | ENSG00000168528:ENSG00000145335:ENSG00000181790 |
| DEG.down | Artery_Tibial | 1746 | 2 | 0.501 | 1.000 | ENSG00000168528:ENSG00000145335 |
| DEG.down | Bladder | 103 | 1 | 0.093 | 1.000 | ENSG00000181790 |
| DEG.down | Brain_Amygdala | 6995 | 3 | 0.985 | 1.000 | ENSG00000060688:ENSG00000197081:ENSG00000077514 |
| DEG.down | Brain_Anterior_cingulate_cortex_BA24 | 6003 | 3 | 0.956 | 1.000 | ENSG00000060688:ENSG00000197081:ENSG00000077514 |
| DEG.down | Brain_Caudate_basal_ganglia | 6174 | 3 | 0.963 | 1.000 | ENSG00000060688:ENSG00000197081:ENSG00000077514 |
| DEG.down | Brain_Cerebellar_Hemisphere | 2381 | 1 | 0.911 | 1.000 | ENSG00000197081 |
| DEG.down | Brain_Cerebellum | 2483 | 2 | 0.702 | 1.000 | ENSG00000168528:ENSG00000197081 |
| DEG.down | Brain_Cortex | 4957 | 3 | 0.887 | 1.000 | ENSG00000060688:ENSG00000197081:ENSG00000077514 |
| DEG.down | Brain_Frontal_Cortex_BA9 | 4429 | 3 | 0.827 | 1.000 | ENSG00000060688:ENSG00000197081:ENSG00000077514 |
| DEG.down | Brain_Hippocampus | 6806 | 4 | 0.933 | 1.000 | ENSG00000060688:ENSG00000168528:ENSG00000197081:ENSG00000077514 |
| DEG.down | Brain_Hypothalamus | 5303 | 4 | 0.785 | 1.000 | ENSG00000060688:ENSG00000168528:ENSG00000197081:ENSG00000077514 |

| | | | | | | |
|---|---|---|---|---|---|---|
| DEG.down | Brain_Nucleus_accumbens_basal_ganglia | 5801 | 3 | 0.947 | 1.000 | ENSG00000060688:ENSG00000197081:ENSG00000077514 |
| DEG.down | Brain_Putamen_basal_ganglia | 7081 | 3 | 0.986 | 1.000 | ENSG00000060688:ENSG00000197081:ENSG00000077514 |
| DEG.down | Brain_Spinal_cord_cervical_c-1 | 4547 | 3 | 0.842 | 1.000 | ENSG00000168528:ENSG00000197081:ENSG00000077514 |
| DEG.down | Brain_Substantia_nigra | 6489 | 4 | 0.912 | 1.000 | ENSG00000060688:ENSG00000168528:ENSG00000197081:ENSG00000077514 |
| DEG.down | Breast_Mammary_Tissue | 912 | 5 | 0.001 | 0.066 | ENSG00000121769:ENSG00000157103:ENSG00000145335:ENSG00000181790:ENSG00000054938 |
| DEG.down | Cells_Cultured_fibroblasts | 1713 | 1 | 0.818 | 1.000 | ENSG00000121769 |
| DEG.down | Cells_EBV-transformed_lymphocytes | 2063 | 0 | 1.000 | 1.000 | |
| DEG.down | Cervix_Ectocervix | 2 | 0 | 1.000 | 1.000 | |
| DEG.down | Cervix_Endocervix | 5 | 0 | 1.000 | 1.000 | |
| DEG.down | Colon_Sigmoid | 1530 | 3 | 0.171 | 1.000 | ENSG00000168528:ENSG00000157103:ENSG00000181790 |
| DEG.down | Colon_Transverse | 1404 | 4 | 0.039 | 1.000 | ENSG00000121769:ENSG00000157103:ENSG00000145335:ENSG00000181790 |
| DEG.down | Esophagus_Gastroesophageal_Junction | 1406 | 4 | 0.039 | 1.000 | ENSG00000168528:ENSG00000157103:ENSG00000145335:ENSG00000181790 |
| DEG.down | Esophagus_Mucosa | 3122 | 4 | 0.339 | 1.000 | ENSG00000121769:ENSG00000242441:ENSG00000145335:ENSG00000181790 |
| DEG.down | Esophagus_Muscularis | 1582 | 4 | 0.056 | 1.000 | ENSG00000168528:ENSG00000157103:ENSG00000145335:ENSG00000181790 |
| DEG.down | Fallopian_Tube | 1 | 0 | 1.000 | 1.000 | |
| DEG.down | Heart_Atrial_Appendage | 7365 | 6 | 0.761 | 1.000 | ENSG00000060688:ENSG00000168528:ENSG00000157103:ENSG00000145335:ENSG00000197081:ENSG00000077514 |

| | | | | | | |
|---|---|---|---|---|---|---|
| DEG.down | Heart_Left_Ventricle | 8514 | 6 | 0.893 | 1.000 | ENSG00000060688:ENSG00000168528:ENSG00000157103:ENSG00000145335:ENSG00000197081:ENSG00000077514 |
| DEG.down | Kidney_Cortex | 5049 | 5 | 0.542 | 1.000 | ENSG00000060688:ENSG00000242441:ENSG00000157103:ENSG00000145335:ENSG00000197081 |
| DEG.down | Kidney_Medulla | 0 | 0 | 1.000 | 1.000 | |
| DEG.down | Liver | 7196 | 5 | 0.874 | 1.000 | ENSG00000060688:ENSG00000121766:ENSG00000121769:ENSG00000197081:ENSG00000077514 |
| DEG.down | Lung | 820 | 2 | 0.180 | 1.000 | ENSG00000145335:ENSG00000181790 |
| DEG.down | Minor_Salivary_Gland | 1666 | 2 | 0.476 | 1.000 | ENSG00000121769:ENSG00000181790 |
| DEG.down | Muscle_Skeletal | 6093 | 4 | 0.879 | 1.000 | ENSG00000060688:ENSG00000121766:ENSG00000157103:ENSG00000077514 |
| DEG.down | Nerve_Tibial | 784 | 2 | 0.168 | 1.000 | ENSG00000121769:ENSG00000168528 |
| DEG.down | Ovary | 1429 | 2 | 0.396 | 1.000 | ENSG00000168528:ENSG00000181790 |
| DEG.down | Pancreas | 8671 | 4 | 0.990 | 1.000 | ENSG00000060688:ENSG00000121766:ENSG00000121769:ENSG00000197081 |
| DEG.down | Pituitary | 1752 | 3 | 0.226 | 1.000 | ENSG00000121769:ENSG00000242441:ENSG00000197081 |
| DEG.down | Prostate | 698 | 1 | 0.490 | 1.000 | ENSG00000181790 |
| DEG.down | Skin_Not_Sun_Exposed_Suprapubic | 2330 | 2 | 0.666 | 1.000 | ENSG00000121769:ENSG00000157103 |
| DEG.down | Skin_Sun_Exposed_Lower_leg | 2123 | 3 | 0.325 | 1.000 | ENSG00000121769:ENSG00000157103:ENSG00000145335 |
| DEG.down | Small_Intestine_Terminal_Ileum | 1257 | 3 | 0.111 | 1.000 | ENSG00000121769:ENSG00000157103:ENSG00000145335 |
| DEG.down | Spleen | 1930 | 3 | 0.272 | 1.000 | ENSG00000168528:ENSG00000157103:ENSG00000145335 |
| DEG.down | Stomach | 2290 | 2 | 0.656 | 1.000 | ENSG00000145335:ENSG00000181790 |
| DEG.down | Testis | 2599 | 1 | 0.930 | 1.000 | ENSG00000145335 |

| | | | | | | |
|---|---|---|---|---|---|---|
| DEG.down | Thyroid | 976 | 5 | 0.002 | 0.090 | ENSG00000121769:ENSG00000242441:ENSG00000157103:ENSG00000145335:ENSG00000181790 |
| DEG.down | Uterus | 788 | 1 | 0.533 | 1.000 | ENSG00000168528 |
| DEG.down | Vagina | 598 | 0 | 1.000 | 1.000 | |
| DEG.down | Whole_Blood | 6321 | 4 | 0.899 | 1.000 | ENSG00000060688:ENSG00000121766:ENSG00000121769:ENSG00000168528 |
| DEG.twoside | Adipose_Subcutaneous | 2996 | 4 | 0.310 | 1.000 | ENSG00000168528:ENSG00000157103:ENSG00000145335:ENSG00000054938 |
| DEG.twoside | Adipose_Visceral_Omentum | 2828 | 4 | 0.272 | 1.000 | ENSG00000121769:ENSG00000168528:ENSG00000157103:ENSG00000181790 |
| DEG.twoside | Adrenal_Gland | 4198 | 4 | 0.585 | 1.000 | ENSG00000084628:ENSG00000121769:ENSG00000145335:ENSG00000077514 |
| DEG.twoside | Artery_Aorta | 3426 | 5 | 0.210 | 1.000 | ENSG00000121769:ENSG00000168528:ENSG00000157103:ENSG00000145335:ENSG00000181790 |
| DEG.twoside | Artery_Coronary | 2456 | 5 | 0.071 | 1.000 | ENSG00000121769:ENSG00000168528:ENSG00000145335:ENSG00000181790:ENSG00000054938 |
| DEG.twoside | Artery_Tibial | 3921 | 4 | 0.525 | 1.000 | ENSG00000121769:ENSG00000168528:ENSG00000145335:ENSG00000054938 |
| DEG.twoside | Bladder | 831 | 2 | 0.184 | 1.000 | ENSG00000181790:ENSG00000077514 |
| DEG.twoside | Brain_Amygdala | 8433 | 8 | 0.587 | 1.000 | ENSG00000204624:ENSG00000084628:ENSG00000060688:ENSG00000157103:ENSG00000145335:ENSG00000197081:ENSG00000181790:ENSG00000077514 |
| DEG.twoside | Brain_Anterior_cingulate_cortex_BA24 | 7869 | 8 | 0.483 | 1.000 | ENSG00000204624:ENSG00000060688:ENSG00000121769:ENSG00000157103:ENSG00000145335:ENSG00000197081:ENSG00000181790:ENSG00000077514 |
| DEG.twoside | Brain_Caudate_basal_ganglia | 7877 | 8 | 0.484 | 1.000 | ENSG00000204624:ENSG00000084628:ENSG00000060688:ENSG00000157103:ENSG000 |

| | | | | | | |
|---|---|---|---|---|---|---|
| | | | | | | 00145335:ENSG00000197081:ENSG0000018 1790:ENSG00000077514 |
| DEG.twoside | Brain_Cerebellar_Hemisphere | 6629 | 6 | 0.641 | 1.000 | ENSG00000204624:ENSG00000084628:ENS G00000157103:ENSG00000145335:ENSG000 00197081:ENSG00000181790 |
| DEG.twoside | Brain_Cerebellum | 6538 | 7 | 0.426 | 1.000 | ENSG00000204624:ENSG00000084628:ENS G00000168528:ENSG00000157103:ENSG000 00145335:ENSG00000197081:ENSG0000018 1790 |
| DEG.twoside | Brain_Cortex | 7116 | 9 | 0.188 | 1.000 | ENSG00000204624:ENSG00000084628:ENS G00000060688:ENSG00000121769:ENSG000 00157103:ENSG00000145335:ENSG0000019 7081:ENSG00000181790:ENSG00000077514 |
| DEG.twoside | Brain_Frontal_Cortex_BA9 | 6851 | 9 | 0.155 | 1.000 | ENSG00000204624:ENSG00000084628:ENS G00000060688:ENSG00000121769:ENSG000 00157103:ENSG00000145335:ENSG0000019 7081:ENSG00000181790:ENSG00000077514 |
| DEG.twoside | Brain_Hippocampus | 8348 | 10 | 0.216 | 1.000 | ENSG00000204624:ENSG00000084628:ENS G00000060688:ENSG00000121769:ENSG000 00168528:ENSG00000157103:ENSG0000014 5335:ENSG00000197081:ENSG00000181790: ENSG00000077514 |
| DEG.twoside | Brain_Hypothalamus | 7282 | 9 | 0.210 | 1.000 | ENSG00000204624:ENSG00000084628:ENS G00000060688:ENSG00000168528:ENSG000 00157103:ENSG00000145335:ENSG0000019 7081:ENSG00000181790:ENSG00000077514 |
| DEG.twoside | Brain_Nucleus_accumbens_b asal_ganglia | 7635 | 8 | 0.439 | 1.000 | ENSG00000204624:ENSG00000084628:ENS G00000060688:ENSG00000157103:ENSG000 00145335:ENSG00000197081:ENSG0000018 1790:ENSG00000077514 |
| DEG.twoside | Brain_Putamen_basal_ganglia | 8427 | 8 | 0.586 | 1.000 | ENSG00000204624:ENSG00000084628:ENS G00000060688:ENSG00000157103:ENSG000 |

| | | | | | | |
|---|---|---|---|---|---|---|
| | | | | | | 00145335:ENSG00000197081:ENSG00000181790:ENSG00000077514 |
| DEG.twoside | Brain_Spinal_cord_cervical_c-1 | 6158 | 6 | 0.553 | 1.000 | ENSG00000084628:ENSG00000168528:ENSG00000157103:ENSG00000145335:ENSG00000197081:ENSG00000077514 |
| DEG.twoside | Brain_Substantia_nigra | 7766 | 10 | 0.145 | 1.000 | ENSG00000204624:ENSG00000084628:ENSG00000060688:ENSG00000121769:ENSG00000168528:ENSG00000157103:ENSG00000145335:ENSG00000197081:ENSG00000181790:ENSG00000077514 |
| DEG.twoside | Breast_Mammary_Tissue | 2340 | 6 | 0.017 | 0.895 | ENSG00000121769:ENSG00000168528:ENSG00000157103:ENSG00000145335:ENSG00000181790:ENSG00000054938 |
| DEG.twoside | Cells_Cultured_fibroblasts | 5399 | 5 | 0.611 | 1.000 | ENSG00000060688:ENSG00000121769:ENSG00000168528:ENSG00000197081:ENSG00000077514 |
| DEG.twoside | Cells_EBV-transformed_lymphocytes | 5896 | 3 | 0.951 | 1.000 | ENSG00000060688:ENSG00000197081:ENSG00000077514 |
| DEG.twoside | Cervix_Ectocervix | 189 | 0 | 1.000 | 1.000 | |
| DEG.twoside | Cervix_Endocervix | 1728 | 0 | 1.000 | 1.000 | |
| DEG.twoside | Colon_Sigmoid | 2794 | 6 | 0.037 | 1.000 | ENSG00000168528:ENSG00000068781:ENSG00000242441:ENSG00000157103:ENSG00000181790:ENSG00000054938 |
| DEG.twoside | Colon_Transverse | 2532 | 8 | 0.001 | 0.061 | ENSG00000121769:ENSG00000168528:ENSG00000068781:ENSG00000242441:ENSG00000157103:ENSG00000145335:ENSG00000181790:ENSG00000054938 |
| DEG.twoside | Esophagus_Gastroesophageal_Junction | 2422 | 8 | 0.001 | 0.045 | ENSG00000168528:ENSG00000068781:ENSG00000242441:ENSG00000138039:ENSG00000157103:ENSG00000145335:ENSG00000181790:ENSG00000054938 |

| | | | | | | |
|---|---|---|---|---|---|---|
| DEG.twoside | Esophagus_Mucosa | 4877 | 5 | 0.506 | 1.000 | ENSG00000121769:ENSG00000168528:ENSG00000242441:ENSG00000145335:ENSG00000181790 |
| DEG.twoside | Esophagus_Muscularis | 2625 | 8 | 0.001 | 0.077 | ENSG00000168528:ENSG00000068781:ENSG00000242441:ENSG00000138039:ENSG00000157103:ENSG00000145335:ENSG00000181790:ENSG00000054938 |
| DEG.twoside | Fallopian_Tube | 452 | 0 | 1.000 | 1.000 | |
| DEG.twoside | Heart_Atrial_Appendage | 7937 | 8 | 0.496 | 1.000 | ENSG00000060688:ENSG00000121769:ENSG00000168528:ENSG00000157103:ENSG00000145335:ENSG00000197081:ENSG00000077514:ENSG00000054938 |
| DEG.twoside | Heart_Left_Ventricle | 8909 | 7 | 0.823 | 1.000 | ENSG00000060688:ENSG00000121769:ENSG00000168528:ENSG00000157103:ENSG00000145335:ENSG00000197081:ENSG00000077514 |
| DEG.twoside | Kidney_Cortex | 5997 | 7 | 0.326 | 1.000 | ENSG00000060688:ENSG00000121769:ENSG00000168528:ENSG00000242441:ENSG00000157103:ENSG00000145335:ENSG00000197081 |
| DEG.twoside | Kidney_Medulla | 93 | 2 | 0.003 | 0.184 | ENSG00000121769:ENSG00000168528 |
| DEG.twoside | Liver | 8286 | 7 | 0.739 | 1.000 | ENSG00000060688:ENSG00000121766:ENSG00000121769:ENSG00000168528:ENSG00000157103:ENSG00000197081:ENSG00000077514 |
| DEG.twoside | Lung | 3849 | 4 | 0.509 | 1.000 | ENSG00000168528:ENSG00000145335:ENSG00000197081:ENSG00000181790 |
| DEG.twoside | Minor_Salivary_Gland | 2951 | 3 | 0.545 | 1.000 | ENSG00000121769:ENSG00000168528:ENSG00000181790 |
| DEG.twoside | Muscle_Skeletal | 6972 | 8 | 0.320 | 1.000 | ENSG00000084628:ENSG00000060688:ENSG00000121766:ENSG00000121769:ENSG00000168528:ENSG00000157103:ENSG00000197081:ENSG00000077514 |

| | | | | | | |
|---|---|---|---|---|---|---|
| DEG.twoside | Nerve_Tibial | 4941 | 6 | 0.314 | 1.000 | ENSG00000060688:ENSG00000121769:ENSG00000168528:ENSG00000138039:ENSG00000157103:ENSG00000145335 |
| DEG.twoside | Ovary | 5103 | 3 | 0.900 | 1.000 | ENSG00000168528:ENSG00000138039:ENSG00000181790 |
| DEG.twoside | Pancreas | 9220 | 4 | 0.995 | 1.000 | ENSG00000060688:ENSG00000121766:ENSG00000121769:ENSG00000197081 |
| DEG.twoside | Pituitary | 5005 | 5 | 0.533 | 1.000 | ENSG00000204624:ENSG00000121769:ENSG00000242441:ENSG00000197081:ENSG00000181790 |
| DEG.twoside | Prostate | 2717 | 3 | 0.485 | 1.000 | ENSG00000168528:ENSG00000181790:ENSG00000054938 |
| DEG.twoside | Skin_Not_Sun_Exposed_Suprapubic | 4367 | 3 | 0.819 | 1.000 | ENSG00000121769:ENSG00000168528:ENSG00000157103 |
| DEG.twoside | Skin_Sun_Exposed_Lower_leg | 4286 | 4 | 0.604 | 1.000 | ENSG00000121769:ENSG00000168528:ENSG00000157103:ENSG00000145335 |
| DEG.twoside | Small_Intestine_Terminal_Ileum | 3112 | 5 | 0.157 | 1.000 | ENSG00000121769:ENSG00000168528:ENSG00000157103:ENSG00000145335:ENSG00000054938 |
| DEG.twoside | Spleen | 4596 | 4 | 0.666 | 1.000 | ENSG00000168528:ENSG00000157103:ENSG00000145335:ENSG00000197081 |
| DEG.twoside | Stomach | 3070 | 5 | 0.150 | 1.000 | ENSG00000168528:ENSG00000068781:ENSG00000145335:ENSG00000181790:ENSG00000054938 |
| DEG.twoside | Testis | 8546 | 6 | 0.895 | 1.000 | ENSG00000204624:ENSG00000060688:ENSG00000242441:ENSG00000145335:ENSG00000077514:ENSG00000204952 |
| DEG.twoside | Thyroid | 4708 | 6 | 0.271 | 1.000 | ENSG00000121769:ENSG00000168528:ENSG00000242441:ENSG00000157103:ENSG00000145335:ENSG00000181790 |
| DEG.twoside | Uterus | 4787 | 8 | 0.056 | 1.000 | ENSG00000060688:ENSG00000121769:ENSG00000168528:ENSG00000068781:ENSG000 |

| | | | | | | |
|---|---|---|---|---|---|---|
| | | | | | | 00242441:ENSG00000145335:ENSG00000077514:ENSG00000054938 |
| DEG.twoside | Vagina | 2611 | 2 | 0.730 | 1.000 | ENSG00000168528:ENSG00000054938 |
| DEG.twoside | Whole_Blood | 7620 | 6 | 0.796 | 1.000 | ENSG00000060688:ENSG00000121766:ENSG00000121769:ENSG00000168528:ENSG00000145335:ENSG00000197081 |

***Table S19. Tissue Specificity of Prioritised Genes from FUMA (using GTEx v8 54 Tissue Types): PANAS Negative***

| Category | GeneSet | N_genes | N_overlap | p | adjP | genes |
|---|---|---|---|---|---|---|
| DEG.up | Adipose_Subcutaneous | 1689 | 1 | 0.998 | 1.000 | ENSG00000181826 |
| DEG.up | Adipose_Visceral_Omentum | 1428 | 2 | 0.960 | 1.000 | ENSG00000181826:ENSG00000111801 |
| DEG.up | Adrenal_Gland | 1204 | 2 | 0.922 | 1.000 | ENSG00000075914:ENSG00000111801 |
| DEG.up | Artery_Aorta | 2103 | 4 | 0.937 | 1.000 | ENSG00000198626:ENSG00000112763:ENSG00000158321:ENSG00000101452 |
| DEG.up | Artery_Coronary | 1593 | 3 | 0.916 | 1.000 | ENSG00000198626:ENSG00000112763:ENSG00000101452 |
| DEG.up | Artery_Tibial | 2175 | 5 | 0.875 | 1.000 | ENSG00000198626:ENSG00000112763:ENSG00000181315:ENSG00000198315:ENSG00000101452 |
| DEG.up | Bladder | 728 | 2 | 0.713 | 1.000 | ENSG00000181315:ENSG00000101452 |
| DEG.up | Brain_Amygdala | 1438 | 2 | 0.961 | 1.000 | ENSG00000164398:ENSG00000185046 |
| DEG.up | Brain_Anterior_cingulate_cortex_BA24 | 1866 | 3 | 0.959 | 1.000 | ENSG00000197410:ENSG00000164398:ENSG00000185046 |
| DEG.up | Brain_Caudate_basal_ganglia | 1703 | 3 | 0.937 | 1.000 | ENSG00000197410:ENSG00000164398:ENSG00000185046 |
| DEG.up | Brain_Cerebellar_Hemisphere | 4248 | 12 | 0.808 | 1.000 | ENSG00000198626:ENSG00000196345:ENSG00000144792:ENSG00000186448:ENSG00000158987:ENSG00000164398:ENSG00000112763:ENSG00000124613:ENSG00000196812:ENSG00000137185:ENSG00000137338:ENSG00000185046 |
| DEG.up | Brain_Cerebellum | 4055 | 12 | 0.750 | 1.000 | ENSG00000198626:ENSG00000196345:ENSG00000144792:ENSG00000186448:ENSG00000158987:ENSG00000164398:ENSG00000112763:ENSG00000124613:ENSG00000196812:ENSG00000137185:ENSG00000137338:ENSG00000185046 |

| | | | | | | |
|---|---|---|---|---|---|---|
| DEG.up | Brain_Cortex | 2159 | 5 | 0.871 | 1.000 | ENSG00000197410:ENSG00000164398:ENSG00000096654:ENSG00000171811:ENSG00000185046 |
| DEG.up | Brain_Frontal_Cortex_BA9 | 2422 | 5 | 0.927 | 1.000 | ENSG00000197410:ENSG00000158985:ENSG00000164398:ENSG00000096654:ENSG00000185046 |
| DEG.up | Brain_Hippocampus | 1542 | 2 | 0.972 | 1.000 | ENSG00000164398:ENSG00000185046 |
| DEG.up | Brain_Hypothalamus | 1979 | 3 | 0.970 | 1.000 | ENSG00000164398:ENSG00000171811:ENSG00000185046 |
| DEG.up | Brain_Nucleus_accumbens_basal_ganglia | 1834 | 3 | 0.956 | 1.000 | ENSG00000197410:ENSG00000164398:ENSG00000185046 |
| DEG.up | Brain_Putamen_basal_ganglia | 1346 | 2 | 0.949 | 1.000 | ENSG00000164398:ENSG00000185046 |
| DEG.up | Brain_Spinal_cord_cervical_c-1 | 1611 | 3 | 0.919 | 1.000 | ENSG00000181826:ENSG00000233822:ENSG00000185046 |
| DEG.up | Brain_Substantia_nigra | 1277 | 2 | 0.937 | 1.000 | ENSG00000164398:ENSG00000185046 |
| DEG.up | Breast_Mammary_Tissue | 1428 | 2 | 0.960 | 1.000 | ENSG00000181826:ENSG00000158321 |
| DEG.up | Cells_Cultured_fibroblasts | 3686 | 8 | 0.950 | 1.000 | ENSG00000196345:ENSG00000186448:ENSG00000075914:ENSG00000171566:ENSG00000146109:ENSG00000196787:ENSG00000187626:ENSG00000189134 |
| DEG.up | Cells_EBV-transformed_lymphocytes | 3833 | 17 | 0.137 | 1.000 | ENSG00000075914:ENSG00000171566:ENSG00000158985:ENSG00000158987:ENSG00000026950:ENSG00000111801:ENSG00000146109:ENSG00000124635:ENSG00000196787:ENSG00000096654:ENSG00000185130:ENSG00000196747:ENSG00000184357:ENSG00000197153:ENSG00000124657:ENSG00000196812:ENSG00000187626 |
| DEG.up | Cervix_Ectocervix | 187 | 0 | 1.000 | 1.000 | |
| DEG.up | Cervix_Endocervix | 1723 | 9 | 0.126 | 1.000 | ENSG00000075914:ENSG00000171566:ENSG00000026950:ENSG00000111801:ENSG00000112763:ENSG00000096654:ENSG00000198315:ENSG00000187626:ENSG00000101452 |

| | | | | | | |
|---|---|---|---|---|---|---|
| DEG.up | Colon_Sigmoid | 1264 | 4 | 0.629 | 1.000 | ENSG00000130940:ENSG00000197410:ENSG00000137338:ENSG00000101452 |
| DEG.up | Colon_Transverse | 1128 | 4 | 0.537 | 1.000 | ENSG00000130940:ENSG00000154274:ENSG00000197410:ENSG00000112812 |
| DEG.up | Esophagus_Gastroesophageal_Junction | 1016 | 1 | 0.971 | 1.000 | ENSG00000101452 |
| DEG.up | Esophagus_Mucosa | 1755 | 1 | 0.998 | 1.000 | ENSG00000112812 |
| DEG.up | Esophagus_Muscularis | 1043 | 2 | 0.876 | 1.000 | ENSG00000198626:ENSG00000101452 |
| DEG.up | Fallopian_Tube | 451 | 1 | 0.788 | 1.000 | ENSG00000112763 |
| DEG.up | Heart_Atrial_Appendage | 572 | 1 | 0.861 | 1.000 | ENSG00000198626 |
| DEG.up | Heart_Left_Ventricle | 395 | 1 | 0.742 | 1.000 | ENSG00000198626 |
| DEG.up | Kidney_Cortex | 948 | 3 | 0.630 | 1.000 | ENSG00000154274:ENSG00000171564:ENSG00000197279 |
| DEG.up | Kidney_Medulla | 93 | 0 | 1.000 | 1.000 | |
| DEG.up | Liver | 1090 | 2 | 0.891 | 1.000 | ENSG00000154274:ENSG00000171564 |
| DEG.up | Lung | 3029 | 9 | 0.719 | 1.000 | ENSG00000130940:ENSG00000181826:ENSG00000158985:ENSG00000026950:ENSG00000111801:ENSG00000112763:ENSG00000124635:ENSG00000112812:ENSG00000171811 |
| DEG.up | Minor_Salivary_Gland | 1285 | 5 | 0.443 | 1.000 | ENSG00000130940:ENSG00000154274:ENSG00000112812:ENSG00000196812:ENSG00000158321 |
| DEG.up | Muscle_Skeletal | 879 | 1 | 0.953 | 1.000 | ENSG00000171811 |
| DEG.up | Nerve_Tibial | 4157 | 10 | 0.924 | 1.000 | ENSG00000196345:ENSG00000144792:ENSG00000186448:ENSG00000163812:ENSG00000181826:ENSG00000158987:ENSG00000181315:ENSG00000198315:ENSG00000137338:ENSG00000158321 |
| DEG.up | Ovary | 3674 | 10 | 0.826 | 1.000 | ENSG00000196345:ENSG00000144792:ENSG00000186448:ENSG00000026950:ENSG00000111801:ENSG00000181315:ENSG0000012 |

| | | | | | | |
|---|---|---|---|---|---|---|
| | | | | | | 4613:ENSG00000198315:ENSG00000137185:ENSG00000158321 |
| DEG.up | Pancreas | 549 | 2 | 0.559 | 1.000 | ENSG00000154274:ENSG00000112812 |
| DEG.up | Pituitary | 3253 | 8 | 0.884 | 1.000 | ENSG00000196345:ENSG00000144792:ENSG00000111801:ENSG00000112812:ENSG00000096654:ENSG00000197279:ENSG00000137185:ENSG00000171811 |
| DEG.up | Prostate | 2019 | 7 | 0.535 | 1.000 | ENSG00000130940:ENSG00000144792:ENSG00000026950:ENSG00000124635:ENSG00000112812:ENSG00000196812:ENSG00000137185 |
| DEG.up | Skin_Not_Sun_Exposed_Suprapubic | 2037 | 6 | 0.702 | 1.000 | ENSG00000130940:ENSG00000163812:ENSG00000075914:ENSG00000146109:ENSG00000112812:ENSG00000158321 |
| DEG.up | Skin_Sun_Exposed_Lower_leg | 2163 | 6 | 0.757 | 1.000 | ENSG00000130940:ENSG00000163812:ENSG00000075914:ENSG00000146109:ENSG00000112812:ENSG00000158321 |
| DEG.up | Small_Intestine_Terminal_Ileum | 1855 | 7 | 0.442 | 1.000 | ENSG00000154274:ENSG00000158985:ENSG00000026950:ENSG00000111801:ENSG00000112812:ENSG00000184357:ENSG00000196812 |
| DEG.up | Spleen | 2666 | 11 | 0.288 | 1.000 | ENSG00000158985:ENSG00000026950:ENSG00000111801:ENSG00000112763:ENSG00000146109:ENSG00000124635:ENSG00000196787:ENSG00000196747:ENSG00000196812:ENSG00000137185:ENSG00000187626 |
| DEG.up | Stomach | 780 | 5 | 0.124 | 1.000 | ENSG00000130940:ENSG00000154274:ENSG00000197410:ENSG00000171564:ENSG00000112812 |
| DEG.up | Testis | 5947 | 19 | 0.670 | 1.000 | ENSG00000196345:ENSG00000144792:ENSG00000163812:ENSG00000075914:ENSG00000197410:ENSG00000158985:ENSG00000158987:ENSG00000164398:ENSG00000124635: |

| | | | | | | |
|---|---|---|---|---|---|---|
| | | | | | | ENSG00000196787:ENSG00000158553:ENSG00000124613:ENSG00000096654:ENSG00000233822:ENSG00000197279:ENSG00000137185:ENSG00000187626:ENSG00000189134:ENSG00000171811 |
| DEG.up | Thyroid | 3732 | 12 | 0.632 | 1.000 | ENSG00000196345:ENSG00000144792:ENSG00000186448:ENSG00000181826:ENSG00000158985:ENSG00000111801:ENSG00000112812:ENSG00000124613:ENSG00000196812:ENSG00000198315:ENSG00000137185:ENSG00000171811 |
| DEG.up | Uterus | 3999 | 17 | 0.180 | 1.000 | ENSG00000196345:ENSG00000144792:ENSG00000186448:ENSG00000154274:ENSG00000197410:ENSG00000171566:ENSG00000026950:ENSG00000111801:ENSG00000112763:ENSG00000146109:ENSG00000181315:ENSG00000124613:ENSG00000198315:ENSG00000137185:ENSG00000187626:ENSG00000137338:ENSG00000101452 |
| DEG.up | Vagina | 2013 | 4 | 0.921 | 1.000 | ENSG00000130940:ENSG00000026950:ENSG00000111801:ENSG00000124635 |
| DEG.up | Whole_Blood | 1299 | 4 | 0.651 | 1.000 | ENSG00000158985:ENSG00000026950:ENSG00000124635:ENSG00000196747 |
| DEG.down | Adipose_Subcutaneous | 1307 | 2 | 0.942 | 1.000 | ENSG00000130940:ENSG00000198626 |
| DEG.down | Adipose_Visceral_Omentum | 1400 | 3 | 0.863 | 1.000 | ENSG00000130940:ENSG00000112812:ENSG00000158321 |
| DEG.down | Adrenal_Gland | 2994 | 10 | 0.574 | 1.000 | ENSG00000130940:ENSG00000181826:ENSG00000158985:ENSG00000158987:ENSG00000164398:ENSG00000112812:ENSG00000197279:ENSG00000189134:ENSG00000158321:ENSG00000101452 |
| DEG.down | Artery_Aorta | 1323 | 0 | 1.000 | 1.000 | |
| DEG.down | Artery_Coronary | 863 | 1 | 0.950 | 1.000 | ENSG00000197279 |

| | | | | | | |
|---|---|---|---|---|---|---|
| DEG.down | Artery_Tibial | 1746 | 3 | 0.944 | 1.000 | ENSG00000026950:ENSG00000124635:ENSG00000197279 |
| DEG.down | Bladder | 103 | 0 | 1.000 | 1.000 | |
| DEG.down | Brain_Amygdala | 6995 | 21 | 0.800 | 1.000 | ENSG00000198626:ENSG00000196345:ENSG00000186448:ENSG00000163812:ENSG00000075914:ENSG00000163815:ENSG00000181826:ENSG00000171566:ENSG00000158987:ENSG00000026950:ENSG00000111801:ENSG00000112763:ENSG00000182952:ENSG00000146109:ENSG00000181315:ENSG00000124635:ENSG00000196812:ENSG00000198315:ENSG00000137185:ENSG00000187626:ENSG00000101452 |
| DEG.down | Brain_Anterior_cingulate_cortex_BA24 | 6003 | 20 | 0.585 | 1.000 | ENSG00000196345:ENSG00000186448:ENSG00000163812:ENSG00000075914:ENSG00000163815:ENSG00000181826:ENSG00000158987:ENSG00000026950:ENSG00000111801:ENSG00000112763:ENSG00000182952:ENSG00000146109:ENSG00000181315:ENSG00000112812:ENSG00000196812:ENSG00000198315:ENSG00000137185:ENSG00000187626:ENSG00000158321:ENSG00000101452 |
| DEG.down | Brain_Caudate_basal_ganglia | 6174 | 18 | 0.822 | 1.000 | ENSG00000196345:ENSG00000186448:ENSG00000163812:ENSG00000075914:ENSG00000181826:ENSG00000158987:ENSG00000026950:ENSG00000111801:ENSG00000112763:ENSG00000182952:ENSG00000146109:ENSG00000181315:ENSG00000197279:ENSG00000196812:ENSG00000198315:ENSG00000137185:ENSG00000187626:ENSG00000101452 |
| DEG.down | Brain_Cerebellar_Hemisphere | 2381 | 1 | 1.000 | 1.000 | ENSG00000163815 |
| DEG.down | Brain_Cerebellum | 2483 | 3 | 0.993 | 1.000 | ENSG00000163815:ENSG00000158985:ENSG00000197279 |

| | | | | | | |
|---|---|---|---|---|---|---|
| DEG.down | Brain_Cortex | 4957 | 16 | 0.639 | 1.000 | ENSG00000163812:ENSG00000075914:ENSG00000163815:ENSG00000181826:ENSG00000026950:ENSG00000111801:ENSG00000112763:ENSG00000182952:ENSG00000146109:ENSG00000181315:ENSG00000197279:ENSG00000196812:ENSG00000198315:ENSG00000137185:ENSG00000187626:ENSG00000101452 |
| DEG.down | Brain_Frontal_Cortex_BA9 | 4429 | 15 | 0.551 | 1.000 | ENSG00000163812:ENSG00000075914:ENSG00000163815:ENSG00000181826:ENSG00000026950:ENSG00000111801:ENSG00000112763:ENSG00000146109:ENSG00000197279:ENSG00000196812:ENSG00000198315:ENSG00000137185:ENSG00000187626:ENSG00000158321:ENSG00000101452 |
| DEG.down | Brain_Hippocampus | 6806 | 19 | 0.888 | 1.000 | ENSG00000196345:ENSG00000186448:ENSG00000163812:ENSG00000075914:ENSG00000181826:ENSG00000171566:ENSG00000158987:ENSG00000026950:ENSG00000111801:ENSG00000112763:ENSG00000182952:ENSG00000146109:ENSG00000181315:ENSG00000124635:ENSG00000198315:ENSG00000137185:ENSG00000187626:ENSG00000158321:ENSG00000101452 |
| DEG.down | Brain_Hypothalamus | 5303 | 16 | 0.754 | 1.000 | ENSG00000198626:ENSG00000163812:ENSG00000075914:ENSG00000154274:ENSG00000181826:ENSG00000026950:ENSG00000111801:ENSG00000112763:ENSG00000182952:ENSG00000146109:ENSG00000096654:ENSG00000197279:ENSG00000198315:ENSG00000137185:ENSG00000187626:ENSG00000101452 |

| | | | | | | |
|---|---|---|---|---|---|---|
| DEG.down | Brain_Nucleus_accumbens_basal_ganglia | 5801 | 18 | 0.720 | 1.000 | ENSG00000196345:ENSG00000163812:ENSG00000075914:ENSG00000163815:ENSG00000181826:ENSG00000158987:ENSG00000026950:ENSG00000111801:ENSG00000112763:ENSG00000182952:ENSG00000146109:ENSG00000181315:ENSG00000197279:ENSG00000196812:ENSG00000198315:ENSG00000137185:ENSG00000187626:ENSG00000101452 |
| DEG.down | Brain_Putamen_basal_ganglia | 7081 | 20 | 0.882 | 1.000 | ENSG00000196345:ENSG00000186448:ENSG00000163812:ENSG00000075914:ENSG00000181826:ENSG00000171566:ENSG00000158987:ENSG00000026950:ENSG00000111801:ENSG00000112763:ENSG00000182952:ENSG00000146109:ENSG00000181315:ENSG00000096654:ENSG00000196812:ENSG00000198315:ENSG00000137185:ENSG00000187626:ENSG00000137338:ENSG00000101452 |
| DEG.down | Brain_Spinal_cord_cervical_c-1 | 4547 | 9 | 0.984 | 1.000 | ENSG00000198626:ENSG00000163812:ENSG00000075914:ENSG00000111801:ENSG00000112763:ENSG00000146109:ENSG00000198315:ENSG00000158321:ENSG00000101452 |
| DEG.down | Brain_Substantia_nigra | 6489 | 17 | 0.931 | 1.000 | ENSG00000198626:ENSG00000196345:ENSG00000163812:ENSG00000075914:ENSG00000181826:ENSG00000026950:ENSG00000111801:ENSG00000112763:ENSG00000182952:ENSG00000146109:ENSG00000124635:ENSG00000096654:ENSG00000198315:ENSG00000137185:ENSG00000187626:ENSG00000158321:ENSG00000101452 |
| DEG.down | Breast_Mammary_Tissue | 912 | 3 | 0.603 | 1.000 | ENSG00000198626:ENSG00000124635:ENSG00000185046 |
| DEG.down | Cells_Cultured_fibroblasts | 1713 | 3 | 0.938 | 1.000 | ENSG00000158985:ENSG00000196812:ENSG00000158321 |

| | | | | | | |
|---|---|---|---|---|---|---|
| DEG.down | Cells_EBV-transformed_lymphocytes | 2063 | 2 | 0.995 | 1.000 | ENSG00000130940:ENSG00000181826 |
| DEG.down | Cervix_Ectocervix | 2 | 0 | 1.000 | 1.000 | |
| DEG.down | Cervix_Endocervix | 5 | 0 | 1.000 | 1.000 | |
| DEG.down | Colon_Sigmoid | 1530 | 1 | 0.996 | 1.000 | ENSG00000185046 |
| DEG.down | Colon_Transverse | 1404 | 2 | 0.957 | 1.000 | ENSG00000198626:ENSG00000189134 |
| DEG.down | Esophagus_Gastroesophageal_Junction | 1406 | 3 | 0.865 | 1.000 | ENSG00000124635:ENSG00000197279:ENSG00000185046 |
| DEG.down | Esophagus_Mucosa | 3122 | 5 | 0.987 | 1.000 | ENSG00000196345:ENSG00000163815:ENSG00000158985:ENSG00000198315:ENSG00000137338 |
| DEG.down | Esophagus_Muscularis | 1582 | 3 | 0.913 | 1.000 | ENSG00000124635:ENSG00000197279:ENSG00000185046 |
| DEG.down | Fallopian_Tube | 1 | 0 | 1.000 | 1.000 | |
| DEG.down | Heart_Atrial_Appendage | 7365 | 21 | 0.879 | 1.000 | ENSG00000186448:ENSG00000163812:ENSG00000075914:ENSG00000181826:ENSG00000171566:ENSG00000171564:ENSG00000158985:ENSG00000158987:ENSG00000026950:ENSG00000111801:ENSG00000112763:ENSG00000146109:ENSG00000181315:ENSG00000124635:ENSG00000096654:ENSG00000196812:ENSG00000198315:ENSG00000137185:ENSG00000187626:ENSG00000137338:ENSG00000158321 |
| DEG.down | Heart_Left_Ventricle | 8514 | 22 | 0.971 | 1.000 | ENSG00000196345:ENSG00000186448:ENSG00000163812:ENSG00000075914:ENSG00000181826:ENSG00000171566:ENSG00000158985:ENSG00000158987:ENSG00000026950:ENSG00000111801:ENSG00000112763:ENSG00000182952:ENSG00000146109:ENSG00000124635:ENSG00000096654:ENSG00000196812:ENSG00000198315:ENSG00000137185: |

| | | | | | | |
|---|---|---|---|---|---|---|
| | | | | | | ENSG00000187626:ENSG00000137338:ENSG00000158321:ENSG00000101452 |
| DEG.down | Kidney_Cortex | 5049 | 11 | 0.975 | 1.000 | ENSG00000186448:ENSG00000181826:ENSG00000171566:ENSG00000158987:ENSG00000112763:ENSG00000146109:ENSG00000124635:ENSG00000198315:ENSG00000189134:ENSG00000158321:ENSG00000101452 |
| DEG.down | Kidney_Medulla | 0 | 0 | 1.000 | 1.000 | |
| DEG.down | Liver | 7196 | 23 | 0.687 | 1.000 | ENSG00000130940:ENSG00000196345:ENSG00000186448:ENSG00000163812:ENSG00000075914:ENSG00000163815:ENSG00000181826:ENSG00000171566:ENSG00000158985:ENSG00000158987:ENSG00000026950:ENSG00000111801:ENSG00000112763:ENSG00000182952:ENSG00000146109:ENSG00000124635:ENSG00000096654:ENSG00000196812:ENSG00000198315:ENSG00000137185:ENSG00000187626:ENSG00000137338:ENSG00000101452 |
| DEG.down | Lung | 820 | 1 | 0.942 | 1.000 | ENSG00000189134 |
| DEG.down | Minor_Salivary_Gland | 1666 | 3 | 0.930 | 1.000 | ENSG00000198626:ENSG00000181826:ENSG00000189134 |
| DEG.down | Muscle_Skeletal | 6093 | 19 | 0.717 | 1.000 | ENSG00000163812:ENSG00000163815:ENSG00000181826:ENSG00000171566:ENSG00000158985:ENSG00000158987:ENSG00000026950:ENSG00000111801:ENSG00000112763:ENSG00000146109:ENSG00000181315:ENSG00000096654:ENSG00000196812:ENSG00000198315:ENSG00000137185:ENSG00000187626:ENSG00000137338:ENSG00000158321:ENSG00000101452 |
| DEG.down | Nerve_Tibial | 784 | 3 | 0.500 | 1.000 | ENSG00000130940:ENSG00000124635:ENSG00000185046 |

| | | | | | | |
|---|---|---|---|---|---|---|
| DEG.down | Ovary | 1429 | 2 | 0.960 | 1.000 | ENSG00000130940:ENSG00000124635 |
| DEG.down | Pancreas | 8671 | 24 | 0.935 | 1.000 | ENSG00000130940:ENSG00000198626:ENSG00000186448:ENSG00000163812:ENSG00000075914:ENSG00000163815:ENSG00000181826:ENSG00000171566:ENSG00000158985:ENSG00000158987:ENSG00000026950:ENSG00000111801:ENSG00000112763:ENSG00000182952:ENSG00000146109:ENSG00000181315:ENSG00000096654:ENSG00000196812:ENSG00000198315:ENSG00000137185:ENSG00000187626:ENSG00000137338:ENSG00000158321:ENSG00000101452 |
| DEG.down | Pituitary | 1752 | 3 | 0.944 | 1.000 | ENSG00000130940:ENSG00000163812:ENSG00000181826 |
| DEG.down | Prostate | 698 | 2 | 0.691 | 1.000 | ENSG00000181826:ENSG00000185046 |
| DEG.down | Skin_Not_Sun_Exposed_Suprapubic | 2330 | 3 | 0.989 | 1.000 | ENSG00000196345:ENSG00000137185:ENSG00000137338 |
| DEG.down | Skin_Sun_Exposed_Lower_leg | 2123 | 4 | 0.940 | 1.000 | ENSG00000196345:ENSG00000124635:ENSG00000137185:ENSG00000137338 |
| DEG.down | Small_Intestine_Terminal_Ileum | 1257 | 2 | 0.933 | 1.000 | ENSG00000198626:ENSG00000189134 |
| DEG.down | Spleen | 1930 | 4 | 0.904 | 1.000 | ENSG00000130940:ENSG00000198626:ENSG00000112812:ENSG00000158321 |
| DEG.down | Stomach | 2290 | 3 | 0.988 | 1.000 | ENSG00000158987:ENSG00000124635:ENSG00000189134 |
| DEG.down | Testis | 2599 | 4 | 0.983 | 1.000 | ENSG00000130940:ENSG00000181826:ENSG00000026950:ENSG00000111801 |
| DEG.down | Thyroid | 976 | 2 | 0.851 | 1.000 | ENSG00000130940:ENSG00000189134 |
| DEG.down | Uterus | 788 | 3 | 0.503 | 1.000 | ENSG00000198626:ENSG00000124635:ENSG00000112812 |
| DEG.down | Vagina | 598 | 2 | 0.606 | 1.000 | ENSG00000198626:ENSG00000189134 |
| DEG.down | Whole_Blood | 6321 | 12 | 0.997 | 1.000 | ENSG00000130940:ENSG00000196345:ENSG00000186448:ENSG00000075914:ENSG00000163815:ENSG00000171566:ENSG0000009 |

| | | | | | | |
|---|---|---|---|---|---|---|
| | | | | | | 6654:ENSG00000198315:ENSG00000137185:ENSG00000187626:ENSG00000158321:ENSG00000101452 |
| DEG.twoside | Adipose_Subcutaneous | 2996 | 3 | 0.999 | 1.000 | ENSG00000130940:ENSG00000198626:ENSG00000181826 |
| DEG.twoside | Adipose_Visceral_Omentum | 2828 | 5 | 0.972 | 1.000 | ENSG00000130940:ENSG00000181826:ENSG00000111801:ENSG00000112812:ENSG00000158321 |
| DEG.twoside | Adrenal_Gland | 4198 | 12 | 0.794 | 1.000 | ENSG00000130940:ENSG00000075914:ENSG00000181826:ENSG00000158985:ENSG00000158987:ENSG00000164398:ENSG00000111801:ENSG00000112812:ENSG00000197279:ENSG00000189134:ENSG00000158321:ENSG00000101452 |
| DEG.twoside | Artery_Aorta | 3426 | 4 | 0.999 | 1.000 | ENSG00000198626:ENSG00000112763:ENSG00000158321:ENSG00000101452 |
| DEG.twoside | Artery_Coronary | 2456 | 4 | 0.974 | 1.000 | ENSG00000198626:ENSG00000112763:ENSG00000197279:ENSG00000101452 |
| DEG.twoside | Artery_Tibial | 3921 | 8 | 0.970 | 1.000 | ENSG00000198626:ENSG00000026950:ENSG00000112763:ENSG00000181315:ENSG00000124635:ENSG00000197279:ENSG00000198315:ENSG00000101452 |
| DEG.twoside | Bladder | 831 | 2 | 0.779 | 1.000 | ENSG00000181315:ENSG00000101452 |
| DEG.twoside | Brain_Amygdala | 8433 | 23 | 0.942 | 1.000 | ENSG00000198626:ENSG00000196345:ENSG00000186448:ENSG00000163812:ENSG00000075914:ENSG00000163815:ENSG00000181826:ENSG00000171566:ENSG00000158987:ENSG00000164398:ENSG00000026950:ENSG00000111801:ENSG00000112763:ENSG00000182952:ENSG00000146109:ENSG00000181315:ENSG00000124635:ENSG00000196812:ENSG00000198315:ENSG00000137185:ENS |

| | | | | | | |
|---|---|---|---|---|---|---|
| | | | | | | G00000187626:ENSG00000185046:ENSG00000101452 |
| DEG.twoside | Brain_Anterior_cingulate_cortex_BA24 | 7869 | 23 | 0.860 | 1.000 | ENSG00000196345:ENSG00000186448:ENSG00000163812:ENSG00000075914:ENSG00000163815:ENSG00000181826:ENSG00000197410:ENSG00000158987:ENSG00000164398:ENSG00000026950:ENSG00000111801:ENSG00000112763:ENSG00000182952:ENSG00000146109:ENSG00000181315:ENSG00000112812:ENSG00000196812:ENSG00000198315:ENSG00000137185:ENSG00000187626:ENSG00000158321:ENSG00000185046:ENSG00000101452 |
| DEG.twoside | Brain_Caudate_basal_ganglia | 7877 | 21 | 0.947 | 1.000 | ENSG00000196345:ENSG00000186448:ENSG00000163812:ENSG00000075914:ENSG00000181826:ENSG00000197410:ENSG00000158987:ENSG00000164398:ENSG00000026950:ENSG00000111801:ENSG00000112763:ENSG00000182952:ENSG00000146109:ENSG00000181315:ENSG00000197279:ENSG00000196812:ENSG00000198315:ENSG00000137185:ENSG00000187626:ENSG00000185046:ENSG00000101452 |
| DEG.twoside | Brain_Cerebellar_Hemisphere | 6629 | 13 | 0.997 | 1.000 | ENSG00000198626:ENSG00000196345:ENSG00000144792:ENSG00000186448:ENSG00000163815:ENSG00000158987:ENSG00000164398:ENSG00000112763:ENSG00000124613:ENSG00000196812:ENSG00000137185:ENSG00000137338:ENSG00000185046 |
| DEG.twoside | Brain_Cerebellum | 6538 | 15 | 0.982 | 1.000 | ENSG00000198626:ENSG00000196345:ENSG00000144792:ENSG00000186448:ENSG00000163815:ENSG00000158985:ENSG00000158987:ENSG00000164398:ENSG00000112763: |

| | | | | | | |
|---|---|---|---|---|---|---|
| | | | | | | ENSG00000124613:ENSG00000197279:ENSG00000196812:ENSG00000137185:ENSG00000137338:ENSG00000185046 |
| DEG.twoside | Brain_Cortex | 7116 | 21 | 0.829 | 1.000 | ENSG00000163812:ENSG00000075914:ENSG00000163815:ENSG00000181826:ENSG00000197410:ENSG00000164398:ENSG00000026950:ENSG00000111801:ENSG00000112763:ENSG00000182952:ENSG00000146109:ENSG00000181315:ENSG00000096654:ENSG00000197279:ENSG00000196812:ENSG00000198315:ENSG00000137185:ENSG00000187626:ENSG00000171811:ENSG00000185046:ENSG00000101452 |
| DEG.twoside | Brain_Frontal_Cortex_BA9 | 6851 | 20 | 0.837 | 1.000 | ENSG00000163812:ENSG00000075914:ENSG00000163815:ENSG00000181826:ENSG00000197410:ENSG00000158985:ENSG00000164398:ENSG00000026950:ENSG00000111801:ENSG00000112763:ENSG00000146109:ENSG00000096654:ENSG00000197279:ENSG00000196812:ENSG00000198315:ENSG00000137185:ENSG00000187626:ENSG00000158321:ENSG00000185046:ENSG00000101452 |
| DEG.twoside | Brain_Hippocampus | 8348 | 21 | 0.978 | 1.000 | ENSG00000196345:ENSG00000186448:ENSG00000163812:ENSG00000075914:ENSG00000181826:ENSG00000171566:ENSG00000158987:ENSG00000164398:ENSG00000026950:ENSG00000111801:ENSG00000112763:ENSG00000182952:ENSG00000146109:ENSG00000181315:ENSG00000124635:ENSG00000198315:ENSG00000137185:ENSG00000187626:ENSG00000158321:ENSG00000185046:ENSG00000101452 |

| | | | | | | |
|---|---|---|---|---|---|---|
| DEG.twoside | Brain_Hypothalamus | 7282 | 19 | 0.949 | 1.000 | ENSG00000198626:ENSG00000163812:ENSG00000075914:ENSG00000154274:ENSG00000181826:ENSG00000164398:ENSG00000026950:ENSG00000111801:ENSG00000112763:ENSG00000182952:ENSG00000146109:ENSG00000096654:ENSG00000197279:ENSG00000198315:ENSG00000137185:ENSG00000187626:ENSG00000171811:ENSG00000185046:ENSG00000101452 |
| DEG.twoside | Brain_Nucleus_accumbens_basal_ganglia | 7635 | 21 | 0.920 | 1.000 | ENSG00000196345:ENSG00000163812:ENSG00000075914:ENSG00000163815:ENSG00000181826:ENSG00000197410:ENSG00000158987:ENSG00000164398:ENSG00000026950:ENSG00000111801:ENSG00000112763:ENSG00000182952:ENSG00000146109:ENSG00000181315:ENSG00000197279:ENSG00000196812:ENSG00000198315:ENSG00000137185:ENSG00000187626:ENSG00000185046:ENSG00000101452 |
| DEG.twoside | Brain_Putamen_basal_ganglia | 8427 | 22 | 0.966 | 1.000 | ENSG00000196345:ENSG00000186448:ENSG00000163812:ENSG00000075914:ENSG00000181826:ENSG00000171566:ENSG00000158987:ENSG00000164398:ENSG00000026950:ENSG00000111801:ENSG00000112763:ENSG00000182952:ENSG00000146109:ENSG00000181315:ENSG00000096654:ENSG00000196812:ENSG00000198315:ENSG00000137185:ENSG00000187626:ENSG00000137338:ENSG00000185046:ENSG00000101452 |
| DEG.twoside | Brain_Spinal_cord_cervical_c-1 | 6158 | 12 | 0.996 | 1.000 | ENSG00000198626:ENSG00000163812:ENSG00000075914:ENSG00000181826:ENSG00000111801:ENSG00000112763:ENSG00000146109:ENSG00000233822:ENSG00000198315: |

| | | | | | | |
|---|---|---|---|---|---|---|
| | | | | | | ENSG00000158321:ENSG00000185046:ENSG00000101452 |
| DEG.twoside | Brain_Substantia_nigra | 7766 | 19 | 0.980 | 1.000 | ENSG00000198626:ENSG00000196345:ENSG00000163812:ENSG00000075914:ENSG00000181826:ENSG00000164398:ENSG00000026950:ENSG00000111801:ENSG00000112763:ENSG00000182952:ENSG00000146109:ENSG00000124635:ENSG00000096654:ENSG00000198315:ENSG00000137185:ENSG00000187626:ENSG00000158321:ENSG00000185046:ENSG00000101452 |
| DEG.twoside | Breast_Mammary_Tissue | 2340 | 5 | 0.912 | 1.000 | ENSG00000198626:ENSG00000181826:ENSG00000124635:ENSG00000158321:ENSG00000185046 |
| DEG.twoside | Cells_Cultured_fibroblasts | 5399 | 11 | 0.989 | 1.000 | ENSG00000196345:ENSG00000186448:ENSG00000075914:ENSG00000171566:ENSG00000158985:ENSG00000146109:ENSG00000196787:ENSG00000196812:ENSG00000187626:ENSG00000189134:ENSG00000158321 |
| DEG.twoside | Cells_EBV-transformed_lymphocytes | 5896 | 19 | 0.652 | 1.000 | ENSG00000130940:ENSG00000075914:ENSG00000181826:ENSG00000171566:ENSG00000158985:ENSG00000158987:ENSG00000026950:ENSG00000111801:ENSG00000146109:ENSG00000124635:ENSG00000196787:ENSG00000096654:ENSG00000185130:ENSG00000196747:ENSG00000184357:ENSG00000197153:ENSG00000124657:ENSG00000196812:ENSG00000187626 |
| DEG.twoside | Cervix_Ectocervix | 189 | 0 | 1.000 | 1.000 | |
| DEG.twoside | Cervix_Endocervix | 1728 | 9 | 0.127 | 1.000 | ENSG00000075914:ENSG00000171566:ENSG00000026950:ENSG00000111801:ENSG000 |

| | | | | | | |
|---|---|---|---|---|---|---|
| | | | | | | 00112763:ENSG00000096654:ENSG00000198315:ENSG00000187626:ENSG00000101452 |
| DEG.twoside | Colon_Sigmoid | 2794 | 5 | 0.970 | 1.000 | ENSG00000130940:ENSG00000197410:ENSG00000137338:ENSG00000185046:ENSG00000101452 |
| DEG.twoside | Colon_Transverse | 2532 | 6 | 0.876 | 1.000 | ENSG00000130940:ENSG00000198626:ENSG00000154274:ENSG00000197410:ENSG00000112812:ENSG00000189134 |
| DEG.twoside | Esophagus_Gastroesophageal_Junction | 2422 | 4 | 0.972 | 1.000 | ENSG00000124635:ENSG00000197279:ENSG00000185046:ENSG00000101452 |
| DEG.twoside | Esophagus_Mucosa | 4877 | 6 | 1.000 | 1.000 | ENSG00000196345:ENSG00000163815:ENSG00000158985:ENSG00000112812:ENSG00000198315:ENSG00000137338 |
| DEG.twoside | Esophagus_Muscularis | 2625 | 5 | 0.954 | 1.000 | ENSG00000198626:ENSG00000124635:ENSG00000197279:ENSG00000185046:ENSG00000101452 |
| DEG.twoside | Fallopian_Tube | 452 | 1 | 0.789 | 1.000 | ENSG00000112763 |
| DEG.twoside | Heart_Atrial_Appendage | 7937 | 22 | 0.920 | 1.000 | ENSG00000198626:ENSG00000186448:ENSG00000163812:ENSG00000075914:ENSG00000181826:ENSG00000171566:ENSG00000171564:ENSG00000158985:ENSG00000158987:ENSG00000026950:ENSG00000111801:ENSG00000112763:ENSG00000146109:ENSG00000181315:ENSG00000124635:ENSG00000096654:ENSG00000196812:ENSG00000198315:ENSG00000137185:ENSG00000187626:ENSG00000137338:ENSG00000158321 |
| DEG.twoside | Heart_Left_Ventricle | 8909 | 23 | 0.976 | 1.000 | ENSG00000198626:ENSG00000196345:ENSG00000186448:ENSG00000163812:ENSG00000075914:ENSG00000181826:ENSG00000171566:ENSG00000158985:ENSG00000158987:ENSG00000026950:ENSG00000111801:ENS |

| | | | | | | |
|---|---|---|---|---|---|---|
| | | | | | | G00000112763:ENSG00000182952:ENSG00000146109:ENSG00000124635:ENSG00000096654:ENSG00000196812:ENSG00000198315:ENSG00000137185:ENSG00000187626:ENSG00000137338:ENSG00000158321:ENSG00000101452 |
| DEG.twoside | Kidney_Cortex | 5997 | 14 | 0.972 | 1.000 | ENSG00000186448:ENSG00000154274:ENSG00000181826:ENSG00000171566:ENSG00000171564:ENSG00000158987:ENSG00000112763:ENSG00000146109:ENSG00000124635:ENSG00000197279:ENSG00000198315:ENSG00000189134:ENSG00000158321:ENSG00000101452 |
| DEG.twoside | Kidney_Medulla | 93 | 0 | 1.000 | 1.000 | |
| DEG.twoside | Liver | 8286 | 25 | 0.822 | 1.000 | ENSG00000130940:ENSG00000196345:ENSG00000186448:ENSG00000163812:ENSG00000075914:ENSG00000163815:ENSG00000154274:ENSG00000181826:ENSG00000171566:ENSG00000171564:ENSG00000158985:ENSG00000158987:ENSG00000026950:ENSG00000111801:ENSG00000112763:ENSG00000182952:ENSG00000146109:ENSG00000124635:ENSG00000096654:ENSG00000196812:ENSG00000198315:ENSG00000137185:ENSG00000187626:ENSG00000137338:ENSG00000101452 |
| DEG.twoside | Lung | 3849 | 10 | 0.868 | 1.000 | ENSG00000130940:ENSG00000181826:ENSG00000158985:ENSG00000026950:ENSG00000111801:ENSG00000112763:ENSG00000124635:ENSG00000112812:ENSG00000189134:ENSG00000171811 |

| | | | | | | |
|---|---|---|---|---|---|---|
| DEG.twoside | Minor_Salivary_Gland | 2951 | 8 | 0.804 | 1.000 | ENSG00000130940:ENSG00000198626:ENSG00000154274:ENSG00000181826:ENSG00000112812:ENSG00000196812:ENSG00000189134:ENSG00000158321 |
| DEG.twoside | Muscle_Skeletal | 6972 | 20 | 0.862 | 1.000 | ENSG00000163812:ENSG00000163815:ENSG00000181826:ENSG00000171566:ENSG00000158985:ENSG00000158987:ENSG00000026950:ENSG00000111801:ENSG00000112763:ENSG00000146109:ENSG00000181315:ENSG00000096654:ENSG00000196812:ENSG00000198315:ENSG00000137185:ENSG00000187626:ENSG00000137338:ENSG00000158321:ENSG00000171811:ENSG00000101452 |
| DEG.twoside | Nerve_Tibial | 4941 | 13 | 0.891 | 1.000 | ENSG00000130940:ENSG00000196345:ENSG00000144792:ENSG00000186448:ENSG00000163812:ENSG00000181826:ENSG00000158987:ENSG00000181315:ENSG00000124635:ENSG00000198315:ENSG00000137338:ENSG00000158321:ENSG00000185046 |
| DEG.twoside | Ovary | 5103 | 12 | 0.955 | 1.000 | ENSG00000130940:ENSG00000196345:ENSG00000144792:ENSG00000186448:ENSG00000026950:ENSG00000111801:ENSG00000181315:ENSG00000124635:ENSG00000124613:ENSG00000198315:ENSG00000137185:ENSG00000158321 |
| DEG.twoside | Pancreas | 9220 | 26 | 0.930 | 1.000 | ENSG00000130940:ENSG00000198626:ENSG00000186448:ENSG00000163812:ENSG00000075914:ENSG00000163815:ENSG00000154274:ENSG00000181826:ENSG00000171566:ENSG00000158985:ENSG00000158987:ENSG00000026950:ENSG00000111801:ENSG00000112763:ENSG00000182952:ENSG00000146109:ENSG00000181315:ENSG00000112812: |

| | | | | | | |
|---|---|---|---|---|---|---|
| | | | | | | ENSG00000096654:ENSG00000196812:ENSG00000198315:ENSG00000137185:ENSG00000187626:ENSG00000137338:ENSG00000158321:ENSG00000101452 |
| DEG.twoside | Pituitary | 5005 | 11 | 0.972 | 1.000 | ENSG00000130940:ENSG00000196345:ENSG00000144792:ENSG00000163812:ENSG00000181826:ENSG00000111801:ENSG00000112812:ENSG00000096654:ENSG00000197279:ENSG00000137185:ENSG00000171811 |
| DEG.twoside | Prostate | 2717 | 9 | 0.585 | 1.000 | ENSG00000130940:ENSG00000144792:ENSG00000181826:ENSG00000026950:ENSG00000124635:ENSG00000112812:ENSG00000196812:ENSG00000137185:ENSG00000185046 |
| DEG.twoside | Skin_Not_Sun_Exposed_Suprapubic | 4367 | 9 | 0.976 | 1.000 | ENSG00000130940:ENSG00000196345:ENSG00000163812:ENSG00000075914:ENSG00000146109:ENSG00000112812:ENSG00000137185:ENSG00000137338:ENSG00000158321 |
| DEG.twoside | Skin_Sun_Exposed_Lower_leg | 4286 | 10 | 0.940 | 1.000 | ENSG00000130940:ENSG00000196345:ENSG00000163812:ENSG00000075914:ENSG00000146109:ENSG00000124635:ENSG00000112812:ENSG00000137185:ENSG00000137338:ENSG00000158321 |
| DEG.twoside | Small_Intestine_Terminal_Ileum | 3112 | 9 | 0.750 | 1.000 | ENSG00000198626:ENSG00000154274:ENSG00000158985:ENSG00000026950:ENSG00000111801:ENSG00000112812:ENSG00000184357:ENSG00000196812:ENSG00000189134 |
| DEG.twoside | Spleen | 4596 | 15 | 0.616 | 1.000 | ENSG00000130940:ENSG00000198626:ENSG00000158985:ENSG00000026950:ENSG00000111801:ENSG00000112763:ENSG00000146109:ENSG00000124635:ENSG00000196787:ENSG00000112812:ENSG00000196747:ENSG00000196812:ENSG00000137185:ENSG00000187626:ENSG00000158321 |

| | | | | | | |
|---|---|---|---|---|---|---|
| DEG.twoside | Stomach | 3070 | 8 | 0.839 | 1.000 | ENSG00000130940:ENSG00000154274:ENSG00000197410:ENSG00000171564:ENSG00000158987:ENSG00000124635:ENSG00000112812:ENSG00000189134 |
| DEG.twoside | Testis | 8546 | 23 | 0.952 | 1.000 | ENSG00000130940:ENSG00000196345:ENSG00000144792:ENSG00000163812:ENSG00000075914:ENSG00000181826:ENSG00000197410:ENSG00000158985:ENSG00000158987:ENSG00000164398:ENSG00000026950:ENSG00000111801:ENSG00000124635:ENSG00000196787:ENSG00000158553:ENSG00000124613:ENSG00000096654:ENSG00000233822:ENSG00000197279:ENSG00000137185:ENSG00000187626:ENSG00000189134:ENSG00000171811 |
| DEG.twoside | Thyroid | 4708 | 14 | 0.760 | 1.000 | ENSG00000130940:ENSG00000196345:ENSG00000144792:ENSG00000186448:ENSG00000181826:ENSG00000158985:ENSG00000111801:ENSG00000112812:ENSG00000124613:ENSG00000196812:ENSG00000198315:ENSG00000137185:ENSG00000189134:ENSG00000171811 |
| DEG.twoside | Uterus | 4787 | 20 | 0.170 | 1.000 | ENSG00000198626:ENSG00000196345:ENSG00000144792:ENSG00000186448:ENSG00000154274:ENSG00000197410:ENSG00000171566:ENSG00000026950:ENSG00000111801:ENSG00000112763:ENSG00000146109:ENSG00000181315:ENSG00000124635:ENSG00000112812:ENSG00000124613:ENSG00000198315:ENSG00000137185:ENSG00000187626:ENSG00000137338:ENSG00000101452 |

| | | | | | | |
|---|---|---|---|---|---|---|
| DEG.twoside | Vagina | 2611 | 6 | 0.894 | 1.000 | ENSG00000130940:ENSG00000198626:ENSG00000026950:ENSG00000111801:ENSG00000124635:ENSG00000189134 |
| DEG.twoside | Whole_Blood | 7620 | 16 | 0.997 | 1.000 | ENSG00000130940:ENSG00000196345:ENSG00000186448:ENSG00000075914:ENSG00000163815:ENSG00000171566:ENSG00000158985:ENSG00000026950:ENSG00000124635:ENSG00000096654:ENSG00000196747:ENSG00000198315:ENSG00000137185:ENSG00000187626:ENSG00000158321:ENSG00000101452 |

***Table S20. Tissue Specificity of Prioritised Genes from FUMA (using GTEx v8 54 Tissue Types): Executive Function (RFFT)***

| Category | GeneSet | N_genes | N_overlap | p | adjP | genes |
|---|---|---|---|---|---|---|
| DEG.up | Adipose_Subcutaneous | 1689 | 2 | 0.735 | 1.000 | ENSG00000173482:ENSG00000099864 |
| DEG.up | Adipose_Visceral_Omentum | 1428 | 2 | 0.643 | 1.000 | ENSG00000173482:ENSG00000099864 |
| DEG.up | Adrenal_Gland | 1204 | 1 | 0.846 | 1.000 | ENSG00000175497 |
| DEG.up | Artery_Aorta | 2103 | 3 | 0.626 | 1.000 | ENSG00000110841:ENSG00000170412:ENSG00000173482 |
| DEG.up | Artery_Coronary | 1593 | 1 | 0.918 | 1.000 | ENSG00000170412 |
| DEG.up | Artery_Tibial | 2175 | 1 | 0.969 | 1.000 | ENSG00000170412 |
| DEG.up | Bladder | 728 | 0 | 1.000 | 1.000 | |
| DEG.up | Brain_Amygdala | 1438 | 7 | 0.004 | 0.237 | ENSG00000175497:ENSG00000175161:ENSG00000120088:ENSG00000186868:ENSG00000073969:ENSG00000099864:ENSG00000186732 |
| DEG.up | Brain_Anterior_cingulate_cortex_BA24 | 1866 | 7 | 0.018 | 0.949 | ENSG00000175497:ENSG00000175161:ENSG00000120088:ENSG00000186868:ENSG00000073969:ENSG00000099864:ENSG00000186732 |
| DEG.up | Brain_Caudate_basal_ganglia | 1703 | 5 | 0.106 | 1.000 | ENSG00000175497:ENSG00000175161:ENSG00000186868:ENSG00000073969:ENSG00000099864 |
| DEG.up | Brain_Cerebellar_Hemisphere | 4248 | 12 | 0.014 | 0.741 | ENSG00000175161:ENSG00000225190:ENSG00000120088:ENSG00000185294:ENSG00000186868:ENSG00000256762:ENSG00000120071:ENSG00000228696:ENSG00000238083:ENSG00000073969:ENSG00000170412:ENSG00000099864 |
| DEG.up | Brain_Cerebellum | 4055 | 11 | 0.026 | 1.000 | ENSG00000175161:ENSG00000225190:ENSG00000120088:ENSG00000185294:ENSG00000186868:ENSG00000256762:ENSG00000120071:ENSG00000238083:ENSG00000073969:ENSG00000170412:ENSG00000099864 |

| | | | | | | |
|---|---|---|---|---|---|---|
| DEG.up | Brain_Cortex | 2159 | 7 | 0.036 | 1.000 | ENSG00000175497:ENSG00000175161:ENSG00000120088:ENSG00000186868:ENSG00000073969:ENSG00000099864:ENSG00000186732 |
| DEG.up | Brain_Frontal_Cortex_BA9 | 2422 | 7 | 0.061 | 1.000 | ENSG00000175497:ENSG00000175161:ENSG00000120088:ENSG00000186868:ENSG00000073969:ENSG00000099864:ENSG00000186732 |
| DEG.up | Brain_Hippocampus | 1542 | 7 | 0.006 | 0.347 | ENSG00000175497:ENSG00000175161:ENSG00000120088:ENSG00000186868:ENSG00000073969:ENSG00000099864:ENSG00000186732 |
| DEG.up | Brain_Hypothalamus | 1979 | 6 | 0.069 | 1.000 | ENSG00000175497:ENSG00000175161:ENSG00000186868:ENSG00000073969:ENSG00000108379:ENSG00000099864 |
| DEG.up | Brain_Nucleus_accumbens_basal_ganglia | 1834 | 5 | 0.134 | 1.000 | ENSG00000175497:ENSG00000175161:ENSG00000186868:ENSG00000073969:ENSG00000099864 |
| DEG.up | Brain_Putamen_basal_ganglia | 1346 | 5 | 0.047 | 1.000 | ENSG00000175497:ENSG00000175161:ENSG00000186868:ENSG00000073969:ENSG00000099864 |
| DEG.up | Brain_Spinal_cord_cervical_c-1 | 1611 | 6 | 0.029 | 1.000 | ENSG00000175497:ENSG00000175161:ENSG00000104537:ENSG00000186868:ENSG00000099864:ENSG00000186732 |
| DEG.up | Brain_Substantia_nigra | 1277 | 5 | 0.039 | 1.000 | ENSG00000175497:ENSG00000175161:ENSG00000104537:ENSG00000186868:ENSG00000099864 |
| DEG.up | Breast_Mammary_Tissue | 1428 | 2 | 0.643 | 1.000 | ENSG00000173482:ENSG00000099864 |
| DEG.up | Cells_Cultured_fibroblasts | 3686 | 4 | 0.833 | 1.000 | ENSG00000029153:ENSG00000110841:ENSG00000073969:ENSG00000173482 |
| DEG.up | Cells_EBV-transformed_lymphocytes | 3833 | 3 | 0.947 | 1.000 | ENSG00000035720:ENSG00000029153:ENSG00000165935 |
| DEG.up | Cervix_Ectocervix | 187 | 0 | 1.000 | 1.000 | |
| DEG.up | Cervix_Endocervix | 1723 | 2 | 0.745 | 1.000 | ENSG00000120071:ENSG00000099864 |
| DEG.up | Colon_Sigmoid | 1264 | 1 | 0.860 | 1.000 | ENSG00000104537 |

| | | | | | | |
|---|---|---|---|---|---|---|
| DEG.up | Colon_Transverse | 1128 | 3 | 0.238 | 1.000 | ENSG00000104537:ENSG00000159314:ENSG00000099812 |
| DEG.up | Esophagus_Gastroesophageal_Junction | 1016 | 0 | 1.000 | 1.000 | |
| DEG.up | Esophagus_Mucosa | 1755 | 4 | 0.267 | 1.000 | ENSG00000029153:ENSG00000165935:ENSG00000159314:ENSG00000225190 |
| DEG.up | Esophagus_Muscularis | 1043 | 1 | 0.801 | 1.000 | ENSG00000104537 |
| DEG.up | Fallopian_Tube | 451 | 1 | 0.496 | 1.000 | ENSG00000099864 |
| DEG.up | Heart_Atrial_Appendage | 572 | 0 | 1.000 | 1.000 | |
| DEG.up | Heart_Left_Ventricle | 395 | 0 | 1.000 | 1.000 | |
| DEG.up | Kidney_Cortex | 948 | 3 | 0.168 | 1.000 | ENSG00000035720:ENSG00000170412:ENSG00000099812 |
| DEG.up | Kidney_Medulla | 93 | 1 | 0.131 | 1.000 | ENSG00000170412 |
| DEG.up | Liver | 1090 | 5 | 0.021 | 1.000 | ENSG00000104537:ENSG00000100652:ENSG00000108379:ENSG00000170412:ENSG00000186732 |
| DEG.up | Lung | 3029 | 4 | 0.689 | 1.000 | ENSG00000110841:ENSG00000159314:ENSG00000173482:ENSG00000099812 |
| DEG.up | Minor_Salivary_Gland | 1285 | 2 | 0.584 | 1.000 | ENSG00000029153:ENSG00000159314 |
| DEG.up | Muscle_Skeletal | 879 | 2 | 0.383 | 1.000 | ENSG00000186868:ENSG00000170412 |
| DEG.up | Nerve_Tibial | 4157 | 2 | 0.992 | 1.000 | ENSG00000175161:ENSG00000099864 |
| DEG.up | Ovary | 3674 | 4 | 0.831 | 1.000 | ENSG00000110841:ENSG00000120071:ENSG00000170412:ENSG00000099864 |
| DEG.up | Pancreas | 549 | 0 | 1.000 | 1.000 | |
| DEG.up | Pituitary | 3253 | 6 | 0.360 | 1.000 | ENSG00000120088:ENSG00000186868:ENSG00000238083:ENSG00000073969:ENSG00000099864:ENSG00000099812 |
| DEG.up | Prostate | 2019 | 2 | 0.823 | 1.000 | ENSG00000170412:ENSG00000099812 |
| DEG.up | Skin_Not_Sun_Exposed_Suprapubic | 2037 | 3 | 0.603 | 1.000 | ENSG00000029153:ENSG00000159314:ENSG00000108379 |

| | | | | | | |
|---|---|---|---|---|---|---|
| DEG.up | Skin_Sun_Exposed_Lower_leg | 2163 | 5 | 0.218 | 1.000 | ENSG00000029153:ENSG00000159314:ENSG00000225190:ENSG00000120071:ENSG00000108379 |
| DEG.up | Small_Intestine_Terminal_Ileum | 1855 | 4 | 0.301 | 1.000 | ENSG00000035720:ENSG00000104537:ENSG00000159314:ENSG00000099812 |
| DEG.up | Spleen | 2666 | 2 | 0.925 | 1.000 | ENSG00000035720:ENSG00000159314 |
| DEG.up | Stomach | 780 | 2 | 0.328 | 1.000 | ENSG00000170412:ENSG00000099812 |
| DEG.up | Testis | 5947 | 8 | 0.714 | 1.000 | ENSG00000035720:ENSG00000104537:ENSG00000165935:ENSG00000185294:ENSG00000120071:ENSG00000176681:ENSG00000238083:ENSG00000108379 |
| DEG.up | Thyroid | 3732 | 4 | 0.841 | 1.000 | ENSG00000110841:ENSG00000120071:ENSG00000170412:ENSG00000173482 |
| DEG.up | Uterus | 3999 | 6 | 0.575 | 1.000 | ENSG00000104537:ENSG00000120088:ENSG00000120071:ENSG00000238083:ENSG00000173482:ENSG00000099864 |
| DEG.up | Vagina | 2013 | 4 | 0.357 | 1.000 | ENSG00000029153:ENSG00000159314:ENSG00000225190:ENSG00000120071 |
| DEG.up | Whole_Blood | 1299 | 3 | 0.309 | 1.000 | ENSG00000035720:ENSG00000159314:ENSG00000225190 |
| DEG.down | Adipose_Subcutaneous | 1307 | 3 | 0.312 | 1.000 | ENSG00000175161:ENSG00000108379:ENSG00000170412 |
| DEG.down | Adipose_Visceral_Omentum | 1400 | 3 | 0.351 | 1.000 | ENSG00000029153:ENSG00000186868:ENSG00000170412 |
| DEG.down | Adrenal_Gland | 2994 | 4 | 0.680 | 1.000 | ENSG00000159314:ENSG00000186868:ENSG00000170412:ENSG00000099864 |
| DEG.down | Artery_Aorta | 1323 | 3 | 0.319 | 1.000 | ENSG00000159314:ENSG00000186868:ENSG00000099864 |
| DEG.down | Artery_Coronary | 863 | 2 | 0.374 | 1.000 | ENSG00000175161:ENSG00000186868 |
| DEG.down | Artery_Tibial | 1746 | 3 | 0.494 | 1.000 | ENSG00000159314:ENSG00000186868:ENSG00000099864 |
| DEG.down | Bladder | 103 | 0 | 1.000 | 1.000 | |

| | | | | | | |
|---|---|---|---|---|---|---|
| DEG.down | Brain_Amygdala | 6995 | 7 | 0.946 | 1.000 | ENSG00000110841:ENSG00000159314:ENSG00000225190:ENSG00000120071:ENSG00000238083:ENSG00000170412:ENSG00000173482 |
| DEG.down | Brain_Anterior_cingulate_cortex_BA24 | 6003 | 6 | 0.929 | 1.000 | ENSG00000110841:ENSG00000159314:ENSG00000225190:ENSG00000120071:ENSG00000170412:ENSG00000173482 |
| DEG.down | Brain_Caudate_basal_ganglia | 6174 | 7 | 0.870 | 1.000 | ENSG00000029153:ENSG00000110841:ENSG00000159314:ENSG00000225190:ENSG00000120071:ENSG00000170412:ENSG00000173482 |
| DEG.down | Brain_Cerebellar_Hemisphere | 2381 | 2 | 0.889 | 1.000 | ENSG00000110841:ENSG00000173482 |
| DEG.down | Brain_Cerebellum | 2483 | 3 | 0.740 | 1.000 | ENSG00000110841:ENSG00000173482:ENSG00000186732 |
| DEG.down | Brain_Cortex | 4957 | 5 | 0.902 | 1.000 | ENSG00000159314:ENSG00000225190:ENSG00000120071:ENSG00000170412:ENSG00000173482 |
| DEG.down | Brain_Frontal_Cortex_BA9 | 4429 | 5 | 0.831 | 1.000 | ENSG00000159314:ENSG00000225190:ENSG00000120071:ENSG00000170412:ENSG00000173482 |
| DEG.down | Brain_Hippocampus | 6806 | 6 | 0.973 | 1.000 | ENSG00000110841:ENSG00000159314:ENSG00000225190:ENSG00000120071:ENSG00000170412:ENSG00000173482 |
| DEG.down | Brain_Hypothalamus | 5303 | 5 | 0.934 | 1.000 | ENSG00000110841:ENSG00000159314:ENSG00000225190:ENSG00000120071:ENSG00000170412 |
| DEG.down | Brain_Nucleus_accumbens_basal_ganglia | 5801 | 7 | 0.817 | 1.000 | ENSG00000029153:ENSG00000110841:ENSG00000159314:ENSG00000225190:ENSG00000120071:ENSG00000170412:ENSG00000173482 |
| DEG.down | Brain_Putamen_basal_ganglia | 7081 | 8 | 0.893 | 1.000 | ENSG00000110841:ENSG00000159314:ENSG00000225190:ENSG00000120071:ENSG00000238083:ENSG00000108379:ENSG00000170412:ENSG00000173482 |

| | | | | | | |
|---|---|---|---|---|---|---|
| DEG.down | Brain_Spinal_cord_cervical_c-1 | 4547 | 6 | 0.712 | 1.000 | ENSG00000110841:ENSG00000159314:ENSG00000225190:ENSG00000120071:ENSG00000170412:ENSG00000173482 |
| DEG.down | Brain_Substantia_nigra | 6489 | 8 | 0.814 | 1.000 | ENSG00000029153:ENSG00000110841:ENSG00000159314:ENSG00000225190:ENSG00000120071:ENSG00000238083:ENSG00000170412:ENSG00000173482 |
| DEG.down | Breast_Mammary_Tissue | 912 | 1 | 0.755 | 1.000 | ENSG00000029153 |
| DEG.down | Cells_Cultured_fibroblasts | 1713 | 4 | 0.252 | 1.000 | ENSG00000159314:ENSG00000238083:ENSG00000108379:ENSG00000099864 |
| DEG.down | Cells_EBV-transformed_lymphocytes | 2063 | 2 | 0.832 | 1.000 | ENSG00000225190:ENSG00000170412 |
| DEG.down | Cervix_Ectocervix | 2 | 0 | 1.000 | 1.000 | |
| DEG.down | Cervix_Endocervix | 5 | 0 | 1.000 | 1.000 | |
| DEG.down | Colon_Sigmoid | 1530 | 3 | 0.406 | 1.000 | ENSG00000159314:ENSG00000108379:ENSG00000170412 |
| DEG.down | Colon_Transverse | 1404 | 2 | 0.634 | 1.000 | ENSG00000029153:ENSG00000186868 |
| DEG.down | Esophagus_Gastroesophageal_Junction | 1406 | 5 | 0.055 | 1.000 | ENSG00000159314:ENSG00000186868:ENSG00000073969:ENSG00000108379:ENSG00000170412 |
| DEG.down | Esophagus_Mucosa | 3122 | 3 | 0.871 | 1.000 | ENSG00000170412:ENSG00000173482:ENSG00000099864 |
| DEG.down | Esophagus_Muscularis | 1582 | 5 | 0.083 | 1.000 | ENSG00000159314:ENSG00000186868:ENSG00000073969:ENSG00000108379:ENSG00000170412 |
| DEG.down | Fallopian_Tube | 1 | 0 | 1.000 | 1.000 | |
| DEG.down | Heart_Atrial_Appendage | 7365 | 7 | 0.966 | 1.000 | ENSG00000159314:ENSG00000225190:ENSG00000186868:ENSG00000120071:ENSG00000238083:ENSG00000073969:ENSG00000170412 |
| DEG.down | Heart_Left_Ventricle | 8514 | 7 | 0.993 | 1.000 | ENSG00000159314:ENSG00000225190:ENSG00000186868:ENSG00000120071:ENSG00000073969:ENSG00000170412:ENSG00000099864 |

| | | | | | | |
|---|---|---|---|---|---|---|
| DEG.down | Kidney_Cortex | 5049 | 4 | 0.967 | 1.000 | ENSG00000225190:ENSG00000120071:ENSG00000073969:ENSG00000108379 |
| DEG.down | Kidney_Medulla | 0 | 0 | 1.000 | 1.000 | |
| DEG.down | Liver | 7196 | 8 | 0.905 | 1.000 | ENSG00000110841:ENSG00000159314:ENSG00000225190:ENSG00000120071:ENSG00000238083:ENSG00000073969:ENSG00000173482:ENSG00000099864 |
| DEG.down | Lung | 820 | 2 | 0.350 | 1.000 | ENSG00000186868:ENSG00000108379 |
| DEG.down | Minor_Salivary_Gland | 1666 | 2 | 0.728 | 1.000 | ENSG00000173482:ENSG00000099864 |
| DEG.down | Muscle_Skeletal | 6093 | 5 | 0.976 | 1.000 | ENSG00000110841:ENSG00000159314:ENSG00000120071:ENSG00000238083:ENSG00000099864 |
| DEG.down | Nerve_Tibial | 784 | 3 | 0.111 | 1.000 | ENSG00000029153:ENSG00000186868:ENSG00000108379 |
| DEG.down | Ovary | 1429 | 4 | 0.163 | 1.000 | ENSG00000159314:ENSG00000186868:ENSG00000073969:ENSG00000108379 |
| DEG.down | Pancreas | 8671 | 8 | 0.984 | 1.000 | ENSG00000110841:ENSG00000159314:ENSG00000225190:ENSG00000120071:ENSG00000238083:ENSG00000073969:ENSG00000173482:ENSG00000099864 |
| DEG.down | Pituitary | 1752 | 2 | 0.754 | 1.000 | ENSG00000029153:ENSG00000170412 |
| DEG.down | Prostate | 698 | 2 | 0.282 | 1.000 | ENSG00000029153:ENSG00000108379 |
| DEG.down | Skin_Not_Sun_Exposed_Suprapubic | 2330 | 4 | 0.470 | 1.000 | ENSG00000120088:ENSG00000186868:ENSG00000173482:ENSG00000099812 |
| DEG.down | Skin_Sun_Exposed_Lower_leg | 2123 | 1 | 0.966 | 1.000 | ENSG00000099812 |
| DEG.down | Small_Intestine_Terminal_Ileum | 1257 | 2 | 0.572 | 1.000 | ENSG00000175161:ENSG00000186868 |
| DEG.down | Spleen | 1930 | 1 | 0.953 | 1.000 | ENSG00000108379 |
| DEG.down | Stomach | 2290 | 4 | 0.456 | 1.000 | ENSG00000175161:ENSG00000159314:ENSG00000186868:ENSG00000099864 |

| | | | | | | |
|---|---|---|---|---|---|---|
| DEG.down | Testis | 2599 | 4 | 0.561 | 1.000 | ENSG00000110841:ENSG00000225190:ENSG00000170412:ENSG00000099812 |
| DEG.down | Thyroid | 976 | 1 | 0.778 | 1.000 | ENSG00000186868 |
| DEG.down | Uterus | 788 | 3 | 0.113 | 1.000 | ENSG00000029153:ENSG00000186868:ENSG00000108379 |
| DEG.down | Vagina | 598 | 1 | 0.599 | 1.000 | ENSG00000186868 |
| DEG.down | Whole_Blood | 6321 | 2 | 1.000 | 1.000 | ENSG00000120071:ENSG00000073969 |
| DEG.twoside | Adipose_Subcutaneous | 2996 | 5 | 0.475 | 1.000 | ENSG00000175161:ENSG00000108379:ENSG00000170412:ENSG00000173482:ENSG00000099864 |
| DEG.twoside | Adipose_Visceral_Omentum | 2828 | 5 | 0.422 | 1.000 | ENSG00000029153:ENSG00000186868:ENSG00000170412:ENSG00000173482:ENSG00000099864 |
| DEG.twoside | Adrenal_Gland | 4198 | 5 | 0.789 | 1.000 | ENSG00000175497:ENSG00000159314:ENSG00000186868:ENSG00000170412:ENSG00000099864 |
| DEG.twoside | Artery_Aorta | 3426 | 6 | 0.411 | 1.000 | ENSG00000110841:ENSG00000159314:ENSG00000186868:ENSG00000170412:ENSG00000173482:ENSG00000099864 |
| DEG.twoside | Artery_Coronary | 2456 | 3 | 0.733 | 1.000 | ENSG00000175161:ENSG00000186868:ENSG00000170412 |
| DEG.twoside | Artery_Tibial | 3921 | 4 | 0.870 | 1.000 | ENSG00000159314:ENSG00000186868:ENSG00000170412:ENSG00000099864 |
| DEG.twoside | Bladder | 831 | 0 | 1.000 | 1.000 | |
| DEG.twoside | Brain_Amygdala | 8433 | 14 | 0.371 | 1.000 | ENSG00000175497:ENSG00000175161:ENSG00000110841:ENSG00000159314:ENSG00000225190:ENSG00000120088:ENSG00000186868:ENSG00000120071:ENSG00000238083:ENSG00000073969:ENSG00000170412:ENSG00000173482:ENSG00000099864:ENSG00000186732 |
| DEG.twoside | Brain_Anterior_cingulate_cortex_BA24 | 7869 | 13 | 0.391 | 1.000 | ENSG00000175497:ENSG00000175161:ENSG00000110841:ENSG00000159314:ENSG000002 |

| | | | | | | |
|---|---|---|---|---|---|---|
| | | | | | | 25190:ENSG00000120088:ENSG00000186868:ENSG00000120071:ENSG00000073969:ENSG00000170412:ENSG00000173482:ENSG00000099864:ENSG00000186732 |
| DEG.twoside | Brain_Caudate_basal_ganglia | 7877 | 12 | 0.546 | 1.000 | ENSG00000175497:ENSG00000175161:ENSG00000029153:ENSG00000110841:ENSG00000159314:ENSG00000225190:ENSG00000186868:ENSG00000120071:ENSG00000073969:ENSG00000170412:ENSG00000173482:ENSG00000099864 |
| DEG.twoside | Brain_Cerebellar_Hemisphere | 6629 | 14 | 0.080 | 1.000 | ENSG00000175161:ENSG00000110841:ENSG00000225190:ENSG00000120088:ENSG00000185294:ENSG00000186868:ENSG00000256762:ENSG00000120071:ENSG00000228696:ENSG00000238083:ENSG00000073969:ENSG00000170412:ENSG00000173482:ENSG00000099864 |
| DEG.twoside | Brain_Cerebellum | 6538 | 14 | 0.072 | 1.000 | ENSG00000175161:ENSG00000110841:ENSG00000225190:ENSG00000120088:ENSG00000185294:ENSG00000186868:ENSG00000256762:ENSG00000120071:ENSG00000238083:ENSG00000073969:ENSG00000170412:ENSG00000173482:ENSG00000099864:ENSG00000186732 |
| DEG.twoside | Brain_Cortex | 7116 | 12 | 0.368 | 1.000 | ENSG00000175497:ENSG00000175161:ENSG00000159314:ENSG00000225190:ENSG00000120088:ENSG00000186868:ENSG00000120071:ENSG00000073969:ENSG00000170412:ENSG00000173482:ENSG00000099864:ENSG00000186732 |
| DEG.twoside | Brain_Frontal_Cortex_BA9 | 6851 | 12 | 0.310 | 1.000 | ENSG00000175497:ENSG00000175161:ENSG00000159314:ENSG00000225190:ENSG00000120088:ENSG00000186868:ENSG00000120071:ENSG00000073969:ENSG00000170412:ENSG0 |

| | | | | | | |
|---|---|---|---|---|---|---|
| | | | | | | 0000173482:ENSG00000099864:ENSG00000186732 |
| DEG.twoside | Brain_Hippocampus | 8348 | 13 | 0.502 | 1.000 | ENSG00000175497:ENSG00000175161:ENSG00000110841:ENSG00000159314:ENSG00000225190:ENSG00000120088:ENSG00000186868:ENSG00000120071:ENSG00000073969:ENSG00000170412:ENSG00000173482:ENSG00000099864:ENSG00000186732 |
| DEG.twoside | Brain_Hypothalamus | 7282 | 11 | 0.561 | 1.000 | ENSG00000175497:ENSG00000175161:ENSG00000110841:ENSG00000159314:ENSG00000225190:ENSG00000186868:ENSG00000120071:ENSG00000073969:ENSG00000108379:ENSG00000170412:ENSG00000099864 |
| DEG.twoside | Brain_Nucleus_accumbens_basal_ganglia | 7635 | 12 | 0.489 | 1.000 | ENSG00000175497:ENSG00000175161:ENSG00000029153:ENSG00000110841:ENSG00000159314:ENSG00000225190:ENSG00000186868:ENSG00000120071:ENSG00000073969:ENSG00000170412:ENSG00000173482:ENSG00000099864 |
| DEG.twoside | Brain_Putamen_basal_ganglia | 8427 | 13 | 0.520 | 1.000 | ENSG00000175497:ENSG00000175161:ENSG00000110841:ENSG00000159314:ENSG00000225190:ENSG00000186868:ENSG00000120071:ENSG00000238083:ENSG00000073969:ENSG00000108379:ENSG00000170412:ENSG00000173482:ENSG00000099864 |
| DEG.twoside | Brain_Spinal_cord_cervical_c-1 | 6158 | 12 | 0.178 | 1.000 | ENSG00000175497:ENSG00000175161:ENSG00000104537:ENSG00000110841:ENSG00000159314:ENSG00000225190:ENSG00000186868:ENSG00000120071:ENSG00000170412:ENSG00000173482:ENSG00000099864:ENSG00000186732 |
| DEG.twoside | Brain_Substantia_nigra | 7766 | 13 | 0.368 | 1.000 | ENSG00000175497:ENSG00000175161:ENSG00000104537:ENSG00000029153:ENSG000001 |

| | | | | | | |
|---|---|---|---|---|---|---|
| | | | | | | 10841:ENSG00000159314:ENSG00000225190:ENSG00000186868:ENSG00000120071:ENSG00000238083:ENSG00000170412:ENSG00000173482:ENSG00000099864 |
| DEG.twoside | Breast_Mammary_Tissue | 2340 | 3 | 0.701 | 1.000 | ENSG00000029153:ENSG00000173482:ENSG00000099864 |
| DEG.twoside | Cells_Cultured_fibroblasts | 5399 | 8 | 0.589 | 1.000 | ENSG00000029153:ENSG00000110841:ENSG00000159314:ENSG00000238083:ENSG00000073969:ENSG00000108379:ENSG00000173482:ENSG00000099864 |
| DEG.twoside | Cells_EBV-transformed_lymphocytes | 5896 | 5 | 0.969 | 1.000 | ENSG00000035720:ENSG00000029153:ENSG00000165935:ENSG00000225190:ENSG00000170412 |
| DEG.twoside | Cervix_Ectocervix | 189 | 0 | 1.000 | 1.000 | |
| DEG.twoside | Cervix_Endocervix | 1728 | 2 | 0.747 | 1.000 | ENSG00000120071:ENSG00000099864 |
| DEG.twoside | Colon_Sigmoid | 2794 | 4 | 0.622 | 1.000 | ENSG00000104537:ENSG00000159314:ENSG00000108379:ENSG00000170412 |
| DEG.twoside | Colon_Transverse | 2532 | 5 | 0.328 | 1.000 | ENSG00000104537:ENSG00000029153:ENSG00000159314:ENSG00000186868:ENSG00000099812 |
| DEG.twoside | Esophagus_Gastroesophageal_Junction | 2422 | 5 | 0.294 | 1.000 | ENSG00000159314:ENSG00000186868:ENSG00000073969:ENSG00000108379:ENSG00000170412 |
| DEG.twoside | Esophagus_Mucosa | 4877 | 7 | 0.627 | 1.000 | ENSG00000029153:ENSG00000165935:ENSG00000159314:ENSG00000225190:ENSG00000170412:ENSG00000173482:ENSG00000099864 |
| DEG.twoside | Esophagus_Muscularis | 2625 | 6 | 0.191 | 1.000 | ENSG00000104537:ENSG00000159314:ENSG00000186868:ENSG00000073969:ENSG00000108379:ENSG00000170412 |
| DEG.twoside | Fallopian_Tube | 452 | 1 | 0.497 | 1.000 | ENSG00000099864 |

| | | | | | | |
|---|---|---|---|---|---|---|
| DEG.twoside | Heart_Atrial_Appendage | 7937 | 7 | 0.984 | 1.000 | ENSG00000159314:ENSG00000225190:ENSG00000186868:ENSG00000120071:ENSG00000238083:ENSG00000073969:ENSG00000170412 |
| DEG.twoside | Heart_Left_Ventricle | 8909 | 7 | 0.997 | 1.000 | ENSG00000159314:ENSG00000225190:ENSG00000186868:ENSG00000120071:ENSG00000073969:ENSG00000170412:ENSG00000099864 |
| DEG.twoside | Kidney_Cortex | 5997 | 7 | 0.847 | 1.000 | ENSG00000035720:ENSG00000225190:ENSG00000120071:ENSG00000073969:ENSG00000108379:ENSG00000170412:ENSG00000099812 |
| DEG.twoside | Kidney_Medulla | 93 | 1 | 0.131 | 1.000 | ENSG00000170412 |
| DEG.twoside | Liver | 8286 | 13 | 0.487 | 1.000 | ENSG00000104537:ENSG00000110841:ENSG00000100652:ENSG00000159314:ENSG00000225190:ENSG00000120071:ENSG00000238083:ENSG00000073969:ENSG00000108379:ENSG00000170412:ENSG00000173482:ENSG00000099864:ENSG00000186732 |
| DEG.twoside | Lung | 3849 | 6 | 0.533 | 1.000 | ENSG00000110841:ENSG00000159314:ENSG00000186868:ENSG00000108379:ENSG00000173482:ENSG00000099812 |
| DEG.twoside | Minor_Salivary_Gland | 2951 | 4 | 0.668 | 1.000 | ENSG00000029153:ENSG00000159314:ENSG00000173482:ENSG00000099864 |
| DEG.twoside | Muscle_Skeletal | 6972 | 7 | 0.945 | 1.000 | ENSG00000110841:ENSG00000159314:ENSG00000186868:ENSG00000120071:ENSG00000238083:ENSG00000170412:ENSG00000099864 |
| DEG.twoside | Nerve_Tibial | 4941 | 5 | 0.900 | 1.000 | ENSG00000175161:ENSG00000029153:ENSG00000186868:ENSG00000108379:ENSG00000099864 |
| DEG.twoside | Ovary | 5103 | 8 | 0.514 | 1.000 | ENSG00000110841:ENSG00000159314:ENSG00000186868:ENSG00000120071:ENSG00000073969:ENSG00000108379:ENSG00000170412:ENSG00000099864 |

| | | | | | | |
|---|---|---|---|---|---|---|
| DEG.twoside | Pancreas | 9220 | 8 | 0.993 | 1.000 | ENSG00000110841:ENSG00000159314:ENSG00000225190:ENSG00000120071:ENSG00000238083:ENSG00000073969:ENSG00000173482:ENSG00000099864 |
| DEG.twoside | Pituitary | 5005 | 8 | 0.489 | 1.000 | ENSG00000029153:ENSG00000120088:ENSG00000186868:ENSG00000238083:ENSG00000073969:ENSG00000170412:ENSG00000099864:ENSG00000099812 |
| DEG.twoside | Prostate | 2717 | 4 | 0.598 | 1.000 | ENSG00000029153:ENSG00000108379:ENSG00000170412:ENSG00000099812 |
| DEG.twoside | Skin_Not_Sun_Exposed_Suprapubic | 4367 | 7 | 0.494 | 1.000 | ENSG00000029153:ENSG00000159314:ENSG00000120088:ENSG00000186868:ENSG00000108379:ENSG00000173482:ENSG00000099812 |
| DEG.twoside | Skin_Sun_Exposed_Lower_leg | 4286 | 6 | 0.650 | 1.000 | ENSG00000029153:ENSG00000159314:ENSG00000225190:ENSG00000120071:ENSG00000108379:ENSG00000099812 |
| DEG.twoside | Small_Intestine_Terminal_Ileum | 3112 | 6 | 0.320 | 1.000 | ENSG00000175161:ENSG00000035720:ENSG00000104537:ENSG00000159314:ENSG00000186868:ENSG00000099812 |
| DEG.twoside | Spleen | 4596 | 3 | 0.982 | 1.000 | ENSG00000035720:ENSG00000159314:ENSG00000108379 |
| DEG.twoside | Stomach | 3070 | 6 | 0.308 | 1.000 | ENSG00000175161:ENSG00000159314:ENSG00000186868:ENSG00000170412:ENSG00000099864:ENSG00000099812 |
| DEG.twoside | Testis | 8546 | 12 | 0.694 | 1.000 | ENSG00000035720:ENSG00000104537:ENSG00000165935:ENSG00000110841:ENSG00000225190:ENSG00000185294:ENSG00000120071:ENSG00000176681:ENSG00000238083:ENSG00000108379:ENSG00000170412:ENSG00000099812 |
| DEG.twoside | Thyroid | 4708 | 5 | 0.872 | 1.000 | ENSG00000110841:ENSG00000186868:ENSG00000120071:ENSG00000170412:ENSG00000173482 |

| | | | | | | |
|---|---|---|---|---|---|---|
| DEG.twoside | Uterus | 4787 | 9 | 0.275 | 1.000 | ENSG00000104537:ENSG00000029153:ENSG00000120088:ENSG00000186868:ENSG00000120071:ENSG00000238083:ENSG00000108379:ENSG00000173482:ENSG00000099864 |
| DEG.twoside | Vagina | 2611 | 5 | 0.353 | 1.000 | ENSG00000029153:ENSG00000159314:ENSG00000225190:ENSG00000186868:ENSG00000120071 |
| DEG.twoside | Whole_Blood | 7620 | 5 | 0.998 | 1.000 | ENSG00000035720:ENSG00000159314:ENSG00000225190:ENSG00000120071:ENSG00000073969 |

***Table S21. Tissue Specificity of Prioritised Genes from FUMA (using GTEx v8 54 Tissue Types): Learning & Memory (One-Card Learning Task).***

| Category | GeneSet | N_genes | N_overlap | p | adjP | genes |
|---|---|---|---|---|---|---|
| DEG.up | Adipose_Subcutaneous | 1689 | 1 | 0.906 | 1.000 | ENSG00000118508 |
| DEG.up | Adipose_Visceral_Omentum | 1428 | 1 | 0.863 | 1.000 | ENSG00000175745 |
| DEG.up | Adrenal_Gland | 1204 | 1 | 0.810 | 1.000 | ENSG00000175745 |
| DEG.up | Artery_Aorta | 2103 | 3 | 0.544 | 1.000 | ENSG00000118508:ENSG00000118058:ENSG00000102780 |
| DEG.up | Artery_Coronary | 1593 | 2 | 0.640 | 1.000 | ENSG00000118508:ENSG00000102780 |
| DEG.up | Artery_Tibial | 2175 | 6 | 0.061 | 1.000 | ENSG00000118655:ENSG00000113391:ENSG00000118508:ENSG00000118058:ENSG00000102780:ENSG00000023516 |
| DEG.up | Bladder | 728 | 1 | 0.629 | 1.000 | ENSG00000175745 |
| DEG.up | Brain_Amygdala | 1438 | 1 | 0.865 | 1.000 | ENSG00000175745 |
| DEG.up | Brain_Anterior_cingulate_cortex_BA24 | 1866 | 2 | 0.727 | 1.000 | ENSG00000175745:ENSG00000255384 |
| DEG.up | Brain_Caudate_basal_ganglia | 1703 | 0 | 1.000 | 1.000 | |
| DEG.up | Brain_Cerebellar_Hemisphere | 4248 | 14 | 0.000 | 0.015 | ENSG00000081026:ENSG00000116793:ENSG00000081019:ENSG00000188761:ENSG00000134262:ENSG00000118655:ENSG00000175745:ENSG00000113391:ENSG00000164209:ENSG00000110344:ENSG00000255384:ENSG00000118058:ENSG00000102780:ENSG00000023516 |
| DEG.up | Brain_Cerebellum | 4055 | 13 | 0.001 | 0.040 | ENSG00000081026:ENSG00000116793:ENSG00000081019:ENSG00000134262:ENSG00000118655:ENSG00000175745:ENSG00000113391:ENSG00000164209:ENSG00000110344:ENSG00000255384:ENSG00000118058:ENSG00000102780:ENSG00000023516 |
| DEG.up | Brain_Cortex | 2159 | 2 | 0.801 | 1.000 | ENSG00000175745:ENSG00000255384 |

| | | | | | | |
|---|---|---|---|---|---|---|
| DEG.up | Brain_Frontal_Cortex_BA9 | 2422 | 3 | 0.645 | 1.000 | ENSG00000175745:ENSG00000255384:ENSG00000023516 |
| DEG.up | Brain_Hippocampus | 1542 | 0 | 1.000 | 1.000 | |
| DEG.up | Brain_Hypothalamus | 1979 | 1 | 0.939 | 1.000 | ENSG00000164512 |
| DEG.up | Brain_Nucleus_accumbens_basal_ganglia | 1834 | 3 | 0.448 | 1.000 | ENSG00000164512:ENSG00000255384:ENSG00000023516 |
| DEG.up | Brain_Putamen_basal_ganglia | 1346 | 0 | 1.000 | 1.000 | |
| DEG.up | Brain_Spinal_cord_cervical_c-1 | 1611 | 1 | 0.895 | 1.000 | ENSG00000023516 |
| DEG.up | Brain_Substantia_nigra | 1277 | 0 | 1.000 | 1.000 | |
| DEG.up | Breast_Mammary_Tissue | 1428 | 0 | 1.000 | 1.000 | |
| DEG.up | Cells_Cultured_fibroblasts | 3686 | 8 | 0.100 | 1.000 | ENSG00000134242:ENSG00000118655:ENSG00000175745:ENSG00000113391:ENSG00000164209:ENSG00000118508:ENSG00000110344:ENSG00000023516 |
| DEG.up | Cells_EBV-transformed_lymphocytes | 3833 | 9 | 0.052 | 1.000 | ENSG00000116793:ENSG00000081019:ENSG00000134242:ENSG00000134262:ENSG00000118655:ENSG00000164209:ENSG00000110344:ENSG00000118058:ENSG00000023516 |
| DEG.up | Cervix_Ectocervix | 187 | 0 | 1.000 | 1.000 | |
| DEG.up | Cervix_Endocervix | 1723 | 4 | 0.193 | 1.000 | ENSG00000081019:ENSG00000175745:ENSG00000113391:ENSG00000118058 |
| DEG.up | Colon_Sigmoid | 1264 | 0 | 1.000 | 1.000 | |
| DEG.up | Colon_Transverse | 1128 | 3 | 0.188 | 1.000 | ENSG00000188761:ENSG00000118508:ENSG00000101076 |
| DEG.up | Esophagus_Gastroesophageal_Junction | 1016 | 1 | 0.752 | 1.000 | ENSG00000102780 |
| DEG.up | Esophagus_Mucosa | 1755 | 1 | 0.915 | 1.000 | ENSG00000188761 |
| DEG.up | Esophagus_Muscularis | 1043 | 0 | 1.000 | 1.000 | |
| DEG.up | Fallopian_Tube | 451 | 1 | 0.456 | 1.000 | ENSG00000175745 |
| DEG.up | Heart_Atrial_Appendage | 572 | 1 | 0.540 | 1.000 | ENSG00000163492 |
| DEG.up | Heart_Left_Ventricle | 395 | 1 | 0.413 | 1.000 | ENSG00000163492 |

| | | | | | | |
|---|---|---|---|---|---|---|
| DEG.up | Kidney_Cortex | 948 | 1 | 0.727 | 1.000 | ENSG00000101076 |
| DEG.up | Kidney_Medulla | 93 | 0 | 1.000 | 1.000 | |
| DEG.up | Liver | 1090 | 1 | 0.777 | 1.000 | ENSG00000101076 |
| DEG.up | Lung | 3029 | 6 | 0.206 | 1.000 | ENSG00000081026:ENSG00000134242:ENSG00000175745:ENSG00000118508:ENSG00000160654:ENSG00000102780 |
| DEG.up | Minor_Salivary_Gland | 1285 | 0 | 1.000 | 1.000 | |
| DEG.up | Muscle_Skeletal | 879 | 0 | 1.000 | 1.000 | |
| DEG.up | Nerve_Tibial | 4157 | 7 | 0.310 | 1.000 | ENSG00000134262:ENSG00000175745:ENSG00000113391:ENSG00000118508:ENSG00000110344:ENSG00000118058:ENSG00000023516 |
| DEG.up | Ovary | 3674 | 4 | 0.752 | 1.000 | ENSG00000081019:ENSG00000134262:ENSG00000175745:ENSG00000118058 |
| DEG.up | Pancreas | 549 | 2 | 0.165 | 1.000 | ENSG00000188761:ENSG00000101076 |
| DEG.up | Pituitary | 3253 | 4 | 0.654 | 1.000 | ENSG00000081026:ENSG00000116793:ENSG00000134262:ENSG00000175745 |
| DEG.up | Prostate | 2019 | 1 | 0.943 | 1.000 | ENSG00000175745 |
| DEG.up | Skin_Not_Sun_Exposed_Suprapubic | 2037 | 0 | 1.000 | 1.000 | |
| DEG.up | Skin_Sun_Exposed_Lower_leg | 2163 | 0 | 1.000 | 1.000 | |
| DEG.up | Small_Intestine_Terminal_Ileum | 1855 | 5 | 0.094 | 1.000 | ENSG00000134242:ENSG00000188761:ENSG00000164512:ENSG00000160654:ENSG00000101076 |
| DEG.up | Spleen | 2666 | 5 | 0.277 | 1.000 | ENSG00000134242:ENSG00000134262:ENSG00000164512:ENSG00000118508:ENSG00000160654 |
| DEG.up | Stomach | 780 | 2 | 0.279 | 1.000 | ENSG00000188761:ENSG00000101076 |
| DEG.up | Testis | 5947 | 6 | 0.855 | 1.000 | ENSG00000116793:ENSG00000134262:ENSG00000164512:ENSG00000248483:ENSG00000185261:ENSG00000118508 |
| DEG.up | Thyroid | 3732 | 3 | 0.901 | 1.000 | ENSG00000081026:ENSG00000134262:ENSG00000175745 |

| | | | | | | |
|---|---|---|---|---|---|---|
| DEG.up | Uterus | 3999 | 9 | 0.066 | 1.000 | ENSG00000081019:ENSG00000134262:ENSG00000175745:ENSG00000113391:ENSG00000118508:ENSG00000110344:ENSG00000118058:ENSG00000102780:ENSG00000023516 |
| DEG.up | Vagina | 2013 | 2 | 0.767 | 1.000 | ENSG00000175745:ENSG00000102780 |
| DEG.up | Whole_Blood | 1299 | 4 | 0.091 | 1.000 | ENSG00000134242:ENSG00000164512:ENSG00000118508:ENSG00000160654 |
| DEG.down | Adipose_Subcutaneous | 1307 | 1 | 0.836 | 1.000 | ENSG00000175745 |
| DEG.down | Adipose_Visceral_Omentum | 1400 | 0 | 1.000 | 1.000 | |
| DEG.down | Adrenal_Gland | 2994 | 4 | 0.583 | 1.000 | ENSG00000081026:ENSG00000118655:ENSG00000118058:ENSG00000102780 |
| DEG.down | Artery_Aorta | 1323 | 1 | 0.840 | 1.000 | ENSG00000175745 |
| DEG.down | Artery_Coronary | 863 | 1 | 0.693 | 1.000 | ENSG00000175745 |
| DEG.down | Artery_Tibial | 1746 | 2 | 0.691 | 1.000 | ENSG00000134242:ENSG00000175745 |
| DEG.down | Bladder | 103 | 0 | 1.000 | 1.000 | |
| DEG.down | Brain_Amygdala | 6995 | 9 | 0.630 | 1.000 | ENSG00000116793:ENSG00000081019:ENSG00000134262:ENSG00000118655:ENSG00000164209:ENSG00000118508:ENSG00000110344:ENSG00000118058:ENSG00000102780 |
| DEG.down | Brain_Anterior_cingulate_cortex_BA24 | 6003 | 7 | 0.738 | 1.000 | ENSG00000081019:ENSG00000134262:ENSG00000118655:ENSG00000118508:ENSG00000110344:ENSG00000118058:ENSG00000102780 |
| DEG.down | Brain_Caudate_basal_ganglia | 6174 | 7 | 0.769 | 1.000 | ENSG00000081026:ENSG00000081019:ENSG00000118655:ENSG00000164209:ENSG00000118508:ENSG00000110344:ENSG00000118058 |
| DEG.down | Brain_Cerebellar_Hemisphere | 2381 | 1 | 0.967 | 1.000 | ENSG00000118508 |

| | | | | | | |
|---|---|---|---|---|---|---|
| DEG.down | Brain_Cerebellum | 2483 | 1 | 0.972 | 1.000 | ENSG00000118508 |
| DEG.down | Brain_Cortex | 4957 | 5 | 0.832 | 1.000 | ENSG00000081019:ENSG00000118655:ENSG00000118508:ENSG00000110344:ENSG00000102780 |
| DEG.down | Brain_Frontal_Cortex_BA9 | 4429 | 4 | 0.877 | 1.000 | ENSG00000118655:ENSG00000118508:ENSG00000110344:ENSG00000102780 |
| DEG.down | Brain_Hippocampus | 6806 | 8 | 0.743 | 1.000 | ENSG00000081026:ENSG00000081019:ENSG00000134262:ENSG00000118655:ENSG00000118508:ENSG00000110344:ENSG00000118058:ENSG00000102780 |
| DEG.down | Brain_Hypothalamus | 5303 | 6 | 0.754 | 1.000 | ENSG00000081019:ENSG00000118655:ENSG00000118508:ENSG00000110344:ENSG00000118058:ENSG00000102780 |
| DEG.down | Brain_Nucleus_accumbens_basal_ganglia | 5801 | 5 | 0.927 | 1.000 | ENSG00000081019:ENSG00000118655:ENSG00000118508:ENSG00000110344:ENSG00000118058 |
| DEG.down | Brain_Putamen_basal_ganglia | 7081 | 9 | 0.648 | 1.000 | ENSG00000081026:ENSG00000116793:ENSG00000081019:ENSG00000134262:ENSG00000118655:ENSG00000164209:ENSG00000118508:ENSG00000110344:ENSG00000118058 |
| DEG.down | Brain_Spinal_cord_cervical_c-1 | 4547 | 5 | 0.763 | 1.000 | ENSG00000116793:ENSG00000081019:ENSG00000118508:ENSG00000118058:ENSG00000102780 |
| DEG.down | Brain_Substantia_nigra | 6489 | 8 | 0.682 | 1.000 | ENSG00000116793:ENSG00000081019:ENSG00000134262:ENSG00000118655:ENSG00000118508:ENSG00000110344:ENSG00000118058:ENSG00000102780 |
| DEG.down | Breast_Mammary_Tissue | 912 | 0 | 1.000 | 1.000 | |
| DEG.down | Cells_Cultured_fibroblasts | 1713 | 0 | 1.000 | 1.000 | |
| DEG.down | Cells_EBV-transformed_lymphocytes | 2063 | 1 | 0.946 | 1.000 | ENSG00000118508 |
| DEG.down | Cervix_Ectocervix | 2 | 0 | 1.000 | 1.000 | |
| DEG.down | Cervix_Endocervix | 5 | 0 | 1.000 | 1.000 | |

| | | | | | | |
|---|---|---|---|---|---|---|
| DEG.down | Colon_Sigmoid | 1530 | 1 | 0.882 | 1.000 | ENSG00000134242 |
| DEG.down | Colon_Transverse | 1404 | 1 | 0.858 | 1.000 | ENSG00000175745 |
| DEG.down | Esophagus_Gastroesophageal_Junction | 1406 | 1 | 0.858 | 1.000 | ENSG00000116793 |
| DEG.down | Esophagus_Mucosa | 3122 | 3 | 0.812 | 1.000 | ENSG00000116793:ENSG00000118508:ENSG00000118058 |
| DEG.down | Esophagus_Muscularis | 1582 | 1 | 0.890 | 1.000 | ENSG00000116793 |
| DEG.down | Fallopian_Tube | 1 | 0 | 1.000 | 1.000 | |
| DEG.down | Heart_Atrial_Appendage | 7365 | 12 | 0.241 | 1.000 | ENSG00000081026:ENSG00000116793:ENSG00000081019:ENSG00000134262:ENSG00000118655:ENSG00000113391:ENSG00000164209:ENSG00000118508:ENSG00000110344:ENSG00000118058:ENSG00000102780:ENSG00000023516 |
| DEG.down | Heart_Left_Ventricle | 8514 | 11 | 0.635 | 1.000 | ENSG00000081026:ENSG00000116793:ENSG00000081019:ENSG00000134262:ENSG00000113391:ENSG00000164209:ENSG00000118508:ENSG00000110344:ENSG00000118058:ENSG00000102780:ENSG00000023516 |
| DEG.down | Kidney_Cortex | 5049 | 10 | 0.107 | 1.000 | ENSG00000116793:ENSG00000081019:ENSG00000134262:ENSG00000118655:ENSG00000113391:ENSG00000164209:ENSG00000110344:ENSG00000118058:ENSG00000102780:ENSG00000023516 |
| DEG.down | Kidney_Medulla | 0 | 0 | 1.000 | 1.000 | |
| DEG.down | Liver | 7196 | 11 | 0.349 | 1.000 | ENSG00000081026:ENSG00000116793:ENSG00000081019:ENSG00000134262:ENSG00000118655:ENSG00000175745:ENSG00000113391:ENSG00000164209:ENSG00000110344:ENSG00000118058:ENSG00000023516 |
| DEG.down | Lung | 820 | 0 | 1.000 | 1.000 | |

| | | | | | | |
|---|---|---|---|---|---|---|
| DEG.down | Minor_Salivary_Gland | 1666 | 3 | 0.386 | 1.000 | ENSG00000116793:ENSG00000134242:ENSG00000175745 |
| DEG.down | Muscle_Skeletal | 6093 | 8 | 0.598 | 1.000 | ENSG00000081026:ENSG00000116793:ENSG00000134262:ENSG00000113391:ENSG00000118508:ENSG00000118058:ENSG00000102780:ENSG00000023516 |
| DEG.down | Nerve_Tibial | 784 | 0 | 1.000 | 1.000 | |
| DEG.down | Ovary | 1429 | 0 | 1.000 | 1.000 | |
| DEG.down | Pancreas | 8671 | 10 | 0.800 | 1.000 | ENSG00000116793:ENSG00000081019:ENSG00000134262:ENSG00000175745:ENSG00000113391:ENSG00000164209:ENSG00000118508:ENSG00000110344:ENSG00000118058:ENSG00000023516 |
| DEG.down | Pituitary | 1752 | 2 | 0.693 | 1.000 | ENSG00000118655:ENSG00000118508 |
| DEG.down | Prostate | 698 | 0 | 1.000 | 1.000 | |
| DEG.down | Skin_Not_Sun_Exposed_Suprapubic | 2330 | 3 | 0.617 | 1.000 | ENSG00000081026:ENSG00000116793:ENSG00000175745 |
| DEG.down | Skin_Sun_Exposed_Lower_leg | 2123 | 3 | 0.550 | 1.000 | ENSG00000081026:ENSG00000116793:ENSG00000175745 |
| DEG.down | Small_Intestine_Terminal_Ileum | 1257 | 0 | 1.000 | 1.000 | |
| DEG.down | Spleen | 1930 | 2 | 0.745 | 1.000 | ENSG00000081026:ENSG00000102780 |
| DEG.down | Stomach | 2290 | 1 | 0.962 | 1.000 | ENSG00000118655 |
| DEG.down | Testis | 2599 | 4 | 0.464 | 1.000 | ENSG00000134242:ENSG00000175745:ENSG00000113391:ENSG00000102780 |
| DEG.down | Thyroid | 976 | 0 | 1.000 | 1.000 | |
| DEG.down | Uterus | 788 | 1 | 0.659 | 1.000 | ENSG00000134242 |

| | | | | | | |
|---|---|---|---|---|---|---|
| DEG.down | Vagina | 598 | 0 | 1.000 | 1.000 | |
| DEG.down | Whole_Blood | 6321 | 5 | 0.959 | 1.000 | ENSG00000113391:ENSG00000164209:ENSG00000110344:ENSG00000118058:ENSG00000023516 |
| DEG.twoside | Adipose_Subcutaneous | 2996 | 2 | 0.927 | 1.000 | ENSG00000175745:ENSG00000118508 |
| DEG.twoside | Adipose_Visceral_Omentum | 2828 | 1 | 0.984 | 1.000 | ENSG00000175745 |
| DEG.twoside | Adrenal_Gland | 4198 | 5 | 0.690 | 1.000 | ENSG00000081026:ENSG00000118655:ENSG00000175745:ENSG00000118058:ENSG00000102780 |
| DEG.twoside | Artery_Aorta | 3426 | 4 | 0.697 | 1.000 | ENSG00000175745:ENSG00000118508:ENSG00000118058:ENSG00000102780 |
| DEG.twoside | Artery_Coronary | 2456 | 3 | 0.655 | 1.000 | ENSG00000175745:ENSG00000118508:ENSG00000102780 |
| DEG.twoside | Artery_Tibial | 3921 | 8 | 0.132 | 1.000 | ENSG00000134242:ENSG00000118655:ENSG00000175745:ENSG00000113391:ENSG00000118508:ENSG00000118058:ENSG00000102780:ENSG00000023516 |
| DEG.twoside | Bladder | 831 | 1 | 0.679 | 1.000 | ENSG00000175745 |
| DEG.twoside | Brain_Amygdala | 8433 | 10 | 0.761 | 1.000 | ENSG00000116793:ENSG00000081019:ENSG00000134262:ENSG00000118655:ENSG00000175745:ENSG00000164209:ENSG00000118508:ENSG00000110344:ENSG00000118058:ENSG00000102780 |
| DEG.twoside | Brain_Anterior_cingulate_cortex_BA24 | 7869 | 9 | 0.793 | 1.000 | ENSG00000081019:ENSG00000134262:ENSG00000118655:ENSG00000175745:ENSG00000118508:ENSG00000110344:ENSG00000255384:ENSG00000118058:ENSG00000102780 |
| DEG.twoside | Brain_Caudate_basal_ganglia | 7877 | 7 | 0.953 | 1.000 | ENSG00000081026:ENSG00000081019:ENSG00000118655:ENSG00000164209:ENSG00000118508:ENSG00000110344:ENSG00000118058 |
| DEG.twoside | Brain_Cerebellar_Hemisphere | 6629 | 15 | 0.009 | 0.506 | ENSG00000081026:ENSG00000116793:ENSG00000081019:ENSG00000188761:ENSG00000134262:ENSG00000118655:ENSG00000175745:ENSG00000113391:ENSG |

| | | | | | | |
|---|---|---|---|---|---|---|
| | | | | | | 00000164209:ENSG00000118508:ENSG00000110344:ENSG00000255384:ENSG00000118058:ENSG00000102780:ENSG00000023516 |
| DEG.two side | Brain_Cerebellum | 6538 | 14 | 0.023 | 1.000 | ENSG00000081026:ENSG00000116793:ENSG00000081019:ENSG00000134262:ENSG00000118655:ENSG00000175745:ENSG00000113391:ENSG00000164209:ENSG00000118508:ENSG00000110344:ENSG00000255384:ENSG00000118058:ENSG00000102780:ENSG00000023516 |
| DEG.two side | Brain_Cortex | 7116 | 7 | 0.896 | 1.000 | ENSG00000081019:ENSG00000118655:ENSG00000175745:ENSG00000118508:ENSG00000110344:ENSG00000255384:ENSG00000102780 |
| DEG.two side | Brain_Frontal_Cortex_BA9 | 6851 | 7 | 0.867 | 1.000 | ENSG00000118655:ENSG00000175745:ENSG00000118508:ENSG00000110344:ENSG00000255384:ENSG00000102780:ENSG00000023516 |
| DEG.two side | Brain_Hippocampus | 8348 | 8 | 0.933 | 1.000 | ENSG00000081026:ENSG00000081019:ENSG00000134262:ENSG00000118655:ENSG00000118508:ENSG00000110344:ENSG00000118058:ENSG00000102780 |
| DEG.two side | Brain_Hypothalamus | 7282 | 7 | 0.912 | 1.000 | ENSG00000081019:ENSG00000118655:ENSG00000164512:ENSG00000118508:ENSG00000110344:ENSG00000118058:ENSG00000102780 |
| DEG.two side | Brain_Nucleus_accumbens_basal_ganglia | 7635 | 8 | 0.867 | 1.000 | ENSG00000081019:ENSG00000118655:ENSG00000164512:ENSG00000118508:ENSG00000110344:ENSG00000255384:ENSG00000118058:ENSG00000023516 |
| DEG.two side | Brain_Putamen_basal_ganglia | 8427 | 9 | 0.869 | 1.000 | ENSG00000081026:ENSG00000116793:ENSG00000081019:ENSG00000134262:ENSG00000118655:ENSG00000164209:ENSG00000118508:ENSG00000110344:ENSG00000118058 |
| DEG.two side | Brain_Spinal_cord_cervical_c-1 | 6158 | 6 | 0.881 | 1.000 | ENSG00000116793:ENSG00000081019:ENSG00000118508:ENSG00000118058:ENSG00000102780:ENSG00000023516 |

| | | | | | | |
|---|---|---|---|---|---|---|
| DEG.two side | Brain_Substantia_nigra | 7766 | 8 | 0.882 | 1.000 | ENSG00000116793:ENSG00000081019:ENSG00000134262:ENSG00000118655:ENSG00000118508:ENSG00000110344:ENSG00000118058:ENSG00000102780 |
| DEG.two side | Breast_Mammary_Tissue | 2340 | 0 | 1.000 | 1.000 | |
| DEG.two side | Cells_Cultured_fibroblasts | 5399 | 8 | 0.435 | 1.000 | ENSG00000134242:ENSG00000118655:ENSG00000175745:ENSG00000113391:ENSG00000164209:ENSG00000118508:ENSG00000110344:ENSG00000023516 |
| DEG.two side | Cells_EBV-transformed_lymphocytes | 5896 | 10 | 0.235 | 1.000 | ENSG00000116793:ENSG00000081019:ENSG00000134242:ENSG00000134262:ENSG00000118655:ENSG00000164209:ENSG00000118508:ENSG00000110344:ENSG00000118058:ENSG00000023516 |
| DEG.two side | Cervix_Ectocervix | 189 | 0 | 1.000 | 1.000 | |
| DEG.two side | Cervix_Endocervix | 1728 | 4 | 0.194 | 1.000 | ENSG00000081019:ENSG00000175745:ENSG00000113391:ENSG00000118058 |
| DEG.two side | Colon_Sigmoid | 2794 | 1 | 0.983 | 1.000 | ENSG00000134242 |
| DEG.two side | Colon_Transverse | 2532 | 4 | 0.443 | 1.000 | ENSG00000188761:ENSG00000175745:ENSG00000118508:ENSG00000101076 |
| DEG.two side | Esophagus_Gastroesophageal_Junction | 2422 | 2 | 0.853 | 1.000 | ENSG00000116793:ENSG00000102780 |
| DEG.two side | Esophagus_Mucosa | 4877 | 4 | 0.923 | 1.000 | ENSG00000116793:ENSG00000188761:ENSG00000118508:ENSG00000118058 |
| DEG.two side | Esophagus_Muscularis | 2625 | 1 | 0.977 | 1.000 | ENSG00000116793 |
| DEG.two side | Fallopian_Tube | 452 | 1 | 0.457 | 1.000 | ENSG00000175745 |
| DEG.two side | Heart_Atrial_Appendage | 7937 | 13 | 0.215 | 1.000 | ENSG00000081026:ENSG00000116793:ENSG00000081019:ENSG00000134262:ENSG00000118655:ENSG00000163492:ENSG00000113391:ENSG00000164209:ENSG00000118508:ENSG00000110344:ENSG00000118058:ENSG00000102780:ENSG00000023516 |

| | | | | | | |
|---|---|---|---|---|---|---|
| DEG.two side | Heart_Left_Ventricle | 8909 | 12 | 0.561 | 1.000 | ENSG00000081026:ENSG00000116793:ENSG00000081019:ENSG00000134262:ENSG00000163492:ENSG00000113391:ENSG00000164209:ENSG00000118508:ENSG00000110344:ENSG00000118058:ENSG00000102780:ENSG00000023516 |
| DEG.two side | Kidney_Cortex | 5997 | 11 | 0.140 | 1.000 | ENSG00000116793:ENSG00000081019:ENSG00000134262:ENSG00000118655:ENSG00000113391:ENSG00000164209:ENSG00000110344:ENSG00000118058:ENSG00000102780:ENSG00000023516:ENSG00000101076 |
| DEG.two side | Kidney_Medulla | 93 | 0 | 1.000 | 1.000 | |
| DEG.two side | Liver | 8286 | 12 | 0.425 | 1.000 | ENSG00000081026:ENSG00000116793:ENSG00000081019:ENSG00000134262:ENSG00000118655:ENSG00000175745:ENSG00000113391:ENSG00000164209:ENSG00000110344:ENSG00000118058:ENSG00000023516:ENSG00000101076 |
| DEG.two side | Lung | 3849 | 6 | 0.409 | 1.000 | ENSG00000081026:ENSG00000134242:ENSG00000175745:ENSG00000118508:ENSG00000160654:ENSG00000102780 |
| DEG.two side | Minor_Salivary_Gland | 2951 | 3 | 0.778 | 1.000 | ENSG00000116793:ENSG00000134242:ENSG00000175745 |
| DEG.two side | Muscle_Skeletal | 6972 | 8 | 0.772 | 1.000 | ENSG00000081026:ENSG00000116793:ENSG00000134262:ENSG00000113391:ENSG00000118508:ENSG00000118058:ENSG00000102780:ENSG00000023516 |
| DEG.two side | Nerve_Tibial | 4941 | 7 | 0.502 | 1.000 | ENSG00000134262:ENSG00000175745:ENSG00000113391:ENSG00000118508:ENSG00000110344:ENSG00000118058:ENSG00000023516 |
| DEG.two side | Ovary | 5103 | 4 | 0.940 | 1.000 | ENSG00000081019:ENSG00000134262:ENSG00000175745:ENSG00000118058 |
| DEG.two side | Pancreas | 9220 | 12 | 0.628 | 1.000 | ENSG00000116793:ENSG00000081019:ENSG00000188761:ENSG00000134262:ENSG00000175745:ENSG00000113391:ENSG00000164209:ENSG00000118508:ENSG |

| | | | | | | |
|---|---|---|---|---|---|---|
| | | | | | | 00000110344:ENSG00000118058:ENSG00000023516:ENSG00000101076 |
| DEG.two side | Pituitary | 5005 | 6 | 0.695 | 1.000 | ENSG00000081026:ENSG00000116793:ENSG00000134262:ENSG00000118655:ENSG00000175745:ENSG00000118508 |
| DEG.two side | Prostate | 2717 | 1 | 0.980 | 1.000 | ENSG00000175745 |
| DEG.two side | Skin_Not_Sun_Exposed_Suprapubic | 4367 | 3 | 0.953 | 1.000 | ENSG00000081026:ENSG00000116793:ENSG00000175745 |
| DEG.two side | Skin_Sun_Exposed_Lower_leg | 4286 | 3 | 0.948 | 1.000 | ENSG00000081026:ENSG00000116793:ENSG00000175745 |
| DEG.two side | Small_Intestine_Terminal_Ileum | 3112 | 5 | 0.403 | 1.000 | ENSG00000134242:ENSG00000188761:ENSG00000164512:ENSG00000160654:ENSG00000101076 |
| DEG.two side | Spleen | 4596 | 7 | 0.417 | 1.000 | ENSG00000081026:ENSG00000134242:ENSG00000134262:ENSG00000164512:ENSG00000118508:ENSG00000160654:ENSG00000102780 |
| DEG.two side | Stomach | 3070 | 3 | 0.802 | 1.000 | ENSG00000188761:ENSG00000118655:ENSG00000101076 |
| DEG.two side | Testis | 8546 | 10 | 0.780 | 1.000 | ENSG00000116793:ENSG00000134242:ENSG00000134262:ENSG00000164512:ENSG00000175745:ENSG00000113391:ENSG00000248483:ENSG00000185261:ENSG00000118508:ENSG00000102780 |
| DEG.two side | Thyroid | 4708 | 3 | 0.970 | 1.000 | ENSG00000081026:ENSG00000134262:ENSG00000175745 |
| DEG.two side | Uterus | 4787 | 10 | 0.079 | 1.000 | ENSG00000081019:ENSG00000134242:ENSG00000134262:ENSG00000175745:ENSG00000113391:ENSG00000118508:ENSG00000110344:ENSG00000118058:ENSG00000102780:ENSG00000023516 |
| DEG.two side | Vagina | 2611 | 2 | 0.882 | 1.000 | ENSG00000175745:ENSG00000102780 |
| DEG.two side | Whole_Blood | 7620 | 9 | 0.752 | 1.000 | ENSG00000134242:ENSG00000164512:ENSG00000113391:ENSG00000164209:ENSG00000118508:ENSG0000 |

| | | | | | | |
|---|---|---|---|---|---|---|
| | | | | | | 0160654:ENSG00000110344:ENSG00000118058:ENSG 00000023516 |

***Table S22. Tissue Specificity of Prioritised Genes from FUMA (using GTEx v8 54 Tissue Types): Working Memory (One-Back Task).***

| Category | GeneSet | N_genes | N_overlap | p | adjP | genes |
|---|---|---|---|---|---|---|
| DEG.up | Adipose_Subcutaneous | 1689 | 1 | 0.662 | 1.000 | ENSG00000135678 |
| DEG.up | Adipose_Visceral_Omentum | 1428 | 1 | 0.597 | 1.000 | ENSG00000135678 |
| DEG.up | Adrenal_Gland | 1204 | 0 | 1.000 | 1.000 | |
| DEG.up | Artery_Aorta | 2103 | 1 | 0.745 | 1.000 | ENSG00000131018 |
| DEG.up | Artery_Coronary | 1593 | 0 | 1.000 | 1.000 | |
| DEG.up | Artery_Tibial | 2175 | 2 | 0.391 | 1.000 | ENSG00000071909:ENSG00000131018 |
| DEG.up | Bladder | 728 | 0 | 1.000 | 1.000 | |
| DEG.up | Brain_Amygdala | 1438 | 1 | 0.600 | 1.000 | ENSG00000160460 |
| DEG.up | Brain_Anterior_cingulate_cortex_BA24 | 1866 | 1 | 0.700 | 1.000 | ENSG00000160460 |
| DEG.up | Brain_Caudate_basal_ganglia | 1703 | 1 | 0.665 | 1.000 | ENSG00000160460 |
| DEG.up | Brain_Cerebellar_Hemisphere | 4248 | 2 | 0.773 | 1.000 | ENSG00000131018:ENSG00000160460 |
| DEG.up | Brain_Cerebellum | 4055 | 2 | 0.747 | 1.000 | ENSG00000131018:ENSG00000160460 |
| DEG.up | Brain_Cortex | 2159 | 1 | 0.755 | 1.000 | ENSG00000160460 |
| DEG.up | Brain_Frontal_Cortex_BA9 | 2422 | 1 | 0.796 | 1.000 | ENSG00000160460 |
| DEG.up | Brain_Hippocampus | 1542 | 1 | 0.627 | 1.000 | ENSG00000160460 |
| DEG.up | Brain_Hypothalamus | 1979 | 1 | 0.722 | 1.000 | ENSG00000160460 |
| DEG.up | Brain_Nucleus_accumbens_basal_ganglia | 1834 | 1 | 0.694 | 1.000 | ENSG00000160460 |
| DEG.up | Brain_Putamen_basal_ganglia | 1346 | 1 | 0.575 | 1.000 | ENSG00000160460 |
| DEG.up | Brain_Spinal_cord_cervical_c-1 | 1611 | 0 | 1.000 | 1.000 | |
| DEG.up | Brain_Substantia_nigra | 1277 | 1 | 0.555 | 1.000 | ENSG00000160460 |
| DEG.up | Breast_Mammary_Tissue | 1428 | 1 | 0.597 | 1.000 | ENSG00000135678 |
| DEG.up | Cells_Cultured_fibroblasts | 3686 | 1 | 0.920 | 1.000 | ENSG00000135678 |

| | | | | | | |
|---|---|---|---|---|---|---|
| DEG.up | Cells_EBV-transformed_lymphocytes | 3833 | 1 | 0.928 | 1.000 | ENSG00000071909 |
| DEG.up | Cervix_Ectocervix | 187 | 0 | 1.000 | 1.000 | |
| DEG.up | Cervix_Endocervix | 1723 | 0 | 1.000 | 1.000 | |
| DEG.up | Colon_Sigmoid | 1264 | 0 | 1.000 | 1.000 | |
| DEG.up | Colon_Transverse | 1128 | 0 | 1.000 | 1.000 | |
| DEG.up | Esophagus_Gastroesophageal_Junction | 1016 | 0 | 1.000 | 1.000 | |
| DEG.up | Esophagus_Mucosa | 1755 | 0 | 1.000 | 1.000 | |
| DEG.up | Esophagus_Muscularis | 1043 | 0 | 1.000 | 1.000 | |
| DEG.up | Fallopian_Tube | 451 | 0 | 1.000 | 1.000 | |
| DEG.up | Heart_Atrial_Appendage | 572 | 0 | 1.000 | 1.000 | |
| DEG.up | Heart_Left_Ventricle | 395 | 0 | 1.000 | 1.000 | |
| DEG.up | Kidney_Cortex | 948 | 1 | 0.449 | 1.000 | ENSG00000071909 |
| DEG.up | Kidney_Medulla | 93 | 0 | 1.000 | 1.000 | |
| DEG.up | Liver | 1090 | 0 | 1.000 | 1.000 | |
| DEG.up | Lung | 3029 | 2 | 0.575 | 1.000 | ENSG00000131018:ENSG00000135678 |
| DEG.up | Minor_Salivary_Gland | 1285 | 0 | 1.000 | 1.000 | |
| DEG.up | Muscle_Skeletal | 879 | 0 | 1.000 | 1.000 | |
| DEG.up | Nerve_Tibial | 4157 | 2 | 0.761 | 1.000 | ENSG00000184465:ENSG00000160460 |
| DEG.up | Ovary | 3674 | 2 | 0.690 | 1.000 | ENSG00000131018:ENSG00000184465 |
| DEG.up | Pancreas | 549 | 0 | 1.000 | 1.000 | |
| DEG.up | Pituitary | 3253 | 4 | 0.121 | 1.000 | ENSG00000071909:ENSG00000131018:ENSG00000160460:ENSG00000167578 |
| DEG.up | Prostate | 2019 | 1 | 0.730 | 1.000 | ENSG00000184465 |
| DEG.up | Skin_Not_Sun_Exposed_Suprapubic | 2037 | 1 | 0.733 | 1.000 | ENSG00000135678 |
| DEG.up | Skin_Sun_Exposed_Lower_leg | 2163 | 1 | 0.756 | 1.000 | ENSG00000135678 |
| DEG.up | Small_Intestine_Terminal_Ileum | 1855 | 0 | 1.000 | 1.000 | |
| DEG.up | Spleen | 2666 | 1 | 0.829 | 1.000 | ENSG00000167578 |

| | | | | | | |
|---|---|---|---|---|---|---|
| DEG.up | Stomach | 780 | 0 | 1.000 | 1.000 | |
| DEG.up | Testis | 5947 | 2 | 0.922 | 1.000 | ENSG00000071909:ENSG00000184465 |
| DEG.up | Thyroid | 3732 | 3 | 0.407 | 1.000 | ENSG00000071909:ENSG00000131018:ENSG00000184465 |
| DEG.up | Uterus | 3999 | 2 | 0.739 | 1.000 | ENSG00000131018:ENSG00000184465 |
| DEG.up | Vagina | 2013 | 1 | 0.729 | 1.000 | ENSG00000184465 |
| DEG.up | Whole_Blood | 1299 | 0 | 1.000 | 1.000 | |
| DEG.down | Adipose_Subcutaneous | 1307 | 1 | 0.564 | 1.000 | ENSG00000160460 |
| DEG.down | Adipose_Visceral_Omentum | 1400 | 1 | 0.590 | 1.000 | ENSG00000160460 |
| DEG.down | Adrenal_Gland | 2994 | 2 | 0.568 | 1.000 | ENSG00000131018:ENSG00000160460 |
| DEG.down | Artery_Aorta | 1323 | 2 | 0.191 | 1.000 | ENSG00000135678:ENSG00000160460 |
| DEG.down | Artery_Coronary | 863 | 1 | 0.418 | 1.000 | ENSG00000160460 |
| DEG.down | Artery_Tibial | 1746 | 2 | 0.290 | 1.000 | ENSG00000135678:ENSG00000160460 |
| DEG.down | Bladder | 103 | 0 | 1.000 | 1.000 | |
| DEG.down | Brain_Amygdala | 6995 | 3 | 0.865 | 1.000 | ENSG00000131018:ENSG00000184465:ENSG00000135678 |
| DEG.down | Brain_Anterior_cingulate_cortex_BA24 | 6003 | 2 | 0.925 | 1.000 | ENSG00000184465:ENSG00000135678 |
| DEG.down | Brain_Caudate_basal_ganglia | 6174 | 2 | 0.934 | 1.000 | ENSG00000184465:ENSG00000135678 |
| DEG.down | Brain_Cerebellar_Hemisphere | 2381 | 1 | 0.790 | 1.000 | ENSG00000135678 |
| DEG.down | Brain_Cerebellum | 2483 | 1 | 0.805 | 1.000 | ENSG00000135678 |

| | | | | | | |
|---|---|---|---|---|---|---|
| DEG.down | Brain_Cortex | 4957 | 2 | 0.850 | 1.000 | ENSG00000184465:ENSG00000135678 |
| DEG.down | Brain_Frontal_Cortex_BA9 | 4429 | 2 | 0.795 | 1.000 | ENSG00000184465:ENSG00000135678 |
| DEG.down | Brain_Hippocampus | 6806 | 2 | 0.959 | 1.000 | ENSG00000184465:ENSG00000135678 |
| DEG.down | Brain_Hypothalamus | 5303 | 2 | 0.880 | 1.000 | ENSG00000184465:ENSG00000135678 |
| DEG.down | Brain_Nucleus_accumbens_basal_ganglia | 5801 | 2 | 0.914 | 1.000 | ENSG00000184465:ENSG00000135678 |
| DEG.down | Brain_Putamen_basal_ganglia | 7081 | 2 | 0.967 | 1.000 | ENSG00000184465:ENSG00000135678 |
| DEG.down | Brain_Spinal_cord_cervical_c-1 | 4547 | 2 | 0.808 | 1.000 | ENSG00000131018:ENSG00000184465 |
| DEG.down | Brain_Substantia_nigra | 6489 | 3 | 0.820 | 1.000 | ENSG00000131018:ENSG00000184465:ENSG00000135678 |
| DEG.down | Breast_Mammary_Tissue | 912 | 1 | 0.436 | 1.000 | ENSG00000160460 |
| DEG.down | Cells_Cultured_fibroblasts | 1713 | 3 | 0.080 | 1.000 | ENSG00000131018:ENSG00000130024:ENSG00000167578 |
| DEG.down | Cells_EBV-transformed_lymphocytes | 2063 | 2 | 0.365 | 1.000 | ENSG00000131018:ENSG00000135678 |
| DEG.down | Cervix_Ectocervix | 2 | 0 | 1.000 | 1.000 | |
| DEG.down | Cervix_Endocervix | 5 | 0 | 1.000 | 1.000 | |
| DEG.down | Colon_Sigmoid | 1530 | 2 | 0.239 | 1.000 | ENSG00000135678:ENSG00000160460 |
| DEG.down | Colon_Transverse | 1404 | 2 | 0.210 | 1.000 | ENSG00000131018:ENSG00000160460 |
| DEG.down | Esophagus_Gastroesophageal_Junction | 1406 | 1 | 0.591 | 1.000 | ENSG00000160460 |

| | | | | | | |
|---|---|---|---|---|---|---|
| DEG.down | Esophagus_Mucosa | 3122 | 2 | 0.593 | 1.000 | ENSG00000131018:ENSG00000135678 |
| DEG.down | Esophagus_Muscularis | 1582 | 2 | 0.251 | 1.000 | ENSG00000135678:ENSG00000160460 |
| DEG.down | Fallopian_Tube | 1 | 0 | 1.000 | 1.000 | |
| DEG.down | Heart_Atrial_Appendage | 7365 | 6 | 0.267 | 1.000 | ENSG00000131018:ENSG00000184465:ENSG00000135678:ENSG00000160460:ENSG00000167578:ENSG00000269858 |
| DEG.down | Heart_Left_Ventricle | 8514 | 7 | 0.217 | 1.000 | ENSG00000131018:ENSG00000184465:ENSG00000130024:ENSG00000135678:ENSG00000160460:ENSG00000167578:ENSG00000269858 |
| DEG.down | Kidney_Cortex | 5049 | 1 | 0.973 | 1.000 | ENSG00000130024 |
| DEG.down | Kidney_Medulla | 0 | 0 | 1.000 | 1.000 | |
| DEG.down | Liver | 7196 | 5 | 0.467 | 1.000 | ENSG00000131018:ENSG00000184465:ENSG00000130024:ENSG00000167578:ENSG00000269858 |
| DEG.down | Lung | 820 | 0 | 1.000 | 1.000 | |
| DEG.down | Minor_Salivary_Gland | 1666 | 2 | 0.271 | 1.000 | ENSG00000131018:ENSG00000135678 |
| DEG.down | Muscle_Skeletal | 6093 | 6 | 0.130 | 1.000 | ENSG00000131018:ENSG00000184465:ENSG00000135678:ENSG00000160460:ENSG00000167578:ENSG00000269858 |
| DEG.down | Nerve_Tibial | 784 | 0 | 1.000 | 1.000 | |
| DEG.down | Ovary | 1429 | 0 | 1.000 | 1.000 | |
| DEG.down | Pancreas | 8671 | 6 | 0.451 | 1.000 | ENSG00000131018:ENSG00000184465:ENSG00000130024:ENSG00000135678:ENSG00000167578:ENSG00000269858 |

| | | | | | | |
|---|---|---|---|---|---|---|
| DEG.down | Pituitary | 1752 | 1 | 0.676 | 1.000 | ENSG00000135678 |
| DEG.down | Prostate | 698 | 1 | 0.353 | 1.000 | ENSG00000160460 |
| DEG.down | Skin_Not_Sun_Exposed_Suprapubic | 2330 | 1 | 0.783 | 1.000 | ENSG00000269858 |
| DEG.down | Skin_Sun_Exposed_Lower_leg | 2123 | 1 | 0.749 | 1.000 | ENSG00000160460 |
| DEG.down | Small_Intestine_Terminal_Ileum | 1257 | 1 | 0.549 | 1.000 | ENSG00000131018 |
| DEG.down | Spleen | 1930 | 0 | 1.000 | 1.000 | |
| DEG.down | Stomach | 2290 | 1 | 0.776 | 1.000 | ENSG00000160460 |
| DEG.down | Testis | 2599 | 1 | 0.820 | 1.000 | ENSG00000135678 |
| DEG.down | Thyroid | 976 | 1 | 0.459 | 1.000 | ENSG00000160460 |
| DEG.down | Uterus | 788 | 0 | 1.000 | 1.000 | |
| DEG.down | Vagina | 598 | 1 | 0.311 | 1.000 | ENSG00000160460 |
| DEG.down | Whole_Blood | 6321 | 3 | 0.802 | 1.000 | ENSG00000131018:ENSG00000130024:ENSG00000135678 |
| DEG.twoside | Adipose_Subcutaneous | 2996 | 2 | 0.569 | 1.000 | ENSG00000135678:ENSG00000160460 |
| DEG.twoside | Adipose_Visceral_Omentum | 2828 | 2 | 0.535 | 1.000 | ENSG00000135678:ENSG00000160460 |
| DEG.twoside | Adrenal_Gland | 4198 | 2 | 0.766 | 1.000 | ENSG00000131018:ENSG00000160460 |
| DEG.twoside | Artery_Aorta | 3426 | 3 | 0.351 | 1.000 | ENSG00000131018:ENSG00000135678:ENSG00000160460 |

| | | | | | | |
|---|---|---|---|---|---|---|
| DEG.two side | Artery_Coronary | 2456 | 1 | 0.801 | 1.000 | ENSG00000160460 |
| DEG.two side | Artery_Tibial | 3921 | 4 | 0.203 | 1.000 | ENSG00000071909:ENSG00000131018:ENSG00000135678:ENSG00000160460 |
| DEG.two side | Bladder | 831 | 0 | 1.000 | 1.000 | |
| DEG.two side | Brain_Amygdala | 8433 | 4 | 0.841 | 1.000 | ENSG00000131018:ENSG00000184465:ENSG00000135678:ENSG00000160460 |
| DEG.two side | Brain_Anterior_cingulate_cortex_BA24 | 7869 | 3 | 0.923 | 1.000 | ENSG00000184465:ENSG00000135678:ENSG00000160460 |
| DEG.two side | Brain_Caudate_basal_ganglia | 7877 | 3 | 0.924 | 1.000 | ENSG00000184465:ENSG00000135678:ENSG00000160460 |
| DEG.two side | Brain_Cerebellar_Hemisphere | 6629 | 3 | 0.833 | 1.000 | ENSG00000131018:ENSG00000135678:ENSG00000160460 |
| DEG.two side | Brain_Cerebellum | 6538 | 3 | 0.824 | 1.000 | ENSG00000131018:ENSG00000135678:ENSG00000160460 |
| DEG.two side | Brain_Cortex | 7116 | 3 | 0.875 | 1.000 | ENSG00000184465:ENSG00000135678:ENSG00000160460 |
| DEG.two side | Brain_Frontal_Cortex_BA9 | 6851 | 3 | 0.853 | 1.000 | ENSG00000184465:ENSG00000135678:ENSG00000160460 |
| DEG.two side | Brain_Hippocampus | 8348 | 3 | 0.946 | 1.000 | ENSG00000184465:ENSG00000135678:ENSG00000160460 |
| DEG.two side | Brain_Hypothalamus | 7282 | 3 | 0.887 | 1.000 | ENSG00000184465:ENSG00000135678:ENSG00000160460 |
| DEG.two side | Brain_Nucleus_accumbens_basal_ganglia | 7635 | 3 | 0.910 | 1.000 | ENSG00000184465:ENSG00000135678:ENSG00000160460 |
| DEG.two side | Brain_Putamen_basal_ganglia | 8427 | 3 | 0.949 | 1.000 | ENSG00000184465:ENSG00000135678:ENSG00000160460 |
| DEG.two side | Brain_Spinal_cord_cervical_c-1 | 6158 | 2 | 0.933 | 1.000 | ENSG00000131018:ENSG00000184465 |
| DEG.two side | Brain_Substantia_nigra | 7766 | 4 | 0.773 | 1.000 | ENSG00000131018:ENSG00000184465:ENSG00000135678:ENSG00000160460 |

| | | | | | | |
|---|---|---|---|---|---|---|
| DEG.two side | Breast_Mammary_Tissue | 2340 | 2 | 0.429 | 1.000 | ENSG00000135678:ENSG00000160460 |
| DEG.two side | Cells_Cultured_fibroblasts | 5399 | 4 | 0.430 | 1.000 | ENSG00000131018:ENSG00000130024:ENSG00000135678:ENSG00000167578 |
| DEG.two side | Cells_EBV-transformed_lymphocytes | 5896 | 3 | 0.753 | 1.000 | ENSG00000071909:ENSG00000131018:ENSG00000135678 |
| DEG.two side | Cervix_Ectocervix | 189 | 0 | 1.000 | 1.000 | |
| DEG.two side | Cervix_Endocervix | 1728 | 0 | 1.000 | 1.000 | |
| DEG.two side | Colon_Sigmoid | 2794 | 2 | 0.528 | 1.000 | ENSG00000135678:ENSG00000160460 |
| DEG.two side | Colon_Transverse | 2532 | 2 | 0.472 | 1.000 | ENSG00000131018:ENSG00000160460 |
| DEG.two side | Esophagus_Gastroesophageal_Junction | 2422 | 1 | 0.796 | 1.000 | ENSG00000160460 |
| DEG.two side | Esophagus_Mucosa | 4877 | 2 | 0.843 | 1.000 | ENSG00000131018:ENSG00000135678 |
| DEG.two side | Esophagus_Muscularis | 2625 | 2 | 0.492 | 1.000 | ENSG00000135678:ENSG00000160460 |
| DEG.two side | Fallopian_Tube | 452 | 0 | 1.000 | 1.000 | |
| DEG.two side | Heart_Atrial_Appendage | 7937 | 6 | 0.344 | 1.000 | ENSG00000131018:ENSG00000184465:ENSG00000135678:ENSG00000160460:ENSG00000167578:ENSG00000269858 |
| DEG.two side | Heart_Left_Ventricle | 8909 | 7 | 0.263 | 1.000 | ENSG00000131018:ENSG00000184465:ENSG00000130024:ENSG00000135678:ENSG00000160460:ENSG00000167578:ENSG00000269858 |
| DEG.two side | Kidney_Cortex | 5997 | 2 | 0.925 | 1.000 | ENSG00000071909:ENSG00000130024 |
| DEG.two side | Kidney_Medulla | 93 | 0 | 1.000 | 1.000 | |

| | | | | | | |
|---|---|---|---|---|---|---|
| DEG.two side | Liver | 8286 | 5 | 0.630 | 1.000 | ENSG00000131018:ENSG00000184465:ENSG00000130024:ENSG00000167578:ENSG00000269858 |
| DEG.two side | Lung | 3849 | 2 | 0.717 | 1.000 | ENSG00000131018:ENSG00000135678 |
| DEG.two side | Minor_Salivary_Gland | 2951 | 2 | 0.560 | 1.000 | ENSG00000131018:ENSG00000135678 |
| DEG.two side | Muscle_Skeletal | 6972 | 6 | 0.219 | 1.000 | ENSG00000131018:ENSG00000184465:ENSG00000135678:ENSG00000160460:ENSG00000167578:ENSG00000269858 |
| DEG.two side | Nerve_Tibial | 4941 | 2 | 0.849 | 1.000 | ENSG00000184465:ENSG00000160460 |
| DEG.two side | Ovary | 5103 | 2 | 0.863 | 1.000 | ENSG00000131018:ENSG00000184465 |
| DEG.two side | Pancreas | 9220 | 6 | 0.533 | 1.000 | ENSG00000131018:ENSG00000184465:ENSG00000130024:ENSG00000135678:ENSG00000167578:ENSG00000269858 |
| DEG.two side | Pituitary | 5005 | 5 | 0.165 | 1.000 | ENSG00000071909:ENSG00000131018:ENSG00000135678:ENSG00000160460:ENSG00000167578 |
| DEG.two side | Prostate | 2717 | 2 | 0.512 | 1.000 | ENSG00000184465:ENSG00000160460 |
| DEG.two side | Skin_Not_Sun_Exposed_Suprapubic | 4367 | 2 | 0.787 | 1.000 | ENSG00000135678:ENSG00000269858 |
| DEG.two side | Skin_Sun_Exposed_Lower_leg | 4286 | 2 | 0.777 | 1.000 | ENSG00000135678:ENSG00000160460 |
| DEG.two side | Small_Intestine_Terminal_Ileum | 3112 | 1 | 0.876 | 1.000 | ENSG00000131018 |
| DEG.two side | Spleen | 4596 | 1 | 0.961 | 1.000 | ENSG00000167578 |
| DEG.two side | Stomach | 3070 | 1 | 0.872 | 1.000 | ENSG00000160460 |
| DEG.two side | Testis | 8546 | 3 | 0.953 | 1.000 | ENSG00000071909:ENSG00000184465:ENSG00000135678 |

| | | | | | | |
|---|---|---|---|---|---|---|
| DEG.two side | Thyroid | 4708 | 4 | 0.319 | 1.000 | ENSG00000071909:ENSG00000131018:ENSG00000184465:ENSG00000160460 |
| DEG.two side | Uterus | 4787 | 2 | 0.834 | 1.000 | ENSG00000131018:ENSG00000184465 |
| DEG.two side | Vagina | 2611 | 2 | 0.489 | 1.000 | ENSG00000184465:ENSG00000160460 |
| DEG.two side | Whole_Blood | 7620 | 3 | 0.909 | 1.000 | ENSG00000131018:ENSG00000130024:ENSG00000135678 |

***Table S23. Tissue Specificity of Prioritised Genes from FUMA (using GTEx v8 54 Tissue Types): Reaction Time (Identification Task)***

| Category | GeneSet | N_genes | N_overlap | p | adjP | genes |
|---|---|---|---|---|---|---|
| DEG.up | Adipose_Subcutaneous | 1689 | 0 | 1.000 | 1.000 | |
| DEG.up | Adipose_Visceral_Omentum | 1428 | 0 | 1.000 | 1.000 | |
| DEG.up | Adrenal_Gland | 1204 | 3 | 0.344 | 1.000 | ENSG00000148795:ENSG00000166275:ENSG00000148842 |
| DEG.up | Artery_Aorta | 2103 | 2 | 0.892 | 1.000 | ENSG00000148842:ENSG00000076685 |
| DEG.up | Artery_Coronary | 1593 | 1 | 0.944 | 1.000 | ENSG00000148842 |
| DEG.up | Artery_Tibial | 2175 | 5 | 0.318 | 1.000 | ENSG00000117614:ENSG00000146085:ENSG00000148842:ENSG00000076685:ENSG00000156374 |
| DEG.up | Bladder | 728 | 1 | 0.722 | 1.000 | ENSG00000167653 |
| DEG.up | Brain_Amygdala | 1438 | 4 | 0.233 | 1.000 | ENSG00000163517:ENSG00000188191:ENSG00000148798:ENSG00000087495 |
| DEG.up | Brain_Anterior_cingulate_cortex_BA24 | 1866 | 5 | 0.214 | 1.000 | ENSG00000163517:ENSG00000188191:ENSG00000148798:ENSG00000127588:ENSG00000087495 |
| DEG.up | Brain_Caudate_basal_ganglia | 1703 | 4 | 0.337 | 1.000 | ENSG00000163517:ENSG00000188191:ENSG00000148798:ENSG00000087495 |
| DEG.up | Brain_Cerebellar_Hemisphere | 4248 | 8 | 0.454 | 1.000 | ENSG00000188191:ENSG00000164815:ENSG00000148842:ENSG00000148798:ENSG00000148835:ENSG00000127586:ENSG00000127588:ENSG00000087495 |
| DEG.up | Brain_Cerebellum | 4055 | 8 | 0.398 | 1.000 | ENSG00000188191:ENSG00000148842:ENSG00000148798:ENSG00000148835:ENSG00000007376:ENSG00000127586:ENSG00000127588:ENSG00000087495 |
| DEG.up | Brain_Cortex | 2159 | 6 | 0.160 | 1.000 | ENSG00000163517:ENSG00000188191:ENSG00000148798:ENSG00000162006:ENSG00000127588:ENSG00000087495 |
| DEG.up | Brain_Frontal_Cortex_BA9 | 2422 | 5 | 0.407 | 1.000 | ENSG00000163517:ENSG00000188191:ENSG00000148798:ENSG00000127588:ENSG00000087495 |
| DEG.up | Brain_Hippocampus | 1542 | 4 | 0.273 | 1.000 | ENSG00000163517:ENSG00000188191:ENSG00000148798:ENSG00000087495 |

| | | | | | | |
|---|---|---|---|---|---|---|
| DEG.up | Brain_Hypothalamus | 1979 | 4 | 0.449 | 1.000 | ENSG00000163517:ENSG00000188191:ENSG00000148798:ENSG00000087495 |
| DEG.up | Brain_Nucleus_accumbens_basal_ganglia | 1834 | 4 | 0.390 | 1.000 | ENSG00000163517:ENSG00000188191:ENSG00000148798:ENSG00000087495 |
| DEG.up | Brain_Putamen_basal_ganglia | 1346 | 4 | 0.199 | 1.000 | ENSG00000163517:ENSG00000188191:ENSG00000148798:ENSG00000087495 |
| DEG.up | Brain_Spinal_cord_cervical_c-1 | 1611 | 8 | 0.005 | 0.266 | ENSG00000188672:ENSG00000163517:ENSG00000031691:ENSG00000188191:ENSG00000148798:ENSG00000127588:ENSG00000105357:ENSG00000087495 |
| DEG.up | Brain_Substantia_nigra | 1277 | 4 | 0.174 | 1.000 | ENSG00000163517:ENSG00000188191:ENSG00000148798:ENSG00000087495 |
| DEG.up | Breast_Mammary_Tissue | 1428 | 1 | 0.923 | 1.000 | ENSG00000130193 |
| DEG.up | Cells_Cultured_fibroblasts | 3686 | 8 | 0.293 | 1.000 | ENSG00000124813:ENSG00000031691:ENSG00000164815:ENSG00000160886:ENSG00000076685:ENSG00000156374:ENSG00000148835:ENSG00000007376 |
| DEG.up | Cells_EBV-transformed_lymphocytes | 3833 | 6 | 0.673 | 1.000 | ENSG00000031691:ENSG00000164815:ENSG00000156374:ENSG00000148835:ENSG00000007376:ENSG00000127586 |
| DEG.up | Cervix_Ectocervix | 187 | 0 | 1.000 | 1.000 | |
| DEG.up | Cervix_Endocervix | 1723 | 3 | 0.581 | 1.000 | ENSG00000164815:ENSG00000166275:ENSG00000156374 |
| DEG.up | Colon_Sigmoid | 1264 | 1 | 0.896 | 1.000 | ENSG00000156374 |
| DEG.up | Colon_Transverse | 1128 | 0 | 1.000 | 1.000 | |
| DEG.up | Esophagus_Gastroesophageal_Junction | 1016 | 0 | 1.000 | 1.000 | |
| DEG.up | Esophagus_Mucosa | 1755 | 5 | 0.180 | 1.000 | ENSG00000167653:ENSG00000160886:ENSG00000130193:ENSG00000076685:ENSG00000105357 |
| DEG.up | Esophagus_Muscularis | 1043 | 0 | 1.000 | 1.000 | |
| DEG.up | Fallopian_Tube | 451 | 0 | 1.000 | 1.000 | |
| DEG.up | Heart_Atrial_Appendage | 572 | 0 | 1.000 | 1.000 | |
| DEG.up | Heart_Left_Ventricle | 395 | 0 | 1.000 | 1.000 | |
| DEG.up | Kidney_Cortex | 948 | 1 | 0.813 | 1.000 | ENSG00000148795 |

| | | | | | | |
|---|---|---|---|---|---|---|
| DEG.up | Kidney_Medulla | 93 | 0 | 1.000 | 1.000 | |
| DEG.up | Liver | 1090 | 2 | 0.568 | 1.000 | ENSG00000146085:ENSG00000130193 |
| DEG.up | Lung | 3029 | 1 | 0.997 | 1.000 | ENSG00000105357 |
| DEG.up | Minor_Salivary_Gland | 1285 | 6 | 0.021 | 1.000 | ENSG00000187010:ENSG00000188672:ENSG00000124813:ENSG00000167653:ENSG00000076685:ENSG00000105357 |
| DEG.up | Muscle_Skeletal | 879 | 1 | 0.789 | 1.000 | ENSG00000105357 |
| DEG.up | Nerve_Tibial | 4157 | 4 | 0.949 | 1.000 | ENSG00000163517:ENSG00000124813:ENSG00000031691:ENSG00000164815 |
| DEG.up | Ovary | 3674 | 5 | 0.788 | 1.000 | ENSG00000146085:ENSG00000031691:ENSG00000188191:ENSG00000148795:ENSG00000127586 |
| DEG.up | Pancreas | 549 | 0 | 1.000 | 1.000 | |
| DEG.up | Pituitary | 3253 | 5 | 0.683 | 1.000 | ENSG00000188672:ENSG00000188191:ENSG00000148842:ENSG00000148798:ENSG00000087495 |
| DEG.up | Prostate | 2019 | 2 | 0.877 | 1.000 | ENSG00000124813:ENSG00000167653 |
| DEG.up | Skin_Not_Sun_Exposed_Suprapubic | 2037 | 3 | 0.698 | 1.000 | ENSG00000167653:ENSG00000160886:ENSG00000105357 |
| DEG.up | Skin_Sun_Exposed_Lower_leg | 2163 | 3 | 0.738 | 1.000 | ENSG00000167653:ENSG00000130193:ENSG00000105357 |
| DEG.up | Small_Intestine_Terminal_Ileum | 1855 | 2 | 0.844 | 1.000 | ENSG00000124813:ENSG00000105357 |
| DEG.up | Spleen | 2666 | 3 | 0.858 | 1.000 | ENSG00000187010:ENSG00000007376:ENSG00000127586 |
| DEG.up | Stomach | 780 | 1 | 0.747 | 1.000 | ENSG00000167653 |
| DEG.up | Testis | 5947 | 9 | 0.743 | 1.000 | ENSG00000188672:ENSG00000163517:ENSG00000164815:ENSG00000160886:ENSG00000148795:ENSG00000076685:ENSG00000156374:ENSG00000148835:ENSG00000127586 |
| DEG.up | Thyroid | 3732 | 5 | 0.800 | 1.000 | ENSG00000130193:ENSG00000148795:ENSG00000166275:ENSG00000076685:ENSG00000105357 |
| DEG.up | Uterus | 3999 | 3 | 0.980 | 1.000 | ENSG00000188191:ENSG00000156374:ENSG00000127586 |

| | | | | | | |
|---|---|---|---|---|---|---|
| DEG.up | Vagina | 2013 | 4 | 0.462 | 1.000 | ENSG00000124813:ENSG00000167653:ENSG00000160886:ENSG00000105357 |
| DEG.up | Whole_Blood | 1299 | 2 | 0.666 | 1.000 | ENSG00000187010:ENSG00000188672 |
| DEG.down | Adipose_Subcutaneous | 1307 | 4 | 0.185 | 1.000 | ENSG00000163517:ENSG00000148795:ENSG00000105357:ENSG00000087495 |
| DEG.down | Adipose_Visceral_Omentum | 1400 | 4 | 0.218 | 1.000 | ENSG00000163517:ENSG00000124813:ENSG00000167653:ENSG00000105357 |
| DEG.down | Adrenal_Gland | 2994 | 5 | 0.605 | 1.000 | ENSG00000163517:ENSG00000188191:ENSG00000148835:ENSG00000127586:ENSG00000105357 |
| DEG.down | Artery_Aorta | 1323 | 3 | 0.401 | 1.000 | ENSG00000124813:ENSG00000160886:ENSG00000105357 |
| DEG.down | Artery_Coronary | 863 | 2 | 0.442 | 1.000 | ENSG00000160886:ENSG00000105357 |
| DEG.down | Artery_Tibial | 1746 | 4 | 0.354 | 1.000 | ENSG00000188191:ENSG00000160886:ENSG00000130193:ENSG00000105357 |
| DEG.down | Bladder | 103 | 0 | 1.000 | 1.000 | |
| DEG.down | Brain_Amygdala | 6995 | 12 | 0.575 | 1.000 | ENSG00000117614:ENSG00000117616:ENSG00000146085:ENSG00000164815:ENSG00000130193:ENSG00000166272:ENSG00000148842:ENSG00000076685:ENSG00000156374:ENSG00000148835:ENSG00000007376:ENSG00000127586 |
| DEG.down | Brain_Anterior_cingulate_cortex_BA24 | 6003 | 9 | 0.755 | 1.000 | ENSG00000117614:ENSG00000117616:ENSG00000164815:ENSG00000160886:ENSG00000166272:ENSG00000076685:ENSG00000156374:ENSG00000148835:ENSG00000127586 |
| DEG.down | Brain_Caudate_basal_ganglia | 6174 | 7 | 0.946 | 1.000 | ENSG00000117614:ENSG00000164815:ENSG00000166272:ENSG00000076685:ENSG00000156374:ENSG00000148835:ENSG00000127586 |
| DEG.down | Brain_Cerebellar_Hemisphere | 2381 | 1 | 0.988 | 1.000 | ENSG00000166272 |
| DEG.down | Brain_Cerebellum | 2483 | 2 | 0.940 | 1.000 | ENSG00000117614:ENSG00000166272 |

| | | | | | | |
|---|---|---|---|---|---|---|
| DEG.down | Brain_Cortex | 4957 | 5 | 0.955 | 1.000 | ENSG00000117614:ENSG00000160886:ENSG00000166272:ENSG00000076685:ENSG00000156374 |
| DEG.down | Brain_Frontal_Cortex_BA9 | 4429 | 6 | 0.811 | 1.000 | ENSG00000117614:ENSG00000160886:ENSG00000166272:ENSG00000076685:ENSG00000156374:ENSG00000127586 |
| DEG.down | Brain_Hippocampus | 6806 | 8 | 0.945 | 1.000 | ENSG00000117614:ENSG00000146085:ENSG00000164815:ENSG00000166272:ENSG00000076685:ENSG00000156374:ENSG00000148835:ENSG00000127586 |
| DEG.down | Brain_Hypothalamus | 5303 | 8 | 0.735 | 1.000 | ENSG00000117614:ENSG00000124813:ENSG00000160886:ENSG00000166272:ENSG00000076685:ENSG00000156374:ENSG00000148835:ENSG00000127586 |
| DEG.down | Brain_Nucleus_accumbens_basal_ganglia | 5801 | 6 | 0.963 | 1.000 | ENSG00000117614:ENSG00000166272:ENSG00000076685:ENSG00000156374:ENSG00000148835:ENSG00000127586 |
| DEG.down | Brain_Putamen_basal_ganglia | 7081 | 9 | 0.916 | 1.000 | ENSG00000117614:ENSG00000117616:ENSG00000146085:ENSG00000164815:ENSG00000166272:ENSG00000076685:ENSG00000156374:ENSG00000148835:ENSG00000127586 |
| DEG.down | Brain_Spinal_cord_cervical_c-1 | 4547 | 6 | 0.832 | 1.000 | ENSG00000164815:ENSG00000160886:ENSG00000166272:ENSG00000148835:ENSG00000007376:ENSG00000127586 |
| DEG.down | Brain_Substantia_nigra | 6489 | 12 | 0.445 | 1.000 | ENSG00000117614:ENSG00000124813:ENSG00000146085:ENSG00000164815:ENSG00000160886:ENSG00000130193:ENSG00000166272:ENSG00000076685:ENSG00000156374:ENSG00000148835:ENSG00000007376:ENSG00000127586 |
| DEG.down | Breast_Mammary_Tissue | 912 | 2 | 0.471 | 1.000 | ENSG00000163517:ENSG00000148795 |
| DEG.down | Cells_Cultured_fibroblasts | 1713 | 3 | 0.577 | 1.000 | ENSG00000163517:ENSG00000188191:ENSG00000130193 |
| DEG.down | Cells_EBV-transformed_lymphocytes | 2063 | 4 | 0.482 | 1.000 | ENSG00000163517:ENSG00000166272:ENSG00000166275:ENSG00000148842 |

| | | | | | | |
|---|---|---|---|---|---|---|
| DEG.down | Cervix_Ectocervix | 2 | 0 | 1.000 | 1.000 | |
| DEG.down | Cervix_Endocervix | 5 | 0 | 1.000 | 1.000 | |
| DEG.down | Colon_Sigmoid | 1530 | 3 | 0.498 | 1.000 | ENSG00000167653:ENSG00000105357:ENSG00000087495 |
| DEG.down | Colon_Transverse | 1404 | 2 | 0.708 | 1.000 | ENSG00000148798:ENSG00000087495 |
| DEG.down | Esophagus_Gastroesophageal_Junction | 1406 | 5 | 0.090 | 1.000 | ENSG00000167653:ENSG00000130193:ENSG00000148795:ENSG00000148798:ENSG00000105357 |
| DEG.down | Esophagus_Mucosa | 3122 | 6 | 0.456 | 1.000 | ENSG00000124813:ENSG00000188191:ENSG00000148795:ENSG00000166275:ENSG00000148842:ENSG00000148798 |
| DEG.down | Esophagus_Muscularis | 1582 | 4 | 0.288 | 1.000 | ENSG00000167653:ENSG00000148795:ENSG00000127586:ENSG00000105357 |
| DEG.down | Fallopian_Tube | 1 | 0 | 1.000 | 1.000 | |
| DEG.down | Heart_Atrial_Appendage | 7365 | 12 | 0.664 | 1.000 | ENSG00000117614:ENSG00000117616:ENSG00000188191:ENSG00000164815:ENSG00000130193:ENSG00000166272:ENSG00000076685:ENSG00000156374:ENSG00000148835:ENSG00000007376:ENSG00000127586:ENSG00000087495 |
| DEG.down | Heart_Left_Ventricle | 8514 | 17 | 0.254 | 1.000 | ENSG00000117614:ENSG00000117616:ENSG00000183726:ENSG00000163517:ENSG00000146085:ENSG00000031691:ENSG00000188191:ENSG00000164815:ENSG00000130193:ENSG00000166272:ENSG00000166275:ENSG00000076685:ENSG00000156374:ENSG00000148835:ENSG00000007376:ENSG00000127586:ENSG00000087495 |
| DEG.down | Kidney_Cortex | 5049 | 9 | 0.520 | 1.000 | ENSG00000124813:ENSG00000031691:ENSG00000164815:ENSG00000166272:ENSG00000076685:ENSG00000156374:ENSG00000148835:ENSG00000127586:ENSG00000105357 |

| | | | | | | |
|---|---|---|---|---|---|---|
| DEG.down | Kidney_Medulla | 0 | 0 | 1.000 | 1.000 | |
| DEG.down | Liver | 7196 | 11 | 0.754 | 1.000 | ENSG00000117614:ENSG00000183726:ENSG00000163517:ENSG00000031691:ENSG00000188191:ENSG00000164815:ENSG00000148842:ENSG00000076685:ENSG00000148835:ENSG00000007376:ENSG00000127586 |
| DEG.down | Lung | 820 | 2 | 0.416 | 1.000 | ENSG00000163517:ENSG00000148795 |
| DEG.down | Minor_Salivary_Gland | 1666 | 2 | 0.795 | 1.000 | ENSG00000188191:ENSG00000148795 |
| DEG.down | Muscle_Skeletal | 6093 | 10 | 0.640 | 1.000 | ENSG00000117616:ENSG00000183726:ENSG00000031691:ENSG00000188191:ENSG00000164815:ENSG00000160886:ENSG00000130193:ENSG00000148835:ENSG00000007376:ENSG00000127586 |
| DEG.down | Nerve_Tibial | 784 | 1 | 0.749 | 1.000 | ENSG00000188191 |
| DEG.down | Ovary | 1429 | 1 | 0.923 | 1.000 | ENSG00000130193 |
| DEG.down | Pancreas | 8671 | 17 | 0.287 | 1.000 | ENSG00000117614:ENSG00000117616:ENSG00000183726:ENSG00000146085:ENSG00000031691:ENSG00000188191:ENSG00000164815:ENSG00000167653:ENSG00000166272:ENSG00000166275:ENSG00000148842:ENSG00000076685:ENSG00000156374:ENSG00000148835:ENSG00000007376:ENSG00000127586:ENSG00000105357 |
| DEG.down | Pituitary | 1752 | 0 | 1.000 | 1.000 | |
| DEG.down | Prostate | 698 | 1 | 0.707 | 1.000 | ENSG00000160886 |
| DEG.down | Skin_Not_Sun_Exposed_Suprapubic | 2330 | 3 | 0.784 | 1.000 | ENSG00000188191:ENSG00000148795:ENSG00000087495 |
| DEG.down | Skin_Sun_Exposed_Lower_leg | 2123 | 5 | 0.300 | 1.000 | ENSG00000188191:ENSG00000148795:ENSG00000166275:ENSG00000148798:ENSG00000087495 |

| | | | | | | |
|---|---|---|---|---|---|---|
| DEG.down | Small_Intestine_Terminal_Ileum | 1257 | 3 | 0.369 | 1.000 | ENSG00000163517:ENSG00000148795:ENSG00000087495 |
| DEG.down | Spleen | 1930 | 3 | 0.661 | 1.000 | ENSG00000163517:ENSG00000146085:ENSG00000148842 |
| DEG.down | Stomach | 2290 | 2 | 0.919 | 1.000 | ENSG00000163517:ENSG00000188191 |
| DEG.down | Testis | 2599 | 5 | 0.471 | 1.000 | ENSG00000183726:ENSG00000167653:ENSG00000130193:ENSG00000105357:ENSG00000087495 |
| DEG.down | Thyroid | 976 | 1 | 0.823 | 1.000 | ENSG00000160886 |
| DEG.down | Uterus | 788 | 1 | 0.751 | 1.000 | ENSG00000105357 |
| DEG.down | Vagina | 598 | 2 | 0.275 | 1.000 | ENSG00000148798:ENSG00000087495 |
| DEG.down | Whole_Blood | 6321 | 11 | 0.551 | 1.000 | ENSG00000117614:ENSG00000163517:ENSG00000146085:ENSG00000188191:ENSG00000164815:ENSG00000130193:ENSG00000166272:ENSG00000166275:ENSG00000156374:ENSG00000148835:ENSG00000127586 |
| DEG.twoside | Adipose_Subcutaneous | 2996 | 4 | 0.783 | 1.000 | ENSG00000163517:ENSG00000148795:ENSG00000105357:ENSG00000087495 |
| DEG.twoside | Adipose_Visceral_Omentum | 2828 | 4 | 0.740 | 1.000 | ENSG00000163517:ENSG00000124813:ENSG00000167653:ENSG00000105357 |
| DEG.twoside | Adrenal_Gland | 4198 | 8 | 0.440 | 1.000 | ENSG00000163517:ENSG00000188191:ENSG00000148795:ENSG00000166275:ENSG00000148842:ENSG00000148835:ENSG00000127586:ENSG00000105357 |
| DEG.twoside | Artery_Aorta | 3426 | 5 | 0.730 | 1.000 | ENSG00000124813:ENSG00000160886:ENSG00000148842:ENSG00000076685:ENSG00000105357 |
| DEG.twoside | Artery_Coronary | 2456 | 3 | 0.815 | 1.000 | ENSG00000160886:ENSG00000148842:ENSG00000105357 |
| DEG.twoside | Artery_Tibial | 3921 | 9 | 0.218 | 1.000 | ENSG00000117614:ENSG00000146085:ENSG00000188191:ENSG00000160886:ENSG00000130193:ENSG00000148842:ENSG00000076685:ENSG00000156374:ENSG00000105357 |

| | | | | | | |
|---|---|---|---|---|---|---|
| DEG.two side | Bladder | 831 | 1 | 0.769 | 1.000 | ENSG00000167653 |
| DEG.two side | Brain_Amygdala | 8433 | 16 | 0.362 | 1.000 | ENSG00000117614:ENSG00000117616:ENSG00000163517:ENSG00000146085:ENSG00000188191:ENSG00000164815:ENSG00000130193:ENSG00000166272:ENSG00000148842:ENSG00000076685:ENSG00000148798:ENSG00000156374:ENSG00000148835:ENSG00000007376:ENSG00000127586:ENSG00000087495 |
| DEG.two side | Brain_Anterior_cingulate_cortex_BA24 | 7869 | 14 | 0.505 | 1.000 | ENSG00000117614:ENSG00000117616:ENSG00000163517:ENSG00000188191:ENSG00000164815:ENSG00000160886:ENSG00000166272:ENSG00000076685:ENSG00000148798:ENSG00000156374:ENSG00000148835:ENSG00000127586:ENSG00000127588:ENSG00000087495 |
| DEG.two side | Brain_Caudate_basal_ganglia | 7877 | 11 | 0.867 | 1.000 | ENSG00000117614:ENSG00000163517:ENSG00000188191:ENSG00000164815:ENSG00000166272:ENSG00000076685:ENSG00000148798:ENSG00000156374:ENSG00000148835:ENSG00000127586:ENSG00000087495 |
| DEG.two side | Brain_Cerebellar_Hemisphere | 6629 | 9 | 0.862 | 1.000 | ENSG00000188191:ENSG00000164815:ENSG00000166272:ENSG00000148842:ENSG00000148798:ENSG00000148835:ENSG00000127586:ENSG00000127588:ENSG00000087495 |
| DEG.two side | Brain_Cerebellum | 6538 | 10 | 0.741 | 1.000 | ENSG00000117614:ENSG00000188191:ENSG00000166272:ENSG00000148842:ENSG00000148798:ENSG00000148835:ENSG00000007376:ENSG00000127586:ENSG00000127588:ENSG00000087495 |
| DEG.two side | Brain_Cortex | 7116 | 11 | 0.738 | 1.000 | ENSG00000117614:ENSG00000163517:ENSG00000188191:ENSG00000160886:ENSG00000166272:ENSG00000076685:ENSG00000148798:ENSG00000156374:ENSG00000162006:ENSG00000127588:ENSG00000087495 |
| DEG.two side | Brain_Frontal_Cortex_BA9 | 6851 | 11 | 0.680 | 1.000 | ENSG00000117614:ENSG00000163517:ENSG00000188191:ENSG00000160886:ENSG00000166272:ENSG00000076685:ENSG00000148798:ENSG00000156374:ENSG00000127586:ENSG00000127588:ENSG00000087495 |

| | | | | | | |
|---|---|---|---|---|---|---|
| DEG.two side | Brain_Hippocampus | 8348 | 12 | 0.850 | 1.000 | ENSG00000117614:ENSG00000163517:ENSG00000146085:ENSG00000188191:ENSG00000164815:ENSG00000166272:ENSG00000076685:ENSG00000148798:ENSG00000156374:ENSG00000148835:ENSG00000127586:ENSG00000087495 |
| DEG.two side | Brain_Hypothalamus | 7282 | 12 | 0.645 | 1.000 | ENSG00000117614:ENSG00000163517:ENSG00000124813:ENSG00000188191:ENSG00000160886:ENSG00000166272:ENSG00000076685:ENSG00000148798:ENSG00000156374:ENSG00000148835:ENSG00000127586:ENSG00000087495 |
| DEG.two side | Brain_Nucleus_accumbens_basal_ganglia | 7635 | 10 | 0.910 | 1.000 | ENSG00000117614:ENSG00000163517:ENSG00000188191:ENSG00000166272:ENSG00000076685:ENSG00000148798:ENSG00000156374:ENSG00000148835:ENSG00000127586:ENSG00000087495 |
| DEG.two side | Brain_Putamen_basal_ganglia | 8427 | 13 | 0.765 | 1.000 | ENSG00000117614:ENSG00000117616:ENSG00000163517:ENSG00000146085:ENSG00000188191:ENSG00000164815:ENSG00000166272:ENSG00000076685:ENSG00000148798:ENSG00000156374:ENSG00000148835:ENSG00000127586:ENSG00000087495 |
| DEG.two side | Brain_Spinal_cord_cervical_c-1 | 6158 | 14 | 0.137 | 1.000 | ENSG00000188672:ENSG00000163517:ENSG00000031691:ENSG00000188191:ENSG00000164815:ENSG00000160886:ENSG00000166272:ENSG00000148798:ENSG00000148835:ENSG00000007376:ENSG00000127586:ENSG00000127588:ENSG00000105357:ENSG00000087495 |
| DEG.two side | Brain_Substantia_nigra | 7766 | 16 | 0.220 | 1.000 | ENSG00000117614:ENSG00000163517:ENSG00000124813:ENSG00000146085:ENSG00000188191:ENSG00000164815:ENSG00000160886:ENSG00000130193:ENSG00000166272:ENSG00000076685:ENSG00000148798:ENSG00000156374:ENSG00000148835:ENSG00000007376:ENSG00000127586:ENSG00000087495 |
| DEG.two side | Breast_Mammary_Tissue | 2340 | 3 | 0.787 | 1.000 | ENSG00000163517:ENSG00000130193:ENSG00000148795 |

| | | | | | | |
|---|---|---|---|---|---|---|
| DEG.two side | Cells_Cultured_fibroblasts | 5399 | 11 | 0.312 | 1.000 | ENSG00000163517:ENSG00000124813:ENSG00000031691:ENSG00000188191:ENSG00000164815:ENSG00000160886:ENSG00000130193:ENSG00000076685:ENSG00000156374:ENSG00000148835:ENSG00000007376 |
| DEG.two side | Cells_EBV-transformed_lymphocytes | 5896 | 10 | 0.591 | 1.000 | ENSG00000163517:ENSG00000031691:ENSG00000164815:ENSG00000166272:ENSG00000166275:ENSG00000148842:ENSG00000156374:ENSG00000148835:ENSG00000007376:ENSG00000127586 |
| DEG.two side | Cervix_Ectocervix | 189 | 0 | 1.000 | 1.000 | |
| DEG.two side | Cervix_Endocervix | 1728 | 3 | 0.583 | 1.000 | ENSG00000164815:ENSG00000166275:ENSG00000156374 |
| DEG.two side | Colon_Sigmoid | 2794 | 4 | 0.731 | 1.000 | ENSG00000167653:ENSG00000156374:ENSG00000105357:ENSG00000087495 |
| DEG.two side | Colon_Transverse | 2532 | 2 | 0.945 | 1.000 | ENSG00000148798:ENSG00000087495 |
| DEG.two side | Esophagus_Gastroesophageal_Junction | 2422 | 5 | 0.407 | 1.000 | ENSG00000167653:ENSG00000130193:ENSG00000148795:ENSG00000148798:ENSG00000105357 |
| DEG.two side | Esophagus_Mucosa | 4877 | 11 | 0.195 | 1.000 | ENSG00000124813:ENSG00000188191:ENSG00000167653:ENSG00000160886:ENSG00000130193:ENSG00000148795:ENSG00000166275:ENSG00000148842:ENSG00000076685:ENSG00000148798:ENSG00000105357 |
| DEG.two side | Esophagus_Muscularis | 2625 | 4 | 0.682 | 1.000 | ENSG00000167653:ENSG00000148795:ENSG00000127586:ENSG00000105357 |
| DEG.two side | Fallopian_Tube | 452 | 0 | 1.000 | 1.000 | |
| DEG.two side | Heart_Atrial_Appendage | 7937 | 12 | 0.783 | 1.000 | ENSG00000117614:ENSG00000117616:ENSG00000188191:ENSG00000164815:ENSG00000130193:ENSG00000166272:ENSG00000076685:ENSG00000156374:ENSG00000148835:ENSG00000007376:ENSG00000127586:ENSG00000087495 |
| DEG.two side | Heart_Left_Ventricle | 8909 | 17 | 0.339 | 1.000 | ENSG00000117614:ENSG00000117616:ENSG00000183726:ENSG00000163517:ENSG00000146085:ENSG0000003 |

| | | | | | | |
|---|---|---|---|---|---|---|
| | | | | | | 1691:ENSG00000188191:ENSG00000164815:ENSG00000130193:ENSG00000166272:ENSG00000166275:ENSG00000076685:ENSG00000156374:ENSG00000148835:ENSG00000007376:ENSG00000127586:ENSG00000087495 |
| DEG.two side | Kidney_Cortex | 5997 | 10 | 0.616 | 1.000 | ENSG00000124813:ENSG00000031691:ENSG00000164815:ENSG00000166272:ENSG00000148795:ENSG00000076685:ENSG00000156374:ENSG00000148835:ENSG00000127586:ENSG00000105357 |
| DEG.two side | Kidney_Medulla | 93 | 0 | 1.000 | 1.000 | |
| DEG.two side | Liver | 8286 | 13 | 0.737 | 1.000 | ENSG00000117614:ENSG00000183726:ENSG00000163517:ENSG00000146085:ENSG00000031691:ENSG00000188191:ENSG00000164815:ENSG00000130193:ENSG00000148842:ENSG00000076685:ENSG00000148835:ENSG00000007376:ENSG00000127586 |
| DEG.two side | Lung | 3849 | 3 | 0.975 | 1.000 | ENSG00000163517:ENSG00000148795:ENSG00000105357 |
| DEG.two side | Minor_Salivary_Gland | 2951 | 8 | 0.123 | 1.000 | ENSG00000187010:ENSG00000188672:ENSG00000124813:ENSG00000188191:ENSG00000167653:ENSG00000148795:ENSG00000076685:ENSG00000105357 |
| DEG.two side | Muscle_Skeletal | 6972 | 11 | 0.707 | 1.000 | ENSG00000117616:ENSG00000183726:ENSG00000031691:ENSG00000188191:ENSG00000164815:ENSG00000160886:ENSG00000130193:ENSG00000148835:ENSG00000007376:ENSG00000127586:ENSG00000105357 |
| DEG.two side | Nerve_Tibial | 4941 | 5 | 0.954 | 1.000 | ENSG00000163517:ENSG00000124813:ENSG00000031691:ENSG00000188191:ENSG00000164815 |
| DEG.two side | Ovary | 5103 | 6 | 0.910 | 1.000 | ENSG00000146085:ENSG00000031691:ENSG00000188191:ENSG00000130193:ENSG00000148795:ENSG00000127586 |
| DEG.two side | Pancreas | 9220 | 17 | 0.413 | 1.000 | ENSG00000117614:ENSG00000117616:ENSG00000183726:ENSG00000146085:ENSG00000031691:ENSG00000188191:ENSG00000164815:ENSG00000167653:ENSG00000166272:ENSG00000166275:ENSG00000148842:ENSG000 |

| | | | | | | |
|---|---|---|---|---|---|---|
| | | | | | | 00076685:ENSG00000156374:ENSG00000148835:ENSG00000007376:ENSG00000127586:ENSG00000105357 |
| DEG.two side | Pituitary | 5005 | 5 | 0.958 | 1.000 | ENSG00000188672:ENSG00000188191:ENSG00000148842:ENSG00000148798:ENSG00000087495 |
| DEG.two side | Prostate | 2717 | 3 | 0.867 | 1.000 | ENSG00000124813:ENSG00000167653:ENSG00000160886 |
| DEG.two side | Skin_Not_Sun_Exposed_Suprapubic | 4367 | 6 | 0.799 | 1.000 | ENSG00000188191:ENSG00000167653:ENSG00000160886:ENSG00000148795:ENSG00000105357:ENSG00000087495 |
| DEG.two side | Skin_Sun_Exposed_Lower_leg | 4286 | 8 | 0.465 | 1.000 | ENSG00000188191:ENSG00000167653:ENSG00000130193:ENSG00000148795:ENSG00000166275:ENSG00000148798:ENSG00000105357:ENSG00000087495 |
| DEG.two side | Small_Intestine_Terminal_Ileum | 3112 | 5 | 0.642 | 1.000 | ENSG00000163517:ENSG00000124813:ENSG00000148795:ENSG00000105357:ENSG00000087495 |
| DEG.two side | Spleen | 4596 | 6 | 0.841 | 1.000 | ENSG00000187010:ENSG00000163517:ENSG00000146085:ENSG00000148842:ENSG00000007376:ENSG00000127586 |
| DEG.two side | Stomach | 3070 | 3 | 0.918 | 1.000 | ENSG00000163517:ENSG00000188191:ENSG00000167653 |
| DEG.two side | Testis | 8546 | 14 | 0.669 | 1.000 | ENSG00000183726:ENSG00000188672:ENSG00000163517:ENSG00000164815:ENSG00000167653:ENSG00000160886:ENSG00000130193:ENSG00000148795:ENSG00000076685:ENSG00000156374:ENSG00000148835:ENSG00000127586:ENSG00000105357:ENSG00000087495 |
| DEG.two side | Thyroid | 4708 | 6 | 0.859 | 1.000 | ENSG00000160886:ENSG00000130193:ENSG00000148795:ENSG00000166275:ENSG00000076685:ENSG00000105357 |
| DEG.two side | Uterus | 4787 | 4 | 0.980 | 1.000 | ENSG00000188191:ENSG00000156374:ENSG00000127586:ENSG00000105357 |
| DEG.two side | Vagina | 2611 | 6 | 0.289 | 1.000 | ENSG00000124813:ENSG00000167653:ENSG00000160886:ENSG00000148798:ENSG00000105357:ENSG00000087495 |

| | | | | | | |
|---|---|---|---|---|---|---|
| DEG.two side | Whole_Blood | 7620 | 13 | 0.586 | 1.000 | ENSG00000117614:ENSG00000187010:ENSG00000188672:ENSG00000163517:ENSG00000146085:ENSG00000188191:ENSG00000164815:ENSG00000130193:ENSG00000166272:ENSG00000166275:ENSG00000156374:ENSG00000148835:ENSG00000127586 |

*Table S24. MR CAUSE: Delta expected log pointwise posterior density (ELPD) results for all phenotypes.*

| Model 1 | Model 2 | Delta ELPD | SE Delta ELPD | Z-score | Exposure_Outcome | p-value |
|---|---|---|---|---|---|---|
| | | | Exposure: GCA | | | |
| null | sharing | -11.554 | 3.401 | -3.397 | GCA_DEP | 3.40E-04 |
| null | causal | -17.346 | 5.028 | -3.450 | GCA_DEP | 2.80E-04 |
| sharing | causal | -5.792 | 1.648 | -3.514 | GCA_DEP | 2.21E-04 |
| null | sharing | -0.712 | 1.336 | -0.533 | GCA_ANX | 2.97E-01 |
| null | causal | -1.401 | 2.414 | -0.580 | GCA_ANX | 2.81E-01 |
| sharing | causal | -0.689 | 1.184 | -0.582 | GCA_ANX | 2.80E-01 |
| null | sharing | -2.571 | 1.454 | -1.767 | GCA_WELLBEING | 3.86E-02 |
| null | causal | -6.541 | 3.421 | -1.912 | GCA_WELLBEING | 2.79E-02 |
| sharing | causal | -3.971 | 1.978 | -2.008 | GCA_WELLBEING | 2.23E-02 |
| null | sharing | -2.853 | 2.124 | -1.343 | GCA_PANASNEG | 8.96E-02 |
| null | causal | -4.704 | 3.399 | -1.384 | GCA_PANASNEG | 8.32E-02 |
| sharing | causal | -1.851 | 1.373 | -1.348 | GCA_PANASNEG | 8.89E-02 |
| null | sharing | 0.178 | 0.429 | 0.416 | GCA_PANASPOS | 6.61E-01 |
| null | causal | 0.180 | 1.581 | 0.114 | GCA_PANASPOS | 5.45E-01 |
| sharing | causal | 0.002 | 1.162 | 0.002 | GCA_PANASPOS | 5.01E-01 |
| | | | Exposure: Depression | | | |
| null | sharing | -12.227 | 3.593 | -3.403 | DEP_GCA | 3.33E-04 |
| null | causal | -17.856 | 5.196 | -3.437 | DEP_GCA | 2.95E-04 |
| sharing | causal | -5.630 | 1.632 | -3.449 | DEP_GCA | 2.81E-04 |
| null | sharing | -0.224 | 0.865 | -0.259 | DEP_RFFT | 3.98E-01 |
| null | causal | -0.842 | 2.064 | -0.408 | DEP_RFFT | 3.42E-01 |
| sharing | causal | -0.618 | 1.234 | -0.501 | DEP_RFFT | 3.08E-01 |
| null | sharing | 0.515 | 0.110 | 4.679 | DEP_ONEBACK | 1.00E+00 |
| null | causal | 1.602 | 0.216 | 7.426 | DEP_ONEBACK | 1.00E+00 |
| sharing | causal | 1.087 | 0.115 | 9.460 | DEP_ONEBACK | 1.00E+00 |
| null | sharing | 0.459 | 0.383 | 1.198 | DEP_RT | 8.85E-01 |
| null | causal | 1.195 | 0.927 | 1.288 | DEP_RT | 9.01E-01 |
| sharing | causal | 0.735 | 0.594 | 1.238 | DEP_RT | 8.92E-01 |
| null | sharing | 0.314 | 0.370 | 0.849 | DEP_OCL | 8.02E-01 |

| | | | | | | |
|---|---|---|---|---|---|---|
| null | causal | 0.116 | 1.484 | 0.078 | DEP_OCL | 5.31E-01 |
| sharing | causal | -0.198 | 1.127 | -0.176 | DEP_OCL | 4.30E-01 |
| Exposure: Wellbeing | | | | | | |
| null | sharing | -3.574 | 1.774 | -2.015 | WELLBEING_GCA | 2.19E-02 |
| null | causal | -7.793 | 3.687 | -2.113 | WELLBEING_GCA | 1.73E-02 |
| sharing | causal | -4.219 | 1.927 | -2.190 | WELLBEING_GCA | 1.43E-02 |
| null | sharing | 0.175 | 0.802 | 0.218 | WELLBEING_RFFT | 5.86E-01 |
| null | causal | 0.424 | 1.518 | 0.279 | WELLBEING_RFFT | 6.10E-01 |
| sharing | causal | 0.249 | 0.804 | 0.310 | WELLBEING_RFFT | 6.22E-01 |
| null | sharing | 0.576 | 0.168 | 3.426 | WELLBEING_ONEBACK | 1.00E+00 |
| null | causal | 1.320 | 0.306 | 4.311 | WELLBEING_ONEBACK | 1.00E+00 |
| sharing | causal | 0.744 | 0.158 | 4.696 | WELLBEING_ONEBACK | 1.00E+00 |
| null | sharing | 0.527 | 0.097 | 5.422 | WELLBEING_RT | 1.00E+00 |
| null | causal | 1.471 | 0.143 | 10.290 | WELLBEING_RT | 1.00E+00 |
| sharing | causal | 0.944 | 0.055 | 17.241 | WELLBEING_RT | 1.00E+00 |
| null | sharing | 0.018 | 0.767 | 0.024 | WELLBEING_OCL | 5.10E-01 |
| null | causal | -0.342 | 1.908 | -0.179 | WELLBEING_OCL | 4.29E-01 |
| sharing | causal | -0.361 | 1.175 | -0.307 | WELLBEING_OCL | 3.79E-01 |
| Exposure: Anxiety | | | | | | |
| null | sharing | 0.163 | 0.043 | 3.789 | ANX_GCA | 1.00E+00 |
| null | causal | 1.061 | 0.337 | 3.149 | ANX_GCA | 9.99E-01 |
| sharing | causal | 0.898 | 0.322 | 2.792 | ANX_GCA | 9.97E-01 |
| null | sharing | 0.222 | 0.076 | 2.904 | ANX_RFFT | 9.98E-01 |
| null | causal | 0.816 | 0.342 | 2.386 | ANX_RFFT | 9.91E-01 |
| sharing | causal | 0.594 | 0.277 | 2.139 | ANX_RFFT | 9.84E-01 |
| null | sharing | 0.095 | 0.055 | 1.719 | ANX_ONEBACK | 9.57E-01 |
| null | causal | 0.420 | 0.663 | 0.634 | ANX_ONEBACK | 7.37E-01 |
| sharing | causal | 0.326 | 0.617 | 0.528 | ANX_ONEBACK | 7.01E-01 |
| null | sharing | 0.132 | 0.065 | 2.026 | ANX_RT | 9.79E-01 |
| null | causal | 0.701 | 0.556 | 1.260 | ANX_RT | 8.96E-01 |
| sharing | causal | 0.569 | 0.506 | 1.125 | ANX_RT | 8.70E-01 |
| null | sharing | 0.231 | 0.074 | 3.097 | ANX_OCL | 9.99E-01 |
| null | causal | 0.892 | 0.289 | 3.086 | ANX_OCL | 9.99E-01 |

| | | | | | | |
|---|---|---|---|---|---|---|
| sharing | causal | 0.662 | 0.223 | 2.964 | ANX_OCL | 9.98E-01 |
| Exposure: PANAS Negative | | | | | | |
| null | sharing | 0.187 | 0.084 | 2.219 | PANASNEG_GCA | 9.87E-01 |
| null | causal | 0.579 | 0.699 | 0.829 | PANASNEG_GCA | 7.96E-01 |
| sharing | causal | 0.392 | 0.618 | 0.635 | PANASNEG_GCA | 7.37E-01 |
| null | sharing | -0.366 | 0.523 | -0.701 | PANASNEG_RFFT | 2.42E-01 |
| null | causal | -1.658 | 2.146 | -0.773 | PANASNEG_RFFT | 2.20E-01 |
| sharing | causal | -1.291 | 1.651 | -0.782 | PANASNEG_RFFT | 2.17E-01 |
| null | sharing | 0.192 | 0.115 | 1.675 | PANASNEG_ONEBACK | 9.53E-01 |
| null | causal | 0.893 | 0.497 | 1.797 | PANASNEG_ONEBACK | 9.64E-01 |
| sharing | causal | 0.701 | 0.399 | 1.758 | PANASNEG_ONEBACK | 9.61E-01 |
| null | sharing | 0.231 | 0.051 | 4.493 | PANASNEG_RT | 1.00E+00 |
| null | causal | 0.894 | 0.506 | 1.766 | PANASNEG_RT | 9.61E-01 |
| sharing | causal | 0.662 | 0.468 | 1.417 | PANASNEG_RT | 9.22E-01 |
| null | sharing | 0.082 | 0.245 | 0.333 | PANASNEG_OCL | 6.30E-01 |
| null | causal | 0.089 | 1.287 | 0.069 | PANASNEG_OCL | 5.28E-01 |
| sharing | causal | 0.007 | 1.060 | 0.007 | PANASNEG_OCL | 5.03E-01 |
| Exposure: PANAS Positive | | | | | | |
| null | sharing | 0.187 | 0.050 | 3.762 | PANASPOS_GCA | 1.00E+00 |
| null | causal | 0.684 | 0.498 | 1.374 | PANASPOS_GCA | 9.15E-01 |
| sharing | causal | 0.497 | 0.452 | 1.100 | PANASPOS_GCA | 8.64E-01 |
| null | sharing | -0.003 | 0.217 | -0.013 | PANASPOS_RFFT | 4.95E-01 |
| null | causal | -0.007 | 1.361 | -0.005 | PANASPOS_RFFT | 4.98E-01 |
| sharing | causal | -0.004 | 1.157 | -0.003 | PANASPOS_RFFT | 4.99E-01 |
| null | sharing | 0.248 | 0.053 | 4.691 | PANASPOS_ONEBACK | 1.00E+00 |
| null | causal | 1.036 | 0.131 | 7.909 | PANASPOS_ONEBACK | 1.00E+00 |
| sharing | causal | 0.788 | 0.098 | 8.073 | PANASPOS_ONEBACK | 1.00E+00 |
| null | sharing | 0.168 | 0.076 | 2.200 | PANASPOS_RT | 9.86E-01 |
| null | causal | 0.935 | 0.393 | 2.379 | PANASPOS_RT | 9.91E-01 |
| sharing | causal | 0.767 | 0.329 | 2.329 | PANASPOS_RT | 9.90E-01 |
| null | sharing | 0.157 | 0.077 | 2.029 | PANASPOS_OCL | 9.79E-01 |
| null | causal | 0.737 | 0.575 | 1.282 | PANASPOS_OCL | 9.00E-01 |
| sharing | causal | 0.580 | 0.509 | 1.139 | PANASPOS_OCL | 8.73E-01 |

Model 1/Model 2=The models being compared; Delta ELPD = Estimated difference in ELPD. If value is negative, model 2 is a better fit; SE DELTA ELPD=Standard error of Delta ELPD; Z-score (Delta ELPD/SE Delta ELPD)=A z-score that can be compared to a normal distribution to test if the difference in model fit is significant.

***Table S25. MR CAUSE: Posterior distribution estimates of parameters under causal and sharing models for all phenotypes.***

| Model | Gamma (95% CI) | Eta (95% CI) | Q (95% CI) | Exposure_Outcome |
|---|---|---|---|---|
| | | Exposure: GCA | | |
| Sharing | NA | -0.16 (-0.26, -0.1) | 0.4 (0.17, 0.64) | GCA_DEP |
| Causal | -0.1 (-0.13, -0.06) | 0 (-0.5, 0.41) | 0.03 (0, 0.25) | GCA_DEP |
| Sharing | NA | -1.43 (-5.68, 0.9) | 0.06 (0, 0.27) | GCA_ANX |
| Causal | -0.18 (-0.4, 0.05) | -0.21 (-6.27, 4.8) | 0.03 (0, 0.24) | GCA_ANX |
| Sharing | NA | 0.04 (0, 0.12) | 0.18 (0.01, 0.46) | GCA_WELLBEING |
| Causal | 0.02 (0.01, 0.03) | 0 (-0.17, 0.16) | 0.03 (0, 0.25) | GCA_WELLBEING |
| Sharing | NA | -0.25 (-0.72, -0.06) | 0.14 (0.01, 0.37) | GCA_PANASNEG |
| Causal | -0.06 (-0.11, -0.01) | 0.01 (-0.98, 1.06) | 0.03 (0, 0.24) | GCA_PANASNEG |
| Sharing | NA | -0.13 (-0.82, 0.84) | 0.04 (0, 0.25) | GCA_PANASPOS |
| Causal | -0.03 (-0.07, 0.01) | 0.03 (-0.74, 1.04) | 0.03 (0, 0.24) | GCA_PANASPOS |
| | | Exposure: Depression | | |
| Sharing | NA | -0.12 (-0.19, -0.07) | 0.41 (0.18, 0.64) | DEP_GCA |
| Causal | -0.07 (-0.09, -0.04) | 0 (-0.34, 0.33) | 0.03 (0, 0.25) | DEP_GCA |
| Sharing | NA | -0.2 (-0.95, 0.36) | 0.06 (0, 0.28) | DEP_RFFT |
| Causal | -0.04 (-0.08, 0.01) | 0.01 (-0.8, 0.76) | 0.04 (0, 0.25) | DEP_RFFT |
| Sharing | NA | 0.01 (-1.05, 1.04) | 0.02 (0, 0.2) | DEP_ONEBACK |
| Causal | 0 (-0.05, 0.05) | -0.03 (-0.99, 0.74) | 0.04 (0, 0.26) | DEP_ONEBACK |
| Sharing | NA | -0.15 (-1.4, 0.76) | 0.02 (0, 0.21) | DEP_RT |
| Causal | -0.01 (-0.06, 0.03) | -0.04 (-1.11, 0.86) | 0.03 (0, 0.23) | DEP_RT |
| Sharing | NA | -0.12 (-0.96, 0.86) | 0.04 (0, 0.24) | DEP_OCL |
| Causal | -0.03 (-0.07, 0.02) | -0.01 (-0.79, 0.44) | 0.04 (0, 0.25) | DEP_OCL |
| | | Exposure: Wellbeing | | |
| Sharing | NA | 0.36 (0.07, 0.88) | 0.21 (0.01, 0.49) | WELLBEING_GCA |
| Causal | 0.16 (0.08, 0.24) | 0 (-1.33, 1.28) | 0.03 (0, 0.25) | WELLBEING_GCA |
| Sharing | NA | 0.93 (-2.8, 4.46) | 0.03 (0, 0.22) | WELLBEING_RFFT |
| Causal | 0.1 (-0.12, 0.33) | -0.58 (-3.79, 2.85) | 0.04 (0, 0.25) | WELLBEING_RFFT |
| Sharing | NA | -0.15 (-3.8, 3.34) | 0.02 (0, 0.2) | WELLBEING_ONEBACK |
| Causal | -0.01 (-0.17, 0.14) | -0.04 (-3.63, 3.21) | 0.03 (0, 0.24) | WELLBEING_ONEBACK |
| Sharing | NA | 0.03 (-2.92, 3.55) | 0.02 (0, 0.2) | WELLBEING_RT |

| | | | | |
|---|---|---|---|---|
| Causal | 0 (-0.14, 0.14) | 0.05 (-2.65, 3.48) | 0.03 (0, 0.24) | WELLBEING_RT |
| Sharing | NA | 0.71 (-1.91, 3.19) | 0.05 (0, 0.26) | WELLBEING_OCL |
| Causal | 0.11 (-0.06, 0.28) | -0.07 (-3.12, 2.86) | 0.04 (0, 0.25) | WELLBEING_OCL |
| | | Exposure: Anxiety | | |
| Sharing | NA | 0 (-0.13, 0.13) | 0.06 (0, 0.27) | ANX_GCA |
| Causal | 0 (-0.04, 0.04) | 0 (-0.13, 0.13) | 0.06 (0, 0.3) | ANX_GCA |
| Sharing | NA | -0.01 (-0.26, 0.25) | 0.05 (0, 0.26) | ANX_RFFT |
| Causal | -0.01 (-0.06, 0.04) | -0.01 (-0.26, 0.25) | 0.05 (0, 0.27) | ANX_RFFT |
| Sharing | NA | 0.03 (-0.32, 0.37) | 0.06 (0, 0.29) | ANX_ONEBACK |
| Causal | 0.05 (-0.07, 0.17) | 0 (-0.34, 0.35) | 0.06 (0, 0.3) | ANX_ONEBACK |
| Sharing | NA | -0.02 (-0.27, 0.25) | 0.06 (0, 0.28) | ANX_RT |
| Causal | -0.03 (-0.1, 0.05) | 0 (-0.26, 0.26) | 0.06 (0, 0.29) | ANX_RT |
| Sharing | NA | 0.01 (-0.35, 0.37) | 0.05 (0, 0.26) | ANX_OCL |
| Causal | 0.01 (-0.07, 0.09) | 0 (-0.35, 0.36) | 0.06 (0, 0.28) | ANX_OCL |
| | | Exposure: PANAS Negative | | |
| Sharing | NA | -0.06 (-0.52, 0.45) | 0.05 (0, 0.27) | PANASNEG_GCA |
| Causal | -0.05 (-0.15, 0.06) | 0 (-0.49, 0.49) | 0.05 (0, 0.28) | PANASNEG_GCA |
| Sharing | NA | -0.35 (-1.08, 0.63) | 0.09 (0, 0.35) | PANASNEG_RFFT |
| Causal | -0.17 (-0.33, 0) | 0 (-0.93, 0.96) | 0.06 (0, 0.28) | PANASNEG_RFFT |
| Sharing | NA | -0.1 (-1.05, 0.88) | 0.05 (0, 0.26) | PANASNEG_ONEBACK |
| Causal | -0.05 (-0.23, 0.14) | -0.02 (-1.02, 0.95) | 0.06 (0, 0.29) | PANASNEG_ONEBACK |
| Sharing | NA | -0.05 (-0.88, 0.85) | 0.05 (0, 0.26) | PANASNEG_RT |
| Causal | -0.05 (-0.21, 0.12) | 0.01 (-0.85, 0.89) | 0.05 (0, 0.28) | PANASNEG_RT |
| Sharing | NA | -0.22 (-1.12, 0.87) | 0.06 (0, 0.29) | PANASNEG_OCL |
| Causal | -0.11 (-0.28, 0.06) | 0 (-1.01, 1.06) | 0.05 (0, 0.28) | PANASNEG_OCL |
| | | Exposure: PANAS Positive | | |
| Sharing | NA | 0.04 (-0.52, 0.58) | 0.05 (0, 0.27) | PANASPOS_GCA |
| Causal | 0.05 (-0.1, 0.19) | 0 (-0.54, 0.56) | 0.06 (0, 0.28) | PANASPOS_GCA |
| Sharing | NA | 0.25 (-0.92, 1.27) | 0.07 (0, 0.3) | PANASPOS_RFFT |
| Causal | 0.17 (-0.1, 0.43) | 0.02 (-1.11, 1.15) | 0.06 (0, 0.3) | PANASPOS_RFFT |
| Sharing | NA | 0.02 (-1.09, 1.14) | 0.05 (0, 0.26) | PANASPOS_ONEBACK |
| Causal | 0.01 (-0.24, 0.27) | 0 (-1.1, 1.14) | 0.06 (0, 0.28) | PANASPOS_ONEBACK |

| | | | | |
|---|---|---|---|---|
| Sharing | NA | 0.06 (-0.91, 1.03) | 0.06 (0, 0.27) | PANASPOS_RT |
| Causal | 0.04 (-0.19, 0.28) | 0.02 (-0.96, 1.02) | 0.06 (0, 0.29) | PANASPOS_RT |
| Sharing | NA | -0.07 (-0.97, 0.9) | 0.06 (0, 0.28) | PANASPOS_OCL |
| Causal | -0.07 (-0.31, 0.17) | 0.01 (-0.94, 0.97) | 0.06 (0, 0.29) | PANASPOS_OCL |

Gamma=Estimate of causal effect of the exposure on the outcome; Eta=Estimate of the effect of a heritable shared factor on the outcome (i.e., effect of correlated pleiotropy); Q=Proportion of variants that act on the exposure and the outcome via a heritable shared factor (i.e., proportion of variants displaying correlated pleiotropy).

#### SUPPLEMENTARY FIGURES

***Figure S1. Flow Diagram of Lifelines Genetic Data*** [Taken from (Giollabhui et al., 2024)]

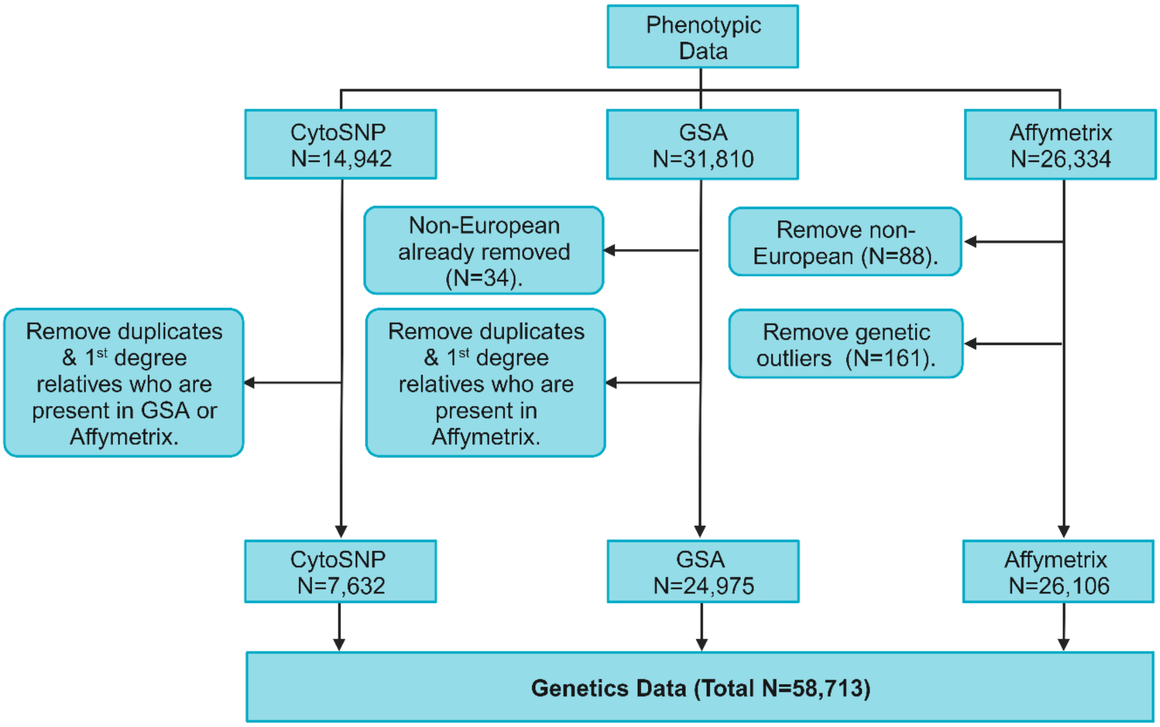

Note: Phenotypic data includes individuals in Lifelines aged ≥ 18 years and excludes individuals who self-report any one of the following health conditions: Alzheimer's disease, dementia, epilepsy, multiple sclerosis, Parkinson's disease, stroke. Lists of duplicates and 1st-degree relatives to remove between chips were provided by Lifelines. Duplicates and 1st-degree relatives were removed from CytoSNP due to its poorer imputation quality compared to the other chips. Genetic outliers and non-European individuals were also provided by Lifelines. Figure created using *Biorender.com*.

***Figure S2. Manhattan plot of GWAS on PANAS positive and negative subscales (N=57,946). Note: variants with MAF < 0.01 not excluded.***

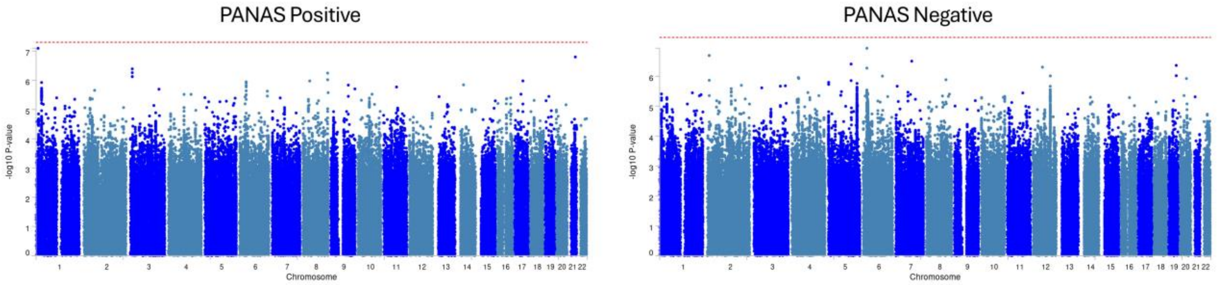

***Figure S3. Manhattan plot of GWAS on cognitive task performance (N range= N=35,729-36,783). Note: variants with MAF < 0.01 not excluded.***

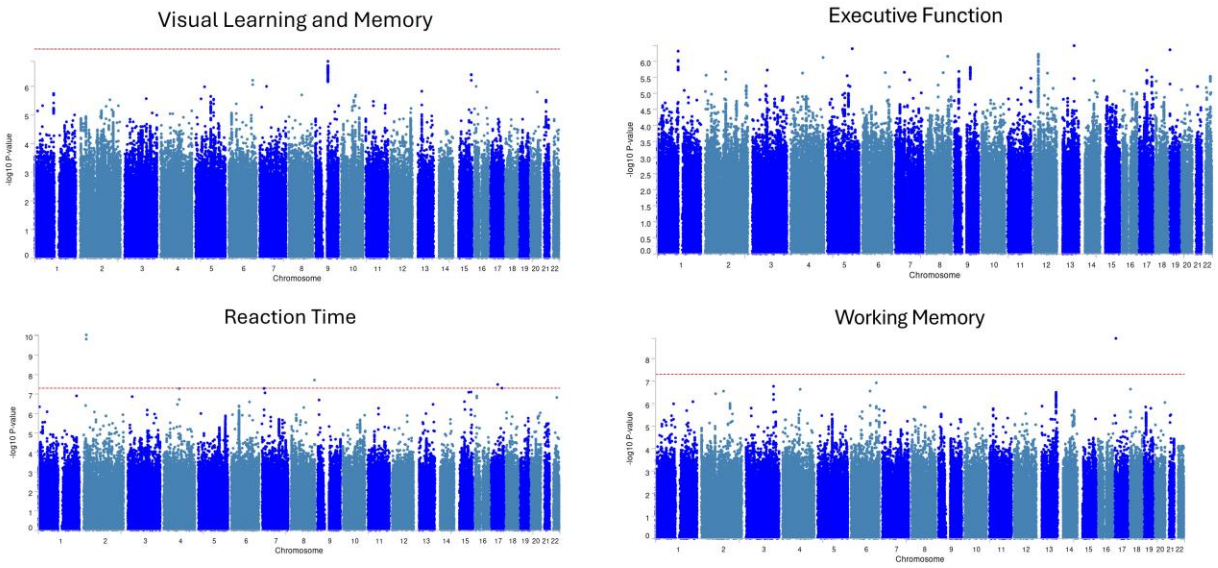

***Figure S4. Locus zoom plot of region around rs2920287.***

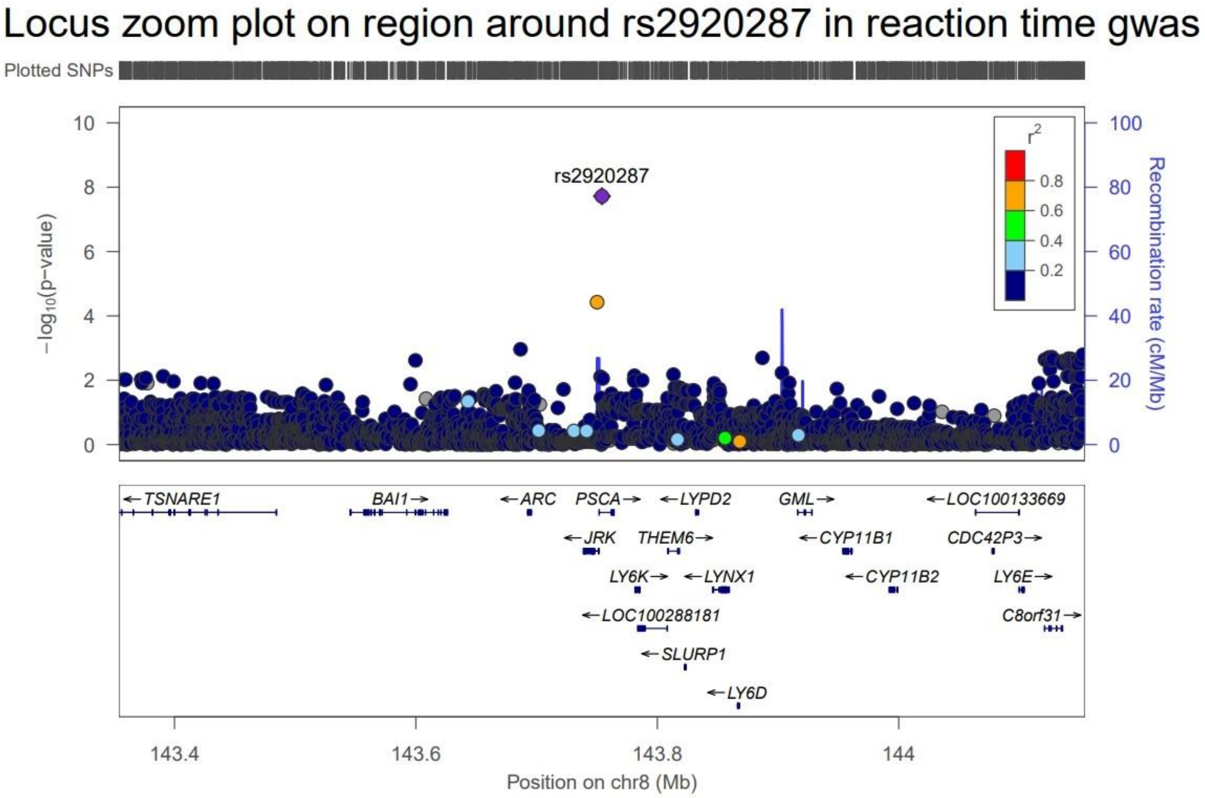

***Figure S5. Tissue specificity of prioritised genes for each phenotype.***

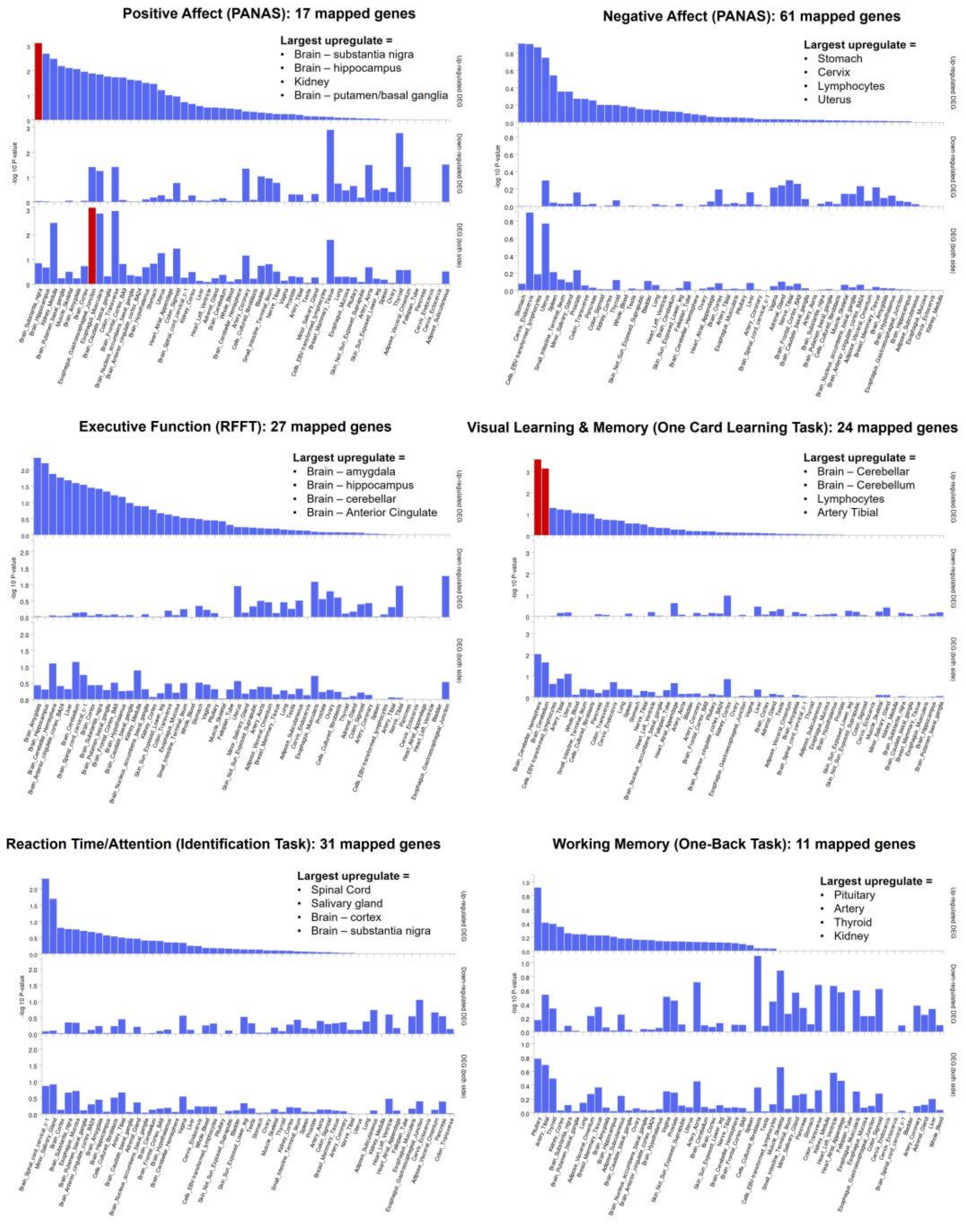

*Note: positional mapping of suggestive [p< 5x10-6] SNPs in FUMA and GTEx v8. Significantly enriched DEG sets (Bonferroni-corrected p<0.05) are highlighted in red.*

***Figure S6. Tissue specificity of prioritised genes for each Lifelines phenotype (using positional mapping, eQTL mapping, and chromatin interaction mapping of suggestive [p< 5x10$^{-6}$] SNPs in FUMA; and GTEx v8).***

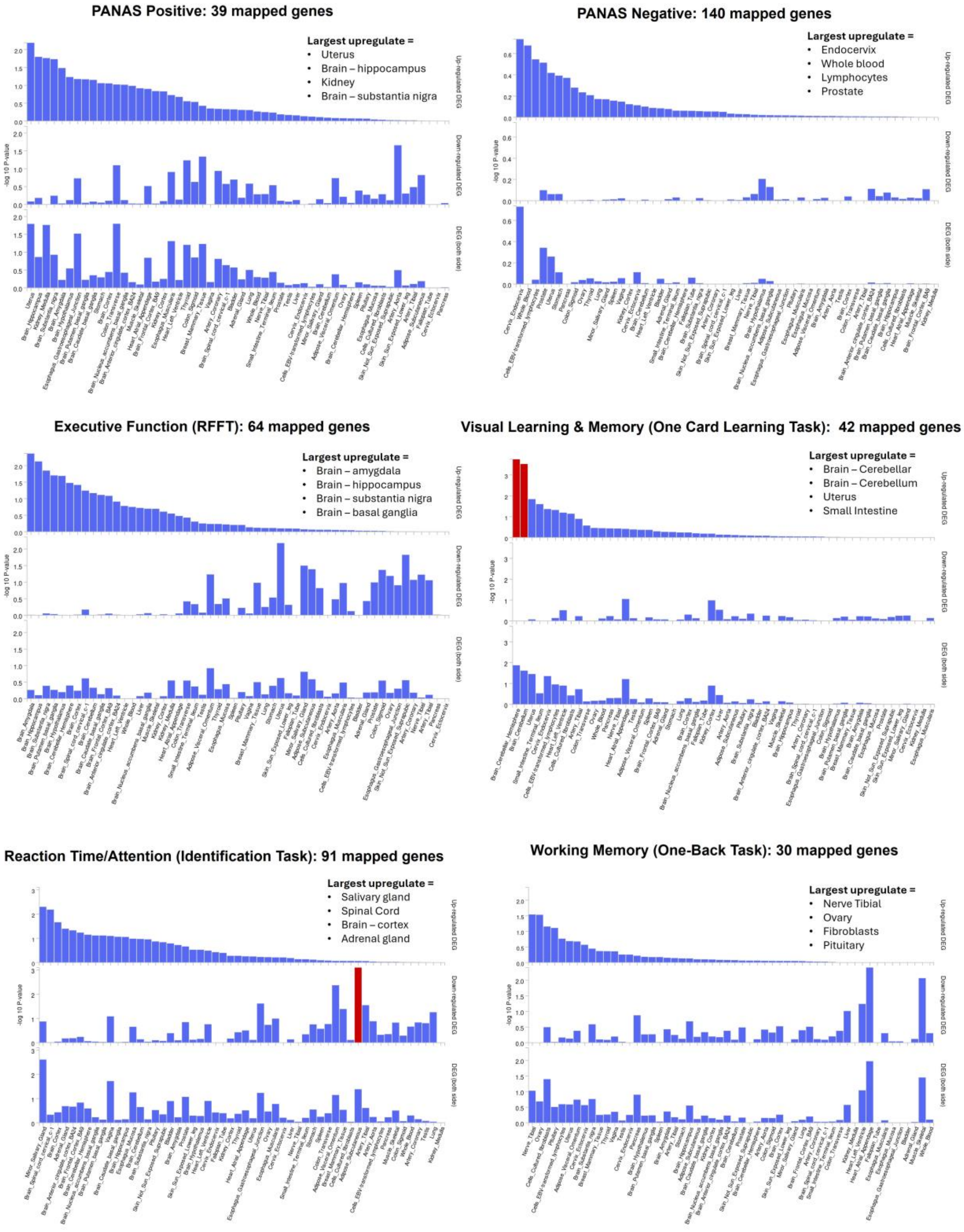

***Figure S7. MR sensitivity plots: GCA on PANAS Negative.*** *Graphs show (A) scatter plot of results from four MR methods, (B) funnel plot showing each SNP causal estimate against its precision (asymmetry may indicate directional pleiotropy), (C) leave-one-out plot showing inverse-variance weighted estimates after removing each individual SNP in turn, (D) forest plot of causal estimates for each SNP.*

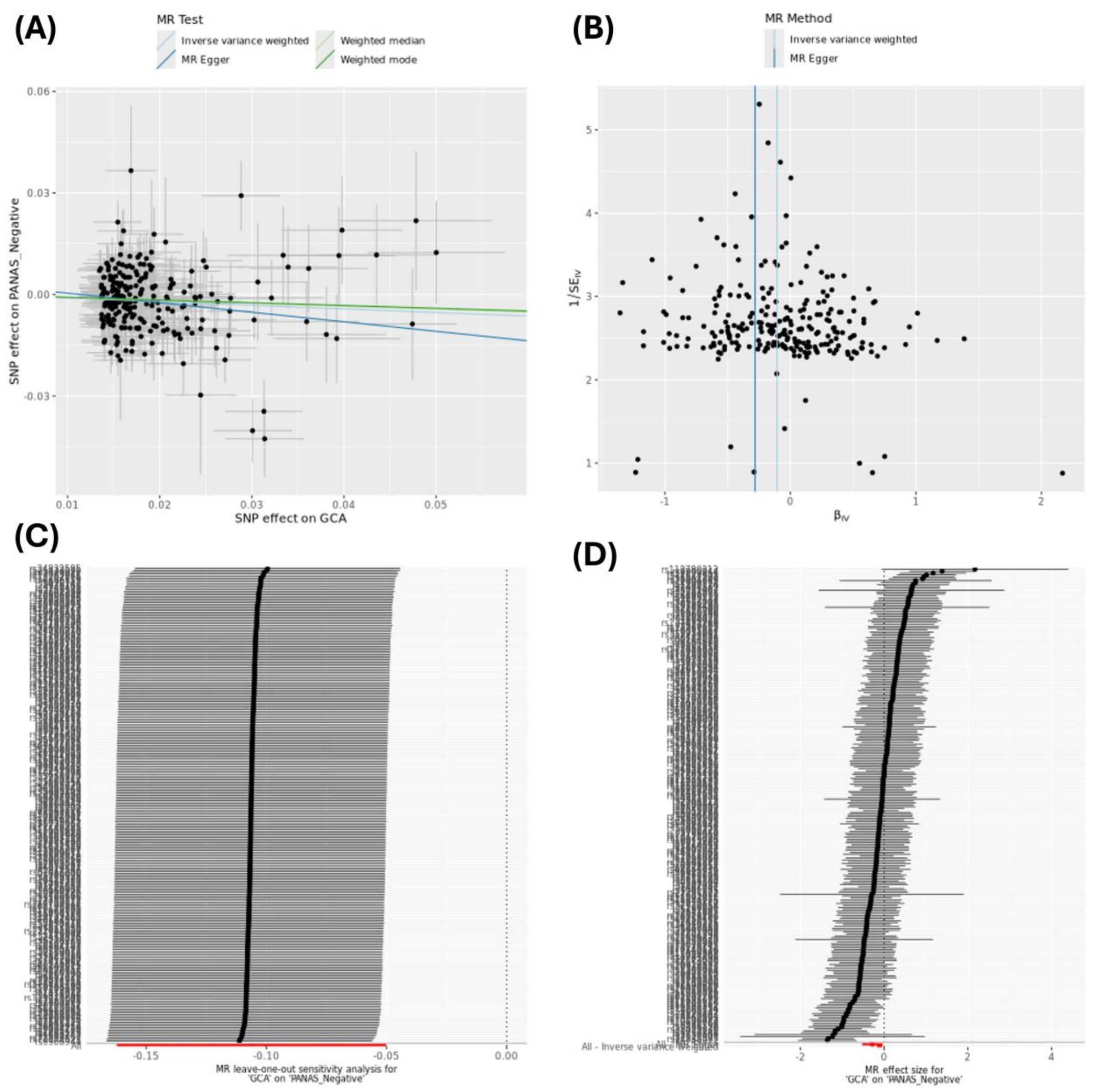

***Figure S8. MR sensitivity plots: GCA on PANAS Positive.*** *Graphs show (A) scatter plot of results from four MR methods, (B) funnel plot showing each SNP causal estimate against its precision (asymmetry may indicate directional pleiotropy), (C) leave-one-out plot showing inverse-variance weighted estimates after removing each individual SNP in turn, (D) forest plot of causal estimates for each SNP.*

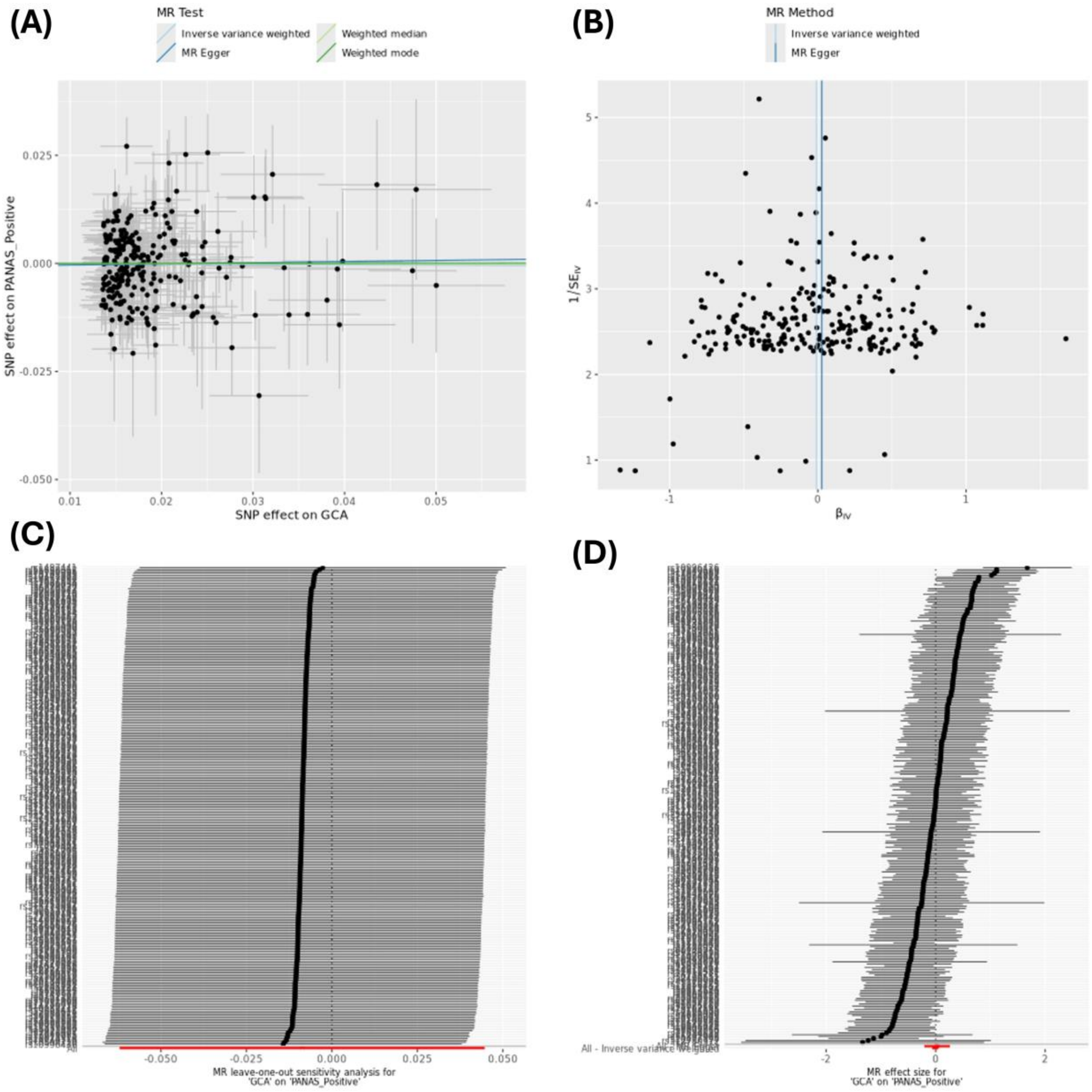

***Figure S9. MR sensitivity plots: GCA on Depression.*** *Graphs show (A) scatter plot of results from four MR methods, (B) funnel plot showing each SNP causal estimate against its precision (asymmetry may indicate directional pleiotropy), (C) leave-one-out plot showing inverse-variance weighted estimates after removing each individual SNP in turn, (D) forest plot of causal estimates for each SNP.*

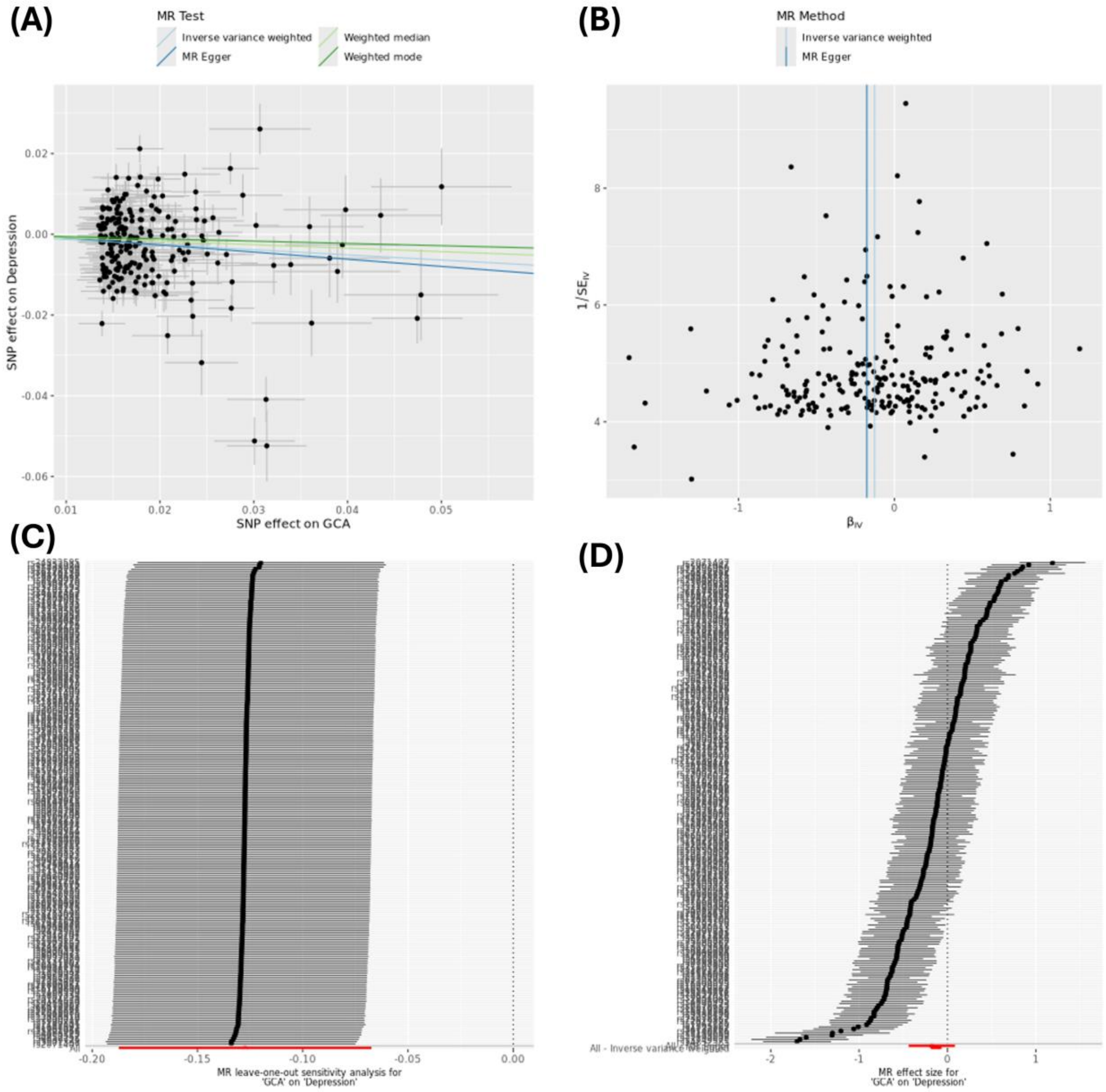

***Figure S10. MR sensitivity plots: GCA on Anxiety.*** *Graphs show (A) scatter plot of results from four MR methods, (B) funnel plot showing each SNP causal estimate against its precision (asymmetry may indicate directional pleiotropy), (C) leave-one-out plot showing inverse-variance weighted estimates after removing each individual SNP in turn, (D) forest plot of causal estimates for each SNP.*

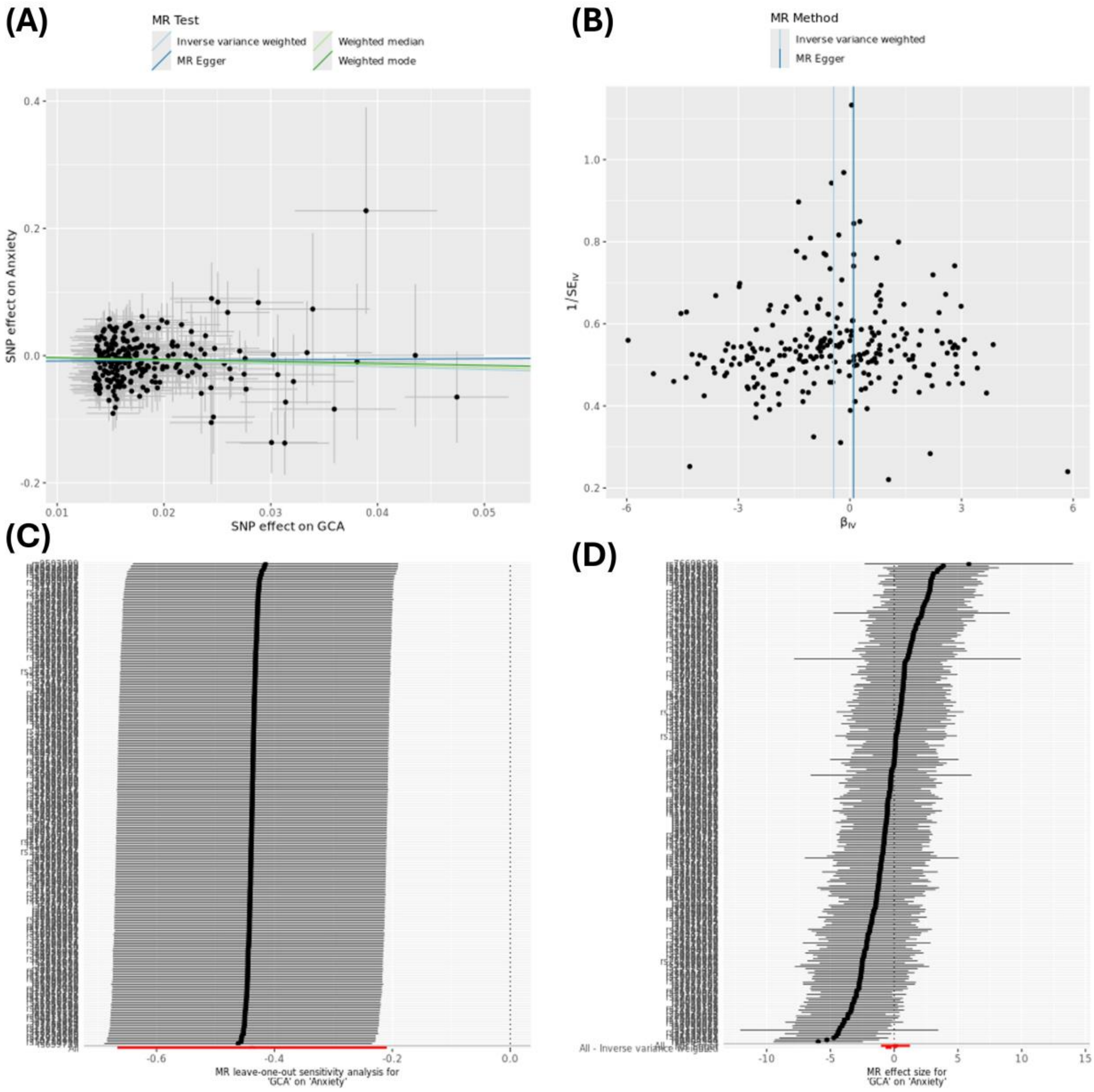

***Figure S11. MR sensitivity plots: GCA on Wellbeing.*** *Graphs show (A) scatter plot of results from four MR methods, (B) funnel plot showing each SNP causal estimate against its precision (asymmetry may indicate directional pleiotropy), (C) leave-one-out plot showing inverse-variance weighted estimates after removing each individual SNP in turn, (D) forest plot of causal estimates for each SNP.*

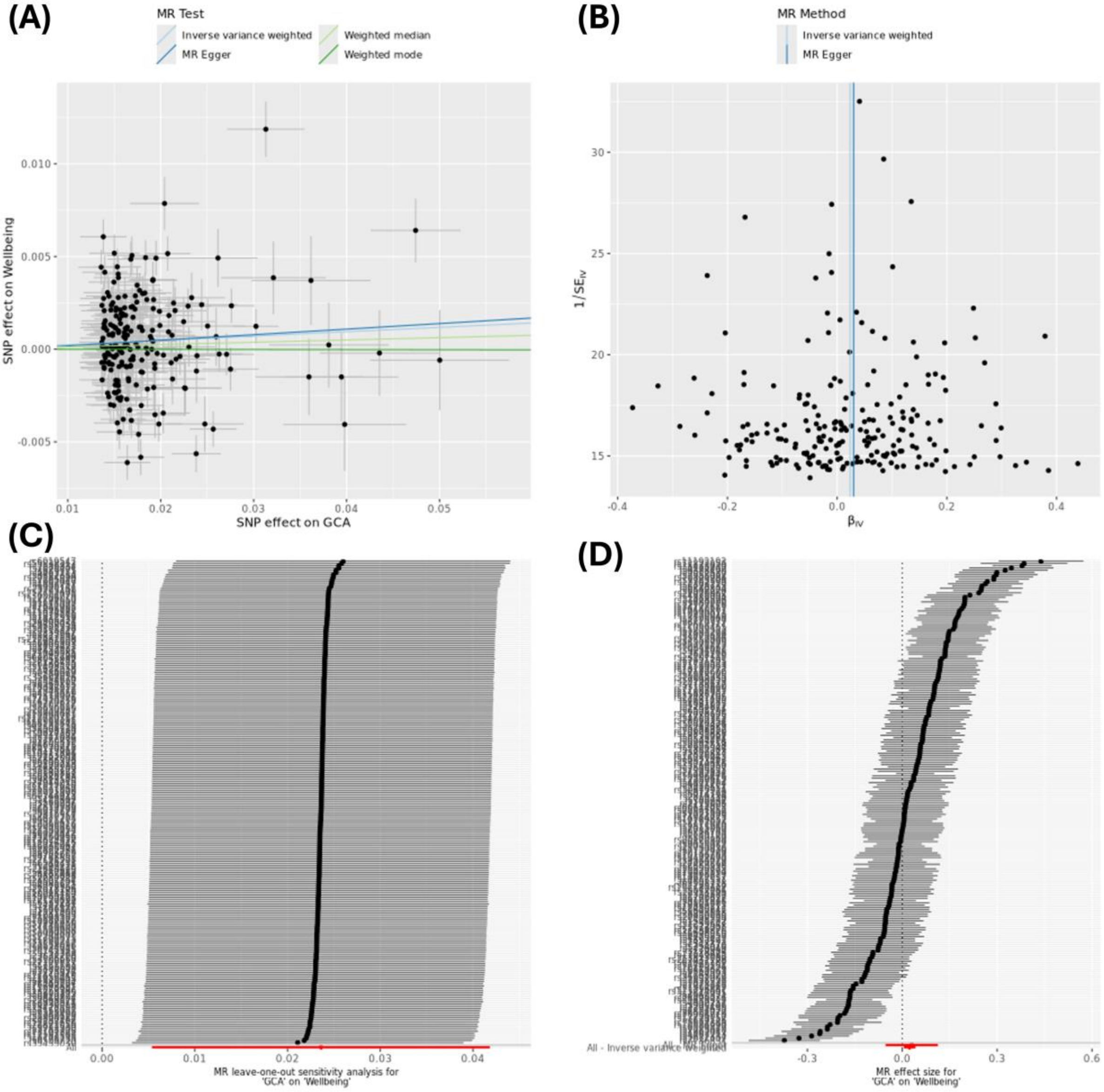

***Figure S12. MR sensitivity plots: Depression on GCA.*** *Graphs show (A) scatter plot of results from four MR methods, (B) funnel plot showing each SNP causal estimate against its precision (asymmetry may indicate directional pleiotropy), (C) leave-one-out plot showing inverse-variance weighted estimates after removing each individual SNP in turn, (D) forest plot of causal estimates for each SNP.*

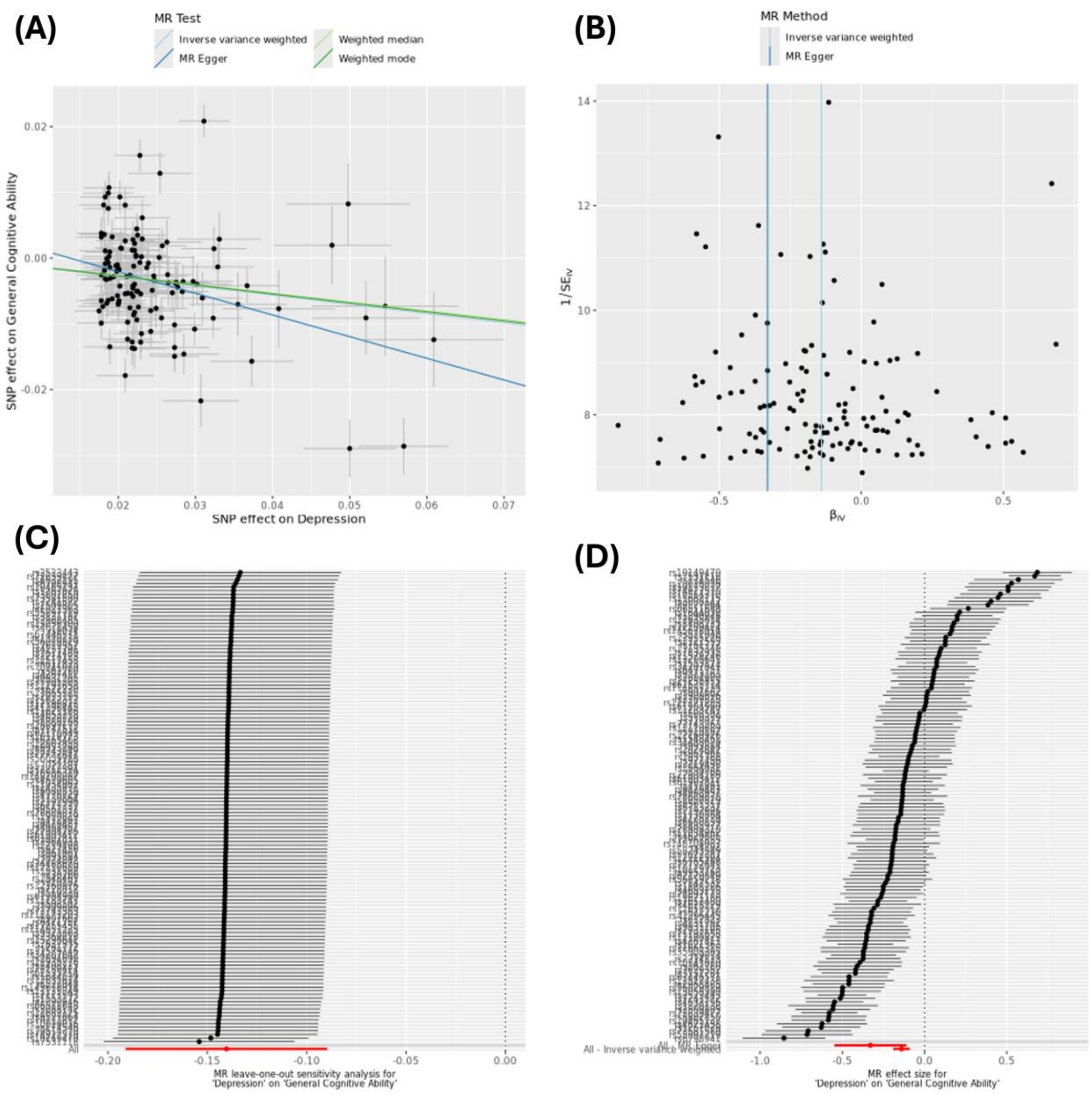

***Figure S13. MR sensitivity plots: Wellbeing on GCA.*** *Graphs show (A) scatter plot of results from four MR methods, (B) funnel plot showing each SNP causal estimate against its precision (asymmetry may indicate directional pleiotropy), (C) leave-one-out plot showing inverse-variance weighted estimates after removing each individual SNP in turn, (D) forest plot of causal estimates for each SNP.*

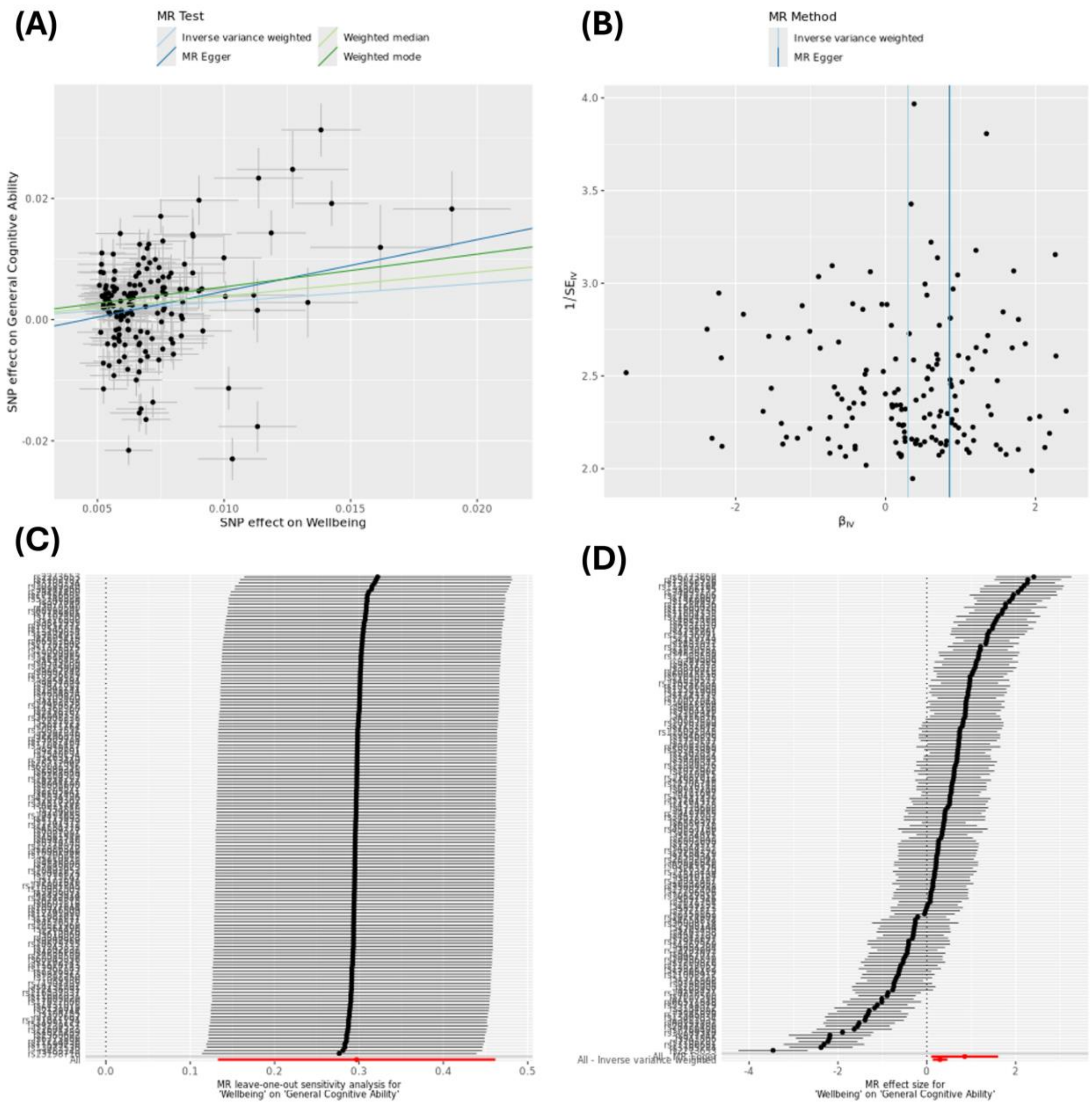

***Figure S14. MR sensitivity plots: PANAS Negative on GCA.*** *Graphs show (A) scatter plot of results from four MR methods, (B) funnel plot showing each SNP causal estimate against its precision (asymmetry may indicate directional pleiotropy), (C) leave-one-out plot showing inverse-variance weighted estimates after removing each individual SNP in turn, (D) forest plot of causal estimates for each SNP.*

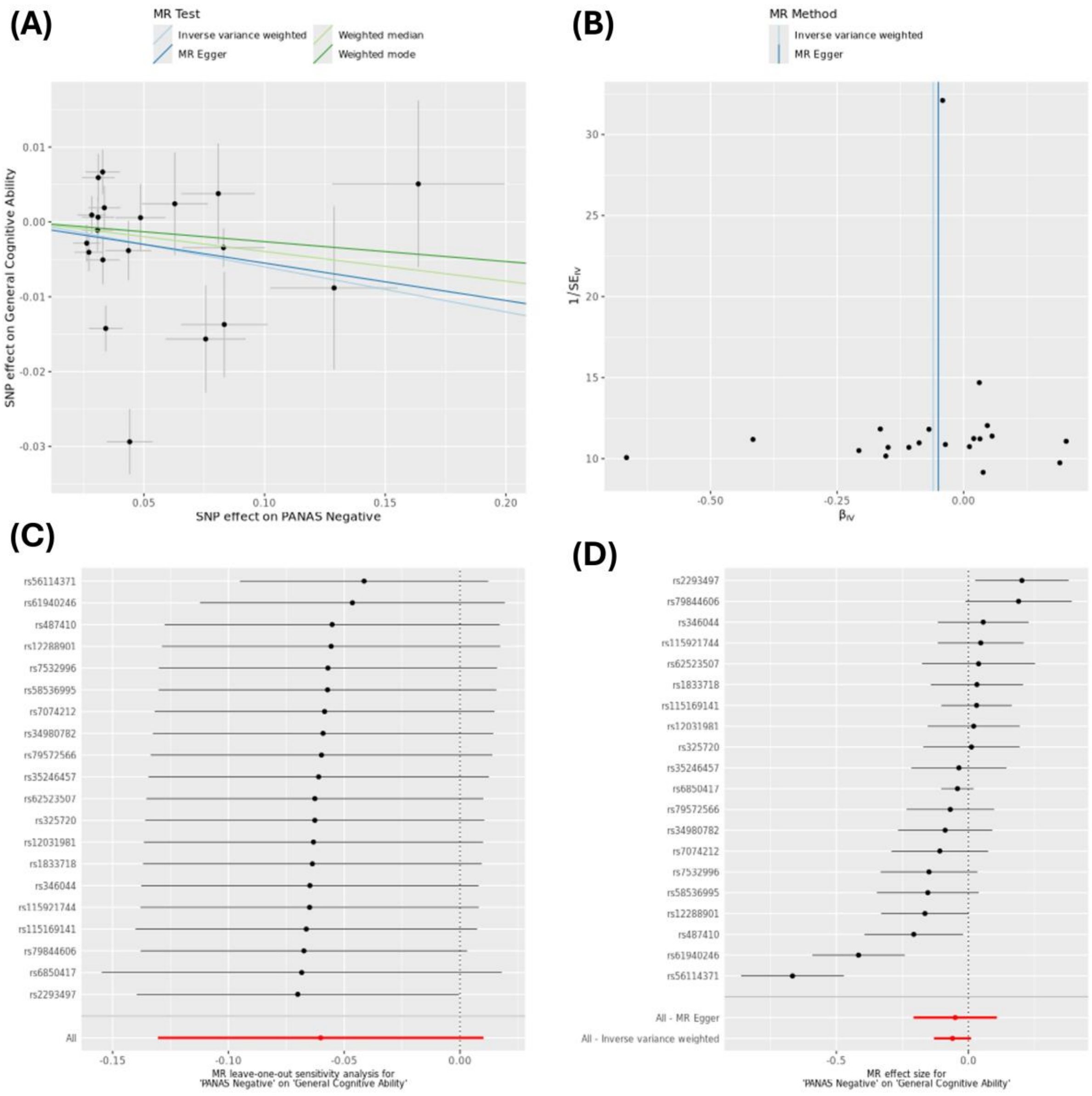

***Figure S15. MR sensitivity plots: PANAS Positive on GCA.*** *Graphs show (A) scatter plot of results from four MR methods, (B) funnel plot showing each SNP causal estimate against its precision (asymmetry may indicate directional pleiotropy), (C) leave-one-out plot showing inverse-variance weighted estimates after removing each individual SNP in turn, (D) forest plot of causal estimates for each SNP.*

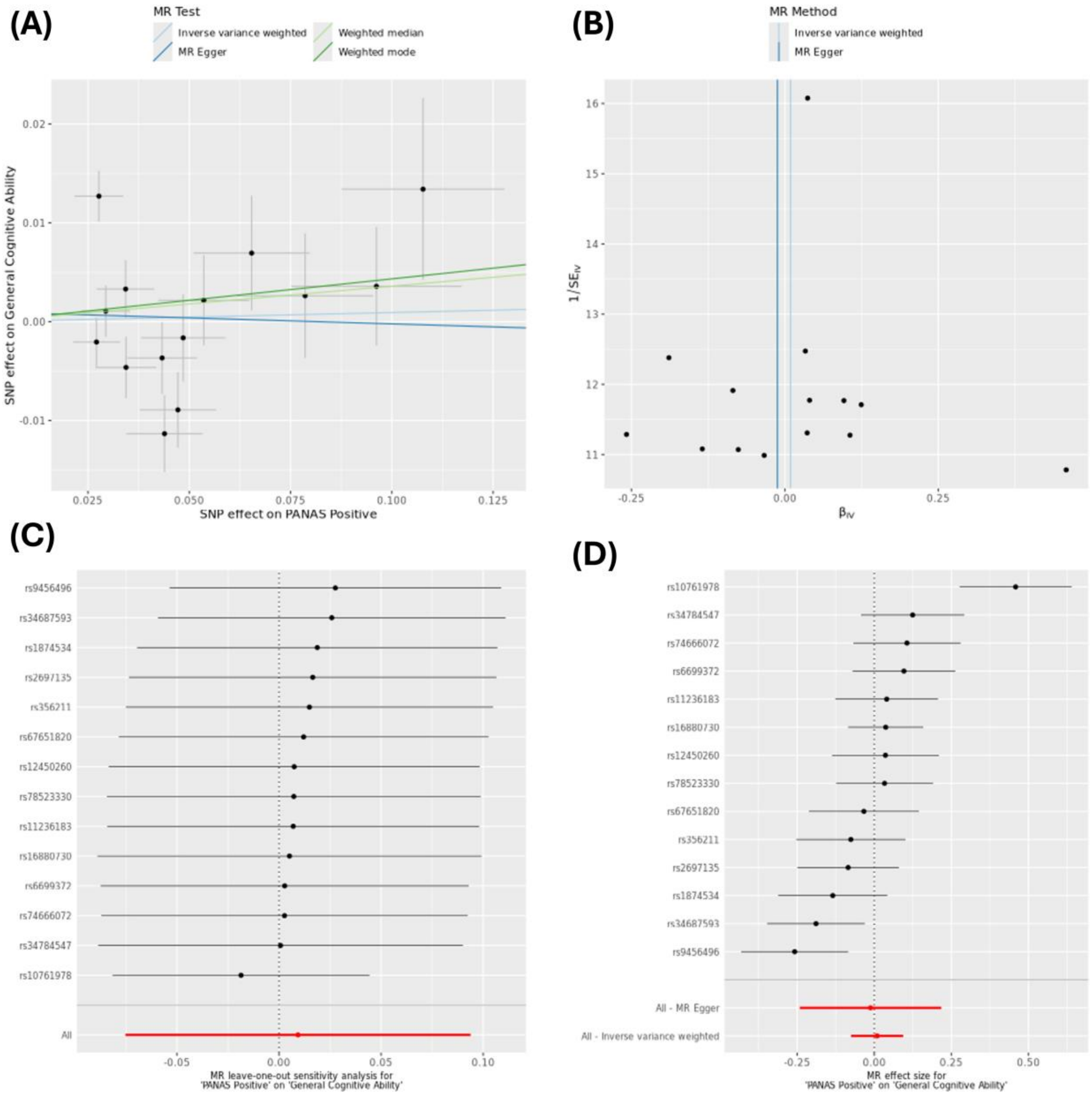

***Figure S16. Comparison of Mendelian randomization results using GWAS data used in this study, alongside population and within-sibship GWAS estimates from Howe et al., (2022).***

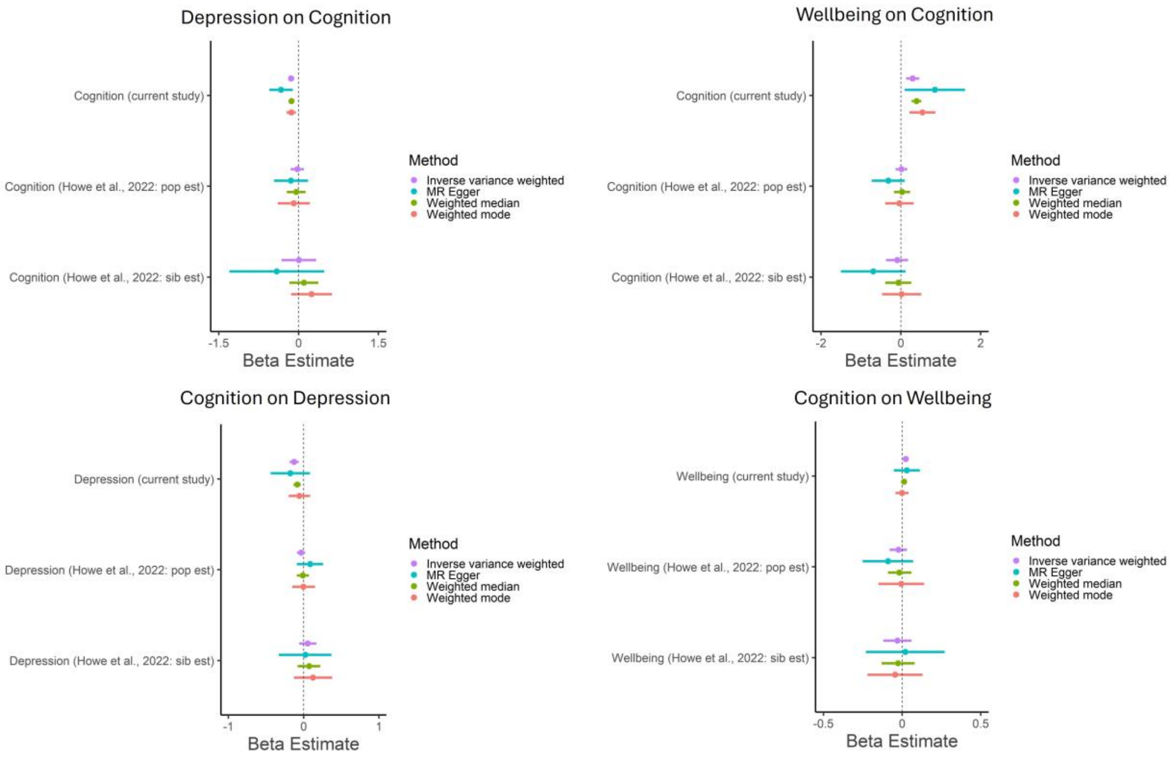
